# Combining genomic and epidemiological data to compare the transmissibility of SARS-CoV-2 lineages

**DOI:** 10.1101/2021.07.01.21259859

**Authors:** Mary E. Petrone, Jessica E. Rothman, Mallery I. Breban, Isabel M. Ott, Alexis Russell, Erica Lasek-Nesselquist, Kevin Kelly, Greg Omerza, Nicholas Renzette, Anne E. Watkins, Chaney C. Kalinich, Tara Alpert, Anderson F. Brito, Rebecca Earnest, Irina R. Tikhonova, Christopher Castaldi, John P. Kelly, Matthew Shudt, Jonathan Plitnick, Erasmus Schneider, Steven Murphy, Caleb Neal, Eva Laszlo, Ahmad Altajar, Claire Pearson, Anthony Muyombwe, Randy Downing, Jafar Razeq, Linda Niccolai, Madeline S. Wilson, Margaret L. Anderson, Jianhui Wang, Chen Liu, Pei Hui, Shrikant Mane, Bradford P. Taylor, William P. Hanage, Marie L. Landry, David R. Peaper, Kaya Bilguvar, Joseph R. Fauver, Chantal B.F. Vogels, Lauren M. Gardner, Virginia E. Pitzer, Kirsten St. George, Mark D. Adams, Nathan D. Grubaugh

## Abstract

Emerging SARS-CoV-2 variants have shaped the second year of the COVID-19 pandemic and the public health discourse around effective control measures. Evaluating the public health threat posed by a new variant is essential for appropriately adapting response efforts when community transmission is detected. However, this assessment requires that a true comparison can be made between the new variant and its predecessors because factors other than the virus genotype may influence spread and transmission. In this study, we develop a framework that integrates genomic surveillance data to estimate the relative effective reproduction number (R_t_) of co-circulating lineages. We use Connecticut, a state in the northeastern United States in which the SARS-CoV-2 variants B.1.1.7 and B.1.526 co-circulated in early 2021, as a case study for implementing this framework. We find that the R_t_ of B.1.1.7 was 6-10% larger than that of B.1.526 in Connecticut in the midst of a COVID-19 vaccination campaign. To assess the generalizability of this framework, we apply it to genomic surveillance data from New York City and observe the same trend. Finally, we use discrete phylogeography to demonstrate that while both variants were introduced into Connecticut at comparable frequencies, clades that resulted from introductions of B.1.1.7 were larger than those resulting from B.1.526 introductions. Our framework, which uses open-source methods requiring minimal computational resources, may be used to monitor near real-time variant dynamics in a myriad of settings.

## Introduction

The emergence of novel SARS-CoV-2 variants has shaped the second year of the COVID-19 pandemic^1–3^ and illustrated the role of genomic epidemiology in facilitating an appropriate, effective, and timely public health response^4^. In particular, genomic epidemiology can determine the source and frequency of new variant introductions into a community, thus indicating where additional surveillance is needed. However, this assessment requires the prior establishment of a robust genomic surveillance system. Once community transmission is documented, the efficacy of control methods should be re-evaluated by assessing the public health risk posed by the variant in comparison to other variants in circulation. This second objective is particularly challenging because factors other than the virus genotype influence its transmission and spread^5–7^. Specifically, competition between virus lineages and local levels of immunity could impact the relative success of a new variant compared to its predecessors. Therefore, we should compare lineages that have emerged concurrently in the same human and virus population as the variant under scrutiny. For an accurate risk assessment of emerging variants, we must control for the epidemiological context. Instances in which these criteria are met are both rare and exceptionally informative.

At the beginning of 2021 two variants of public health significance synchronously emerged in Connecticut, a United States (US) state with high rates of SARS-CoV-2 genomic surveillance. SARS-CoV-2 lineage B.1.526 (designated as variant ‘iota’) was detected in New York in December 2020^8^. Shortly thereafter, cases of B.1.1.7 (‘alpha’), the variant first characterized in the United Kingdom, were identified in the northeastern US. Due to evidence collected in the United Kingdom that this variant was more transmissible than other lineages, B.1.1.7 was expected to become dominant in the US by March^9–11^. Instead, B.1.526 co-circulated in New York with B.1.1.7 and may have slowed the decline of COVID-19 incidence in New York City^12^. Both variants were initially detected in Connecticut within the first two weeks of January 2021, likely introduced by infected travelers, and continued to co-circulate in the state for months^10^.

In this study, we develop a framework to measure the relative transmissibility of B.1.1.7 and B.1.526 by combining epidemiological and genomic data collected in Connecticut between January and May 2021. We first measure the relative transmissibility of these variants by modeling their growth rates and time-varying effective reproduction numbers following their emergence. Both metrics indicate that B.1.1.7 was 6-10% more transmissible than B.1.526 when these variants circulated in the same population. Interestingly, these findings are consistent with the relationship we observed in New York City where B.1.526 was established before B.1.1.7. We next estimate the timing, number, and clade size following sustained introductions of each variant into Connecticut to determine whether the apparent fitness advantage we observed for B.1.1.7 could be attributed to a higher rate of introductions over our study period rather than higher fitness. We use discrete phylogeography to infer the source and number of introductions for each variant and find that both were introduced at comparable rates, but the size of clades precipitated by introductions of B.1.1.7 were on average larger than those formed from introductions of B.1.526. The concordance of our epidemiological and phylodynamic results indicate that B.1.1.7 had a fitness advantage over B.1.526 when potentially confounding factors were controlled. Through this case study, we demonstrate that our framework, which utilizes open-source computational methods, is a robust and useful tool for continually monitoring the dynamics of SARS-CoV-2 variants in near real time.

## Results

### Rapid rise in B.1.1.7 and B.1.526 prevalence in Connecticut and New York City

The rapid spread of SARS-CoV-2 lineages B.1.1.7 in the United Kingdom^13^ and B.1.526 (including sublineages B.1.526.1 and B.1.526.2) in New York City^12,14^ suggested that these variants have a competitive advantage over other SARS-CoV-2 lineages. Both variants are defined by key amino acid substitutions in the spike protein that may contribute to this advantage. We therefore hypothesized that B.1.1.7 and B.1.526 would compete for dominance in Connecticut soon after they emerged. To test this hypothesis, we measured the daily frequencies and growth rates of B.1.1.7 and B.1.526 in Connecticut and compared these patterns to those observed in New York City (**Fig. 1**). Our analysis revealed that B.1.1.7 and B.1.526 displaced nearly all other lineages circulating in both regions within three months of emergence. Moreover, the frequency of B.1.1.7 grew at a faster rate than B.1.526.

**Figure 1:**
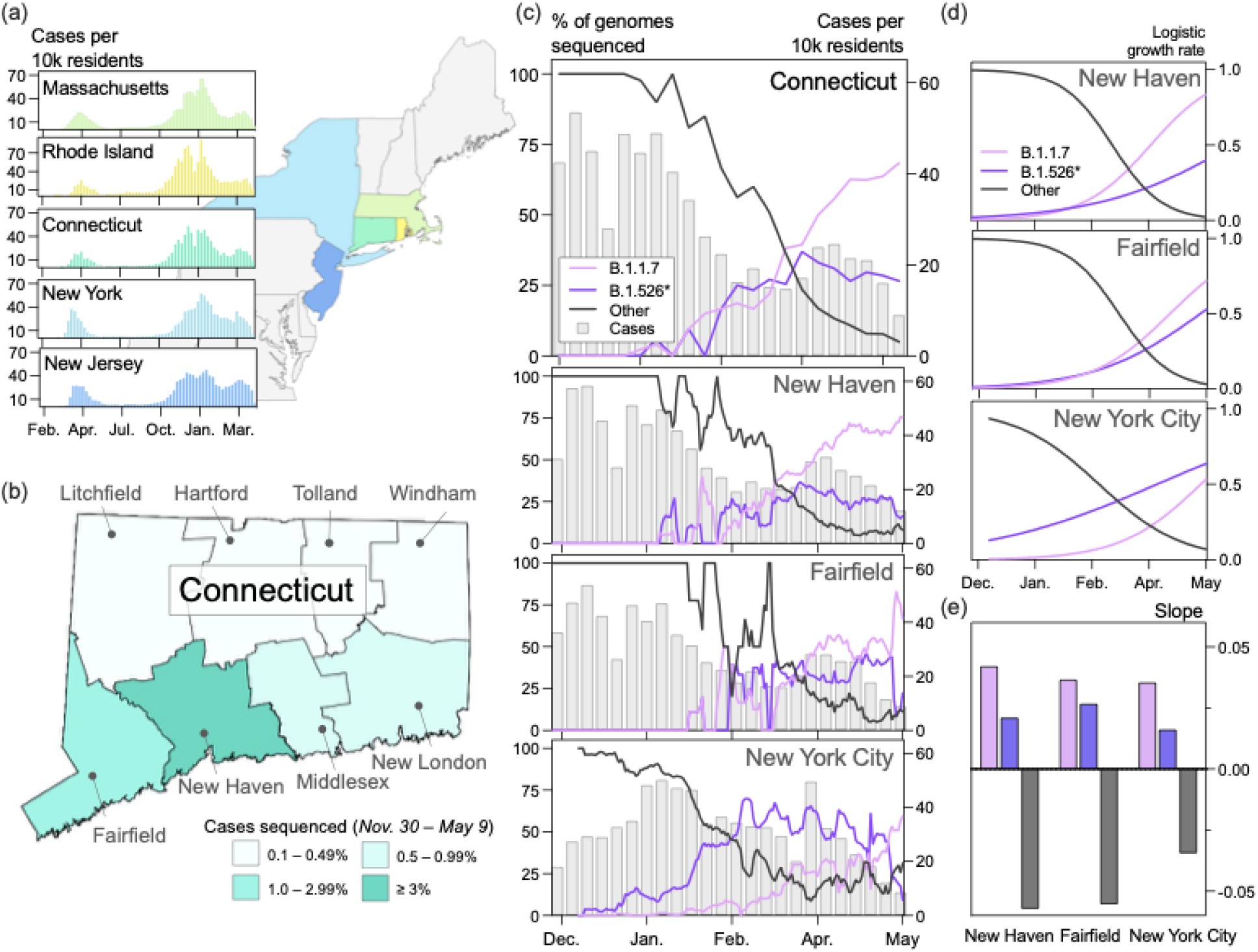
B.1.1.7 and B.1.526^*^ dominated the circulating SARS-CoV-2 populations in Connecticut and New York City in early 2021. **(a)** Trends in COVID-19 incidence were consistent across northeastern states throughout the pandemic. (*map*) Connecticut (teal) is bordered by New York, Rhode Island, and Massachusetts. New York City is less than 50 miles from Fairfield County. Weekly COVID-19 incidence was tabulated according to the Johns Hopkins COVID-19 portal (https://github.com/CSSEGISandData/COVID-19). Shapefile source: United States Census Bureau. **(b)** New Haven County led the state in the percentage of COVID-19 cases sequenced between November 30, 2020 and May 9, 2021 (3.33%). During this period, 0.51% of COVID-19 cases in New York City were sequenced. Genomes that were collected through targeted variant screening (e.g., spike-gene target failure) were excluded from this analysis. Shapefile source: the Connecticut Department of Energy & Environmental Protection (DEEP) Geographic Information Systems Open Data Website. **(c)** Together, B.1.1.7 and B.1.526* variants displaced nearly all other SARS-CoV-2 lineages in New Haven County (n = 2,086), Fairfield County (n = 612), and New York City (n = 4,528). The lineages of sequenced viruses were assigned using pangolin v.2.4.2. The lineages B.1.526, B.1.526.1, and B.1.526.2 were assigned to the general lineage category ‘B.1.526^*^’. We calculated a 7-day rolling average for the proportion of B.1.1.7, B.1526^*^, and ‘other’ SARS-CoV-2 lineages sequenced in our dataset. **(d)** Logistic regression of the growth rates per lineage using Rv.4.0.1. Line colors correspond to the legend in (c). (**e**) Slope of logistic growth shown in (d). Bar colors correspond to the legend in (c).

Unlike the situation in New York City, which may be the origin of B.1.526, B.1.1.7 and B.1.526 emerged concurrently in Connecticut through infected travelers. Connecticut is a state in the northeast US, bordered by Rhode Island, Massachusetts, and New York (**Fig. 1a**, *map*). These states experienced synchronous waves of COVID-19 incidence throughout the pandemic (**Fig. 1a**, *graphs*). We first detected B.1.1.7 in Connecticut on January 6, 2021 (sample collection date) in New Haven County ^10^, and we detected the first B.1.526 soon after on January 14, 2021.

As of early May 2021, B.1.526 contains two sublineages, and SARS-CoV-2 sequences belonging to this clade can be classified as either B.1.526, B.1.526.1, or B.1.526.2 (**Supplementary Fig. 1**). This clade has a poorly resolved phylogenetic relationship and the lineages have different patterns of three key amino acid substitutions in the spike gene: L452R, S477N, and E484K (**Supplementary Fig. 1a**)^12,14–17^. B.1.526 does not include the L452R substitution, but ∼20% include S477N and ∼75% include E484K^3,18^; B.1.526.1 only includes L452R; only includes S477N. The CDC classified B.1.526 and B.1.526.1 as ‘Variants of Interest’ (VOI), but did not include B.1.526.2 in this category^15^. Even with these ostensibly important molecular differences, all three lineages had similar epidemiological dynamics and co-circulated in Connecticut since February 2021 at relatively equal frequencies (**Supplementary Fig. 1b**). We therefore elected to analyze the dynamics of B.1.526, B.1.526.1, and B.1.526.2 collectively, which we hereafter refer to as B.1.526^*^.

To compare the relative growth rates of B.1.1.7 and B.1.526^*^ over time, we collected and sequenced 2,951 whole SARS-CoV-2 genomes from Connecticut between November 30, 2020 and May 9, 2021 using an unbiased sampling approach. Specifically, we excluded genomes that were targeted for sequencing because of spike-gene target failure or any other anomaly. We assigned PANGO lineages to each genome^19^ and created a general lineage classification with three categories: ‘B.1.1.7’, ‘B.1.526^*^’, or ‘other’. The ‘other’ lineages primarily include B.1.2, B.1.517, B.1.575, and B.1.243, but they also include low frequencies of several ‘Variants of Concern’ (VOCs) and VOIs (**Supplementary Table 1**). We calculated a rolling 7-day average for each general lineage classification to mitigate the impact of daily reporting trends.

In southern Connecticut, B.1.1.7 and B.1.526^*^ collectively rose to above 50% prevalence by March 2021, but the relative prevalence of these variants differed across the region (**Fig. 1c**). Due to the close proximity of New Haven and Fairfield counties to New York City and the large volume of travelers between New York City and southern Connecticut, we hypothesized that the frequency patterns in Connecticut would reflect those observed in New York City. We therefore modeled the logistic growth of each variant across locations (**Fig. 1d**). While the growth rates of B.1.1.7 and B.1.526* were comparable and consistently higher than all other lineages (**Fig. 1e**), we observed heterogeneity in the relative growth of these variants. The estimated logistic growth rate of B.1.1.7 was twice that of B.1.526* in New Haven County (B.1.1.7 = 0.042, B.1.526^*^ = 0.021) and New York City (B.1.1.7 = 0.035, B.1.526^*^ = 0.016). The rate of B.1.1.7. growth was 1.37 times that of B.1.526^*^ in Fairfield County (B.1.1.7 = 0.037, B.1.526^*^ = 0.028). These findings suggest that B.1.1.7 and B.1.526* had a competitive fitness advantage over their predecessors, and, once established, B.1.1.7 may have spread more quickly than B.1.526^*^. This pattern was particularly noticeable in New York City, where B.1.526^*^ emerged first but increased in frequency more slowly than B.1.1.7 (**Fig. 1c**).

### Evidence that B.1.1.7 is more transmissible than B.1.526*

The relative changes in frequency and growth rates reflected by our sequencing data indicated that B.1.1.7 and B.1.526^*^ outcompeted other co-circulating SARS-CoV-2 lineages (**Fig. 1**). They also provided some evidence that the prevalence of B.1.1.7 increased at a faster rate than that of B.1.526^*^ in three different populations. However, these observations did not account for COVID-19 incidence in each population. Over the duration of our study period, the weekly number of reported COVID-19 cases in Connecticut declined, peaking at 53 cases per 10,000 residents and falling to 9 cases per 10,000 residents with fluctuations in between. To more accurately measure the relative transmissibility of B.1.1.7 and B.1.526^*^, we combined the frequency estimates from our genomic data with daily reported COVID-19 cases and estimated the effective reproduction numbers (R_t_), which quantifies the average number of secondary cases from a primary infection, for each variant (**Fig. 2**). In New Haven County, Connecticut, we found that B.1.1.7 was 6-10% more transmissible than B.1.526^*^ (**Fig. 2c**). We obtained consistent albeit noisier results in New York City, providing further evidence that B.1.1.7 was more transmissible than B.1.526* (**Fig. 2f**).

**Figure 2:**
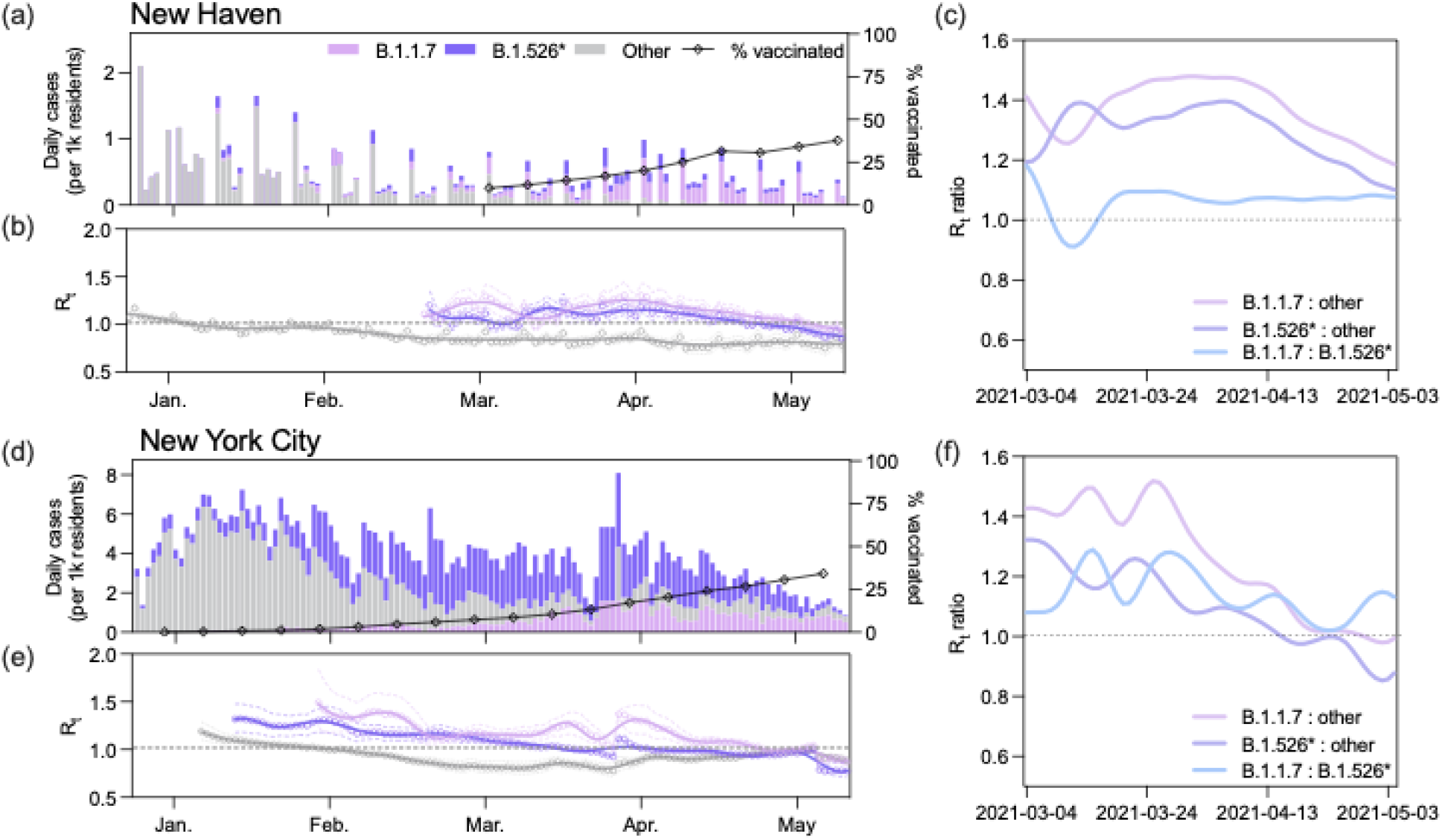
B.1.1.7 had a larger effective reproduction number (R_t_) than B.1.526 during the COVID-19 vaccination campaign. (**a, d**) Daily incidence and full vaccination rates (2 weeks post last dose) of B.1.1.7, B.1.526^*^, and other circulating lineages in New Haven County (a) and New York City (d). Daily cases were assigned to one of three lineage categories (‘B.1.1.7’, ‘B.1.526^*^’, and ‘other’) according to the 7-day rolling average of variant frequency among sequenced cases. ‘B.1.526^*^’ includes the sublineages B.1.526, B.1.526.1, and B.1.526.2. (**b, e**) Time-varying effective reproduction numbers (R_t_) were calculated using the R package *epiEstim*. An R_t_ value above 1 indicates that an infected individual will, on average, infect more than 1 additional person. We assumed a serial interval of mean 5.2 days and standard deviation of 4 days for all lineages. We used a smoothing spline to smooth the daily R_t_ curves (line) with the package, *stat* in R v4.0.1. Non-smoothed estimates are shown as individual points. 0.025 and 0.975 quantiles are shown as dotted lines (**c, f**) Ratios of estimated R_t_ between March 4 and May 4, 2021 calculated using the splines shown in (b) and (e).

We estimated R_t_ for B.1.1.7 and B.1.526^*^ by extrapolating the variant frequencies among sequenced cases to the total number of reported cases in New Haven County (**Fig. 2a**). We selected New Haven County because we sequenced a higher percentage of cases compared to other counties in Connecticut (**Fig. 1b**), providing us with better estimates. We assumed that the 7-day rolling average of B.1.1.7 and B.1.526^*^ in our dataset was representative of the true prevalence of these variants in the population because these datasets were compiled using genomes collected from the same sources. Therefore, we assumed that any biases introduced through subsampling would be systematic across all lineages. However, we also calculated a Jeffreys interval for daily variant frequencies and used the 0.025 and 0.975 quantiles to compute R_t_ and improve the robustness of our analysis (**Supplementary Fig. 2**).

In New Haven County, our R_t_ estimates for B.1.1.7 and B.1.526^*^ followed similar decreasing trajectories as COVID-19 vaccination rates increased, though B.1.1.7 consistently had a higher R_t_ (**Fig 2b**). The R_t_ for both B.1.1.7 and B.1.526* were above 1 between March 4, when our estimates began, and the end of April, when fully vaccinated rates reached ∼25%. An R_t_ value above 1 indicates that on average an infected individual infects more than one additional person. The R_t_ estimates for ‘other’ lineages fell below 1 in early January. Notably, the R_t_ for B.1.526* decreased to below 1 about one week earlier than that for B.1.1.7 (**Fig. 2b**). To directly compare the transmissibility of B.1.1.7 and B.1.526^*^, we calculated the ratio of R_t_ for each lineage over time (**Fig. 2c**). Once our estimates stabilized at the end of March, the R_t_ of B.1.1.7 was consistently higher than that of B.1.526^*^ (range: 1.057-1.10). These findings indicate that B.1.1.7 was approximately 6-10% more transmissible than B.1.526^*^ when both variants circulated concurrently in the same population. The ratio of R_t_ estimates calculated using the lower and upper quantiles of our Jeffreys intervals also exhibited this pattern (**Supplementary Fig. 2**). We observed a similar relationship in New York City, though with larger fluctuations (range: 1.02-1.29). The consistency of these findings suggests that B.1.1.7 was more transmissible than B.1.526^*^even when B.1.526^*^ emerged before B.1.1.7.

### Association of B.1.1.7 introductions with larger phylogenetic clusters than B.1.526* introductions

We next considered the possibility that the apparent increased transmissibility of B.1.1.7 compared to B.1.526^*^ was due to the number and timing of the introductions of each variant into Connecticut. More frequent introductions of B.1.1.7 could artificially inflate our R_t_ estimates (**Fig. 2**). To assess this possibility, we used a Bayesian phylogeographic method to quantify the number, timing, and source of observed introductions of both variants into Connecticut (**Fig. 3**). We found that while B.1.1.7 was not introduced more often into the state than B.1.526^*^, rather the clusters resulting from each introduction were on average larger than those produced by B.1.526^*^ introductions. These observations are in agreement with our epidemiological findings and further support the likelihood that B.1.1.7 spread more rapidly than B.1.526^*^.

**Figure 3:**
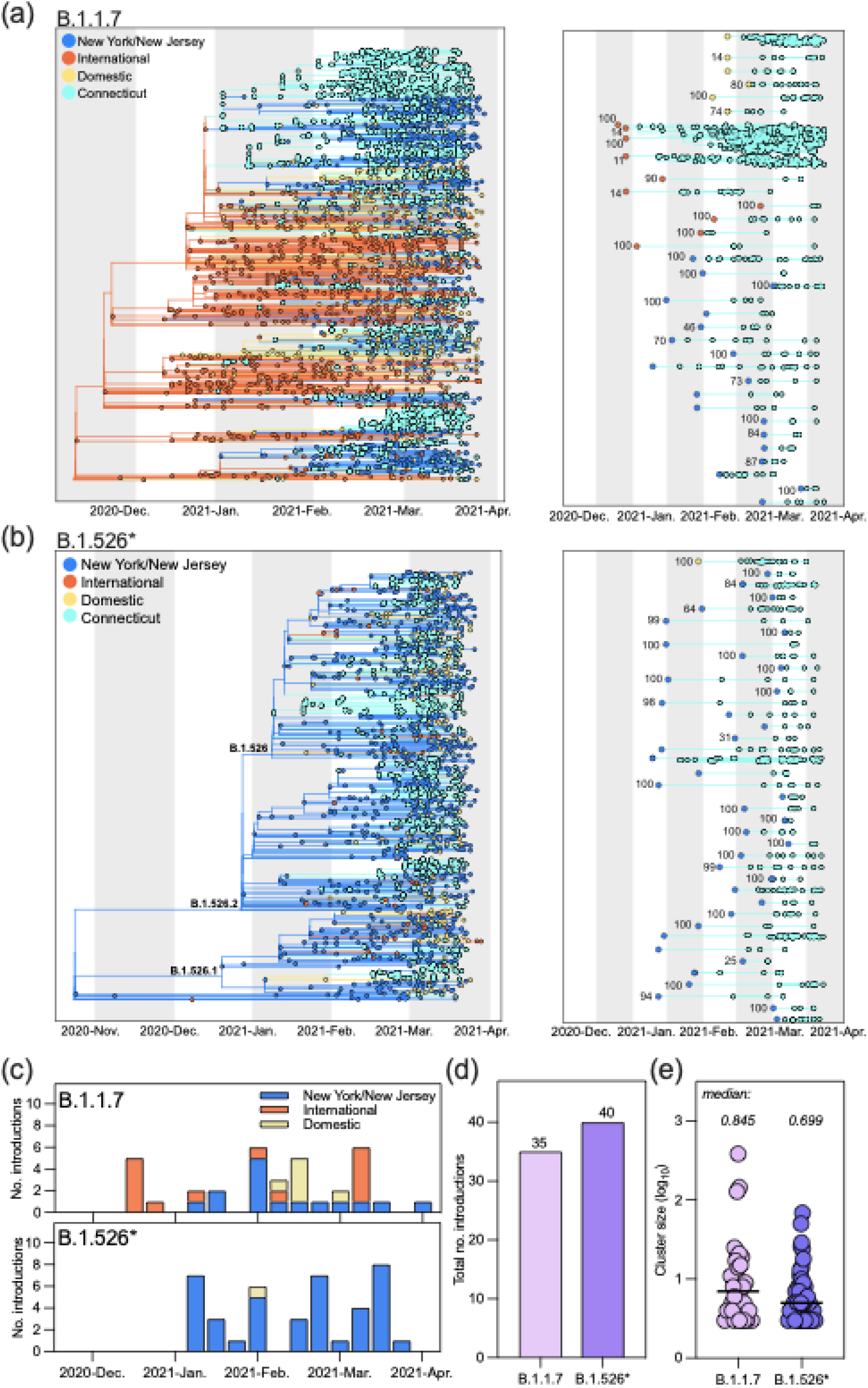
B.1.1.7 was introduced into Connecticut at a similar frequency as B.1.526^*^ but was associated with larger cluster sizes. (**a, b**) Discrete phylogeography of B.1.1.7 (*a*) and B.1.526^*^ (*b*). Tips and nodes were assigned one of four possible locations: Connecticut, New York/New Jersey, domestic, and international. The phylogeographic analysis was performed in BEAST^21^ using a time-resolved tree as the fixed topology ^22^. Bootstrap values for each clade are shown at each ancestral node (*right*) and were obtained by constructing individual maximum likelihood trees with 1000 ultrafast bootstraps in IQTree^24^. Clades without a support value were part of polytomies. (**c**) We summed the number of sustained introductions for each variant by week. We defined sustained introductions as Connecticut-only clades containing at least 3 tips related by a non-Connecticut ancestor with at least 0.7 posterior probability for the inferred location. Bar colors indicate the source of introduction. (**d**) There were more sustained introductions of B.1.526^*^ than B.1.1.7 into Connecticut. (**e**) The size of B.1.1.7 clades in Connecticut was on average larger than B.1.526^*^ clades in Connecticut. We calculated the log_10_ size of Connecticut clades shown in (a) and (b). The horizontal line denotes the median log cluster size.

To begin our phylogeographic analysis, we combined our SARS-CoV-2 genomic data with randomly sampled publicly available B.1.1.7 and B.1.526* genomes (gisaid.org) from outside of Connecticut, New York, and New Jersey, normalizing by reported deaths per location (see *Methods*). We did this independently 5 times for each variant to account for any potential biases from the subsampling process (∼17,000 and ∼13,000 genomes for each B.1.1.7 and B.1.526^*^ subsample, respectively; **Supplementary Table 2**), and constructed the ten, corresponding time-resolved trees with TreeTime^20^. We next performed discrete phylogeographic reconstruction over the time-resolved trees in BEAST^21,22^. We inferred the ancestral geographic states according to four discrete geographic categories: Connecticut, New York/New Jersey, domestic, and international. We chose to combine New York and New Jersey into one region because of the large volume of commuters and visitors who travel from northern New Jersey to New York City.

Due to the notably different geographic distribution of these two variants (**Fig. 3a,b**), we expected the source of introductions for each to also differ. Because B.1.526^*^ was first identified in New York and the majority of genomes from this variant family were sequenced in New York, we hypothesized New York would be the main source of B.1.526^*^ introductions into Connecticut. B.1.1.7 spread widely in the United Kingdom and Europe, and the first case of B.1.1.7 in Connecticut was associated with international travel^23^, indicating that early introduction would likely come from international sources. We anticipated that international sources would drive the initial introductions of B.1.1.7 into Connecticut until this variant was established in the US.

We found that the sources and size of B.1.1.7 and B.1.526^*^ sustained introductions differed between variants and throughout the study period (**Fig. 3c-e**). We defined a sustained introduction as a transition from a location outside of Connecticut into Connecticut in which (***1***) the resulting clade contained at least 3 tips and (***2***) the posterior probability of the ancestral, outside-Connecticut node was at least 0.7. As we expected, New York/New Jersey was a main source of introductions of B.1.526^*^ into Connecticut, accounting for all but one of the 40 independent introductions (**Fig. 3c-e**). The sources of introductions of B.1.1.7 were heterogeneous, including international and domestic sources throughout the study period (**Fig. 3c-e**). The relatively limited role of New York/New Jersey in the spread of B.1.1.7 into Connecticut may be due to the lower prevalence of this variant in New York City for the majority of our study period (**Fig. 1c**). Our phylogeographic analysis also revealed that although B.1.1.7 was introduced slightly less frequently than B.1.526* (**Fig. 3d**), B.1.1.7 introductions led to larger clusters (**Fig. 3e**). These patterns were consistent across all five phylodynamic replicates and with our estimates of R_t_ suggesting that B.1.1.7 has a fitness advantage over B.1.526* (**Supplementary Fig. 3, Fig. 2c**).

## Discussion

In this study, we developed and implemented a novel framework that combines epidemiological and genomic data to quantify the relative fitness of SARS-CoV-2 variants of public health significance. This framework, which measures changes in frequencies and estimates the effective reproduction number for individual variants, can be used to monitor the epidemiological dynamics of variants. Our approach is more informative than the current practice of tracking variant prevalence because it accounts for both the change in lineage frequencies and the number of incident cases. We demonstrated the utility of our framework by applying it to the dynamics of B.1.1.7 and B.1.526^*^ in Connecticut. In doing so, we found that while both B.1.1.7 and B.1.526^*^ likely had a fitness advantage over other lineages (**Fig. 1**), B.1.1.7 was 6-10% more transmissible than B.1.526^*^ (**Fig. 2**). Notably, the transmissibility of B.1.1.7 decreased at a slower rate than that of B.1.526^*^ as COVID-19 vaccination rates increased (**Fig. 2**). These conclusions were consistent with those of our phylogeographic analysis (**Fig. 3**), the typical albeit more computationally-intensive method for evaluating the dynamics of virus transmission and spread.

Our framework facilitates the continual monitoring of SARS-CoV-2 variant epidemiology across different settings, which is critical for informing public health policy decisions that must be made with readily available data. We also used our framework to analyze the dynamics of B.1.1.7 and B.1.526^*^ in New York City, where both variants also co-circulated. Our results were consistent with those from Connecticut (**Fig. 2**) but had some discrepancies with previous reports of relative growth rates in New York City. Specifically, West *et al*. reported that B.1.526 with the spike E484K substitution was growing at a faster rate than B.1.1.7 in New York City between December 2020 and March 2021^12^. We also observed this rapid rise in B.1.526^*^ prevalence during that time period (**Fig. 1d**); however, we found that the growth rate of B.1.526^*^ slowed shortly thereafter (**Fig. 1d**), and B.1.1.7 became the dominant circulating lineage in April (**Fig. 1c**). Moreover, we estimated that the effective reproduction number of B.1.1.7 was equal to or greater than that of B.1.526^*^ by February (**Fig. 2e**), an early indicator of the eventual rise in B.1.1.7 prevalence a few months later. This second finding is particularly crucial because it illustrates that while variant frequencies at specific time points may not be indicative of variant fitness, the changes in these frequencies can reveal relative variant transmission dynamics when combined with daily incidence data. Therefore, our framework can be used to accurately compare the fitness of competing variants soon after they emerge.

The epidemiological findings from our case study also have broader public health implications as new SARS-CoV-2 variants continue to emerge worldwide. The sources of introductions of novel variants reflect their global distribution (**Fig. 3a-c**), which will likely change over time. This heterogeneity poses a serious obstacle to control and prevention efforts because it limits the efficacy of policies that target specific points of entry. For variants that are prevalent on multiple continents like B.1.1.7 and, more recently, B.1.617.2 (‘delta’), testing, contact tracing, and vaccination campaigns within communities will likely prove more efficient in limiting their spread than targeting a specific subset of travelers. Once local transmission of a new variant has been established, assessing the public health threat is both challenging and necessarily retrospective. However, a robust genomic surveillance infrastructure coupled with the application of our framework would enable the monitoring of variant epidemiology in close to real time. The phylodynamic methods we applied in this study can be run on a desktop computer in a few hours, making the collation of representative datasets the rate limiting step. Expanding genomic surveillance efforts would remove this barrier and promote a rapid and efficient response to outbreaks caused by new variants.

There were some limitations to the epidemiological findings we have presented. First, we were not able to directly measure the secondary attack rates of individuals infected with B.1.1.7 or one of the B.1.526^*^ sublineages. Collecting this information requires extensive contact tracing and sequencing of all secondary infections that are not available in Connecticut. Instead, we assumed that biases introduced by the method we employed in this study would be systematic across SARS-CoV-2 lineages so that estimates of the relative transmissibility of B.1.1.7 and B.1.526^*^ would be unaffected. Second, we used a small subset of publicly available SARS-CoV-2 genomes for our phylodynamic analyses to make them computationally tractable. Incorporating a small proportion of available data into our analyses may have introduced biases, but by demonstrating the reproducibility of our findings with independent replicates (**Supplemental Fig. 3**), we substantially mitigated this issue. Finally, the scope of our study was limited to Connecticut and, in some cases, New York City, which may impinge upon the generalizability of our findings. However, our objective was to directly compare the fitness of B.1.1.7 and B.1.526^*^, and Connecticut is one of few locations with a robust genomic surveillance infrastructure where these variants emerged concurrently.

Here, we present a framework that uses genomic data to estimate the effective reproduction number of individual virus lineages as a measure of relative transmission and fitness. In applying our framework to Connecticut, this study is the first to directly compare the fitness of B.1.1.7 and B.1.526* in a setting where they emerged concurrently. Moreover, our findings highlight that many factors influence a variant’s success including the timing of introduction, the existing virus population, host immunity, and advantageous amino acid substitutions. As new SARS-CoV-2 variants emerge, it will be critical to assess the magnitude of the role that each of these elements play in precipitating local outbreaks so that appropriate, effective, and immediate steps may be taken to control further SARS-CoV-2 transmission.

## Methods

### Ethics

#### Yale University

The Institutional Review Board from the Yale University Human Research Protection Program determined that the RT-qPCR testing and sequencing of de-identified remnant COVID-19 clinical samples obtained from clinical partners conducted in this study is not research involving human subjects (IRB Protocol ID: 2000028599).

#### Jackson Laboratory

The Institutional Review Board of The Jackson Laboratory determined that use of de-identified residual COVID-19 clinical samples obtained from the Clinical Genomics Laboratory for RT-qPCR testing and sequencing for this study is not research involving human subjects (IRB Determination: 2020-NHSR-021).

#### New York State Department of Health, Wadsworth Center

Residual portions of respiratory specimens from individuals who tested positive for SARS-CoV-2 by RT-PCR were obtained from the Wadsworth Center and partnering clinical laboratories. This work was approved by the New York State Department of Health Institutional Review Board, under study numbers 02-054 and 07-022.

### Reported COVID-19 case data

We used daily reported cases compiled by the Johns Hopkins COVID-19 portal (https://github.com/CSSEGISandData/COVID-19). We summed the number of incident cases by week by state for Massachusetts, New York, Rhode Island, New Jersey, and Connecticut, and we aggregated incident cases by week by county for New Haven, Fairfield, and Westchester. We visualized these data using Prism v.9.0.2 (plots) and Rv.1.2 (maps). For the latter, we obtained the shapefiles from the United States Census Bureau (east coast) and the Connecticut Department of Energy & Environmental Protection (DEEP) Geographic Information Systems Open Data Website (Connecticut).

### SARS-CoV-2 sequencing and consensus generation

#### Yale University

We received clinical samples from confirmed SARS-CoV-2 positive individuals from routine testing provided by Yale New Haven Hospital, Yale Pathology Laboratory, “Yale Campus Study”, Connecticut Department of Public Health, and Murphy Medical Associates. These samples were sent as either nasal swabs in viral transport media, raw saliva, or extracted and purified RNA. For the former two, we extracted RNA from 300µL of the original sample using the MagMAX viral/pathogen nucleic acid isolation kit, eluting in 75µl of elution buffer. We tested the extracted nucleic acid using our ‘variant of concern’ RT-qPCR assay to determine the SARS-CoV-2 viral RNA load ^25^. Samples with cycle thresholds <35 were prepared for sequencing using the Illumina COVIDSeq Test RUO version to synthesize cDNA, and generate and tagment amplicons. Amplicons were pooled and cleaned before quantification with Qubit High Sensitivity dsDNA kit. The resulting libraries were sequenced using a 2×100 or 2×150 approach on an Illumina NovaSeq at the Yale Center for Genomic Analysis. Each sample was given at least 1 million reads. Samples were typically processed in sets of 94 with negative controls incorporated during the RNA extraction, cDNA synthesis, and amplicon generation steps.

Using BWA-MEM v.0.7.15^26^, we aligned reads to the Wuhan-Hu-1 reference genomes (GenBank MN908937.3). With iVar v1.2.1^27^ and SAMtools^28^, we trimmed sequencing adaptors, masked primer sequences, and called bases by simple majority (>50% frequency) at each site to generate consensus genomes. An ambiguous ‘N’ was used when fewer than 10 reads were present at a site. In all cases, negative controls were analyzed and confirmed to consist of at least 95% Ns. We used pangolin v.2.4.2^29^ to assign lineages^19^.

#### Jackson Laboratory

Clinical samples were received in The Jackson Laboratory Clinical Genomics Laboratory (CGL) as part of a statewide COVID-19 surveillance program, with the majority of samples representing asymptomatic screening of nursing home and assisted living facility residents and staff. Total nucleic acids were extracted from anterior nares swabs in viral transport media or saline (200µL) using the MagMAX Viral RNA Isolation kit (ThermoFisher) on a KingFisher Flex purification system. Samples were tested for the presence of SARS-CoV-2 RNA using the TaqPath COVID-19 Combo Kit (ThermoFisher). Samples with cycle thresholds ≤30 for the N gene target were prepared for sequencing using the Illumina COVIDSeq Test kit. Sequencing was performed on an Illumina NovaSeq or NextSeq in the CGL. Data analysis was performed using the DRAGEN COVID Lineage App in BaseSpace Sequence Hub. Sequences with >80% of bases with non-N basecalls and ≥1500-fold median coverage were considered successful and were submitted to GISAID. Lineages were assigned using pangolin v.2.4.2^29^and the most current version of the pangoLEARN assignment algorithm.

#### New York State Department of Health, Wadsworth Center

Respiratory swabs positive for SARS-CoV-2 were sent to the Wadsworth Center from collaborating clinical laboratories across New York State as part of an enhanced genomic surveillance program initiated by the New York State Department of Health in December 2020. Specimens were required to have a real-time cycle threshold value less than 30. Nucleic acid extraction was performed on a Roche MagNAPure 96 (Roche, Indianapolis, IN) and RNA was processed for whole genome sequencing with a modified ARTIC3 protocol (http://artic.network/ncov-2019) in the Applied Genomics Technology Core at the Wadsworth Center, as previously described ^10^. Lineage was determined by GISAID using pangolin software ^29^, updated June 7, 2021. Daily relative frequency of variants within New York City was determined based on sample collection date and patient residence within Bronx, Kings, New York, Queens, or Richmond counties. Any specimens that were sequenced as a result of pre-screening for specific mutations or clinical/epidemiological criteria were removed from the analysis.

### Percent of COVID-19 cases sequenced

To calculate the percent of cases sequenced in each county, we tabulated the number of genomes collected from the state with available county-level data. Though this level of geographic resolution was only available for genomes sequenced by our laboratory and the Jackson Laboratory for Genomic Medicine, these two sources have generated the vast majority of genomes for the state of Connecticut. For New York City, NY, we used genomes generated by the Wadsworth Center. Using the case data described above, we summed the number of cases reported by each county between November 30, 2020 and May 9, 2021, and divided the total number of genomes generated for each county within the same timeframe by that sum.

### Frequency of SARS-CoV-2 variants among sequenced cases

To assess the frequency of circulating lineages, we selected genomes that were sequenced through a non-biased sampling approach. Specifically, we excluded genomes that were screened and sequenced through a targeted S-gene target failure surveillance system. As with the dataset we used to measure the percent of cases sequenced by county, these genomes were generated by our laboratory, Jackson Laboratory, and the Wadsworth Center. We organized these genomes into three categories using Pangolin v.2.4.2^29^: B.1.1.7, B.1.1526^*^, and ‘other’. We then tabulated the number of genomes in each category by week and calculated the percent of the total number of genomes for that week.

### Distribution of SARS-CoV-2 variants among cases

We obtained estimates of the distribution of cases attributed to each lineage category by multiplying the frequency of that category by the number of cases reported in the same week. In doing so, we assumed that the sequencing frequencies described above were representative of the virus population circulating in New Haven and Fairfield counties, and New York City (all counties). We also assumed that the number of reported cases for each county was representative of the true number of infections in that region.

To account for any uncertainty in our assumption that the sampling frequencies were representative of cases per county, we began by calculating p, a 7-day rolling average for the proportion of sequenced cases for each lineage category. This produced daily proportion estimates. To further account for any uncertainty, for each p, we calculated a Jeffreys interval, which is a Bayesian, equal-tailed interval of the form^30^:

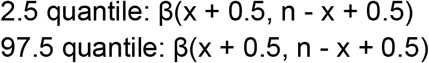

where β represents the beta distribution, x represents the 7-day rolling average of sequences of a specific lineage, and n represents the 7-day rolling average of sequences for all lineages. Our measure of interest, p, is calculated by x/n. The Jeffreys intervals were calculated using the package “DescTools” in R v4.0.1.

### Logistic regression

We computed logistic growth models for each lineage category in each county using the frequency estimates described above.

### Effective reproduction number

Using p, and the 0.025 and 0.975 quantiles from the Jeffreys interval, we multiplied these values by the number of reported cases per day. These three potential case counts were used to calculate the reproduction number (R_t_), the mean number of secondary cases generated by a typical primary case at time t in a population, curves per lineage (**Supplementary Fig. 2**). Because there is no consensus in the literature as to the precise serial interval for each variant, we used an uncertain serial interval with mean of 5.2 days and standard deviation of 4 days^31–33^. Through the uncertain serial interval, multiple distributions were explored where the mean was allowed to vary from 2.2 to 8.2 days, and the standard deviation varied from 2.5 to 5.5 days. From each of these R_t_ distributions, we selected the mean R_t_ to represent a given lineage’s instantaneous effective reproductive number per day. Further, for the R_t_ distribution calculated from p, we also computed the 0.025 and 0.975 quantiles (**Supplementary Fig. 2**). All of the R_t_ estimates were calculated using the “EpiEstim” package in R v4.0.1. Finally, we used a smoothing spline to smooth the daily R_t_ curves with the package “stat” in R v4.0.1.

### COVID-19 vaccination rates

We obtained vaccination data for New York City from data.cdc.gov and for Connecticut from data.ct.gov (COVID-19 Vaccinations by Town and Age Group).

### SARS-CoV-2 genome selection for phylogenetic analysis

We downsampled both B.1.1.7 and B.1.526* datasets using COVID-19 death counts. We elected to normalize genome counts to the number of deaths because deaths are less likely to be under-reported than cases^34^. We obtained daily death counts for all countries and US states from the Johns Hopkins COVID-19 portal (https://github.com/CSSEGISandData/COVID-19). We summed the cumulative number of deaths for each state or country between October 1, 2020 and April 23, 2021 because we assumed that deaths could not be attributed to either variant before October 1. Because country-level death data were not reported for countries within the UK, we calculated the total number of genomes to sample from the UK with the method described above, and then calculated the distribution of genomes by UK country based on population size. To calculate the number of genomes we would include in our final datasets from each region, we first calculated the ratio of variant genomes to deaths. In doing so, we assumed that the number of variant genomes sequenced by each country or state was proportional to the total number of genomes sequenced in those places.

For non-US sampling, if the number of variant genomes sequenced comprised less than 1% of cumulative deaths, we included all of the genomes from that location. Otherwise, we selected the number of genomes that corresponded to 1% of reported cumulative deaths. We used a similar approach for US states except we set the minimum threshold to 0.1% of cumulative deaths. In all cases, if a country or state had less than 20 genomes available, we included all of them. For the B.1.526^*^ lineages, we calculated the proportion of each lineage out of the total number of B.1.526^*^ genomes sequenced in each country or state and selected genomes according to this proportion.

We did not downsample Connecticut, New York, or New Jersey for either variant dataset in the first stage of downsampling. Once functional duplicates were removed from these locations, we included 1,408 genomes sequenced by Yale (Connecticut), 497 sequenced by Jackson Laboratory (Connecticut), and 803 sequenced by the Wadsworth Center (New York City) collected between December 1, 2020 and April 23, 2021. We obtained all other genomes for this analysis from GISAID (gisaid.org) (**Supplementary Table 2, 3**). We also applied a modified sampling scheme for B.1.1.7 genomes from Australia, New Zealand, Sint Maarten, Bonaire, Vietnam, or Singapore because these locations reported a negligible number of deaths. For this reason, it was impossible to downsample based on the number of reported deaths. We therefore randomly selected 1% of available genomes from those locations instead. To select the genomes to incorporate into our dataset from the downsampled locations, we randomly selected a weekly set of genomes equal to 1% of deaths per week. Using this workflow, we generated five datasets for each variant to serve as independent replicates for the remainder of our analysis (**Table 1**). In all cases, we excluded genomes containing more than 30% Ns from our selection. Due to the broader global distribution of B.1.1.7, the datasets for this variant were necessarily larger than those for B.1.526^*^.

At that stage, the datasets were still too large to be computationally tractable. We next scaled each dataset by a factor of 0.1 by randomly selecting 10% of genomes by country or state (US only). We did not scale genomes from Connecticut so that the final datasets were not precisely one tenth the size of the original (**Supplementary Table 2**).

### SARS-CoV-2 phylogenetic analysis

#### Sequence alignment and refinement

Having compiled our ten datasets, we aligned the genomes using MAFFT^35^. We then removed gaps and masked problematic sites^36^. We then removed functional duplicates from each dataset to reduce phylogenetic redundancy. We defined a functional duplicate as genomes that shared identical sequences, week of collection, and geographic region. For genomes collected in Connecticut and New York, we defined the geographic region as the county. For genomes collected elsewhere in the US, we defined it as ‘state’. For genomes collected internationally, we defined the geographic region as ‘country’.

#### Maximum likelihood construction

To identify and remove problematic genomes from our datasets, we performed a preliminary phylogenetic analysis in IQTree^24^. Each tree was rooted using a P.1 genome (hCoV-19/Brazi/AM-FIOCRUZ-20842882CA/2020). We performed a root-to-tip analysis in TempEST^37^ and removed outliers with residuals > |0.0015|. We constructed a maximum likelihood tree with each dataset (n = 10) using a GTR substitution model with 1000 ultrafast bootstraps again with IQTree.

#### Time-resolved construction

To avoid computational bottlenecks in our phylogeographic reconstruction, we did not use a Bayesian method to infer the temporal resolution of our maximum likelihood tree. We have previously shown that temporal estimates inferred using TreeTime agree with those inferred from BEAST for B.1.1.7^10^, and we assumed this would also be the case for B.1.526^*^. We used the bootstrapped trees and associated alignments to construct corresponding time-resolved phylogenetic trees with TreeTime v.0.8.0^20^. This method is implemented in an augur pipeline^38^.

#### Discrete phylogeographic analysis

We performed a discrete phylogeographic analysis with the time-resolved trees as the fixed topology using BEAST^21,22^. Specifically, we assigned a location to each of the tree tips from four categories: ‘Connecticut’, ‘New York/New Jersey’, ‘domestic’, and ‘international’. We used an asymmetric substitution model and a strict clock to model location. We ran each tree for 1 million chains and used Tracer v.1.7.1 to confirm that all parameters had achieved ESS values of at least 200.

We identified Connecticut-only clades and their source of introduction using the “exploded tree” script implemented with baltic 0.1.6 (https://github.com/evogytis/baltic). We restricted our subsequent analysis to clades that represented sustained introductions, or clades that were composed of at least 3 tips that were related by a non-Connecticut ancestor with at least 0.7 posterior probability for the inferred location. We aggregated the number of sustained introductions by week and source, and visualized the results using Prism v.9.0.2. We merged the bootstrap values from our original trees with the topology of our geographically-resolved trees using baltic.

## Data Availability

All of the genomic data used for the analyses in this manuscript are available on GISAID (gisaid.org). We gratefully acknowledge all of the laboratories that obtained the clinical specimen and generated the SARS-CoV-2 genomes used in our analyses (acknowledgment tables included in supplement). All files associated with our phylogenetic analysis may be found in our Github repository (https://github.com/grubaughlab/paper_2021_B117vsB1526).

## Acknowledgements

We thank the frontline and essential workers for their service during the pandemic, the groups that continuously make their data available to the public, K. Gangavarapu for technical advice, C. O’Connor for map creation, and our friends and family - particularly V. Parsons, P. Jack, S. Currie, and S. Taylor - for their support. This work was funded by CTSA Grant Number TL1 TR001864 (M.E.P. and T.A.), Fast Grant from Emergent Ventures at the Mercatus Center at George Mason University (N.D.G.) and CDC Contract # 75D30120C09570 (N.D.G.). Initial funding for sequencing at the Wadsworth Center was provided by the New York Community Trust.

## Author contributions

Conceptualization, M.E.P, J.E.R., B.P.T., W.P.H., L.M.G, V.E.P., N.D.G.; Sample acquisition and diagnostic testing, K.K., G.O., N.R., R.E., S.M., C.N., E.L., A.M., R.D., J.R., L.N., M.S.W., M.L.A., J.W., C.L., P.H., M.L.L., D.R.P., M.D.A.; Sequencing and data processing, M.E.P., M.I.B., I.M.O., A.R., E.L-N., K.K., G.O., N.R., A.E.W., C.C.K., T.A., A.F.B., R.E., I.R.T., C.C., J.P.K., M.S., J.P., E.S., J.R.F.; Data analysis and interpretation, M.E.P., J.E.R., A.R., L.M.G., V.E.P., N.D.G.; Supervision, S.M., K.B., J.R.F., C.B.F.V., L.M.G., V.E.P., K.S.G., M.D.A., N.D.G.; Writing - Original Draft, M.E.P., N.D.G.; Writing - Review & Editing, J.E.R., A.R., C.B.F.V., M.D.A.

## Data availability

All of the genomic data used for the analyses in this manuscript are available on GISAID (gisaid.org). We gratefully acknowledge all of the laboratories that obtained the clinical specimen and generated the SARS-CoV-2 genomes used in our analyses (**Supplementary Table 2,3**). All files associated with our phylogenetic analysis may be found in our Github repository (https://github.com/grubaughlab/paper_2021_B117vsB1526).

