## Supplementary Table 3 for "Combining genomic and epidemiological data to compare the transmissibility of SARS-CoV-2 lineages"

We gratefully acknowledge the following Authors from the Originating laboratories responsible for obtaining the specimens, as well as the Submitting laboratories where the genome data were generated and shared via GISAID, on which this research is based.

All Submitters of data may be contacted directly via www.gisaid.org

Authors are sorted alphabetically.

| Accession ID | Originating Laboratory | Submitting Laboratory | Authors |
| --- | --- | --- | --- |
| EPI_ISL_1009654 | DPHL | Delaware Public Health Lab | Gregory Hovan |
| EPI_ISL_1015696, EPI_ISL_1015699, EPI_ISL_1015702, EPI_ISL_1015703 | Columbia University Irving Medical Center | Wadsworth Center, New York State Department of Health | Kirsten St. George, Daryl M. Lamson, Alexis Russel, Matthew Shudt, Melissa A Leisner, Jonathan Plitnick, Navjot Singh, John Kelly, Erasmus Schneider, Erica Lasek-Nesselquist |
| EPI_ISL_1016177, EPI_ISL_1016182, EPI_ISL_1016184, EPI_ISL_1016199, EPI_ISL_1016211, EPI_ISL_1016213 | MONTEFIORE MEDICAL CENTER LABORATORIES | Wadsworth Center, New York State Department of Health | Kirsten St. George, Daryl M. Lamson, Alexis Russel, Matthew Shudt, Melissa A Leisner, Jonathan Plitnick, Navjot Singh, John Kelly, Erasmus Schneider, Erica Lasek-Nesselquist |
| EPI_ISL_1016228, EPI_ISL_1016242, EPI_ISL_1016244, EPI_ISL_1016248 | New York Presbyterian Hospital | Wadsworth Center, New York State Department of Health | Kirsten St. George, Daryl M. Lamson, Alexis Russel, Matthew Shudt, Melissa A Leisner, Jonathan Plitnick, Navjot Singh, John Kelly, Erasmus Schneider, Erica Lasek-Nesselquist |
| EPI_ISL_1016260, EPI_ISL_1016268, EPI_ISL_1016331 | Columbia University Irving Medical Center | Wadsworth Center, New York State Department of Health | Kirsten St. George, Daryl M. Lamson, Alexis Russel, Matthew Shudt, Melissa A Leisner, Jonathan Plitnick, Navjot Singh, John Kelly, Erasmus Schneider, Erica Lasek-Nesselquist |
| EPI_ISL_1017982, EPI_ISL_1017985 | DOHMH Central Harlem | New York City Public Health Laboratory | Jade Wang, et al. |
| EPI_ISL_1017990 | Department of Homeless Services | New York City Public Health Laboratory | Jade Wang, et al. |
| EPI_ISL_1018003 | DOHMH PHL | New York City Public Health Laboratory | Jade Wang, et al. |
| EPI_ISL_1018012 | DOHMH Jamaica | New York City Public Health Laboratory | Jade Wang, et al. |
| EPI_ISL_1018034, EPI_ISL_1018037, EPI_ISL_1018047 | OCME Office Of Chief Medical Examiner | New York City Public Health Laboratory | Jade Wang, et al. |
| EPI_ISL_1020490, EPI_ISL_1020509, EPI_ISL_1020511, EPI_ISL_1020512, EPI_ISL_1020513 | Columbia University Irving Medical Center | Wadsworth Center, New York State Department of Health | Kirsten St. George, Daryl M. Lamson, Alexis Russel, Matthew Shudt, Melissa A Leisner, Jonathan Plitnick, Navjot Singh, John Kelly, Erasmus Schneider, Erica Lasek-Nesselquist |
| EPI_ISL_1020839, EPI_ISL_1020994, EPI_ISL_1037840 | Laboratory Corporation of America | Respiratory Viruses Branch, Division of Viral Diseases, Centers for Disease Control and Prevention | Peter W. Cook, Dakota Howard, Dhwani Batra, Ben L. Rambo-Martin, Clinton R. Paden, Suxiang Tong, Duncan MacCannell |
| EPI_ISL_1038995 | Murphy Medical Associates | Grubaugh Lab - Yale School of Public Health | Joseph Fauver, Tara Alpert, Anderson Brito, Mallery Breban, Anne Wyllie, Chantal Vogels, Mary Petrone, Annie Watkins, Chaney Kalinich, Isabel Ott, Nathan Grubaugh |
| EPI_ISL_1038997 | Fair Haven CHC | Grubaugh Lab - Yale School of Public Health | Joseph Fauver, Tara Alpert, Anderson Brito, Mallery Breban, Anne Wyllie, Chantal Vogels, Mary Petrone, Annie Watkins, Chaney Kalinich, Isabel Ott, Nathan Grubaugh |
| EPI_ISL_1038998 | Yale Pathology Lab | Grubaugh Lab - Yale School of Public Health | Joseph Fauver, Tara Alpert, Anderson Brito, Mallery Breban, Anne Wyllie, Chantal Vogels, Mary Petrone, Annie Watkins, Chaney Kalinich, Isabel Ott, Nathan Grubaugh |
| EPI_ISL_1040861, EPI_ISL_1040900 | NYU Langone Health | Departments of Pathology and Medicine, New York University School of Medicine | Adriana Heguy, Dacia Dimartino, Emily Guzman, Christian Marier, Peter Meyn, Sitharam Ramaswami, Gael Westby, Paul Zappile, Yutong Zhang, Paolo Cotzia, Guiqing Wang |
| EPI_ISL_1041215, EPI_ISL_1041223, EPI_ISL_1041238, EPI_ISL_1041241, EPI_ISL_1041242, EPI_ISL_1041262, EPI_ISL_1041267, EPI_ISL_1041269, EPI_ISL_1041274, EPI_ISL_1041284, EPI_ISL_1041289, EPI_ISL_1041300, EPI_ISL_1041308, EPI_ISL_1041333, EPI_ISL_1041364, EPI_ISL_1041374, EPI_ISL_1041380, EPI_ISL_1041386, EPI_ISL_1041387, EPI_ISL_1041415, EPI_ISL_1041416, EPI_ISL_1041421, EPI_ISL_1041443, EPI_ISL_1041457, EPI_ISL_1041467, EPI_ISL_1041470, EPI_ISL_1041475, EPI_ISL_1041476, EPI_ISL_1041488, EPI_ISL_1041495, EPI_ISL_1041502, EPI_ISL_1041504, EPI_ISL_1041505, EPI_ISL_1041508, EPI_ISL_1041511, EPI_ISL_1041521, EPI_ISL_1041548, EPI_ISL_1041558, EPI_ISL_1041561, EPI_ISL_1041569, EPI_ISL_1041580, EPI_ISL_1041582, EPI_ISL_1041583, EPI_ISL_1041585, EPI_ISL_1041594, EPI_ISL_1041595, EPI_ISL_1041596, EPI_ISL_1041604, EPI_ISL_1041606, EPI_ISL_1041612, EPI_ISL_1041635, EPI_ISL_1041650, EPI_ISL_1041652, EPI_ISL_1041657, EPI_ISL_1041661, EPI_ISL_1041668, EPI_ISL_1041673, EPI_ISL_1041695, EPI_ISL_1041701, EPI_ISL_1041708, EPI_ISL_1041710, EPI_ISL_1041731, EPI_ISL_1041733, EPI_ISL_1041744, EPI_ISL_1041745, EPI_ISL_1041747, EPI_ISL_1041750, EPI_ISL_1041760, EPI_ISL_1041763, EPI_ISL_1041765, EPI_ISL_1041807, EPI_ISL_1041815, EPI_ISL_1041822, EPI_ISL_1041829, EPI_ISL_1041830, EPI_ISL_1041838, EPI_ISL_1041858, EPI_ISL_1041861, EPI_ISL_1041874, EPI_ISL_1041878, EPI_ISL_1041884, EPI_ISL_1041885, EPI_ISL_1041886, EPI_ISL_1041887, EPI_ISL_1041897, EPI_ISL_1041899, EPI_ISL_1041921, EPI_ISL_1041927, EPI_ISL_1041928, EPI_ISL_1041930 | | | |
| see above | Pandemic Response Lab - NYC | Pandemic Response Lab, R&D | Henry Lee, Michael Hammerling, Melissa Hopkins, Cybill del Castillo, William Ward, Pradeep Bugga, Haiping Hao, Jon Laurent |
| EPI_ISL_1046773 | SARS-CoV-2 testing team, National Institute of Infectious Diseases | Pathogen Genomics Center, National Institute of Infectious Diseases | Tsuyoshi Sekizuka, Kentaro Itokawa, Rina Tanaka, Masanori Hashino, Tadashi Kikuchi, Lucky Ronald Runtuwene, Shigeru Kusagawa, Makoto Kuroda |
| EPI_ISL_1050039 | NYU Langone Health | Departments of Pathology and Medicine, New York University School of Medicine | Adriana Heguy, Dacia Dimartino, Emily Guzman, Christian Marier, Peter Meyn, Sitharam Ramaswami, Gael Westby, Paul Zappile, Yutong Zhang, Paolo Cotzia, Guiqing Wang |
| EPI_ISL_1059101 | University of Michigan Clinical Microbiology Laboratory | Lauring Lab, University of Michigan, Department of Microbiology and Immunology | Valesano |
| EPI_ISL_1060772, EPI_ISL_1060780, EPI_ISL_1060782, EPI_ISL_1060784, EPI_ISL_1060785, EPI_ISL_1060788, EPI_ISL_1060800, EPI_ISL_1060801, EPI_ISL_1060802, EPI_ISL_1060804 | NJ Rapid Response Teams | New Jersey Public Health Environmental Laboratories (NJ_PHEL) | Lindsey Bodnar, Shiv Verma, Dana Woell, Byeong Jeong |
| EPI_ISL_1064764, EPI_ISL_1064770 | OCME Office Of Chief Medical Examiner | New York City Public Health Laboratory | Jade Wang, et al. |
| EPI_ISL_1064777 | DOHMH Crown Heights | New York City Public Health Laboratory | Jade Wang, et al. |
| EPI_ISL_1064801 | DOHMH Morrisania | New York City Public Health Laboratory | Jade Wang, et al. |
| EPI_ISL_1064803 | DOHMH Central Harlem | New York City Public Health Laboratory | Jade Wang, et al. |
| EPI_ISL_1064804 | OCME Office Of Chief Medical Examiner | New York City Public Health Laboratory | Jade Wang, et al. |
| EPI_ISL_1064807 | DOHMH Jamaica | New York City Public Health Laboratory | Jade Wang, et al. |
| EPI_ISL_1064815, EPI_ISL_1064817 | OCME Office Of Chief Medical Examiner | New York City Public Health Laboratory | Jade Wang, et al. |
| EPI_ISL_1064823 | DOHMH Riverside | New York City Public Health Laboratory | Jade Wang, et al. |
| EPI_ISL_1064824 | DOHMH Crown Heights | New York City Public Health Laboratory | Jade Wang, et al. |
| EPI_ISL_1064826 | DOHMH PHL | New York City Public Health Laboratory | Jade Wang, et al. |
| EPI_ISL_1064831 | DOHMH Central Harlem | New York City Public Health Laboratory | Jade Wang, et al. |
| EPI_ISL_1064836 | DOHMH Fort Greene | New York City Public Health Laboratory | Jade Wang, et al. |

| | | | |
|---|---|---|---|
| EPI_ISL_1064837 | DOHMH Jamaica | New York City Public Health Laboratory | Jade Wang, et al. |
| EPI_ISL_1064842 | DOHMH Morrisania | New York City Public Health Laboratory | Jade Wang, et al. |
| EPI_ISL_1064843, EPI_ISL_1064846 | OCME Office Of Chief Medical Examiner | New York City Public Health Laboratory | Jade Wang, et al. |
| EPI_ISL_1064848 | DOHMH Jamaica | New York City Public Health Laboratory | Jade Wang, et al. |
| EPI_ISL_1064849 | DOHMH Morrisania | New York City Public Health Laboratory | Jade Wang, et al. |
| EPI_ISL_1064853, EPI_ISL_1064854, EPI_ISL_1064855 | OCME Office Of Chief Medical Examiner | New York City Public Health Laboratory | Jade Wang, et al. |
| EPI_ISL_1064858 | DOHMH Jamaica | New York City Public Health Laboratory | Jade Wang, et al. |
| EPI_ISL_1064863 | OCME Office Of Chief Medical Examiner | New York City Public Health Laboratory | Jade Wang, et al. |
| EPI_ISL_1076611, EPI_ISL_1079543, EPI_ISL_1079930 | Houston Methodist Hospital | Houston Methodist Hospital | S. Wesley Long, Randall J. Olsen, Paul A. Christensen, Sishir Subedi, Robert Olson, James J. Davis, Matthew Ojeda Saavedra, Prasanti Yerramilli, Layne Pruitt, Kristina Reppond, Madison N. Shyer, Jessica Cambric, Ilya J. Finkelstein, Jimmy Gollihar, and James M. Musser |
| EPI_ISL_1080350 | NJ Rapid Response Teams | NJ_PHEL | Lindsey Bodnar, Shiv Verma, Dana Woell, Byeong Jeong |
| EPI_ISL_1081297, EPI_ISL_1081298 | Wyoming Public Health Laboratory | Wyoming Public Health Laboratory | Jim Mildenberger, Wanda Manley, Noah Hull, Taylor Fearing, Lynette Gumbleton, Channing Weber, Ashley Norberg, Chayse Rowley, Marley Goetz, Brian Dominguez, Elliot Thomasson, Cari Sloma, and Rob Christensen |
| EPI_ISL_1087278, EPI_ISL_1087290, EPI_ISL_1087292, EPI_ISL_1087293, EPI_ISL_1087294, EPI_ISL_1087296, EPI_ISL_1087299 | NYU Langone Health | Departments of Pathology and Medicine, New York University School of Medicine | Adriana Heguy, Dacia Dimartino, Emily Guzman, Christian Marier, Peter Meyn, Sitharam Ramaswami, Gael Westby, Paul Zappile, Yutong Zhang, Paolo Cotzia, Guiqing Wang |
| EPI_ISL_1091224 | Croatian Institute of Public Health | Croatian Institute of Public Health | Irena Tabain, Ivana Ferenak |
| EPI_ISL_1092375, EPI_ISL_1092979, EPI_ISL_1092980 | National Virus Reference Laboratory | National Virus Reference Laboratory | Zoe Yandle, Charlene Bennet, Gabriel Gonzalez, Michael Carr, Jonathan Dean, Cillian F De Gascun |
| EPI_ISL_1094370 | NYSDOH Wadsworth Center, Virology Lab | Respiratory Viruses Branch, Division of Viral Diseases, Centers for Disease Control and Prevention | Krista Queen, Yan Li, Ying Tao, Jing Zhang, Anna Uehara, Anna Montmayeur, Clinton R. Paden, Peter W. Cook, Rachel Marine, Mili Sheth, Jasmine Padilla, Sarah Nobles, Mark Burroughs, Lori Rowe, Haibin Wang, Ben L. Rambo-Martin, Dhwani Batra, Justin Lee, Suxiang Tong |
| EPI_ISL_1097791, EPI_ISL_1097795, EPI_ISL_1097803, EPI_ISL_1097823, EPI_ISL_1097826, EPI_ISL_1097831, EPI_ISL_1097845, EPI_ISL_1097846, EPI_ISL_1097859, EPI_ISL_1097864, EPI_ISL_1097871, EPI_ISL_1097879, EPI_ISL_1097887, EPI_ISL_1097896, EPI_ISL_1097901, EPI_ISL_1097907, EPI_ISL_1097908, EPI_ISL_1097918, EPI_ISL_1097922, EPI_ISL_1097925, EPI_ISL_1097930, EPI_ISL_1097936, EPI_ISL_1097953, EPI_ISL_1097958, EPI_ISL_1097965, EPI_ISL_1097976, EPI_ISL_1097979, EPI_ISL_1097994, EPI_ISL_1098005, EPI_ISL_1098017, EPI_ISL_1098018, EPI_ISL_1098020, EPI_ISL_1098026, EPI_ISL_1098029, EPI_ISL_1098043, EPI_ISL_1098051, EPI_ISL_1098055, EPI_ISL_1098057, EPI_ISL_1098070, EPI_ISL_1098074, EPI_ISL_1098077, EPI_ISL_1098081, EPI_ISL_1098085, EPI_ISL_1098100, EPI_ISL_1098101, EPI_ISL_1098105, EPI_ISL_1098108, EPI_ISL_1098115, EPI_ISL_1098137, EPI_ISL_1098138, EPI_ISL_1098140, EPI_ISL_1098149, EPI_ISL_1098161, EPI_ISL_1098163, EPI_ISL_1098176, EPI_ISL_1098181, EPI_ISL_1098186, EPI_ISL_1098191, EPI_ISL_1098195, EPI_ISL_1098204, EPI_ISL_1098235, EPI_ISL_1098246, EPI_ISL_1098254, EPI_ISL_1098267, EPI_ISL_1098271, EPI_ISL_1098275, EPI_ISL_1098281, EPI_ISL_1098301, EPI_ISL_1098303, EPI_ISL_1098310, EPI_ISL_1098311, EPI_ISL_1098312, EPI_ISL_1098326, EPI_ISL_1098327, EPI_ISL_1098332, EPI_ISL_1098339, EPI_ISL_1098343, EPI_ISL_1098348, EPI_ISL_1098358, EPI_ISL_1098378, EPI_ISL_1098383, EPI_ISL_1098403, EPI_ISL_1098404, EPI_ISL_1098421, EPI_ISL_1098425, EPI_ISL_1098428, EPI_ISL_1098429, EPI_ISL_1098432, EPI_ISL_1098433, EPI_ISL_1098437, EPI_ISL_1098438, EPI_ISL_1098448, EPI_ISL_1098455, EPI_ISL_1098467, EPI_ISL_1098469, EPI_ISL_1098473, EPI_ISL_1098476, EPI_ISL_1098489, EPI_ISL_1098493, EPI_ISL_1098504, EPI_ISL_1098510, EPI_ISL_1098514, EPI_ISL_1098519, EPI_ISL_1098525, EPI_ISL_1098526, EPI_ISL_1098536, EPI_ISL_1098542, EPI_ISL_1098552, EPI_ISL_1098555, EPI_ISL_1098562, EPI_ISL_1098564, EPI_ISL_1098576, EPI_ISL_1098586 | | | |
| see above | Pandemic Response Lab - NYC | Pandemic Response Lab, R&D | Henry Lee, Michael Hammerling, Melissa Hopkins, Cybill del Castillo, Shinyoung Clair Kang, William Ward, Pradeep Bugga, Haiping Hao, Jon Laurent |
| EPI_ISL_1098644, EPI_ISL_1098645 | Public Health Virology-Forensic and Scientific Services | Public Health Virology-Forensic and Scientific Services | Son Nguyen |
| EPI_ISL_1109556, EPI_ISL_1109557 | "Dr. Andrija Stampar" Teaching Institute of Public Health | Croatian Institute of Public Health | Irena Tabain, Ivana Ferenak |
| EPI_ISL_1111536, EPI_ISL_1111543, EPI_ISL_1111546 | Wyoming Public Health Laboratory | Wyoming Public Health Laboratory | Jim Mildenberger, Wanda Manley, Noah Hull, Taylor Fearing, Lynette Gumbleton, Channing Weber, Ashley Norberg, Chayse Rowley, Marley Goetz, Brian Dominguez, Elliot Thomasson, Cari Sloma, and Rob Christensen |
| EPI_ISL_1113148, EPI_ISL_1113149 | NJ Rapid Response Teams | New Jersey Public Health and Environmental Laboratories (PHEL) | Lindsey Bodnar, Shiv Verma, Dana Woell, Byeong Jeong |
| EPI_ISL_1114471 | Department of Health and Mental Hygiene Central Harlem | New York City Public Health Laboratory | Jade Wang, et al. |
| EPI_ISL_1114474 | Department of Health and Mental Hygiene Corona | New York City Public Health Laboratory | Jade Wang, et al. |
| EPI_ISL_1114478 | Department of Health and Mental Hygiene Jamaica | New York City Public Health Laboratory | Jade Wang, et al. |
| EPI_ISL_1114482 | Department of Health and Mental Hygiene Public Health Laboratory | New York City Public Health Laboratory | Jade Wang, et al. |
| EPI_ISL_1114495 | Department of Health and Mental Hygiene Fort Greene | New York City Public Health Laboratory | Jade Wang, et al. |
| EPI_ISL_1114499, EPI_ISL_1114502 | Department of Health and Mental Hygiene Crown Heights | New York City Public Health Laboratory | Jade Wang, et al. |
| EPI_ISL_1114515 | Department of Health and Mental Hygiene Corona | New York City Public Health Laboratory | Jade Wang, et al. |
| EPI_ISL_1114516, EPI_ISL_1114517 | Department of Health and Mental Hygiene Crown Heights | New York City Public Health Laboratory | Jade Wang, et al. |
| EPI_ISL_1114521 | Department of Health and Mental Hygiene Public Health Laboratory | New York City Public Health Laboratory | Jade Wang, et al. |
| EPI_ISL_1114524 | Department of Health and Mental Hygiene Chelsea | New York City Public Health Laboratory | Jade Wang, et al. |
| EPI_ISL_1114525 | OCME Office Of Chief Medical Examiner | New York City Public Health Laboratory | Jade Wang, et al. |
| EPI_ISL_1116510, EPI_ISL_1116514, EPI_ISL_1116516, EPI_ISL_1116517, EPI_ISL_1116518, EPI_ISL_1116519, EPI_ISL_1116520, EPI_ISL_1116526, EPI_ISL_1116527, EPI_ISL_1116529, EPI_ISL_1116531 | | | |
| see above | NYU Langone Health | Departments of Pathology and Medicine, New York University School of Medicine | Adriana Heguy, Dacia Dimartino, Emily Guzman, Christian Marier, Peter Meyn, Sitharam Ramaswami, Gael Westby, Paul Zappile, Yutong Zhang, Paolo Cotzia, Guiqing Wang |
| EPI_ISL_1121020 | Wyoming Public Health Laboratory | Wyoming Public Health Laboratory | Jim Mildenberger, Wanda Manley, Noah Hull, Taylor Fearing, Lynette Gumbleton, Channing Weber, Ashley Norberg, Chayse Rowley, Marley Goetz, Brian Dominguez, Elliot Thomasson, Cari Sloma, and Rob Christensen |
| EPI_ISL_1121888 | Department of Health & Mental Hygiene Morrisania | New York City Public Health Laboratory | Jade Wang, et al. |
| EPI_ISL_1121889 | Department of Health & Mental Hygiene Fort Greene | New York City Public Health Laboratory | Jade Wang, et al. |
| EPI_ISL_1121892, EPI_ISL_1121904 | Department of Health & Mental Hygiene Crown Heights | New York City Public Health Laboratory | Jade Wang, et al. |
| EPI_ISL_1121906 | Department of Health & Mental Hygiene Corona | New York City Public Health Laboratory | Jade Wang, et al. |
| EPI_ISL_1121910, EPI_ISL_1121911 | Department of Health & Mental Hygiene PHL | New York City Public Health Laboratory | Jade Wang, et al. |
| EPI_ISL_1121919, EPI_ISL_1121921 | Department of Health & Mental Hygiene Morrisania | New York City Public Health Laboratory | Jade Wang, et al. |
| EPI_ISL_1121929 | Department of Health & Mental Hygiene PHL | New York City Public Health Laboratory | Jade Wang, et al. |
| EPI_ISL_1121940 | Department of Health & Mental Hygiene Morrisania | New York City Public Health Laboratory | Jade Wang, et al. |
| EPI_ISL_1121950 | Department of Health & Mental Hygiene Central Harlem | New York City Public Health Laboratory | Jade Wang, et al. |
| EPI_ISL_1121953, EPI_ISL_1121956 | Department of Health & Mental Hygiene Jamaica | New York City Public Health Laboratory | Jade Wang, et al. |

| | | | |
|---|---|---|---|
| EPI_ISL_1137499, EPI_ISL_1137500 | New Jersey Department of Health, Public Health and Environmental Laboratories | New Jersey Public Health and Environmental Laboratories | Lindsey Bodnar, Shiv K. Verma, Dana Woell, Byeong Jeong |
| EPI_ISL_1153659 | MVZ Labor Dr. Fenner und Kollegen (Standort Hamburg) | Robert Koch Institute | unknown |
| EPI_ISL_1157425 | Northwestern Memorial Hospital | Northwestern University - Ozer Lab | Ramon Lorenzo-Redondo, Lacy M. Simons, Taylor J. Dean, Chad J. Achenbach, Lawrence J. Jennings, Chao Qi, Michael G. Ison, Judd F. Hultquist, Egon A. Ozer |
| EPI_ISL_1158070 | Michigan Department of Health and Human Services, Bureau of Laboratories | Michigan Department of Health and Human Services, Bureau of Laboratories | Blankenship HM, Riner D, Soehnlen MK |
| EPI_ISL_1160452, EPI_ISL_1160546, EPI_ISL_1160662, EPI_ISL_1162392, EPI_ISL_1162393, EPI_ISL_1162860 | Laboratory Corporation of America | Respiratory Viruses Branch, Division of Viral Diseases, Centers for Disease Control and Prevention | Peter W. Cook, Dakota Howard, Dhwani Batra, Ben L. Rambo-Martin, Minoo Agarwal, Eyad Almasri Debbie Boles, Ayla Burns, Nuthawin Charoensri, Oren Cohen, Susan Countryman, Mary Ann Cristobal, Bobbi Croy, Suzanne Dale, Hrushikesh Deshmukh, Amanda Douglas, Vincent Drouillon, Marcia Eisenberg, Howard Engler, Rama Ghatti, Prashant Gupta, Susan Hicks, Jake Humphrey, Lax Iyer, Manoj Jain, Mohan Kolli, Brian Krueger, Tim Kuphal, Stanley Letovsky, Michael Levandoski, Craig Lukasik, Jonathan Meltzer, Brian Norvell, Mindy Nye, Scott Parker, Christos Petropoulos, John Pruitt, Steven Ragan, Scott Ryan, Mike Sapeta, Jana Schroth, Suresh Babu Selvaraju, Goran Stevovic, Amanda Suchanek, Andrea Throop, Lyndon Tilson, Thomas Urban, Joe Voshell, Kimberly Wagner, Jonathan Williams, Mary Williamson, Qian Zeng, Tricia Zwiefelhofer, Clinton R. Paden, Suxiang Tong, Duncan MacCannell |
| EPI_ISL_1163684, EPI_ISL_1163685, EPI_ISL_1163686 | Yale Clinical Virology Lab | Grubaugh Lab - Yale School of Public Health | Joseph Fauver, Tara Alpert, Anderson Brito, Mallery Breban, Anne Wyllie, Chantal Vogels, Mary Petrone, Annie Watkins, Chaney Kalinich, Isabel Ott, Nathan Grubaugh |
| EPI_ISL_1164075 | National Virus Reference Laboratory | National Virus Reference Laboratory | Zoe Yandle, Charlene Bennet, Gabriel Gonzalez, Michael Carr, Jonathan Dean, Cillian F De Gascun |
| EPI_ISL_1164355 | National Public Health Laboratory, National Centre for Infectious Diseases | National Public Health Laboratory, National Centre for Infectious Diseases | Tze Minn Mak, Zhenyang Zhou, Lin Cui, Raymond Tzer Pin Lin |
| EPI_ISL_1164855 | Diagnostic Genomics Lab and Functional Genomics Core University of South Carolina | Diagnostic Genomics Lab and Functional Genomics Core, University of South Carolina | Carolyn Banister, Hao Ji, Diego Altomare, Herr Doktor Professor Genomiklaborsdirektor Universitätkovidenpandemieretter Phillip Buckhaults, Michael Shtutman, Helmut Albrecht |
| EPI_ISL_1168237 | TXDSHS | TXDSHS | Rashmi Tuladhar, Bonnie Oh, Jenny Zhang, Maliha Rahman, Mayela Pedrueza, Anita Pokharel, Myong Koag, Chun Wang, Rachel Lee, Grace Kubin |
| EPI_ISL_1168763, EPI_ISL_1168764 | Yale Pathology Lab | Grubaugh Lab - Yale School of Public Health | Tara Alpert, Joseph Fauver, Chen Liu, Pei Hui, Jianhui Wang, Susan Bell and Han Zhou, Anderson Brito, Mallery Breban, Anne Wyllie, Chantal Vogels, Mary Petrone, Chaney Kalinich, Isabel Ott, Arnau Casanovas, Catherine Muenker, Adam Moore, Alice Lu, Maria Tokuyama, Patrick Wong, Peiwen Lu, Saad Omer, Richard Martinello, Allison Nelson, Shelli Farhadian, Akiko Iwasaki, Charlese Dela Cruz, Albert Ko, Nathan Grubaugh |
| EPI_ISL_1172116, EPI_ISL_1172121, EPI_ISL_1172128, EPI_ISL_1172132, EPI_ISL_1172134, EPI_ISL_1172141, EPI_ISL_1172144, EPI_ISL_1172147, EPI_ISL_1172148, EPI_ISL_1172149, EPI_ISL_1172158, EPI_ISL_1172166, EPI_ISL_1172170, EPI_ISL_1172176, EPI_ISL_1172189, EPI_ISL_1172190, EPI_ISL_1172194, EPI_ISL_1172209, EPI_ISL_1172213, EPI_ISL_1172219, EPI_ISL_1172220, EPI_ISL_1172236, EPI_ISL_1172246, EPI_ISL_1172247, EPI_ISL_1172250, EPI_ISL_1172254, EPI_ISL_1172269, EPI_ISL_1172276, EPI_ISL_1172287, EPI_ISL_1172295, EPI_ISL_1172296, EPI_ISL_1172299, EPI_ISL_1172300, EPI_ISL_1172309, EPI_ISL_1172313, EPI_ISL_1172316, EPI_ISL_1172331, EPI_ISL_1172333, EPI_ISL_1172336, EPI_ISL_1172341, EPI_ISL_1172347, EPI_ISL_1172348, EPI_ISL_1172378, EPI_ISL_1172391, EPI_ISL_1172397, EPI_ISL_1172399, EPI_ISL_1172405, EPI_ISL_1172409, EPI_ISL_1172412, EPI_ISL_1172419, EPI_ISL_1172433, EPI_ISL_1172438, EPI_ISL_1172439, EPI_ISL_1172441, EPI_ISL_1172450, EPI_ISL_1172451, EPI_ISL_1172452, EPI_ISL_1172454, EPI_ISL_1172456, EPI_ISL_1172460, EPI_ISL_1172471, EPI_ISL_1172472, EPI_ISL_1172482, EPI_ISL_1172485, EPI_ISL_1172486, EPI_ISL_1172490, EPI_ISL_1172494, EPI_ISL_1172502, EPI_ISL_1172505, EPI_ISL_1172508, EPI_ISL_1172514, EPI_ISL_1172533, EPI_ISL_1172541, EPI_ISL_1172542, EPI_ISL_1172557, EPI_ISL_1172563, EPI_ISL_1172575, EPI_ISL_1172576, EPI_ISL_1172578, EPI_ISL_1172588, EPI_ISL_1172590, EPI_ISL_1172591, EPI_ISL_1172592, EPI_ISL_1172595, EPI_ISL_1172596, EPI_ISL_1172597, EPI_ISL_1172605, EPI_ISL_1172606, EPI_ISL_1172609, EPI_ISL_1172611, EPI_ISL_1172615, EPI_ISL_1172622, EPI_ISL_1172626, EPI_ISL_1172647, EPI_ISL_1172654, EPI_ISL_1172659, EPI_ISL_1172660, EPI_ISL_1172663, EPI_ISL_1172665, EPI_ISL_1172679, EPI_ISL_1172694, EPI_ISL_1172701, EPI_ISL_1172722, EPI_ISL_1172725, EPI_ISL_1172726, EPI_ISL_1172736, EPI_ISL_1172738, EPI_ISL_1172742, EPI_ISL_1172746, EPI_ISL_1172747, EPI_ISL_1172748, EPI_ISL_1172749, EPI_ISL_1172753, EPI_ISL_1172759, EPI_ISL_1172763, EPI_ISL_1172769, EPI_ISL_1172775, EPI_ISL_1172780, EPI_ISL_1172782, EPI_ISL_1172785, EPI_ISL_1172804, EPI_ISL_1172806, EPI_ISL_1172807, EPI_ISL_1172808, EPI_ISL_1172810, EPI_ISL_1172811, EPI_ISL_1172814, EPI_ISL_1172815, EPI_ISL_1172817, EPI_ISL_1172825, EPI_ISL_1172833, EPI_ISL_1172838, EPI_ISL_1172843, EPI_ISL_1172846, EPI_ISL_1172847, EPI_ISL_1172850, EPI_ISL_1172851, EPI_ISL_1172856, EPI_ISL_1172859, EPI_ISL_1172869, EPI_ISL_1172875, EPI_ISL_1172881, EPI_ISL_1172886, EPI_ISL_1172892, EPI_ISL_1172900, EPI_ISL_1172903, EPI_ISL_1172912, EPI_ISL_1172919, EPI_ISL_1172922, EPI_ISL_1172924, EPI_ISL_1172925, EPI_ISL_1172931, EPI_ISL_1172941, EPI_ISL_1172944, EPI_ISL_1172945, EPI_ISL_1172946, EPI_ISL_1172949, EPI_ISL_1172970, EPI_ISL_1172972, EPI_ISL_1172973, EPI_ISL_1172978, EPI_ISL_1172981, EPI_ISL_1172982, EPI_ISL_1172983, EPI_ISL_1172989, EPI_ISL_1172995, EPI_ISL_1173002, EPI_ISL_1173009, EPI_ISL_1173016, EPI_ISL_1173024, EPI_ISL_1173044, EPI_ISL_1173054, EPI_ISL_1173058, EPI_ISL_1173061, EPI_ISL_1173067, EPI_ISL_1173073, EPI_ISL_1173075, EPI_ISL_1173076, EPI_ISL_1173080, EPI_ISL_1173083, EPI_ISL_1173100, EPI_ISL_1173105, EPI_ISL_1173107, EPI_ISL_1173109, EPI_ISL_1173110, EPI_ISL_1173120, EPI_ISL_1173123, EPI_ISL_1173127, EPI_ISL_1173130, EPI_ISL_1173139, EPI_ISL_1173144, EPI_ISL_1173149, EPI_ISL_1173151, EPI_ISL_1173152, EPI_ISL_1173157 | | | |
| see above | Pandemic Response Lab - NYC | Pandemic Response Lab, R&D | Henry Lee, Michael Hammerling, Melissa Hopkins, Cybill del Castillo, Shinyoung Clair Kang, William Ward, Pradeep Bugga, Haiping Hao, Jon Laurent |
| EPI_ISL_1181668 | DPHL | Delaware Public Health Lab | Gregory Hovan |
| EPI_ISL_1182022 | Yale Clinical Virology Lab | Grubaugh Lab - Yale School of Public Health | Joseph Fauver, Mallery Breban, Isabell Ott, Tara Alpert, Mary Petrone, Anderson Brito, Chantal Vogels, Annie Watkins, Chaney Kalinich, Marie L. Landry, Nathan Grubaugh |
| EPI_ISL_1184102, EPI_ISL_1184103, EPI_ISL_1184106, EPI_ISL_1184109 | New Jersey Department of Health, Public Health and Environmental Laboratories | New Jersey Public Health and Environmental Laboratories | Lindsey Bodnar, Shiv K. Verma, Dana Woell, Byeong Jeong |
| EPI_ISL_1191460 | National Virus Reference Laboratory | National Virus Reference Laboratory | Zoe Yandle, Charlene Bennet, Gabriel Gonzalez, Michael Carr, Jonathan Dean, Cillian F De Gascun |
| EPI_ISL_1195753, EPI_ISL_1195759, EPI_ISL_1195761, EPI_ISL_1195763, EPI_ISL_1195775, EPI_ISL_1195776, EPI_ISL_1195780, EPI_ISL_1195789, EPI_ISL_1195808, EPI_ISL_1195810, EPI_ISL_1195827, EPI_ISL_1195830, EPI_ISL_1195833, EPI_ISL_1195843, EPI_ISL_1195857, EPI_ISL_1195866, EPI_ISL_1195879, EPI_ISL_1195901 | | | |
| see above | NYU Langone Health | Departments of Pathology and Medicine, New York University School of Medicine | Adriana Heguy, Dacia Dimartino, Emily Guzman, Christian Marier, Peter Meyn, Sitharam Ramaswami, Gael Westby, Paul Zappile, Yutong Zhang, Paolo Cotzia, Guiqing Wang |
| EPI_ISL_1197134, EPI_ISL_1197141, EPI_ISL_1197147 | USC Clinical Lab | Los Angeles County PHL | P. Hemarajata et al. |
| EPI_ISL_1197979 | New Mexico Department of Health Scientific Laboratory | Center for Global Health, University of New Mexico Health Sciences Center | Daryl Domman, Kurt Schwalm, Twila Kunde, Joseph Hicks, Anastacia Griego, Michael Edwards, Darrell Dinwiddie |
| EPI_ISL_1200535, EPI_ISL_1200563 | DOHMH Jamaica | New York City Public Health Laboratory | Jade Wang, et al. |
| EPI_ISL_1200564 | DOHMH Chelsea | New York City Public Health Laboratory | Jade Wang, et al. |
| EPI_ISL_1200567, EPI_ISL_1200578, EPI_ISL_1200580 | DOHMH Morrisania | New York City Public Health Laboratory | Jade Wang, et al. |
| EPI_ISL_1200590 | DOHMH Jamaica | New York City Public Health Laboratory | Jade Wang, et al. |
| EPI_ISL_1200604 | DOHMH Morrisania | New York City Public Health Laboratory | Jade Wang, et al. |
| EPI_ISL_1200605 | DOHMH Crown Heights | New York City Public Health Laboratory | Jade Wang, et al. |
| EPI_ISL_1200606 | DOHMH Riverside | New York City Public Health Laboratory | Jade Wang, et al. |
| EPI_ISL_1200610 | OCME Office Of Chief Medical Examiner | New York City Public Health Laboratory | Jade Wang, et al. |
| EPI_ISL_1203819, EPI_ISL_1203826, EPI_ISL_1203827, EPI_ISL_1203828 | The Jackson Laboratory | The Jackson Laboratory | Lloyd M, Sanderson B, Srivastava A, Maurya R, Renzette N, Omerza G, Kelly K, Li L, Wei C L, Adams M |
| EPI_ISL_1205453 | Mayo Clinic & Mayo Clinic Laboratories | Minnesota Department of Health, Public Health Laboratory | Alexandra Lorentz, Jacob Garfin, Matt Plumb, and Xiong Wang |
| EPI_ISL_1218888 | Florida Bureau of Public Health Laboratories | Florida Bureau of Public Health Laboratories | Sarah Schmedes, Jason Blanton |
| EPI_ISL_1221173, EPI_ISL_1221352, EPI_ISL_1221982, EPI_ISL_1222140, EPI_ISL_1222413 | Laboratory Corporation of America | Centers for Disease Control and Prevention Division of Viral Diseases, Pathogen Discovery | Peter W. Cook, Dakota Howard, Dhwani Batra, Ben L. Rambo-Martin, Minoo Agarwal, Eyad Almasri, Debbie Boles, Ayla Burns, Nuthawin Charoensri, Oren Cohen, Susan Countryman, Mary Ann Cristobal, Bobbi Croy, Suzanne Dale, Hrushikesh Deshmukh, Amanda Douglas, Vincent Drouillon, Marcia Eisenberg, Howard Engler, Rama Ghatti, Prashant Gupta, Susan Hicks, Jake Humphrey, Lax Iyer, Manoj Jain, Mohan Kolli, Brian Krueger, Tim Kuphal, Stanley Letovsky, Michael Levandoski, Craig Lukasik, Jonathan Meltzer, Brian Norvell, Mindy Nye, Scott Parker, Christos Petropoulos, John Pruitt, Steven Ragan, Scott Ryan, Mike Sapeta, Jana Schroth, Suresh Babu Selvaraju, Goran Stevovic, Amanda Suchanek, Andrea Throop, Lyndon Tilson, Thomas Urban, Joe Voshell, Kimberly Wagner, Jonathan Williams, Mary Williamson, Qian Zeng, Tricia Zwiefelhofer, Clinton R. Paden, Suxiang Tong, Duncan MacCannell |

| | | | |
|---|---|---|---|
| EPI_ISL_1225597, EPI_ISL_1225608, EPI_ISL_1225609 | Yale Clinical Virology Lab | Grubaugh Lab - Yale School of Public Health | Joseph Fauver, Mallery Breban, Isabell Ott, Tara Alpert, Mary Petrone, Anderson Brito, Chantal Vogels, Annie Watkins, Chaney Kalinich, Marie L. Landry, Nathan Grubaugh |
| EPI_ISL_1225664 | Murphy Medical Association | Grubaugh Lab - Yale School of Public Health | Joseph Fauver, Mallery Breban, Isabell Ott, Tara Alpert, Mary Petrone, Anderson Brito, Chantal Vogels, Annie Watkins, Chaney Kalinich, Caleb Neal, Eva Laszlo, Steven Murphy, Nathan Grubaugh |
| EPI_ISL_1225668, EPI_ISL_1225670 | Yale Clinical Virology Lab | Grubaugh Lab - Yale School of Public Health | Joseph Fauver, Mallery Breban, Isabell Ott, Tara Alpert, Mary Petrone, Anderson Brito, Chantal Vogels, Annie Watkins, Chaney Kalinich, Marie L. Landry, Nathan Grubaugh |
| EPI_ISL_1225699, EPI_ISL_1225701, EPI_ISL_1225713 | Murphy Medical Association | Grubaugh Lab - Yale School of Public Health | Joseph Fauver, Mallery Breban, Isabell Ott, Tara Alpert, Mary Petrone, Anderson Brito, Chantal Vogels, Annie Watkins, Chaney Kalinich, Caleb Neal, Eva Laszlo, Steven Murphy, Nathan Grubaugh |
| EPI_ISL_1225729, EPI_ISL_1225730 | Yale Clinical Virology Lab | Grubaugh Lab - Yale School of Public Health | Joseph Fauver, Mallery Breban, Isabell Ott, Tara Alpert, Mary Petrone, Anderson Brito, Chantal Vogels, Annie Watkins, Chaney Kalinich, Marie L. Landry, Nathan Grubaugh |
| EPI_ISL_1226419, EPI_ISL_1226440, EPI_ISL_1226595 | NYC Pandemic Response Lab | Wadsworth Center, New York State Department of Health | Kirsten St. George, Daryl M. Lamson, Alexis Russel, Matthew Shudt, Melissa A Leisner, Jonathan Plitnick, Navjot Singh, John Kelly, Erasmus Schneider, Erica Lasek-Nesselquist |
| EPI_ISL_1226702, EPI_ISL_1226723, EPI_ISL_1226727 | WESTCHESTER MEDICAL CENTER | Wadsworth Center, New York State Department of Health | Kirsten St. George, Daryl M. Lamson, Alexis Russel, Matthew Shudt, Melissa A Leisner, Jonathan Plitnick, Navjot Singh, John Kelly, Erasmus Schneider, Erica Lasek-Nesselquist |
| EPI_ISL_1226728 | Columbia University Irving Medical Center | Wadsworth Center, New York State Department of Health | Kirsten St. George, Daryl M. Lamson, Alexis Russel, Matthew Shudt, Melissa A Leisner, Jonathan Plitnick, Navjot Singh, John Kelly, Erasmus Schneider, Erica Lasek-Nesselquist |
| EPI_ISL_1226735, EPI_ISL_1226737, EPI_ISL_1226740, EPI_ISL_1226741 | MONTEFIORE MEDICAL CENTER LABORATORIES | Wadsworth Center, New York State Department of Health | Kirsten St. George, Daryl M. Lamson, Alexis Russel, Matthew Shudt, Melissa A Leisner, Jonathan Plitnick, Navjot Singh, John Kelly, Erasmus Schneider, Erica Lasek-Nesselquist |
| EPI_ISL_1226743 | Columbia University Irving Medical Center | Wadsworth Center, New York State Department of Health | Kirsten St. George, Daryl M. Lamson, Alexis Russel, Matthew Shudt, Melissa A Leisner, Jonathan Plitnick, Navjot Singh, John Kelly, Erasmus Schneider, Erica Lasek-Nesselquist |
| EPI_ISL_1226744, EPI_ISL_1226746, EPI_ISL_1226749, EPI_ISL_1226750, EPI_ISL_1226751, EPI_ISL_1226753, EPI_ISL_1226760 | MONTEFIORE MEDICAL CENTER LABORATORIES | Wadsworth Center, New York State Department of Health | Kirsten St. George, Daryl M. Lamson, Alexis Russel, Matthew Shudt, Melissa A Leisner, Jonathan Plitnick, Navjot Singh, John Kelly, Erasmus Schneider, Erica Lasek-Nesselquist |
| EPI_ISL_1226768, EPI_ISL_1226769 | NYC Pandemic Response Lab | Wadsworth Center, New York State Department of Health | Kirsten St. George, Daryl M. Lamson, Alexis Russel, Matthew Shudt, Melissa A Leisner, Jonathan Plitnick, Navjot Singh, John Kelly, Erasmus Schneider, Erica Lasek-Nesselquist |
| EPI_ISL_1226770 | MONTEFIORE MEDICAL CENTER LABORATORIES | Wadsworth Center, New York State Department of Health | Kirsten St. George, Daryl M. Lamson, Alexis Russel, Matthew Shudt, Melissa A Leisner, Jonathan Plitnick, Navjot Singh, John Kelly, Erasmus Schneider, Erica Lasek-Nesselquist |
| EPI_ISL_1226772 | WESTCHESTER MEDICAL CENTER | Wadsworth Center, New York State Department of Health | Kirsten St. George, Daryl M. Lamson, Alexis Russel, Matthew Shudt, Melissa A Leisner, Jonathan Plitnick, Navjot Singh, John Kelly, Erasmus Schneider, Erica Lasek-Nesselquist |
| EPI_ISL_1226776, EPI_ISL_1226780 | MONTEFIORE MEDICAL CENTER LABORATORIES | Wadsworth Center, New York State Department of Health | Kirsten St. George, Daryl M. Lamson, Alexis Russel, Matthew Shudt, Melissa A Leisner, Jonathan Plitnick, Navjot Singh, John Kelly, Erasmus Schneider, Erica Lasek-Nesselquist |
| EPI_ISL_1226782, EPI_ISL_1226784, EPI_ISL_1226786, EPI_ISL_1226787 | WESTCHESTER MEDICAL CENTER | Wadsworth Center, New York State Department of Health | Kirsten St. George, Daryl M. Lamson, Alexis Russel, Matthew Shudt, Melissa A Leisner, Jonathan Plitnick, Navjot Singh, John Kelly, Erasmus Schneider, Erica Lasek-Nesselquist |
| EPI_ISL_1226796, EPI_ISL_1226797 | NYC Pandemic Response Lab | Wadsworth Center, New York State Department of Health | Kirsten St. George, Daryl M. Lamson, Alexis Russel, Matthew Shudt, Melissa A Leisner, Jonathan Plitnick, Navjot Singh, John Kelly, Erasmus Schneider, Erica Lasek-Nesselquist |
| EPI_ISL_1226801, EPI_ISL_1226802, EPI_ISL_1226806, EPI_ISL_1226807 | Columbia University Irving Medical Center | Wadsworth Center, New York State Department of Health | Kirsten St. George, Daryl M. Lamson, Alexis Russel, Matthew Shudt, Melissa A Leisner, Jonathan Plitnick, Navjot Singh, John Kelly, Erasmus Schneider, Erica Lasek-Nesselquist |
| EPI_ISL_1226808 | NYC Pandemic Response Lab | Wadsworth Center, New York State Department of Health | Kirsten St. George, Daryl M. Lamson, Alexis Russel, Matthew Shudt, Melissa A Leisner, Jonathan Plitnick, Navjot Singh, John Kelly, Erasmus Schneider, Erica Lasek-Nesselquist |
| EPI_ISL_1226812 | New York Presbyterian Hospital | Wadsworth Center, New York State Department of Health | Kirsten St. George, Daryl M. Lamson, Alexis Russel, Matthew Shudt, Melissa A Leisner, Jonathan Plitnick, Navjot Singh, John Kelly, Erasmus Schneider, Erica Lasek-Nesselquist |
| EPI_ISL_1226820 | MEMORIAL SLOAN KETTERING CANCER CENTER | Wadsworth Center, New York State Department of Health | Kirsten St. George, Daryl M. Lamson, Alexis Russel, Matthew Shudt, Melissa A Leisner, Jonathan Plitnick, Navjot Singh, John Kelly, Erasmus Schneider, Erica Lasek-Nesselquist |
| EPI_ISL_1226824 | Columbia University Irving Medical Center | Wadsworth Center, New York State Department of Health | Kirsten St. George, Daryl M. Lamson, Alexis Russel, Matthew Shudt, Melissa A Leisner, Jonathan Plitnick, Navjot Singh, John Kelly, Erasmus Schneider, Erica Lasek-Nesselquist |
| EPI_ISL_1226825, EPI_ISL_1226826, EPI_ISL_1226829, EPI_ISL_1226834 | WESTCHESTER MEDICAL CENTER | Wadsworth Center, New York State Department of Health | Kirsten St. George, Daryl M. Lamson, Alexis Russel, Matthew Shudt, Melissa A Leisner, Jonathan Plitnick, Navjot Singh, John Kelly, Erasmus Schneider, Erica Lasek-Nesselquist |
| EPI_ISL_1226873 | SUNY UPSTATE MEDICAL UNIVERSITY | Wadsworth Center, New York State Department of Health | Kirsten St. George, Daryl M. Lamson, Alexis Russel, Matthew Shudt, Melissa A Leisner, Jonathan Plitnick, Navjot Singh, John Kelly, Erasmus Schneider, Erica Lasek-Nesselquist |
| EPI_ISL_1226935 | NYC Pandemic Response Lab | Wadsworth Center, New York State Department of Health | Kirsten St. George, Daryl M. Lamson, Alexis Russel, Matthew Shudt, Melissa A Leisner, Jonathan Plitnick, Navjot Singh, John Kelly, Erasmus Schneider, Erica Lasek-Nesselquist |
| EPI_ISL_1226939, EPI_ISL_1226943, EPI_ISL_1226944, EPI_ISL_1226945, EPI_ISL_1226946, EPI_ISL_1226947, EPI_ISL_1226948, EPI_ISL_1226951, EPI_ISL_1226953, EPI_ISL_1226957, EPI_ISL_1226960, EPI_ISL_1226965, EPI_ISL_1226967, EPI_ISL_1226975 | | | |
| see above | MONTEFIORE MEDICAL CENTER LABORATORIES | Wadsworth Center, New York State Department of Health | Kirsten St. George, Daryl M. Lamson, Alexis Russel, Matthew Shudt, Melissa A Leisner, Jonathan Plitnick, Navjot Singh, John Kelly, Erasmus Schneider, Erica Lasek-Nesselquist |
| EPI_ISL_1226981 | SUNY UPSTATE MEDICAL UNIVERSITY | Wadsworth Center, New York State Department of Health | Kirsten St. George, Daryl M. Lamson, Alexis Russel, Matthew Shudt, Melissa A Leisner, Jonathan Plitnick, Navjot Singh, John Kelly, Erasmus Schneider, Erica Lasek-Nesselquist |
| EPI_ISL_1226985 | BIO-REFERENCE LABORATORIES | Wadsworth Center, New York State Department of Health | Kirsten St. George, Daryl M. Lamson, Alexis Russel, Matthew Shudt, Melissa A Leisner, Jonathan Plitnick, Navjot Singh, John Kelly, Erasmus Schneider, Erica Lasek-Nesselquist |
| EPI_ISL_1226991, EPI_ISL_1226993, EPI_ISL_1226997 | Columbia University Irving Medical Center | Wadsworth Center, New York State Department of Health | Kirsten St. George, Daryl M. Lamson, Alexis Russel, Matthew Shudt, Melissa A Leisner, Jonathan Plitnick, Navjot Singh, John Kelly, Erasmus Schneider, Erica Lasek-Nesselquist |
| EPI_ISL_1226998 | NYC Pandemic Response Lab | Wadsworth Center, New York State Department of Health | Kirsten St. George, Daryl M. Lamson, Alexis Russel, Matthew Shudt, Melissa A Leisner, Jonathan Plitnick, Navjot Singh, John Kelly, Erasmus Schneider, Erica Lasek-Nesselquist |
| EPI_ISL_1227002, EPI_ISL_1227003, EPI_ISL_1227004 | Columbia University Irving Medical Center | Wadsworth Center, New York State Department of Health | Kirsten St. George, Daryl M. Lamson, Alexis Russel, Matthew Shudt, Melissa A Leisner, Jonathan Plitnick, Navjot Singh, John Kelly, Erasmus Schneider, Erica Lasek-Nesselquist |
| EPI_ISL_1227005, EPI_ISL_1227009 | NYC Pandemic Response Lab | Wadsworth Center, New York State Department of Health | Kirsten St. George, Daryl M. Lamson, Alexis Russel, Matthew Shudt, Melissa A Leisner, Jonathan Plitnick, Navjot Singh, John Kelly, Erasmus Schneider, Erica Lasek-Nesselquist |
| EPI_ISL_1227016, EPI_ISL_1227021, EPI_ISL_1227024 | MONTEFIORE MEDICAL CENTER LABORATORIES | Wadsworth Center, New York State Department of Health | Kirsten St. George, Daryl M. Lamson, Alexis Russel, Matthew Shudt, Melissa A Leisner, Jonathan Plitnick, Navjot Singh, John Kelly, Erasmus Schneider, Erica Lasek-Nesselquist |
| EPI_ISL_1227032 | ALBANY MEDICAL CENTER | Wadsworth Center, New York State Department of Health | Kirsten St. George, Daryl M. Lamson, Alexis Russel, Matthew Shudt, Melissa A Leisner, Jonathan Plitnick, Navjot Singh, John Kelly, Erasmus Schneider, |

| | | | |
|---|---|---|---|
| | | | Erica Lasek-Nesselquist |
| EPI_ISL_1227035, EPI_ISL_1227037, EPI_ISL_1227039, EPI_ISL_1227044, EPI_ISL_1227048, EPI_ISL_1227049, EPI_ISL_1227050, EPI_ISL_1227051, EPI_ISL_1227053, EPI_ISL_1227056, EPI_ISL_1227057, EPI_ISL_1227059, EPI_ISL_1227060, EPI_ISL_1227061, EPI_ISL_1227062 | | | |
| see above | MONTEFIORE MEDICAL CENTER LABORATORIES | Wadsworth Center, New York State Department of Health | Kirsten St. George, Daryl M. Lamson, Alexis Russel, Matthew Shudt, Melissa A Leisner, Jonathan Plitnick, Navjot Singh, John Kelly, Erasmus Schneider, Erica Lasek-Nesselquist |
| EPI_ISL_1227068 | Columbia University Irving Medical Center | Wadsworth Center, New York State Department of Health | Kirsten St. George, Daryl M. Lamson, Alexis Russel, Matthew Shudt, Melissa A Leisner, Jonathan Plitnick, Navjot Singh, John Kelly, Erasmus Schneider, Erica Lasek-Nesselquist |
| EPI_ISL_1227070, EPI_ISL_1227071, EPI_ISL_1227073, EPI_ISL_1227075, EPI_ISL_1227083, EPI_ISL_1227084 | MONTEFIORE MEDICAL CENTER LABORATORIES | Wadsworth Center, New York State Department of Health | Kirsten St. George, Daryl M. Lamson, Alexis Russel, Matthew Shudt, Melissa A Leisner, Jonathan Plitnick, Navjot Singh, John Kelly, Erasmus Schneider, Erica Lasek-Nesselquist |
| EPI_ISL_1227087 | NYC Pandemic Response Lab | Wadsworth Center, New York State Department of Health | Kirsten St. George, Daryl M. Lamson, Alexis Russel, Matthew Shudt, Melissa A Leisner, Jonathan Plitnick, Navjot Singh, John Kelly, Erasmus Schneider, Erica Lasek-Nesselquist |
| EPI_ISL_1227090, EPI_ISL_1227093, EPI_ISL_1227094, EPI_ISL_1227097, EPI_ISL_1227098, EPI_ISL_1227099, EPI_ISL_1227103, EPI_ISL_1227104, EPI_ISL_1227108 | MONTEFIORE MEDICAL CENTER LABORATORIES | Wadsworth Center, New York State Department of Health | Kirsten St. George, Daryl M. Lamson, Alexis Russel, Matthew Shudt, Melissa A Leisner, Jonathan Plitnick, Navjot Singh, John Kelly, Erasmus Schneider, Erica Lasek-Nesselquist |
| EPI_ISL_1227109 | NYC Pandemic Response Lab | Wadsworth Center, New York State Department of Health | Kirsten St. George, Daryl M. Lamson, Alexis Russel, Matthew Shudt, Melissa A Leisner, Jonathan Plitnick, Navjot Singh, John Kelly, Erasmus Schneider, Erica Lasek-Nesselquist |
| EPI_ISL_1227112, EPI_ISL_1227125, EPI_ISL_1227127, EPI_ISL_1227128 | MONTEFIORE MEDICAL CENTER LABORATORIES | Wadsworth Center, New York State Department of Health | Kirsten St. George, Daryl M. Lamson, Alexis Russel, Matthew Shudt, Melissa A Leisner, Jonathan Plitnick, Navjot Singh, John Kelly, Erasmus Schneider, Erica Lasek-Nesselquist |
| EPI_ISL_1227132 | WESTCHESTER MEDICAL CENTER | Wadsworth Center, New York State Department of Health | Kirsten St. George, Daryl M. Lamson, Alexis Russel, Matthew Shudt, Melissa A Leisner, Jonathan Plitnick, Navjot Singh, John Kelly, Erasmus Schneider, Erica Lasek-Nesselquist |
| EPI_ISL_1227138, EPI_ISL_1227139, EPI_ISL_1227141, EPI_ISL_1227146, EPI_ISL_1227147, EPI_ISL_1227156, EPI_ISL_1227159, EPI_ISL_1227160 | NYC Pandemic Response Lab | Wadsworth Center, New York State Department of Health | Kirsten St. George, Daryl M. Lamson, Alexis Russel, Matthew Shudt, Melissa A Leisner, Jonathan Plitnick, Navjot Singh, John Kelly, Erasmus Schneider, Erica Lasek-Nesselquist |
| EPI_ISL_1227166 | Columbia University Irving Medical Center | Wadsworth Center, New York State Department of Health | Kirsten St. George, Daryl M. Lamson, Alexis Russel, Matthew Shudt, Melissa A Leisner, Jonathan Plitnick, Navjot Singh, John Kelly, Erasmus Schneider, Erica Lasek-Nesselquist |
| EPI_ISL_1227167, EPI_ISL_1227170, EPI_ISL_1227171 | NYC Pandemic Response Lab | Wadsworth Center, New York State Department of Health | Kirsten St. George, Daryl M. Lamson, Alexis Russel, Matthew Shudt, Melissa A Leisner, Jonathan Plitnick, Navjot Singh, John Kelly, Erasmus Schneider, Erica Lasek-Nesselquist |
| EPI_ISL_1227174 | WESTCHESTER MEDICAL CENTER | Wadsworth Center, New York State Department of Health | Kirsten St. George, Daryl M. Lamson, Alexis Russel, Matthew Shudt, Melissa A Leisner, Jonathan Plitnick, Navjot Singh, John Kelly, Erasmus Schneider, Erica Lasek-Nesselquist |
| EPI_ISL_1227180 | MEMORIAL SLOAN KETTERING CANCER CENTER | Wadsworth Center, New York State Department of Health | Kirsten St. George, Daryl M. Lamson, Alexis Russel, Matthew Shudt, Melissa A Leisner, Jonathan Plitnick, Navjot Singh, John Kelly, Erasmus Schneider, Erica Lasek-Nesselquist |
| EPI_ISL_1227184, EPI_ISL_1227186, EPI_ISL_1227187 | NYC Pandemic Response Lab | Wadsworth Center, New York State Department of Health | Kirsten St. George, Daryl M. Lamson, Alexis Russel, Matthew Shudt, Melissa A Leisner, Jonathan Plitnick, Navjot Singh, John Kelly, Erasmus Schneider, Erica Lasek-Nesselquist |
| EPI_ISL_1227189 | WESTCHESTER MEDICAL CENTER | Wadsworth Center, New York State Department of Health | Kirsten St. George, Daryl M. Lamson, Alexis Russel, Matthew Shudt, Melissa A Leisner, Jonathan Plitnick, Navjot Singh, John Kelly, Erasmus Schneider, Erica Lasek-Nesselquist |
| EPI_ISL_1227191, EPI_ISL_1227192, EPI_ISL_1227194, EPI_ISL_1227199, EPI_ISL_1227202, EPI_ISL_1227203, EPI_ISL_1227204 | NYC Pandemic Response Lab | Wadsworth Center, New York State Department of Health | Kirsten St. George, Daryl M. Lamson, Alexis Russel, Matthew Shudt, Melissa A Leisner, Jonathan Plitnick, Navjot Singh, John Kelly, Erasmus Schneider, Erica Lasek-Nesselquist |
| EPI_ISL_1227207 | WESTCHESTER MEDICAL CENTER | Wadsworth Center, New York State Department of Health | Kirsten St. George, Daryl M. Lamson, Alexis Russel, Matthew Shudt, Melissa A Leisner, Jonathan Plitnick, Navjot Singh, John Kelly, Erasmus Schneider, Erica Lasek-Nesselquist |
| EPI_ISL_1227209 | ALBANY MEDICAL CENTER | Wadsworth Center, New York State Department of Health | Kirsten St. George, Daryl M. Lamson, Alexis Russel, Matthew Shudt, Melissa A Leisner, Jonathan Plitnick, Navjot Singh, John Kelly, Erasmus Schneider, Erica Lasek-Nesselquist |
| EPI_ISL_1227211 | MEMORIAL SLOAN KETTERING CANCER CENTER | Wadsworth Center, New York State Department of Health | Kirsten St. George, Daryl M. Lamson, Alexis Russel, Matthew Shudt, Melissa A Leisner, Jonathan Plitnick, Navjot Singh, John Kelly, Erasmus Schneider, Erica Lasek-Nesselquist |
| EPI_ISL_1227218, EPI_ISL_1227219 | WESTCHESTER MEDICAL CENTER | Wadsworth Center, New York State Department of Health | Kirsten St. George, Daryl M. Lamson, Alexis Russel, Matthew Shudt, Melissa A Leisner, Jonathan Plitnick, Navjot Singh, John Kelly, Erasmus Schneider, Erica Lasek-Nesselquist |
| EPI_ISL_1227221 | NYC Pandemic Response Lab | Wadsworth Center, New York State Department of Health | Kirsten St. George, Daryl M. Lamson, Alexis Russel, Matthew Shudt, Melissa A Leisner, Jonathan Plitnick, Navjot Singh, John Kelly, Erasmus Schneider, Erica Lasek-Nesselquist |
| EPI_ISL_1227223, EPI_ISL_1227225 | WESTCHESTER MEDICAL CENTER | Wadsworth Center, New York State Department of Health | Kirsten St. George, Daryl M. Lamson, Alexis Russel, Matthew Shudt, Melissa A Leisner, Jonathan Plitnick, Navjot Singh, John Kelly, Erasmus Schneider, Erica Lasek-Nesselquist |
| EPI_ISL_1231730, EPI_ISL_1231847, EPI_ISL_1231857 | Fulgent Genetics | Fulgent Genetics | Harry Gao, Mickey Li, John Gao, Joseph Fierro, Benafsh Sapra, Becky Tsai, Yan Meng, Doreen Ng, James Xie |
| EPI_ISL_1235195 | CA DPH Viral and Rickettsial Disease Laboratory | Chan-Zuckerberg Biohub | CZB Cliahub Consortium |
| EPI_ISL_1235709 | Houston Methodist Hospital | Houston Methodist Hospital | S. Wesley Long, Randall J. Olsen, Paul A. Christensen, Sishir Subedi, Robert Olson, James J. Davis, Matthew Ojeda Saavedra, Prasanti Yerramilli, Layne Pruitt, Kristina Reppond, Madison N. Shyer, Jessica Cambric, Ilya J. Finkelstein, Jimmy Gollihar, and James M. Musser |
| EPI_ISL_1240765, EPI_ISL_1240772 | University Hospitals Translational Laboratory (UHTL), University Hospitals | University Hospitals Translational Laboratory (UHTL), University Hospitals | Sadri,N., Alouani,D., Song,X. |
| EPI_ISL_1241837 | PHV-FSS | PHV-FSS | Son Nguyen |
| EPI_ISL_1244858 | Lighthouse Lab in Cambridge | Wellcome Sanger Institute for the COVID-19 Genomics UK (COG-UK) Consortium | Rob Howes, The Lighthouse Lab in Cambridge and Alex Alderton, Roberto Amato, Jeffrey Barrett, Sonia Goncalves, Ewan Harrison, David K. Jackson, Ian Johnston, Dominic Kwiatkowski, Cordelia Langford, John Sillitoe on behalf of the Wellcome Sanger Institute COVID-19 Surveillance Team |
| EPI_ISL_1253798 | Broad Institute Clinical Research Sequencing Platform | Infectious Disease Program, Broad Institute of Harvard and MIT | Lemieux,J.E., Siddle,K.J., Adams,G., Gladden-Young,A., Lagerborg,K., Rudy,M., DeRuff,K., Carter,A., Normandin,E., Bauer,M., Reilly,S., Tomkins-Tinch,C., Loreth,C., Chaluvadi,S., Birren,B.W., Gallagher,G., Smole,S., Park,D.J., MacInnis,B.L., and Sabeti,P.C. |
| EPI_ISL_1254233, EPI_ISL_1254234, EPI_ISL_1254237, EPI_ISL_1254242, EPI_ISL_1254243, EPI_ISL_1254259, EPI_ISL_1254283, EPI_ISL_1254285, EPI_ISL_1254288, EPI_ISL_1254293 | NJDOH, Public Health and Environmental Laboratories | NJ_PHEL | Lindsey Bodnar, Shiv K. Verma, Dana Woell, Byeong Jeong |
| EPI_ISL_1258470, EPI_ISL_1258471, EPI_ISL_1258482, EPI_ISL_1258484, EPI_ISL_1258486, EPI_ISL_1258490, EPI_ISL_1258502, EPI_ISL_1258508, EPI_ISL_1258509, EPI_ISL_1258515, EPI_ISL_1258525, EPI_ISL_1258535, EPI_ISL_1258536, EPI_ISL_1258537, EPI_ISL_1258538, EPI_ISL_1258540, EPI_ISL_1258543, | | | |

| | | | |
|---|---|---|---|
| EPI_ISL_1258550, EPI_ISL_1258553, EPI_ISL_1258589, EPI_ISL_1258637, EPI_ISL_1258638, EPI_ISL_1258640, EPI_ISL_1258641, EPI_ISL_1258643, EPI_ISL_1258660, EPI_ISL_1258672, EPI_ISL_1258681, EPI_ISL_1258683, EPI_ISL_1258684, EPI_ISL_1258685, EPI_ISL_1258716, EPI_ISL_1258717, EPI_ISL_1258718, EPI_ISL_1258719, EPI_ISL_1258723, EPI_ISL_1258725, EPI_ISL_1258726, EPI_ISL_1258728, EPI_ISL_1258730, EPI_ISL_1258731, EPI_ISL_1258733, EPI_ISL_1258734, EPI_ISL_1258736, EPI_ISL_1258737, EPI_ISL_1258739, EPI_ISL_1258741, EPI_ISL_1258744, EPI_ISL_1258746, EPI_ISL_1258755, EPI_ISL_1258756, EPI_ISL_1258760, EPI_ISL_1258767, EPI_ISL_1258768, EPI_ISL_1258769, EPI_ISL_1258770, EPI_ISL_1258774, EPI_ISL_1258776, EPI_ISL_1258777, EPI_ISL_1258786, EPI_ISL_1258795, EPI_ISL_1258796, EPI_ISL_1258799, EPI_ISL_1258802, EPI_ISL_1258804, EPI_ISL_1258806, EPI_ISL_1258807, EPI_ISL_1258811, EPI_ISL_1258816, EPI_ISL_1258820, EPI_ISL_1258822, EPI_ISL_1258827, EPI_ISL_1258828, EPI_ISL_1258830, EPI_ISL_1258831, EPI_ISL_1258835, EPI_ISL_1258838, EPI_ISL_1258839, EPI_ISL_1258840, EPI_ISL_1258841, EPI_ISL_1258842, EPI_ISL_1258843, EPI_ISL_1258849, EPI_ISL_1258854, EPI_ISL_1258860, EPI_ISL_1258868, EPI_ISL_1258870, EPI_ISL_1258873, EPI_ISL_1258884, EPI_ISL_1258885, EPI_ISL_1258888, EPI_ISL_1258890, EPI_ISL_1258892, EPI_ISL_1258902, EPI_ISL_1258903, EPI_ISL_1258904, EPI_ISL_1258907, EPI_ISL_1258908, EPI_ISL_1258913, EPI_ISL_1258915, EPI_ISL_1258919, EPI_ISL_1258926, EPI_ISL_1258927 | | | |
| see above | Pandemic Response Lab - NYC | Pandemic Response Lab, R&D | Henry Lee, Michael Hammerling, Melissa Hopkins, Cybill del Castillo, Shinyoung Clair Kang, William Ward, Pradeep Bugga, Haiping Hao, Jon Laurent |
| EPI_ISL_1261140, EPI_ISL_1261149 | Molecular & Genomic Pathology Laboratory, Thomas Jefferson University Hospital | Molecular & Genomic Pathology Laboratory, Thomas Jefferson University Hospital | Nitika Badjatia, Run Jin, Zi-xuan Wang |
| EPI_ISL_1261814 | DOHMH Morrisania | New York City Public Health Laboratory | Jade Wang, et al. |
| EPI_ISL_1261817 | DOHMH Chelsea | New York City Public Health Laboratory | Jade Wang, et al. |
| EPI_ISL_1261821 | DOHMH Crown Heights | New York City Public Health Laboratory | Jade Wang, et al. |
| EPI_ISL_1261822 | DOHMH Jamaica | New York City Public Health Laboratory | Jade Wang, et al. |
| EPI_ISL_1261824 | DOHMH Central Harlem | New York City Public Health Laboratory | Jade Wang, et al. |
| EPI_ISL_1261828, EPI_ISL_1261829 | OCME Office Of Chief Medical Examiner | New York City Public Health Laboratory | Jade Wang, et al. |
| EPI_ISL_1261831 | DOHMH Jamaica | New York City Public Health Laboratory | Jade Wang, et al. |
| EPI_ISL_1261835, EPI_ISL_1261837 | DOHMH Corona | New York City Public Health Laboratory | Jade Wang, et al. |
| EPI_ISL_1261838 | DOHMH Morrisania | New York City Public Health Laboratory | Jade Wang, et al. |
| EPI_ISL_1261840, EPI_ISL_1261841, EPI_ISL_1261842 | DOHMH Riverside | New York City Public Health Laboratory | Jade Wang, et al. |
| EPI_ISL_1261843 | DOHMH Central Harlem | New York City Public Health Laboratory | Jade Wang, et al. |
| EPI_ISL_1261845 | DOHMH Riverside | New York City Public Health Laboratory | Jade Wang, et al. |
| EPI_ISL_1261848 | DOHMH Morrisania | New York City Public Health Laboratory | Jade Wang, et al. |
| EPI_ISL_1261852 | DOHMH Chelsea | New York City Public Health Laboratory | Jade Wang, et al. |
| EPI_ISL_1261853 | Department of Homeless Services | New York City Public Health Laboratory | Jade Wang, et al. |
| EPI_ISL_1261854 | OCME Office Of Chief Medical Examiner | New York City Public Health Laboratory | Jade Wang, et al. |
| EPI_ISL_1262507, EPI_ISL_1262508, EPI_ISL_1262517, EPI_ISL_1262533, EPI_ISL_1262547, EPI_ISL_1262551, EPI_ISL_1262552, EPI_ISL_1262554, EPI_ISL_1262556, EPI_ISL_1262559, EPI_ISL_1262561, EPI_ISL_1262566, EPI_ISL_1262567, EPI_ISL_1262568 | | | |
| see above | NYU Langone Health | Departments of Pathology and Medicine, New York University School of Medicine | Adriana Heguy, Dacia Dimartino, Emily Guzman, Christian Marier, Peter Meyn, Sitharam Ramaswami, Gael Westby, Paul Zappile, Yutong Zhang, Paolo Cotzia, Guiqing Wang |
| EPI_ISL_1268198 | DPHL | Delaware Public Health Lab | Rebecca Savage |
| EPI_ISL_1273308, EPI_ISL_1273310, EPI_ISL_1273326, EPI_ISL_1273329, EPI_ISL_1273337, EPI_ISL_1273339, EPI_ISL_1273340, EPI_ISL_1273342, EPI_ISL_1273343, EPI_ISL_1273344, EPI_ISL_1273345, EPI_ISL_1273352, EPI_ISL_1273355, EPI_ISL_1273357, EPI_ISL_1273359, EPI_ISL_1273369, EPI_ISL_1273372, EPI_ISL_1273374, EPI_ISL_1273377, EPI_ISL_1273378 | | | |
| see above | The Jackson Laboratory | The Jackson Laboratory | Bergeron D, Renzette N, Adams M, Omerza G, Kelly K, Li L |
| EPI_ISL_1289652 | Michigan Department of Health and Human Services, Bureau of Laboratories | Michigan Department of Health and Human Services, Bureau of Laboratories | Blankenship HM, Riner D, Soehnlen MK |
| EPI_ISL_1292627 | State Laboratories Division, Hawaii State Department of Health | State Laboratories Division, Hawaii State Department of Health | Pamela O'Brien, Drew Kuwazaki, Ayana Garnet, Razvan Sultana, Edward Desmond |
| EPI_ISL_1292995, EPI_ISL_1292996 | Yale Clinical Virology Lab | Grubaugh Lab - Yale School of Public Health | Joseph Fauver, Mallery Breban, Isabell Ott, Tara Alpert, Mary Petrone, Anderson Brito, Chantal Vogels, Annie Watkins, Chaney Kalinich, Jessica Rothman, Marie L. Landry, Nathan Grubaugh |
| EPI_ISL_1293135, EPI_ISL_1293136, EPI_ISL_1293141 | Murphy Medical Association | Grubaugh Lab - Yale School of Public Health | Joseph Fauver, Mallery Breban, Isabell Ott, Tara Alpert, Mary Petrone, Anderson Brito, Chantal Vogels, Annie Watkins, Chaney Kalinich, Jessica Rothman, Caleb Neal, Eva Laszlo, Steven Murphy, Nathan Grubaugh |
| EPI_ISL_1293203 | Yale Clinical Virology Lab | Grubaugh Lab - Yale School of Public Health | Joseph Fauver, Mallery Breban, Isabell Ott, Tara Alpert, Mary Petrone, Anderson Brito, Chantal Vogels, Annie Watkins, Chaney Kalinich, Jessica Rothman, Marie L. Landry, Nathan Grubaugh |
| EPI_ISL_1300107 | Colorado Department of Public Health and Environment | Colorado Department of Public Health and Environment | Laura Bankers, Molly C. Hetherington-Rauth, Diana Ir, Shannon Ely, Shannon R. Matzinger, Sarah Elizabeth Totten, Emily A. Travanty |
| EPI_ISL_1300176, EPI_ISL_1300196, EPI_ISL_1300198, EPI_ISL_1300199, EPI_ISL_1300212, EPI_ISL_1300213, EPI_ISL_1300215, EPI_ISL_1300216, EPI_ISL_1300217, EPI_ISL_1300227, EPI_ISL_1300228, EPI_ISL_1300233, EPI_ISL_1300234, EPI_ISL_1300235, EPI_ISL_1300238, EPI_ISL_1300242, EPI_ISL_1300243, EPI_ISL_1300253, EPI_ISL_1300261, EPI_ISL_1300264, EPI_ISL_1300265, EPI_ISL_1300267, EPI_ISL_1300275 | | | |
| see above | NYU Langone Health | Departments of Pathology and Medicine, New York University School of Medicine | Adriana Heguy, Dacia Dimartino, Emily Guzman, Christian Marier, Peter Meyn, Sitharam Ramaswami, Gael Westby, Paul Zappile, Yutong Zhang, Paolo Cotzia, Guiqing Wang |
| EPI_ISL_1300757, EPI_ISL_1300758, EPI_ISL_1300818, EPI_ISL_1300820, EPI_ISL_1300843, EPI_ISL_1300875, EPI_ISL_1300877, EPI_ISL_1300879, EPI_ISL_1300883, EPI_ISL_1300899, EPI_ISL_1300903, EPI_ISL_1300906, EPI_ISL_1300911, EPI_ISL_1300917, EPI_ISL_1300920, EPI_ISL_1300925, EPI_ISL_1300926, EPI_ISL_1300928 | | | |
| see above | MSHS Clinical Microbiology Laboratories | MSHS Pathogen Surveillance Program | Ana S. Gonzalez-Reiche, Hala Alshammary, Mitchell J. Sullivan, Brianne Ciferri, Ajay Obla, Angela Amoako, Mahmoud Awawda, Daniel Floda, Julia Matthews, Ashley Salimbangon, Levy Sominsky, Katherine Beach, Kayla Russo, Charles Gleason, Shelcie Fabre, Giulio Kleiner, Zenab Khan, Bremy Alburquerque, Adriana van de Guchte, Komal Srivastava, Matthew M. Hernandez, Jayeeta Dutta, Denise Jurczyszak, Nancy Francoeur, Betsaida Salom Melo, Irina Oussenko, Gintaras Deikus, Juan Soto, Shwetha Hara Sridhar, Ying-Chih Wang, Kathryn Twyman, Deena R. Altman, Robert Sebra, Adolfo García-Sastre, Marta Luksza, Gopi Patel, Sarah Schaefer, Melissa Gitman, Michael D. Nowak, Alberto Paniz-Mondolfi, Emilia Mia Sordillo, Viviana Simon, Harm van Bakel |
| EPI_ISL_1302760 | Ministry of Health Turkey | Ministry of Health Turkey | Fatma Bayrakdar, Yasemin Cosgun, Suleyman Yalcin, Gulay Korukluoglu |
| EPI_ISL_1303415, EPI_ISL_1303443, EPI_ISL_1303462 | USC Clinical Lab | Los Angeles County PHL | P. Hemarajata et al. |
| EPI_ISL_1303484 | Flow Diagnostics Lab | Los Angeles County PHL | P. Hemarajata et al. |
| EPI_ISL_1303493 | USC Clinical Lab | Los Angeles County PHL | P. Hemarajata et al. |
| EPI_ISL_1305151 | Houston Methodist Hospital | Houston Methodist Hospital | S. Wesley Long, Randall J. Olsen, Paul A. Christensen, Sishir Subedi, Robert Olson, James J. Davis, Matthew Ojeda Saavedra, Prasanti Yerramilli, Layne Pruitt, Kristina Reppond, Madison N. Shyer, Jessica Cambric, Ilya J. Finkelstein, Jimmy Gollihar, and James M. Musser |
| EPI_ISL_1306254, EPI_ISL_1306256, EPI_ISL_1306274, EPI_ISL_1306283, EPI_ISL_1306286, EPI_ISL_1306296, EPI_ISL_1306299, EPI_ISL_1306307, EPI_ISL_1306312, EPI_ISL_1306315, EPI_ISL_1306316, EPI_ISL_1306331, EPI_ISL_1306332, EPI_ISL_1306333, EPI_ISL_1306335, EPI_ISL_1306347, EPI_ISL_1306353, EPI_ISL_1306364, EPI_ISL_1306373, EPI_ISL_1306379, EPI_ISL_1306381, EPI_ISL_1306382, EPI_ISL_1306388, EPI_ISL_1306391, EPI_ISL_1306394, EPI_ISL_1306395, EPI_ISL_1306398, EPI_ISL_1306412, EPI_ISL_1306419, EPI_ISL_1306422, EPI_ISL_1306426, EPI_ISL_1306429, EPI_ISL_1306436, EPI_ISL_1306438, EPI_ISL_1306440, EPI_ISL_1306453, EPI_ISL_1306457, EPI_ISL_1306458, EPI_ISL_1306471, EPI_ISL_1306474, EPI_ISL_1306477, EPI_ISL_1306478, EPI_ISL_1306485, EPI_ISL_1306492, EPI_ISL_1306493, EPI_ISL_1306508, EPI_ISL_1306513, EPI_ISL_1306518, EPI_ISL_1306528, EPI_ISL_1306539, EPI_ISL_1306545, EPI_ISL_1306546, EPI_ISL_1306549, EPI_ISL_1306550, EPI_ISL_1306553, EPI_ISL_1306558, EPI_ISL_1306568, EPI_ISL_1306575, EPI_ISL_1306580, EPI_ISL_1306587, EPI_ISL_1306590, EPI_ISL_1306607, EPI_ISL_1306616, EPI_ISL_1306617, EPI_ISL_1306626, EPI_ISL_1306627, EPI_ISL_1306628, EPI_ISL_1306634, EPI_ISL_1306640, EPI_ISL_1306644, EPI_ISL_1306648, EPI_ISL_1306661, EPI_ISL_1306662, EPI_ISL_1306667, EPI_ISL_1306679, EPI_ISL_1306680, EPI_ISL_1306695, EPI_ISL_1306698, EPI_ISL_1306701, EPI_ISL_1306703, EPI_ISL_1306705, EPI_ISL_1306712, EPI_ISL_1306724, EPI_ISL_1306735, EPI_ISL_1306746, | | | |

| | | | |
|---|---|---|---|
| EPI_ISL_1306769, EPI_ISL_1306778, EPI_ISL_1306782, EPI_ISL_1306785, EPI_ISL_1306792, EPI_ISL_1306802, EPI_ISL_1306818, EPI_ISL_1306832, EPI_ISL_1306839, EPI_ISL_1306840, EPI_ISL_1306846, EPI_ISL_1306854, EPI_ISL_1306858, EPI_ISL_1306863, EPI_ISL_1306866, EPI_ISL_1306870, EPI_ISL_1306874, EPI_ISL_1306879, EPI_ISL_1306880, EPI_ISL_1306884, EPI_ISL_1306895, EPI_ISL_1306898, EPI_ISL_1306899, EPI_ISL_1306913, EPI_ISL_1306924, EPI_ISL_1306926, EPI_ISL_1306936, EPI_ISL_1306944, EPI_ISL_1306948, EPI_ISL_1306952, EPI_ISL_1306971, EPI_ISL_1306972, EPI_ISL_1306979, EPI_ISL_1306985, EPI_ISL_1306987, EPI_ISL_1306988, EPI_ISL_1306992, EPI_ISL_1307001, EPI_ISL_1307003, EPI_ISL_1307004, EPI_ISL_1307006, EPI_ISL_1307008, EPI_ISL_1307010, EPI_ISL_1307014, EPI_ISL_1307022, EPI_ISL_1307023, EPI_ISL_1307024, EPI_ISL_1307025, EPI_ISL_1307027, EPI_ISL_1307033, EPI_ISL_1307035, EPI_ISL_1307038, EPI_ISL_1307039, EPI_ISL_1307053, EPI_ISL_1307060, EPI_ISL_1307062, EPI_ISL_1307064, EPI_ISL_1307077, EPI_ISL_1307081, EPI_ISL_1307085, EPI_ISL_1307086, EPI_ISL_1307087, EPI_ISL_1307088, EPI_ISL_1307090, EPI_ISL_1307095, EPI_ISL_1307097, EPI_ISL_1307099, EPI_ISL_1307102, EPI_ISL_1307103, EPI_ISL_1307104, EPI_ISL_1307109, EPI_ISL_1307111, EPI_ISL_1307115, EPI_ISL_1307116, EPI_ISL_1307117, EPI_ISL_1307120, EPI_ISL_1307122, EPI_ISL_1307124, EPI_ISL_1307125, EPI_ISL_1307130, EPI_ISL_1307131, EPI_ISL_1307133, EPI_ISL_1307136, EPI_ISL_1307146, EPI_ISL_1307149, EPI_ISL_1307152, EPI_ISL_1307154, EPI_ISL_1307160, EPI_ISL_1307162, EPI_ISL_1307171, EPI_ISL_1307176, EPI_ISL_1307180, EPI_ISL_1307181, EPI_ISL_1307182, EPI_ISL_1307183, EPI_ISL_1307185, EPI_ISL_1307188, EPI_ISL_1307197, EPI_ISL_1307204, EPI_ISL_1307205, EPI_ISL_1307221, EPI_ISL_1307234, EPI_ISL_1307240, EPI_ISL_1307252, EPI_ISL_1307258, EPI_ISL_1307271, EPI_ISL_1307273, EPI_ISL_1307274, EPI_ISL_1307280, EPI_ISL_1307284, EPI_ISL_1307285, EPI_ISL_1307289, EPI_ISL_1307293, EPI_ISL_1307297, EPI_ISL_1307301, EPI_ISL_1307310, EPI_ISL_1307314, EPI_ISL_1307319, EPI_ISL_1307320, EPI_ISL_1307324, EPI_ISL_1307325, EPI_ISL_1307326, EPI_ISL_1307336, EPI_ISL_1307340, EPI_ISL_1307342, EPI_ISL_1307343, EPI_ISL_1307349, EPI_ISL_1307361, EPI_ISL_1307373, EPI_ISL_1307374, EPI_ISL_1307380, EPI_ISL_1307384, EPI_ISL_1307387, EPI_ISL_1307388, EPI_ISL_1307389, EPI_ISL_1307392, EPI_ISL_1307399, EPI_ISL_1307407, EPI_ISL_1307408, EPI_ISL_1307409, EPI_ISL_1307414, EPI_ISL_1307424, EPI_ISL_1307425, EPI_ISL_1307429, EPI_ISL_1307435, EPI_ISL_1307453, EPI_ISL_1307457, EPI_ISL_1307470, EPI_ISL_1307473, EPI_ISL_1307474, EPI_ISL_1307475, EPI_ISL_1307493, EPI_ISL_1307495, EPI_ISL_1307501, EPI_ISL_1307503, EPI_ISL_1307505, EPI_ISL_1307510, EPI_ISL_1307513, EPI_ISL_1307515, EPI_ISL_1307516, EPI_ISL_1307519, EPI_ISL_1307524, EPI_ISL_1307533, EPI_ISL_1307536, EPI_ISL_1307541, EPI_ISL_1307542, EPI_ISL_1307543, EPI_ISL_1307544, EPI_ISL_1307546, EPI_ISL_1307556, EPI_ISL_1307557, EPI_ISL_1307558, EPI_ISL_1307559, EPI_ISL_1307561, EPI_ISL_1307568, EPI_ISL_1307569, EPI_ISL_1307578, EPI_ISL_1307579, EPI_ISL_1307580, EPI_ISL_1307587, EPI_ISL_1307590, EPI_ISL_1307591, EPI_ISL_1307593, EPI_ISL_1307603, EPI_ISL_1307611, EPI_ISL_1307615, EPI_ISL_1307627, EPI_ISL_1307632 | | | |
| see above | Pandemic Response Lab - NYC | Pandemic Response Lab, R&D | Henry Lee, Michael Hammerling, Melissa Hopkins, Cybill del Castillo, Shinyoung Clair Kang, William Ward, Pradeep Bugga, Sol Rey, Dylan Law, Haiping Hao, Jon Laurent |
| EPI_ISL_1311965 | The Ohio State University Applied Microbiology Services Laboratory | The Ohio State University Applied Microbiology Services Laboratory | Seth A. Faith PhD |
| EPI_ISL_1312775 | Molecular & Genomic Pathology Laboratory, Thomas Jefferson University Hospital | Molecular & Genomic Pathology Laboratory, Thomas Jefferson University Hospital | Run Jin, Nitika Badjatia, Zi-xuan Wang |
| EPI_ISL_1313545, EPI_ISL_1313547, EPI_ISL_1313549 | NJDOH, Public Health and Environmental Laboratories | NJ_PHEL | Lindsey Bodnar, Shiv K. Verma, Dana Woell, Byeong Jeong |
| EPI_ISL_1313718 | Utah Public Health Laboratory | Utah Public Health Laboratory | Erin L. Young, Kelly F. Oakeson, Tara Gallagher |
| EPI_ISL_1314086 | University of Wisconsin-Madison AIDS Vaccine Research Laboratories | University of Wisconsin-Madison AIDS Vaccine Research Laboratories | Gage Moreno, Katarina Braun, et al. AIDS Vaccine Research Laboratories |
| EPI_ISL_1314316 | Mayo Clinic & Mayo Clinic Laboratories | Minnesota Department of Health, Public Health Laboratory | Alexandra Lorentz, Jacob Garfin, Matt Plumb, and Xiong Wang |
| EPI_ISL_1315130, EPI_ISL_1315131, EPI_ISL_1315134, EPI_ISL_1315138, EPI_ISL_1315141, EPI_ISL_1315142, EPI_ISL_1315143, EPI_ISL_1315144, EPI_ISL_1315145, EPI_ISL_1315166, EPI_ISL_1315172, EPI_ISL_1315178, EPI_ISL_1315179, EPI_ISL_1315186, EPI_ISL_1315189, EPI_ISL_1315190, EPI_ISL_1315193, EPI_ISL_1315196, EPI_ISL_1315197, EPI_ISL_1315202, EPI_ISL_1315203, EPI_ISL_1315207, EPI_ISL_1315208, EPI_ISL_1315209, EPI_ISL_1315215, EPI_ISL_1315217, EPI_ISL_1315218 | | | |
| see above | The Jackson Laboratory | The Jackson Laboratory | Bergeron D, Renzette N, Adams M, Omerza G, Kelly K, Li L |
| EPI_ISL_1319050, EPI_ISL_1319169, EPI_ISL_1319502, EPI_ISL_1319648, EPI_ISL_1320399, EPI_ISL_1320698, EPI_ISL_1320751, EPI_ISL_1320893 | Laboratory Corporation of America | Centers for Disease Control and Prevention Division of Viral Diseases, Pathogen Discovery | Peter W. Cook, Dakota Howard, Dhwani Batra, Ben L. Rambo-Martin, Minoo Agarwal, Eyad Almasri, Debbie Boles, Ayla Burns, Nuthawin Charoensri, Oren Cohen, Susan Countryman, Mary Ann Cristobal, Bobbi Croy, Suzanne Dale, Hrushikesh Deshmukh, Amanda Douglas, Vincent Drouillon, Marcia Eisenberg, Howard Engler, Rama Ghatti, Prashant Gupta, Susan Hicks, Jake Humphrey, Lax Iyer, Manoj Jain, Mohan Kolli, Brian Krueger, Tim Kuphal, Stanley Letovsky, Michael Levandoski, Craig Lukasik, Jonathan Meltzer, Brian Norvell, Mindy Nye, Scott Parker, Christos Petropoulos, John Pruitt, Steven Ragan, Scott Ryan, Mike Sapeta, Jana Schroth, Suresh Babu Selvaraju, Goran Stevovic, Amanda Suchanek, Andrea Throop, Lyndon Tilson, Thomas Urban, Joe Voshell, Kimberly Wagner, Jonathan Williams, Mary Williamson, Qian Zeng, Tricia Zwiefelhofer, Clinton R. Paden, Suxiang Tong, Duncan MacCannell |
| EPI_ISL_1321493 | Genetica Molecular and Subdepartamento de Virologia ISP Chile | Instituto de Salud Publica de Chile | Javier Tognarelli, Karen Orostica, Barbara Parra, Loredana Arata, Jaime Lagos, Gisselle Barra, Patricia Bustos, Rodrigo Fasce, Andres Castillo, Jorge Fernandez |
| EPI_ISL_1322716 | KU Leuven, Rega Institute, Clinical and Epidemiological Virology | KU Leuven, Rega Institute, Clinical and Epidemiological Virology | Tony Wawina-Bokalanga, Bert Vanmechelen, Joan Marti-Carerras, Piet Maes |
| EPI_ISL_1322748, EPI_ISL_1322767, EPI_ISL_1322782, EPI_ISL_1322785, EPI_ISL_1322786, EPI_ISL_1322795, EPI_ISL_1322801, EPI_ISL_1322803, EPI_ISL_1322808, EPI_ISL_1322813, EPI_ISL_1322814, EPI_ISL_1322822, EPI_ISL_1322823, EPI_ISL_1322824, EPI_ISL_1322827, EPI_ISL_1322829 | | | |
| see above | NYU Langone Health | Departments of Pathology and Medicine, New York University School of Medicine | Adriana Heguy, Dacia Dimartino, Emily Guzman, Christian Marier, Peter Meyn, Sitharam Ramaswami, Gael Westby, Paul Zappile, Yutong Zhang, Paolo Cotzia, Guiqing Wang |
| EPI_ISL_1332719 | Lighthouse Lab in Cambridge | Wellcome Sanger Institute for the COVID-19 Genomics UK (COG-UK) Consortium | Rob Howes, The Lighthouse Lab in Cambridge and Alex Alderton, Roberto Amato, Jeffrey Barrett, Sonia Goncalves, Ewan Harrison, David K. Jackson, Ian Johnston, Dominic Kwiatkowski, Cordelia Langford, John Sillitoe on behalf of the Wellcome Sanger Institute COVID-19 Surveillance Team |
| EPI_ISL_1337464 | DOHMH Chelsea | New York City Public Health Laboratory | Jade Wang, et al. |
| EPI_ISL_1337468 | DOHMH Fort Greene | New York City Public Health Laboratory | Jade Wang, et al. |
| EPI_ISL_1337475 | DOHMH Corona | New York City Public Health Laboratory | Jade Wang, et al. |
| EPI_ISL_1337482 | OCME Office Of Chief Medical Examiner | New York City Public Health Laboratory | Jade Wang, et al. |
| EPI_ISL_1337513 | DOHMH Jamaica | New York City Public Health Laboratory | Jade Wang, et al. |
| EPI_ISL_1337514, EPI_ISL_1337515 | DOHMH Corona | New York City Public Health Laboratory | Jade Wang, et al. |
| EPI_ISL_1337520 | DOHMH Central Harlem | New York City Public Health Laboratory | Jade Wang, et al. |
| EPI_ISL_1337527 | OCME Office Of Chief Medical Examiner | New York City Public Health Laboratory | Jade Wang, et al. |
| EPI_ISL_1337528 | DOHMH Crown Heights | New York City Public Health Laboratory | Jade Wang, et al. |
| EPI_ISL_1337530 | DOHMH PHL | New York City Public Health Laboratory | Jade Wang, et al. |
| EPI_ISL_1337534, EPI_ISL_1337535 | DOHMH Crown Heights | New York City Public Health Laboratory | Jade Wang, et al. |
| EPI_ISL_1337537, EPI_ISL_1337538 | DOHMH Jamaica | New York City Public Health Laboratory | Jade Wang, et al. |
| EPI_ISL_1337540 | DOHMH Morrisania | New York City Public Health Laboratory | Jade Wang, et al. |
| EPI_ISL_1338534, EPI_ISL_1338632, EPI_ISL_1338661, EPI_ISL_1338672, EPI_ISL_1338686, EPI_ISL_1338705, EPI_ISL_1338723, EPI_ISL_1339238, EPI_ISL_1339633, EPI_ISL_1339748 | Laboratory Corporation of America | Centers for Disease Control and Prevention Division of Viral Diseases, Pathogen Discovery | Peter W. Cook, Dakota Howard, Dhwani Batra, Ben L. Rambo-Martin, Minoo Agarwal, Eyad Almasri, Debbie Boles, Ayla Burns, Nuthawin Charoensri, Oren Cohen, Susan Countryman, Mary Ann Cristobal, Bobbi Croy, Suzanne Dale, Hrushikesh Deshmukh, Amanda Douglas, Vincent Drouillon, Marcia Eisenberg, Howard Engler, Rama Ghatti, Prashant Gupta, Susan Hicks, Jake Humphrey, Lax Iyer, Manoj Jain, Mohan Kolli, Brian Krueger, Tim Kuphal, Stanley Letovsky, Michael Levandoski, Craig Lukasik, Jonathan Meltzer, Brian Norvell, Mindy Nye, Scott Parker, Christos Petropoulos, John Pruitt, Steven Ragan, Scott Ryan, Mike Sapeta, Jana Schroth, Suresh Babu Selvaraju, Goran Stevovic, Amanda Suchanek, Andrea Throop, Lyndon Tilson, Thomas Urban, Joe Voshell, Kimberly Wagner, Jonathan Williams, Mary Williamson, Qian Zeng, Tricia Zwiefelhofer, Clinton R. Paden, Suxiang Tong, Duncan MacCannell |
| EPI_ISL_1340173, EPI_ISL_1340175, EPI_ISL_1340190, EPI_ISL_1340202, EPI_ISL_1340204, EPI_ISL_1340207 | NJDOH, Public Health and Environmental Laboratories | New Jersey Public Health and Environmental Laboratories (PHEL) | Lindsey Bodnar, Shiv K. Verma, Dana Woell, Byeong Jeong |
| EPI_ISL_1340884 | New Mexico Department of Health Scientific Laboratory | New Mexico Department of Health Scientific Laboratory | Ellie Johnson, Anastacia Griego-Fisher, D'eldra Malone, Jennifer Benoit |
| EPI_ISL_1364525 | Stanford Health Care | Stanford University School of Medicine, Clinical Virology Laboratory | Daniel Solis, Mamdouh Sibai, Fumiko Yamamoto, Malaya K. Sahoo, Michelle Verghese, ChunHong Huang, James Zehnder, and Benjamin A. Pinsky |
| EPI_ISL_1365482 | Arizona State University | Arizona State University | Peter T. Skidmore, LaRinda A. Holland, Rabia Maqsood, Emily A. Kaelin, Nicholas J. Mellor, Kristina Buss, Joy M. Blain, Neal W Woodbury, Valerie Harris, Joshua LaBaer, Vel Murugan, Efrem S. Lim |

| | | | |
|---|---|---|---|
| EPI_ISL_1365717 | Virginia Division of Consolidated Laboratory Services | Virginia Division of Consolidated Laboratory Services | Virginia DCLS |
| EPI_ISL_1367826 | Michigan Department of Health and Human Services, Bureau of Laboratories | Michigan Department of Health and Human Services, Bureau of Laboratories | Blankenship HM, Riner D, Soehnlen MK |
| EPI_ISL_1369590 | University Hospitals of Geneva, Laboratory of Virology | HUG, Laboratory of Virology and the Health2030 Genome Center | Samuel Cordey, Ana Rita Goncalves, Laurent Kaiser, Lorenzo Cerutti, Henri Pegeot, Melyssa Elies, Deborah Penet, Keith Harshman, Ioannis Xenarios, Emmanouil Dermitzakis |
| EPI_ISL_1373032 | Wyoming Public Health Laboratory | Wyoming Public Health Laboratory | Jim Mildenberger, Wanda Manley, Noah Hull, Taylor Fearing, Lynette Gumbleton, Channing Weber, Ashley Norberg, Chayse Rowley, Marley Goetz, Brian Dominguez, Elliot Thomasson, Cari Sloma, and Rob Christensen |
| EPI_ISL_1378711, EPI_ISL_1378720, EPI_ISL_1378724 | Yale Clinical Virology Lab | Grubaugh Lab - Yale School of Public Health | Joseph Fauver, Mallery Breban, Isabell Ott, Tara Alpert, Mary Petrone, Anderson Brito, Chantal Vogels, Annie Watkins, Chaney Kalinich, Jessica Rothman, Marie L. Landry, Nathan Grubaugh |
| EPI_ISL_1378726 | Grubaugh Lab - Yale School of Public Health | Grubaugh Lab - Yale School of Public Health | Joseph Fauver, Mallery Breban, Isabell Ott, Tara Alpert, Mary Petrone, Anderson Brito, Chantal Vogels, Annie Watkins, Chaney Kalinich, Jessica Rothman, Nathan Grubaugh |
| EPI_ISL_1378728, EPI_ISL_1378730, EPI_ISL_1378736, EPI_ISL_1378738, EPI_ISL_1378741, EPI_ISL_1378744, EPI_ISL_1378745, EPI_ISL_1378746, EPI_ISL_1378748, EPI_ISL_1378752 | Yale Clinical Virology Lab | Grubaugh Lab - Yale School of Public Health | Joseph Fauver, Mallery Breban, Isabell Ott, Tara Alpert, Mary Petrone, Anderson Brito, Chantal Vogels, Annie Watkins, Chaney Kalinich, Jessica Rothman, Marie L. Landry, Nathan Grubaugh |
| EPI_ISL_1378753 | Grubaugh Lab - Yale School of Public Health | Grubaugh Lab - Yale School of Public Health | Joseph Fauver, Mallery Breban, Isabell Ott, Tara Alpert, Mary Petrone, Anderson Brito, Chantal Vogels, Annie Watkins, Chaney Kalinich, Jessica Rothman, Nathan Grubaugh |
| EPI_ISL_1378762, EPI_ISL_1378764 | Yale Clinical Virology Lab | Grubaugh Lab - Yale School of Public Health | Joseph Fauver, Mallery Breban, Isabell Ott, Tara Alpert, Mary Petrone, Anderson Brito, Chantal Vogels, Annie Watkins, Chaney Kalinich, Jessica Rothman, Marie L. Landry, Nathan Grubaugh |
| EPI_ISL_1378768, EPI_ISL_1378790, EPI_ISL_1378799, EPI_ISL_1378809 | Tempus | Grubaugh Lab - Yale School of Public Health | Joseph Fauver, Mallery Breban, Isabell Ott, Tara Alpert, Mary Petrone, Anderson Brito, Chantal Vogels, Annie Watkins, Chaney Kalinich, Jessica Rothman, Matthew J. MacKay, Gaurav Khullar, Jessica Metti, Joel T. Dudley, Megan Nash, Nike Beaubier, Christopher E. Mason, Nathan Grubaugh |
| EPI_ISL_1378825, EPI_ISL_1378826 | Murphy Medical Association | Grubaugh Lab - Yale School of Public Health | Joseph Fauver, Mallery Breban, Isabell Ott, Tara Alpert, Mary Petrone, Anderson Brito, Chantal Vogels, Annie Watkins, Chaney Kalinich, Jessica Rothman, Caleb Neal, Eva Laszlo, Steven Murphy, Nathan Grubaugh |
| EPI_ISL_1379220, EPI_ISL_1379221, EPI_ISL_1379224, EPI_ISL_1379226, EPI_ISL_1379231, EPI_ISL_1379236, EPI_ISL_1379240, EPI_ISL_1379247, EPI_ISL_1379255, EPI_ISL_1379262 | NYP-WCM | New York Genome Center | Michael Zody, Andre Corvelo, Dayna M. Oschwald, Samantha Fennessey, Tom Maniatis, Melissa Cushing, Olivier Elemento, Margaret Elizabeth Ross, Chris Mason, Priya Velu, Hanna Rennert, Arryn Craney, Lars F Westblade |
| EPI_ISL_1379268, EPI_ISL_1379283, EPI_ISL_1379303, EPI_ISL_1379304, EPI_ISL_1379325, EPI_ISL_1379337 | Hackensack Meridian Health | New York Genome Center | Michael Zody, Andre Corvelo, Dayna M. Oschwald, Samantha Fennessey, Tom Maniatis, Liang Chen, Jose Mediavilla, Marcus Cunningham, Kaelea Composto, Kar Chow, David Perlin, Barry Kreiswirth |
| EPI_ISL_1379342, EPI_ISL_1379343, EPI_ISL_1379344, EPI_ISL_1379345, EPI_ISL_1379358, EPI_ISL_1379374, EPI_ISL_1379382, EPI_ISL_1379384, EPI_ISL_1379386 | NYP-WCM | New York Genome Center | Michael Zody, Andre Corvelo, Dayna M. Oschwald, Samantha Fennessey, Tom Maniatis, Melissa Cushing, Olivier Elemento, Margaret Elizabeth Ross, Chris Mason, Priya Velu, Hanna Rennert, Arryn Craney, Lars F Westblade |
| EPI_ISL_1379974 | UCLA Clinical Micro Lab | Los Angeles County PHL | P. Hemarajata et al. |
| EPI_ISL_1381304, EPI_ISL_1381305 | Omics Sciences Laboratory | Omics Sciences Laboratory | Derly Andrade Molina, Rubén Armas González, Gabriel Morey León, Darlyn Amaya, Katheryn Sacheri Viteri, Emily Sulay Saltos Montalvo, Paula Juliana Gavilanes Jarrín, Juan Carlos Fernández Cadena |
| EPI_ISL_1384639 | Anteja laboratorija (UAB Diagnostikos laboratorija) | Vilnius University Hospital Santaros Klinikos, Center of Laboratory Medicine | Dovile Ezerskyte, Daniel Naumovas, Gytis Dudas, Ingrida Olendraite, Rimvydas Norvilas, Ligita Raugaite, Monika Katenaite, Mindaugas Stoskus, Laimonas Griskevicius |
| EPI_ISL_1384891, EPI_ISL_1384892, EPI_ISL_1384904, EPI_ISL_1384905, EPI_ISL_1384907, EPI_ISL_1384908, EPI_ISL_1384909, EPI_ISL_1384916, EPI_ISL_1384927, EPI_ISL_1384929, EPI_ISL_1384937, EPI_ISL_1384938, EPI_ISL_1384939, EPI_ISL_1384940, EPI_ISL_1384942, EPI_ISL_1384944, EPI_ISL_1384945, EPI_ISL_1384958, EPI_ISL_1384960, EPI_ISL_1384962, EPI_ISL_1384966, EPI_ISL_1384978, EPI_ISL_1384985, EPI_ISL_1384998, EPI_ISL_1385003, EPI_ISL_1385006, EPI_ISL_1385008, EPI_ISL_1385010, EPI_ISL_1385011, EPI_ISL_1385017, EPI_ISL_1385018, EPI_ISL_1385029, EPI_ISL_1385041, EPI_ISL_1385043, EPI_ISL_1385047, EPI_ISL_1385056, EPI_ISL_1385067, EPI_ISL_1385070, EPI_ISL_1385078, EPI_ISL_1385087, EPI_ISL_1385089, EPI_ISL_1385091, EPI_ISL_1385095, EPI_ISL_1385102, EPI_ISL_1385103, EPI_ISL_1385105, EPI_ISL_1385106, EPI_ISL_1385115, EPI_ISL_1385116, EPI_ISL_1385118, EPI_ISL_1385119, EPI_ISL_1385120, EPI_ISL_1385121, EPI_ISL_1385123, EPI_ISL_1385125, EPI_ISL_1385128, EPI_ISL_1385130, EPI_ISL_1385139, EPI_ISL_1385143, EPI_ISL_1385150, EPI_ISL_1385152, EPI_ISL_1385156, EPI_ISL_1385161, EPI_ISL_1385162, EPI_ISL_1385163, EPI_ISL_1385166, EPI_ISL_1385171, EPI_ISL_1385177, EPI_ISL_1385179, EPI_ISL_1385185, EPI_ISL_1385188, EPI_ISL_1385189, EPI_ISL_1385192, EPI_ISL_1385193, EPI_ISL_1385196, EPI_ISL_1385200, EPI_ISL_1385202, EPI_ISL_1385203, EPI_ISL_1385206, EPI_ISL_1385212, EPI_ISL_1385219, EPI_ISL_1385221, EPI_ISL_1385231, EPI_ISL_1385232, EPI_ISL_1385236, EPI_ISL_1385239, EPI_ISL_1385242, EPI_ISL_1385243, EPI_ISL_1385244, EPI_ISL_1385245, EPI_ISL_1385249, EPI_ISL_1385255, EPI_ISL_1385257, EPI_ISL_1385266, EPI_ISL_1385273, EPI_ISL_1385276, EPI_ISL_1385279, EPI_ISL_1385291, EPI_ISL_1385298, EPI_ISL_1385310, EPI_ISL_1385322, EPI_ISL_1385329, EPI_ISL_1385333, EPI_ISL_1385343, EPI_ISL_1385347, EPI_ISL_1385351, EPI_ISL_1385356, EPI_ISL_1385361, EPI_ISL_1385363, EPI_ISL_1385364, EPI_ISL_1385366, EPI_ISL_1385370, EPI_ISL_1385375, EPI_ISL_1385376, EPI_ISL_1385393, EPI_ISL_1385394, EPI_ISL_1385395, EPI_ISL_1385401, EPI_ISL_1385402, EPI_ISL_1385403, EPI_ISL_1385413, EPI_ISL_1385414, EPI_ISL_1385415, EPI_ISL_1385416, EPI_ISL_1385420, EPI_ISL_1385425, EPI_ISL_1385434, EPI_ISL_1385436, EPI_ISL_1385456, EPI_ISL_1385457, EPI_ISL_1385473, EPI_ISL_1385481, EPI_ISL_1385492, EPI_ISL_1385495, EPI_ISL_1385500, EPI_ISL_1385504, EPI_ISL_1385514, EPI_ISL_1385517, EPI_ISL_1385521, EPI_ISL_1385531, EPI_ISL_1385535, EPI_ISL_1385547, EPI_ISL_1385550, EPI_ISL_1385552, EPI_ISL_1385554, EPI_ISL_1385560, EPI_ISL_1385562, EPI_ISL_1385564, EPI_ISL_1385577, EPI_ISL_1385579, EPI_ISL_1385584, EPI_ISL_1385589, EPI_ISL_1385591, EPI_ISL_1385592, EPI_ISL_1385597, EPI_ISL_1385603, EPI_ISL_1385606, EPI_ISL_1385616, EPI_ISL_1385617, EPI_ISL_1385625, EPI_ISL_1385628, EPI_ISL_1385640, EPI_ISL_1385647, EPI_ISL_1385652, EPI_ISL_1385654, EPI_ISL_1385660, EPI_ISL_1385665, EPI_ISL_1385667, EPI_ISL_1385674, EPI_ISL_1385679, EPI_ISL_1385687, EPI_ISL_1385695, EPI_ISL_1385696, EPI_ISL_1385698, EPI_ISL_1385702, EPI_ISL_1385703, EPI_ISL_1385709, EPI_ISL_1385712, EPI_ISL_1385728, EPI_ISL_1385731, EPI_ISL_1385742, EPI_ISL_1385743, EPI_ISL_1385746, EPI_ISL_1385754, EPI_ISL_1385760, EPI_ISL_1385761, EPI_ISL_1385776, EPI_ISL_1385780 | | | |
| see above | Pandemic Response Lab - NYC | Pandemic Response Lab, R&D | Henry Lee, Michael Hammerling, Melissa Hopkins, Cybill del Castillo, Shinyoung Clair Kang, William Ward, Pradeep Bugga, Sol Rey, Dylan Law, Haiping Hao, Jon Laurent |
| EPI_ISL_1395744, EPI_ISL_1395747, EPI_ISL_1395748 | Columbia University Irving Medical Center | Wadsworth Center, New York State Department of Health | Kirsten St. George, Daryl M. Lamson, Alexis Russel, Matthew Shudt, Melissa A Leisner, Jonathan Plitnick, Navjot Singh, John Kelly, Erasmus Schneider, Erica Lasek-Nesselquist |
| EPI_ISL_1395750 | NORTH SHORE UNIVERSITY HOSPITAL | Wadsworth Center, New York State Department of Health | Kirsten St. George, Daryl M. Lamson, Alexis Russel, Matthew Shudt, Melissa A Leisner, Jonathan Plitnick, Navjot Singh, John Kelly, Erasmus Schneider, Erica Lasek-Nesselquist |
| EPI_ISL_1395755, EPI_ISL_1395759 | SUNY UPSTATE MEDICAL UNIVERSITY | Wadsworth Center, New York State Department of Health | Kirsten St. George, Daryl M. Lamson, Alexis Russel, Matthew Shudt, Melissa A Leisner, Jonathan Plitnick, Navjot Singh, John Kelly, Erasmus Schneider, Erica Lasek-Nesselquist |
| EPI_ISL_1395760 | NORTH SHORE UNIVERSITY HOSPITAL | Wadsworth Center, New York State Department of Health | Kirsten St. George, Daryl M. Lamson, Alexis Russel, Matthew Shudt, Melissa A Leisner, Jonathan Plitnick, Navjot Singh, John Kelly, Erasmus Schneider, Erica Lasek-Nesselquist |
| EPI_ISL_1395770, EPI_ISL_1395776 | SUNY UPSTATE MEDICAL UNIVERSITY | Wadsworth Center, New York State Department of Health | Kirsten St. George, Daryl M. Lamson, Alexis Russel, Matthew Shudt, Melissa A Leisner, Jonathan Plitnick, Navjot Singh, John Kelly, Erasmus Schneider, Erica Lasek-Nesselquist |
| EPI_ISL_1397515, EPI_ISL_1397528 | New York Presbyterian Hospital | Wadsworth Center, New York State Department of Health | Kirsten St. George, Daryl M. Lamson, Alexis Russel, Matthew Shudt, Melissa A Leisner, Jonathan Plitnick, Navjot Singh, John Kelly, Erasmus Schneider, Erica Lasek-Nesselquist |
| EPI_ISL_1397593, EPI_ISL_1397595, EPI_ISL_1397597, EPI_ISL_1397611 | SUNY UPSTATE MEDICAL UNIVERSITY | Wadsworth Center, New York State Department of Health | Kirsten St. George, Daryl M. Lamson, Alexis Russel, Matthew Shudt, Melissa A Leisner, Jonathan Plitnick, Navjot Singh, John Kelly, Erasmus Schneider, Erica Lasek-Nesselquist |
| EPI_ISL_1397618, EPI_ISL_1397620 | MONTEFIORE MEDICAL CENTER LABORATORIES | Wadsworth Center, New York State Department of Health | Kirsten St. George, Daryl M. Lamson, Alexis Russel, Matthew Shudt, Melissa A Leisner, Jonathan Plitnick, Navjot Singh, John Kelly, Erasmus Schneider, Erica Lasek-Nesselquist |
| EPI_ISL_1397636, EPI_ISL_1397638, EPI_ISL_1397639 | Columbia University Irving Medical Center | Wadsworth Center, New York State Department of Health | Kirsten St. George, Daryl M. Lamson, Alexis Russel, Matthew Shudt, Melissa A Leisner, Jonathan Plitnick, Navjot Singh, John Kelly, Erasmus Schneider, Erica Lasek-Nesselquist |

| | | | |
|---|---|---|---|
| EPI_ISL_1397651, EPI_ISL_1397656, EPI_ISL_1397666, EPI_ISL_1397668, EPI_ISL_1397677, EPI_ISL_1397683, EPI_ISL_1397687 | NORTH SHORE UNIVERSITY HOSPITAL | Wadsworth Center, New York State Department of Health | Kirsten St. George, Daryl M. Lamson, Alexis Russel, Matthew Shudt, Melissa A Leisner, Jonathan Plitnick, Navjot Singh, John Kelly, Erasmus Schneider, Erica Lasek-Nesselquist |
| EPI_ISL_1397749 | ADIRONDACK MEDICAL CENTER | Wadsworth Center, New York State Department of Health | Kirsten St. George, Daryl M. Lamson, Alexis Russel, Matthew Shudt, Melissa A Leisner, Jonathan Plitnick, Navjot Singh, John Kelly, Erasmus Schneider, Erica Lasek-Nesselquist |
| EPI_ISL_1397762 | ALBANY MEDICAL CENTER | Wadsworth Center, New York State Department of Health | Kirsten St. George, Daryl M. Lamson, Alexis Russel, Matthew Shudt, Melissa A Leisner, Jonathan Plitnick, Navjot Singh, John Kelly, Erasmus Schneider, Erica Lasek-Nesselquist |
| EPI_ISL_1397793, EPI_ISL_1397805, EPI_ISL_1397806, EPI_ISL_1397807 | MONTEFIORE MEDICAL CENTER LABORATORIES | Wadsworth Center, New York State Department of Health | Kirsten St. George, Daryl M. Lamson, Alexis Russel, Matthew Shudt, Melissa A Leisner, Jonathan Plitnick, Navjot Singh, John Kelly, Erasmus Schneider, Erica Lasek-Nesselquist |
| EPI_ISL_1397824 | Columbia University Irving Medical Center | Wadsworth Center, New York State Department of Health | Kirsten St. George, Daryl M. Lamson, Alexis Russel, Matthew Shudt, Melissa A Leisner, Jonathan Plitnick, Navjot Singh, John Kelly, Erasmus Schneider, Erica Lasek-Nesselquist |
| EPI_ISL_1397831 | STONY BROOK UNIVERSITY HOSPITAL | Wadsworth Center, New York State Department of Health | Kirsten St. George, Daryl M. Lamson, Alexis Russel, Matthew Shudt, Melissa A Leisner, Jonathan Plitnick, Navjot Singh, John Kelly, Erasmus Schneider, Erica Lasek-Nesselquist |
| EPI_ISL_1397867 | ALBANY MEDICAL CENTER | Wadsworth Center, New York State Department of Health | Kirsten St. George, Daryl M. Lamson, Alexis Russel, Matthew Shudt, Melissa A Leisner, Jonathan Plitnick, Navjot Singh, John Kelly, Erasmus Schneider, Erica Lasek-Nesselquist |
| EPI_ISL_1397873, EPI_ISL_1397881, EPI_ISL_1397882 | MONTEFIORE MEDICAL CENTER LABORATORIES | Wadsworth Center, New York State Department of Health | Kirsten St. George, Daryl M. Lamson, Alexis Russel, Matthew Shudt, Melissa A Leisner, Jonathan Plitnick, Navjot Singh, John Kelly, Erasmus Schneider, Erica Lasek-Nesselquist |
| EPI_ISL_1397888, EPI_ISL_1397897, EPI_ISL_1397900, EPI_ISL_1397904 | Columbia University Irving Medical Center | Wadsworth Center, New York State Department of Health | Kirsten St. George, Daryl M. Lamson, Alexis Russel, Matthew Shudt, Melissa A Leisner, Jonathan Plitnick, Navjot Singh, John Kelly, Erasmus Schneider, Erica Lasek-Nesselquist |
| EPI_ISL_1397909 | NORTHWELL HEALTH LABORATORIES | Wadsworth Center, New York State Department of Health | Kirsten St. George, Daryl M. Lamson, Alexis Russel, Matthew Shudt, Melissa A Leisner, Jonathan Plitnick, Navjot Singh, John Kelly, Erasmus Schneider, Erica Lasek-Nesselquist |
| EPI_ISL_1397928, EPI_ISL_1397937, EPI_ISL_1397939, EPI_ISL_1397942, EPI_ISL_1397948 | WESTCHESTER MEDICAL CENTER | Wadsworth Center, New York State Department of Health | Kirsten St. George, Daryl M. Lamson, Alexis Russel, Matthew Shudt, Melissa A Leisner, Jonathan Plitnick, Navjot Singh, John Kelly, Erasmus Schneider, Erica Lasek-Nesselquist |
| EPI_ISL_1398023 | ALBANY MEDICAL CENTER | Wadsworth Center, New York State Department of Health | Kirsten St. George, Daryl M. Lamson, Alexis Russel, Matthew Shudt, Melissa A Leisner, Jonathan Plitnick, Navjot Singh, John Kelly, Erasmus Schneider, Erica Lasek-Nesselquist |
| EPI_ISL_1398036, EPI_ISL_1398048, EPI_ISL_1398049, EPI_ISL_1398050, EPI_ISL_1398052, EPI_ISL_1398057, EPI_ISL_1398064, EPI_ISL_1398065, EPI_ISL_1398066 | MONTEFIORE MEDICAL CENTER LABORATORIES | Wadsworth Center, New York State Department of Health | Kirsten St. George, Daryl M. Lamson, Alexis Russel, Matthew Shudt, Melissa A Leisner, Jonathan Plitnick, Navjot Singh, John Kelly, Erasmus Schneider, Erica Lasek-Nesselquist |
| EPI_ISL_1398068, EPI_ISL_1398070, EPI_ISL_1398072, EPI_ISL_1398074, EPI_ISL_1398080, EPI_ISL_1398081, EPI_ISL_1398082, EPI_ISL_1398084, EPI_ISL_1398086, EPI_ISL_1398095, EPI_ISL_1398112, EPI_ISL_1398119 | | | |
| see above | NORTHWELL HEALTH LABORATORIES | Wadsworth Center, New York State Department of Health | Kirsten St. George, Daryl M. Lamson, Alexis Russel, Matthew Shudt, Melissa A Leisner, Jonathan Plitnick, Navjot Singh, John Kelly, Erasmus Schneider, Erica Lasek-Nesselquist |
| EPI_ISL_1398134 | MONTEFIORE MEDICAL CENTER LABORATORIES | Wadsworth Center, New York State Department of Health | Kirsten St. George, Daryl M. Lamson, Alexis Russel, Matthew Shudt, Melissa A Leisner, Jonathan Plitnick, Navjot Singh, John Kelly, Erasmus Schneider, Erica Lasek-Nesselquist |
| EPI_ISL_1398145, EPI_ISL_1398169, EPI_ISL_1398172, EPI_ISL_1398173, EPI_ISL_1398178, EPI_ISL_1398186, EPI_ISL_1398187, EPI_ISL_1398188, EPI_ISL_1398190, EPI_ISL_1398192, EPI_ISL_1398194, EPI_ISL_1398196, EPI_ISL_1398202 | | | |
| see above | Columbia University Irving Medical Center | Wadsworth Center, New York State Department of Health | Kirsten St. George, Daryl M. Lamson, Alexis Russel, Matthew Shudt, Melissa A Leisner, Jonathan Plitnick, Navjot Singh, John Kelly, Erasmus Schneider, Erica Lasek-Nesselquist |
| EPI_ISL_1398268 | WHITE PLAINS HOSPITAL CENTER LABORATORY | Wadsworth Center, New York State Department of Health | Kirsten St. George, Daryl M. Lamson, Alexis Russel, Matthew Shudt, Melissa A Leisner, Jonathan Plitnick, Navjot Singh, John Kelly, Erasmus Schneider, Erica Lasek-Nesselquist |
| EPI_ISL_1398285, EPI_ISL_1398300 | NYC Pandemic Response Lab | Wadsworth Center, New York State Department of Health | Kirsten St. George, Daryl M. Lamson, Alexis Russel, Matthew Shudt, Melissa A Leisner, Jonathan Plitnick, Navjot Singh, John Kelly, Erasmus Schneider, Erica Lasek-Nesselquist |
| EPI_ISL_1398313 | URMC LABS | Wadsworth Center, New York State Department of Health | Kirsten St. George, Daryl M. Lamson, Alexis Russel, Matthew Shudt, Melissa A Leisner, Jonathan Plitnick, Navjot Singh, John Kelly, Erasmus Schneider, Erica Lasek-Nesselquist |
| EPI_ISL_1398326, EPI_ISL_1398327, EPI_ISL_1398330 | NYC Pandemic Response Lab | Wadsworth Center, New York State Department of Health | Kirsten St. George, Daryl M. Lamson, Alexis Russel, Matthew Shudt, Melissa A Leisner, Jonathan Plitnick, Navjot Singh, John Kelly, Erasmus Schneider, Erica Lasek-Nesselquist |
| EPI_ISL_1398337, EPI_ISL_1398339 | MONTEFIORE MEDICAL CENTER LABORATORIES | Wadsworth Center, New York State Department of Health | Kirsten St. George, Daryl M. Lamson, Alexis Russel, Matthew Shudt, Melissa A Leisner, Jonathan Plitnick, Navjot Singh, John Kelly, Erasmus Schneider, Erica Lasek-Nesselquist |
| EPI_ISL_1398344, EPI_ISL_1398348, EPI_ISL_1398349 | NYC Pandemic Response Lab | Wadsworth Center, New York State Department of Health | Kirsten St. George, Daryl M. Lamson, Alexis Russel, Matthew Shudt, Melissa A Leisner, Jonathan Plitnick, Navjot Singh, John Kelly, Erasmus Schneider, Erica Lasek-Nesselquist |
| EPI_ISL_1399589, EPI_ISL_1399590 | Instituto Nacional de Saude (INSA) | Instituto Nacional de Saude (INSA) | Borges et al |
| EPI_ISL_1401473, EPI_ISL_1401477, EPI_ISL_1401478, EPI_ISL_1401479, EPI_ISL_1401481, EPI_ISL_1401483, EPI_ISL_1401484, EPI_ISL_1401485, EPI_ISL_1401486, EPI_ISL_1401489, EPI_ISL_1401490, EPI_ISL_1401492, EPI_ISL_1401504, EPI_ISL_1401507, EPI_ISL_1401508, EPI_ISL_1401510, EPI_ISL_1401511, EPI_ISL_1401515, EPI_ISL_1401517, EPI_ISL_1401521, EPI_ISL_1401523, EPI_ISL_1401525, EPI_ISL_1401526, EPI_ISL_1401527, EPI_ISL_1401530, EPI_ISL_1401534, EPI_ISL_1401540, EPI_ISL_1401542, EPI_ISL_1401550, EPI_ISL_1401553, EPI_ISL_1401558 | | | |
| see above | The Jackson Laboratory | The Jackson Laboratory | Bergeron D, Uvalic J, Renzette N, Adams M, Omerza G, Kelly K, Li L |
| EPI_ISL_1403004 | Hospital of the University of Pennsylvania Molecular Pathology Lab | Bushman Lab - University of Pennsylvania | John Everett, Kyle Rodino, Shantan Reddy, Pascha Hokama, Aoife M. Roche, Young Hwang, Abigail Glascock, Scott Sherrill-Mix, Samantha A. Whiteside, Jevon Graham-Wooten, Layla A. Khatib, Ayannah S. Fitzgerald, Arupa Ganguly, Mike Feldman, Brendan Kelly, Ronald G. Collman and Frederic Bushman |
| EPI_ISL_1405013, EPI_ISL_1405016, EPI_ISL_1405026, EPI_ISL_1405027, EPI_ISL_1405033, EPI_ISL_1405034 | NJDOH, Public Health and Environmental Laboratories | New Jersey Public Health and Environmental Laboratories (NJ PHEL) | Lindsey Bodnar, Shiv K. Verma, Dana Woell, Byeong Jeong |
| EPI_ISL_1406656 | Centre De Prelevement COVID RIOM | CHU Clermont-Ferrand, service de virologie | Bisseux Maxime, Mirand Audrey, Combes Patricia, Henquell Cécile |
| EPI_ISL_1406657 | Centre De Prélévement COVID VICHY | CHU Clermont-Ferrand, service de virologie | Bisseux Maxime, Mirand Audrey, Combes Patricia, Henquell Cécile |
| EPI_ISL_1408479 | Viollier AG | Department of Biosystems Science and Engineering, ETH Zürich | Chaoran Chen, Sarah Nadeau, Catharine Aquino, Ivan Topolsky, Philipp Jablonski, Lara Fuhrmann, David Dreifuss, Katharina Jahn, Andreia Cabral de Gouvea, Maria Domenica Moccia, Simon Grüter, Timothy Sykes, Lennart Opitz, Griffin White, Laura Neff, Doris Popovic, Andrea Patrignani, Jay Tracy, Ralph Schlapbach, Christiane Beckmann, Maurice Redondo, Olivier Kobel, Christoph Noppen, Sophie Seidel, Noemie Santamaria de Souza, Niko Beerenwinkel, Tanja Stadler |
| EPI_ISL_1410112 | Lighthouse Lab in Cambridge | Wellcome Sanger Institute for the COVID-19 Genomics UK (COG-UK) Consortium | Rob Howes, The Lighthouse Lab in Cambridge and Alex Alderton, Roberto Amato, Jeffrey Barrett, Sonia Goncalves, Ewan Harrison, David K. Jackson, Ian Johnston, Dominic Kwiatkowski, Cordelia Langford, John Sillitoe on behalf of the Wellcome Sanger Institute COVID-19 Surveillance Team |
| EPI_ISL_1413154, EPI_ISL_1413193, EPI_ISL_1413221, EPI_ISL_1413316, | Broad Institute Clinical Research Sequencing Platform | Infectious Disease Program, Broad Institute of Harvard and MIT | Siddle,K.J., Adams,G., Pearlman,L., Gladden-Young,A., Vicente,G., Blumenstiel,B., DeFelice,M., Lee,M., McGovern,S., Lagerborg,K., Rudy,M., DeRuff,K., Carter,A., Normandin,E., Bauer,M., Reilly,S., Tomkins-Tinch,C., Loreth,C., Chaluvadi,S., Meldrim,J., Granger,B., Lemieux,J.E., Birren,B.W., Sabeti,P.C., |

| | | | |
|---|---|---|---|
| EPI_ISL_1413498 | | | Larkin,K., Dodge,S., Lennon,N., Madoff,L., Brown,C., Gallagher,G., Smole,S., Park,D.J., Gabriel,S., and MacInnis,B.L. |
| EPI_ISL_1422232 | University of Wisconsin-Madison AIDS Vaccine Research Laboratories | University of Wisconsin-Madison AIDS Vaccine Research Laboratories | Gage Moreno, Katarina Braun, et al. AIDS Vaccine Research Laboratories |
| EPI_ISL_1423379, EPI_ISL_1423400, EPI_ISL_1423411, EPI_ISL_1423419, EPI_ISL_1423421, EPI_ISL_1423428, EPI_ISL_1423430, EPI_ISL_1423438, EPI_ISL_1423441, EPI_ISL_1423452, EPI_ISL_1423459, EPI_ISL_1423476, EPI_ISL_1423481, EPI_ISL_1423485, EPI_ISL_1423494, EPI_ISL_1423495, EPI_ISL_1423500, EPI_ISL_1423503 | | | |
| see above | NYU Langone Health | Departments of Pathology and Medicine, New York University School of Medicine | Adriana Heguy, Dacia Dimartino, Emily Guzman, Christian Marier, Peter Meyn, Sitharam Ramaswami, Gael Westby, Paul Zappile, Yutong Zhang, Paolo Cotzia, Guiqing Wang |
| EPI_ISL_1424140 | Oregon State Public Health Laboratory | Oregon State Public Health Laboratory | LaDonna Grenz, Karim Morey, Joel R. Sevinksy, Kevin, G. Libuit, John Fontana, Shane Sevey |
| EPI_ISL_1443675, EPI_ISL_1443676, EPI_ISL_1443680 | Omics Sciences Laboratory | Omics Sciences Laboratory | Derly Andrade Molina, Rubén Armas González, Gabriel Morey León, Darlyn Amaya, Katheryn Sacheri Viteri, Emily Sulay Saltos Montalvo, Paula Juliana Gavilanes Jarrín, Juan Carlos Fernández Cadena |
| EPI_ISL_1443892 | Outre mer | National Reference Center for Viruses of Respiratory Infections, Institut Pasteur, Paris | Marion Barbet, Sylvie Behillil, Méline Bizard,Frédéric Lemoine,Corinne Maufrais,Christophe Malabat, Angela Brisebarre, Camille Capel, Louise Lefrançois, Etienne Simon-Lorière, Vincent Enouf, Maud Vanpeene, Sylvie van der Werf,StéPhanie Guyomard-Rabenirina |
| EPI_ISL_1446204 | Texas Children's Hospital | Texas Children's Microbiome Center | Ruth Ann Luna, Jennifer K. Spinler, James Dunn, James Versalovic, Ila Singh |
| EPI_ISL_1447088 | DOHMH Corona | New York City Public Health Laboratory | Jade Wang, et al. |
| EPI_ISL_1447090 | DOHMH Morrisania | New York City Public Health Laboratory | Jade Wang, et al. |
| EPI_ISL_1447091 | OCME Office Of Chief Medical Examiner | New York City Public Health Laboratory | Jade Wang, et al. |
| EPI_ISL_1447092 | DOHMH Jamaica | New York City Public Health Laboratory | Jade Wang, et al. |
| EPI_ISL_1447100 | OCME Office Of Chief Medical Examiner | New York City Public Health Laboratory | Jade Wang, et al. |
| EPI_ISL_1447102 | Department of Homeless Services | New York City Public Health Laboratory | Jade Wang, et al. |
| EPI_ISL_1447103, EPI_ISL_1447106 | DOHMH Jamaica | New York City Public Health Laboratory | Jade Wang, et al. |
| EPI_ISL_1447108, EPI_ISL_1447110 | DOHMH Morrisania | New York City Public Health Laboratory | Jade Wang, et al. |
| EPI_ISL_1447145 | DOHMH PHL | New York City Public Health Laboratory | Jade Wang, et al. |
| EPI_ISL_1447146 | DOHMH Jamaica | New York City Public Health Laboratory | Jade Wang, et al. |
| EPI_ISL_1447148 | DOHMH Crown Heights | New York City Public Health Laboratory | Jade Wang, et al. |
| EPI_ISL_1447149, EPI_ISL_1447150 | DOHMH PHL | New York City Public Health Laboratory | Jade Wang, et al. |
| EPI_ISL_1447152, EPI_ISL_1447159 | DOHMH Jamaica | New York City Public Health Laboratory | Jade Wang, et al. |
| EPI_ISL_1447160 | DOHMH Corona | New York City Public Health Laboratory | Jade Wang, et al. |
| EPI_ISL_1447163, EPI_ISL_1447164 | DOHMH Jamaica | New York City Public Health Laboratory | Jade Wang, et al. |
| EPI_ISL_1447165, EPI_ISL_1447171 | DOHMH Corona | New York City Public Health Laboratory | Jade Wang, et al. |
| EPI_ISL_1447173 | DOHMH Riverside | New York City Public Health Laboratory | Jade Wang, et al. |
| EPI_ISL_1447176 | DOHMH Fort Greene | New York City Public Health Laboratory | Jade Wang, et al. |
| EPI_ISL_1447177 | OCME Office Of Chief Medical Examiner | New York City Public Health Laboratory | Jade Wang, et al. |
| EPI_ISL_1447179 | DOHMH Jamaica | New York City Public Health Laboratory | Jade Wang, et al. |
| EPI_ISL_1447181 | DOHMH Morrisania | New York City Public Health Laboratory | Jade Wang, et al. |
| EPI_ISL_1447291 | South Eastern Area Laboratory Services (SEALS) | NSW Health Pathology - Institute of Clinical Pathology and Medical Research; Westmead Hospital; University of Sydney | CIDM-PH et al. |
| EPI_ISL_1447510, EPI_ISL_1447512, EPI_ISL_1447514, EPI_ISL_1447520, EPI_ISL_1447531, EPI_ISL_1447532, EPI_ISL_1447534, EPI_ISL_1447536, EPI_ISL_1447542, EPI_ISL_1447546, EPI_ISL_1447549, EPI_ISL_1447551, EPI_ISL_1447552, EPI_ISL_1447554, EPI_ISL_1447556, EPI_ISL_1447558, EPI_ISL_1447562, EPI_ISL_1447563, EPI_ISL_1447566, EPI_ISL_1447571, EPI_ISL_1447576, EPI_ISL_1447578, EPI_ISL_1447582, EPI_ISL_1447592, EPI_ISL_1447597, EPI_ISL_1447598, EPI_ISL_1447599, EPI_ISL_1447600, EPI_ISL_1447601, EPI_ISL_1447602, EPI_ISL_1447605, EPI_ISL_1447607, EPI_ISL_1447608, EPI_ISL_1447609, EPI_ISL_1447610, EPI_ISL_1447615, EPI_ISL_1447621, EPI_ISL_1447622, EPI_ISL_1447625, EPI_ISL_1447645, EPI_ISL_1447650, EPI_ISL_1447653, EPI_ISL_1447654, EPI_ISL_1447655, EPI_ISL_1447658, EPI_ISL_1447665, EPI_ISL_1447667 | | | |
| see above | Yale Clinical Virology Lab | Grubaugh Lab - Yale School of Public Health | Joseph Fauver, Mallery Breban, Isabel Ott, Tara Alpert, Mary Petrone, Anderson Brito, Chantal Vogels, Annie Watkins, Chaney Kalinich, Jessica Rothman, Marie L. Landry, Nathan Grubaugh |
| EPI_ISL_1448609 | University Hospitals of Geneva, Laboratory of Virology | HUG, Laboratory of Virology and the Health2030 Genome Center | Samuel Cordey, Ana Rita Goncalves, Laurent Kaiser, Lorenzo Cerutti, Henri Pegeot, Melyssa Elies, Deborah Penet, Keith Harshman, Ioannis Xenarios, Emmanouil Dermitzakis |
| EPI_ISL_1461303 | URMC LABS | Wadsworth Center, New York State Department of Health | Kirsten St. George, Daryl M. Lamson, Alexis Russell, Matthew Shudt, Melissa A Leisner, Jonathan Plitnick, Catharine Prussing, Navjot Singh, John Kelly, Erasmus Schneider, Erica Lasek-Nesselquist |
| EPI_ISL_1461316 | SUNY UPSTATE MEDICAL UNIVERSITY | Wadsworth Center, New York State Department of Health | Kirsten St. George, Daryl M. Lamson, Alexis Russell, Matthew Shudt, Melissa A Leisner, Jonathan Plitnick, Catharine Prussing, Navjot Singh, John Kelly, Erasmus Schneider, Erica Lasek-Nesselquist |
| EPI_ISL_1461323, EPI_ISL_1461327, EPI_ISL_1461342, EPI_ISL_1461344, EPI_ISL_1461347, EPI_ISL_1461352 | NYC Pandemic Response Lab | Wadsworth Center, New York State Department of Health | Kirsten St. George, Daryl M. Lamson, Alexis Russell, Matthew Shudt, Melissa A Leisner, Jonathan Plitnick, Catharine Prussing, Navjot Singh, John Kelly, Erasmus Schneider, Erica Lasek-Nesselquist |
| EPI_ISL_1461359, EPI_ISL_1461360, EPI_ISL_1461374 | TEMPUS LABS INC | Wadsworth Center, New York State Department of Health | Kirsten St. George, Daryl M. Lamson, Alexis Russell, Matthew Shudt, Melissa A Leisner, Jonathan Plitnick, Catharine Prussing, Navjot Singh, John Kelly, Erasmus Schneider, Erica Lasek-Nesselquist |
| EPI_ISL_1461505 | Michigan Department of Health and Human Services, Bureau of Laboratories | Michigan Department of Health and Human Services, Bureau of Laboratories | Blankenship HM, Riner D, Soehnlen MK |
| EPI_ISL_1462512 | URMC LABS | Wadsworth Center, New York State Department of Health | Kirsten St. George, Daryl M. Lamson, Alexis Russell, Matthew Shudt, Melissa A Leisner, Jonathan Plitnick, Catharine Prussing, Navjot Singh, John Kelly, Erasmus Schneider, Erica Lasek-Nesselquist |
| EPI_ISL_1462542 | SUNY UPSTATE MEDICAL UNIVERSITY | Wadsworth Center, New York State Department of Health | Kirsten St. George, Daryl M. Lamson, Alexis Russell, Matthew Shudt, Melissa A Leisner, Jonathan Plitnick, Catharine Prussing, Navjot Singh, John Kelly, Erasmus Schneider, Erica Lasek-Nesselquist |
| EPI_ISL_1462551 | MONTEFIORE MEDICAL CENTER LABORATORIES | Wadsworth Center, New York State Department of Health | Kirsten St. George, Daryl M. Lamson, Alexis Russell, Matthew Shudt, Melissa A Leisner, Jonathan Plitnick, Catharine Prussing, Navjot Singh, John Kelly, Erasmus Schneider, Erica Lasek-Nesselquist |
| EPI_ISL_1462576, EPI_ISL_1462579 | NORTHWELL HEALTH LABORATORIES | Wadsworth Center, New York State Department of Health | Kirsten St. George, Daryl M. Lamson, Alexis Russell, Matthew Shudt, Melissa A Leisner, Jonathan Plitnick, Catharine Prussing, Navjot Singh, John Kelly, Erasmus Schneider, Erica Lasek-Nesselquist |
| EPI_ISL_1462583 | Laboratory Corporation of America | Centers for Disease Control and Prevention Division of Viral Diseases, Pathogen Discovery | Dakota Howard, Dhwani Batra, Peter W. Cook, Kara Moser, Adrian Paskey, Jason Caravas, Benjamin Rambo-Martin, Shatavia Morrison, Christopher Gulvick, Scott Sammons, Yvette Unoarumhi, Darlene Wagner, Matthew Schmerer, Minoo Agarwal, Eyad Almasri, Debbie Boles, Ayla Burns, Nuthawin Charoensri, Oren Cohen, Susan Countryman, Mary Ann Cristobal, Bobbi Croy, Suzanne Dale, Hrushikesh Deshmukh, Amanda Douglas, Vincent Drouillon, Marcia Eisenberg, Howard Engler, Rama Ghatti, Prashant Gupta, Susan Hicks, Jake Humphrey, Lax Iyer, Manoj Jain, Mohan Kolli, Brian Krueger, Tim Kuphal, Stanley Letovsky, Michael Levandoski, Craig Lukasik, Jonathan Meltzer, Brian Norvell, Mindy Nye, Scott Parker, Christos Petropoulos, John Pruitt, |

| | | | |
|---|---|---|---|
| | | | Steven Ragan, Scott Ryan, Mike Sapeta, Jana Schroth, Suresh Babu Selvaraju, Goran Stevovic, Amanda Suchanek, Andrea Throop, Lyndon Tilson, Thomas Urban, Joe Voshell, Kimberly Wagner, Jonathan Williams, Mary Williamson, Qian Zeng, Tricia Zwiefelhofer, Clinton R. Paden, Duncan MacCannell |
| EPI_ISL_1462589, EPI_ISL_1462594, EPI_ISL_1462599, EPI_ISL_1462604, EPI_ISL_1462609, EPI_ISL_1462618, EPI_ISL_1462620 | NORTHWELL HEALTH LABORATORIES | Wadsworth Center, New York State Department of Health | Kirsten St. George, Daryl M. Lamson, Alexis Russell, Matthew Shudt, Melissa A Leisner, Jonathan Plitnick, Catharine Prussing, Navjot Singh, John Kelly, Erasmus Schneider, Erica Lasek-Nesselquist |
| EPI_ISL_1462838, EPI_ISL_1463087, EPI_ISL_1463100, EPI_ISL_1463112, EPI_ISL_1463118, EPI_ISL_1463119, EPI_ISL_1463187, EPI_ISL_1463208, EPI_ISL_1463318, EPI_ISL_1463321, EPI_ISL_1463400 | | | |
| see above | Laboratory Corporation of America | Centers for Disease Control and Prevention Division of Viral Diseases, Pathogen Discovery | Dakota Howard, Dhwani Batra, Peter W. Cook, Kara Moser, Adrian Paskey, Jason Caravas, Benjamin Rambo-Martin, Shatavia Morrison, Christopher Gulvick, Scott Sammons, Yvette Unoarumhi, Darlene Wagner, Matthew Schmerer, Minoo Agarwal, Eyad Almasri, Debbie Boles, Ayla Burns, Nuthawin Charoensri, Oren Cohen, Susan Countryman, Mary Ann Cristobal, Bobbi Croy, Suzanne Dale, Hrushikesh Deshmukh, Amanda Douglas, Vincent Drouillon, Marcia Eisenberg, Howard Engler, Rama Ghatti, Prashant Gupta, Susan Hicks, Jake Humphrey, Lax Iyer, Manoj Jain, Mohan Kolli, Brian Krueger, Tim Kuphal, Stanley Letovsky, Michael Levandoski, Craig Lukasik, Jonathan Meltzer, Brian Norvell, Mindy Nye, Scott Parker, Christos Petropoulos, John Pruitt, Steven Ragan, Scott Ryan, Mike Sapeta, Jana Schroth, Suresh Babu Selvaraju, Goran Stevovic, Amanda Suchanek, Andrea Throop, Lyndon Tilson, Thomas Urban, Joe Voshell, Kimberly Wagner, Jonathan Williams, Mary Williamson, Qian Zeng, Tricia Zwiefelhofer, Clinton R. Paden, Duncan MacCannell |
| EPI_ISL_1464568 | Labor Dr. Fenner und Kollegen | Heinrich Pette Institute, Leibniz Institute for Experimental Virology | Alexis Robitaille, Thomas Günther, Johannes Knobloch, Martin Aepfelbacher, Nicole Fischer, Adam Grundhoff |
| EPI_ISL_1465000 | Sonora Quest Laboratories | TGen North | "Jolene Bowers, Heather Centner, Chris French, Hayley Yaglom, Ashlyn Pfeiffer, Darrin Lemmer, Dave Engelthaler, The Arizona COVID Genomics Union (ACGU)" |
| EPI_ISL_1465635 | ALBANY MEDICAL CENTER | Wadsworth Center, New York State Department of Health | Kirsten St. George, Daryl M. Lamson, Alexis Russell, Matthew Shudt, Melissa A Leisner, Jonathan Plitnick, Catharine Prussing, Navjot Singh, John Kelly, Erasmus Schneider, Erica Lasek-Nesselquist |
| EPI_ISL_1465668, EPI_ISL_1465671 | MONTEFIORE MEDICAL CENTER LABORATORIES | Wadsworth Center, New York State Department of Health | Kirsten St. George, Daryl M. Lamson, Alexis Russell, Matthew Shudt, Melissa A Leisner, Jonathan Plitnick, Catharine Prussing, Navjot Singh, John Kelly, Erasmus Schneider, Erica Lasek-Nesselquist |
| EPI_ISL_1465692 | SUNY UPSTATE MEDICAL UNIVERSITY | Wadsworth Center, New York State Department of Health | Kirsten St. George, Daryl M. Lamson, Alexis Russell, Matthew Shudt, Melissa A Leisner, Jonathan Plitnick, Catharine Prussing, Navjot Singh, John Kelly, Erasmus Schneider, Erica Lasek-Nesselquist |
| EPI_ISL_1465701, EPI_ISL_1465706, EPI_ISL_1465709, EPI_ISL_1465710 | MONTEFIORE MEDICAL CENTER LABORATORIES | Wadsworth Center, New York State Department of Health | Kirsten St. George, Daryl M. Lamson, Alexis Russell, Matthew Shudt, Melissa A Leisner, Jonathan Plitnick, Catharine Prussing, Navjot Singh, John Kelly, Erasmus Schneider, Erica Lasek-Nesselquist |
| EPI_ISL_1465722 | NYC Pandemic Response Lab | Wadsworth Center, New York State Department of Health | Kirsten St. George, Daryl M. Lamson, Alexis Russell, Matthew Shudt, Melissa A Leisner, Jonathan Plitnick, Catharine Prussing, Navjot Singh, John Kelly, Erasmus Schneider, Erica Lasek-Nesselquist |
| EPI_ISL_1465737 | URMC LABS | Wadsworth Center, New York State Department of Health | Kirsten St. George, Daryl M. Lamson, Alexis Russell, Matthew Shudt, Melissa A Leisner, Jonathan Plitnick, Catharine Prussing, Navjot Singh, John Kelly, Erasmus Schneider, Erica Lasek-Nesselquist |
| EPI_ISL_1465761, EPI_ISL_1465762 | NORTHWELL HEALTH LABORATORIES | Wadsworth Center, New York State Department of Health | Kirsten St. George, Daryl M. Lamson, Alexis Russell, Matthew Shudt, Melissa A Leisner, Jonathan Plitnick, Catharine Prussing, Navjot Singh, John Kelly, Erasmus Schneider, Erica Lasek-Nesselquist |
| EPI_ISL_1465797, EPI_ISL_1465798 | MONTEFIORE MEDICAL CENTER LABORATORIES | Wadsworth Center, New York State Department of Health | Kirsten St. George, Daryl M. Lamson, Alexis Russell, Matthew Shudt, Melissa A Leisner, Jonathan Plitnick, Catharine Prussing, Navjot Singh, John Kelly, Erasmus Schneider, Erica Lasek-Nesselquist |
| EPI_ISL_1465807, EPI_ISL_1465809, EPI_ISL_1465810, EPI_ISL_1465811, EPI_ISL_1465819, EPI_ISL_1465827, EPI_ISL_1465828, EPI_ISL_1465831 | NYC Pandemic Response Lab | Wadsworth Center, New York State Department of Health | Kirsten St. George, Daryl M. Lamson, Alexis Russell, Matthew Shudt, Melissa A Leisner, Jonathan Plitnick, Catharine Prussing, Navjot Singh, John Kelly, Erasmus Schneider, Erica Lasek-Nesselquist |
| EPI_ISL_1465838, EPI_ISL_1465840, EPI_ISL_1465842 | STONY BROOK UNIVERSITY HOSPITAL | Wadsworth Center, New York State Department of Health | Kirsten St. George, Daryl M. Lamson, Alexis Russell, Matthew Shudt, Melissa A Leisner, Jonathan Plitnick, Catharine Prussing, Navjot Singh, John Kelly, Erasmus Schneider, Erica Lasek-Nesselquist |
| EPI_ISL_1465853 | GLENS FALLS HOSPITAL LABORATORY | Wadsworth Center, New York State Department of Health | Kirsten St. George, Daryl M. Lamson, Alexis Russell, Matthew Shudt, Melissa A Leisner, Jonathan Plitnick, Catharine Prussing, Navjot Singh, John Kelly, Erasmus Schneider, Erica Lasek-Nesselquist |
| EPI_ISL_1465860, EPI_ISL_1465862, EPI_ISL_1465871, EPI_ISL_1465873, EPI_ISL_1465878 | TEMPUS LABS INC | Wadsworth Center, New York State Department of Health | Kirsten St. George, Daryl M. Lamson, Alexis Russell, Matthew Shudt, Melissa A Leisner, Jonathan Plitnick, Catharine Prussing, Navjot Singh, John Kelly, Erasmus Schneider, Erica Lasek-Nesselquist |
| EPI_ISL_1470560, EPI_ISL_1470561, EPI_ISL_1470563, EPI_ISL_1470564, EPI_ISL_1470567, EPI_ISL_1470569, EPI_ISL_1470570, EPI_ISL_1470575, EPI_ISL_1470583, EPI_ISL_1470585, EPI_ISL_1470587, EPI_ISL_1470588, EPI_ISL_1470591, EPI_ISL_1470597, EPI_ISL_1470599, EPI_ISL_1470603, EPI_ISL_1470605, EPI_ISL_1470607, EPI_ISL_1470612, EPI_ISL_1470614, EPI_ISL_1470618, EPI_ISL_1470619, EPI_ISL_1470628, EPI_ISL_1470634, EPI_ISL_1470635, EPI_ISL_1470637, EPI_ISL_1470640, EPI_ISL_1470646, EPI_ISL_1470648, EPI_ISL_1470649, EPI_ISL_1470650, EPI_ISL_1470653, EPI_ISL_1470654, EPI_ISL_1470662, EPI_ISL_1470672, EPI_ISL_1470674, EPI_ISL_1470676, EPI_ISL_1470677, EPI_ISL_1470678, EPI_ISL_1470681, EPI_ISL_1470683, EPI_ISL_1470686, EPI_ISL_1470688, EPI_ISL_1470689, EPI_ISL_1470693, EPI_ISL_1470694, EPI_ISL_1470695, EPI_ISL_1470696, EPI_ISL_1470699, EPI_ISL_1470700, EPI_ISL_1470705, EPI_ISL_1470706, EPI_ISL_1470707, EPI_ISL_1470712, EPI_ISL_1470714, EPI_ISL_1470715, EPI_ISL_1470718, EPI_ISL_1470720, EPI_ISL_1470721, EPI_ISL_1470725, EPI_ISL_1470726, EPI_ISL_1470728, EPI_ISL_1470730, EPI_ISL_1470731, EPI_ISL_1470738, EPI_ISL_1470739 | | | |
| see above | The Jackson Laboratory | The Jackson Laboratory | Bergeron D, Renzette N, Adams M, Omerza G, Kelly K, Li L |
| EPI_ISL_1470744, EPI_ISL_1470754, EPI_ISL_1470758, EPI_ISL_1470765, EPI_ISL_1470770, EPI_ISL_1470780, EPI_ISL_1470781, EPI_ISL_1470782, EPI_ISL_1470787, EPI_ISL_1470790, EPI_ISL_1470795, EPI_ISL_1470802, EPI_ISL_1470803, EPI_ISL_1470813, EPI_ISL_1470815, EPI_ISL_1470821, EPI_ISL_1470823, EPI_ISL_1470830, EPI_ISL_1470833, EPI_ISL_1470843, EPI_ISL_1470847, EPI_ISL_1470851, EPI_ISL_1470853, EPI_ISL_1470860, EPI_ISL_1470861, EPI_ISL_1470871, EPI_ISL_1470873, EPI_ISL_1470874, EPI_ISL_1470876, EPI_ISL_1470880, EPI_ISL_1470882, EPI_ISL_1470886, EPI_ISL_1470899, EPI_ISL_1470901, EPI_ISL_1470902, EPI_ISL_1470903, EPI_ISL_1470905, EPI_ISL_1470909, EPI_ISL_1470916, EPI_ISL_1470927, EPI_ISL_1470931, EPI_ISL_1470932, EPI_ISL_1470933, EPI_ISL_1470936, EPI_ISL_1470947, EPI_ISL_1470953, EPI_ISL_1470954, EPI_ISL_1470963, EPI_ISL_1470975, EPI_ISL_1470978, EPI_ISL_1470984, EPI_ISL_1470986, EPI_ISL_1470989, EPI_ISL_1471000, EPI_ISL_1471009, EPI_ISL_1471021, EPI_ISL_1471026, EPI_ISL_1471028, EPI_ISL_1471030, EPI_ISL_1471036, EPI_ISL_1471041, EPI_ISL_1471048, EPI_ISL_1471050, EPI_ISL_1471051, EPI_ISL_1471053, EPI_ISL_1471054, EPI_ISL_1471067, EPI_ISL_1471068, EPI_ISL_1471072, EPI_ISL_1471076, EPI_ISL_1471080, EPI_ISL_1471088, EPI_ISL_1471092, EPI_ISL_1471099, EPI_ISL_1471105, EPI_ISL_1471109, EPI_ISL_1471111, EPI_ISL_1471123, EPI_ISL_1471130, EPI_ISL_1471135, EPI_ISL_1471139, EPI_ISL_1471143, EPI_ISL_1471163, EPI_ISL_1471166, EPI_ISL_1471169, EPI_ISL_1471173, EPI_ISL_1471176, EPI_ISL_1471178, EPI_ISL_1471179, EPI_ISL_1471190, EPI_ISL_1471191, EPI_ISL_1471197, EPI_ISL_1471208, EPI_ISL_1471209, EPI_ISL_1471211, EPI_ISL_1471213, EPI_ISL_1471216, EPI_ISL_1471220, EPI_ISL_1471222, EPI_ISL_1471224, EPI_ISL_1471238, EPI_ISL_1471240, EPI_ISL_1471242, EPI_ISL_1471243, EPI_ISL_1471249, EPI_ISL_1471250, EPI_ISL_1471253, EPI_ISL_1471256, EPI_ISL_1471264, EPI_ISL_1471267, EPI_ISL_1471273, EPI_ISL_1471280, EPI_ISL_1471284, EPI_ISL_1471293, EPI_ISL_1471295, EPI_ISL_1471298, EPI_ISL_1471301, EPI_ISL_1471318, EPI_ISL_1471323, EPI_ISL_1471324, EPI_ISL_1471335, EPI_ISL_1471340, EPI_ISL_1471342, EPI_ISL_1471358, EPI_ISL_1471365, EPI_ISL_1471366, EPI_ISL_1471389, EPI_ISL_1471393, EPI_ISL_1471394, EPI_ISL_1471395, EPI_ISL_1471400, EPI_ISL_1471402, EPI_ISL_1471408, EPI_ISL_1471409, EPI_ISL_1471421, EPI_ISL_1471424, EPI_ISL_1471425, EPI_ISL_1471429, EPI_ISL_1471436, EPI_ISL_1471443, EPI_ISL_1471446, EPI_ISL_1471452, EPI_ISL_1471461, EPI_ISL_1471462, EPI_ISL_1471464, EPI_ISL_1471465, EPI_ISL_1471496, EPI_ISL_1471503, EPI_ISL_1471507, EPI_ISL_1471509, EPI_ISL_1471510, EPI_ISL_1471520, EPI_ISL_1471525, EPI_ISL_1471527, EPI_ISL_1471532, EPI_ISL_1471537, EPI_ISL_1471540, EPI_ISL_1471552, EPI_ISL_1471560, EPI_ISL_1471568, EPI_ISL_1471569, EPI_ISL_1471571, EPI_ISL_1471586, EPI_ISL_1471589, EPI_ISL_1471590, EPI_ISL_1471593, EPI_ISL_1471625, EPI_ISL_1471633, EPI_ISL_1471634, EPI_ISL_1471635, EPI_ISL_1471644, EPI_ISL_1471646, EPI_ISL_1471656, EPI_ISL_1471658, EPI_ISL_1471669, EPI_ISL_1471677, EPI_ISL_1471679, EPI_ISL_1471680, EPI_ISL_1471683, EPI_ISL_1471686, EPI_ISL_1471688, EPI_ISL_1471690, EPI_ISL_1471692, EPI_ISL_1471698, EPI_ISL_1471701, EPI_ISL_1471704, EPI_ISL_1471707, EPI_ISL_1471710, EPI_ISL_1471712, EPI_ISL_1471716, EPI_ISL_1471718, EPI_ISL_1471720, EPI_ISL_1471721, EPI_ISL_1471723, EPI_ISL_1471730, EPI_ISL_1471736, EPI_ISL_1471741, EPI_ISL_1471755, EPI_ISL_1471762, EPI_ISL_1471772, EPI_ISL_1471783, EPI_ISL_1471789, EPI_ISL_1471791, EPI_ISL_1471796, EPI_ISL_1471800, EPI_ISL_1471804, EPI_ISL_1471808, EPI_ISL_1471816, EPI_ISL_1471820, EPI_ISL_1471824, EPI_ISL_1471827, EPI_ISL_1471843, EPI_ISL_1471847, EPI_ISL_1471852, EPI_ISL_1471855, EPI_ISL_1471859, EPI_ISL_1471860, EPI_ISL_1471861, EPI_ISL_1471865, EPI_ISL_1471868, EPI_ISL_1471876, EPI_ISL_1471879, EPI_ISL_1471896, EPI_ISL_1471898, EPI_ISL_1471900, EPI_ISL_1471908, EPI_ISL_1471911, EPI_ISL_1471913, EPI_ISL_1471916, EPI_ISL_1471920, EPI_ISL_1471921, EPI_ISL_1471925, EPI_ISL_1471927, EPI_ISL_1471930, EPI_ISL_1471931, EPI_ISL_1471933, EPI_ISL_1471939, EPI_ISL_1471943, EPI_ISL_1471957, EPI_ISL_1471980, EPI_ISL_1471984, EPI_ISL_1471988, EPI_ISL_1471989, EPI_ISL_1472006, EPI_ISL_1472013, EPI_ISL_1472014, EPI_ISL_1472015, EPI_ISL_1472016, EPI_ISL_1472018, EPI_ISL_1472021, EPI_ISL_1472024, EPI_ISL_1472026, EPI_ISL_1472032, EPI_ISL_1472053, EPI_ISL_1472055, EPI_ISL_1472057, EPI_ISL_1472064, EPI_ISL_1472066, EPI_ISL_1472068, EPI_ISL_1472074 | | | |
| see above | Pandemic Response Lab - NYC | Pandemic Response Lab, R&D | Henry Lee, Michael Hammerling, Melissa Hopkins, Cybill del Castillo, Shinyoung Clair Kang, William Ward, Pradeep Bugga, Sol Rey, Dylan Law, Haiping Hao, Jon Laurent |
| EPI_ISL_1477057 | Hospital General Universitario Gregorio Marañón | Hospital General Universitario Gregorio Marañón | Sergio Buenestado Serrano, Pedro Sola Campoy, Laura Pérez-Lago, Cristina Rodriguez-Grande, Pilar Catalán, Patricia Muñoz, Darío García de Viedma |
| EPI_ISL_1478113 | CA DPH Viral and Rickettsial Disease Laboratory | Chan-Zuckerberg Biohub | CZB Cliahub Consortium |
| EPI_ISL_1481723, EPI_ISL_1481724, EPI_ISL_1481755, EPI_ISL_1481927, EPI_ISL_1481967, EPI_ISL_1482119, EPI_ISL_1482125 | Fulgent Genetics | Centers for Disease Control and Prevention Division of Viral Diseases, Pathogen Discovery | Dakota Howard, Dhwani Batra, Peter W. Cook, Kara Moser, Adrian Paskey, Jason Caravas, Benjamin Rambo-Martin, Shatavia Morrison, Christopher Gulvick, Scott Sammons, Yvette Unoarumhi, Darlene Wagner, Matthew Schmerer, Harry Gao, Mickey Li, John Gao, Joseph Fierro, Benafsh Sapra, Becky Tsai, Yan Meng, Doreen Ng, James Xie, Clinton R. Paden, Duncan MacCannell |
| EPI_ISL_1482288 | NYC Pandemic Response Lab | Wadsworth Center, New York State Department of Health | Kirsten St. George, Daryl M. Lamson, Alexis Russell, Matthew Shudt, Melissa A Leisner, Jonathan Plitnick, Catharine Prussing, Navjot Singh, John Kelly, |

| | | | |
|---|---|---|---|
| | | | Erasmus Schneider, Erica Lasek-Nesselquist |
| EPI_ISL_1482303, EPI_ISL_1482307, EPI_ISL_1482309 | URMC LABS | Wadsworth Center, New York State Department of Health | Kirsten St. George, Daryl M. Lamson, Alexis Russell, Matthew Shudt, Melissa A Leisner, Jonathan Plitnick, Catharine Prussing, Navjot Singh, John Kelly, Erasmus Schneider, Erica Lasek-Nesselquist |
| EPI_ISL_1482343 | MONTEFIORE NEW ROCHELLE HOSPITAL LABORATORY | Wadsworth Center, New York State Department of Health | Kirsten St. George, Daryl M. Lamson, Alexis Russell, Matthew Shudt, Melissa A Leisner, Jonathan Plitnick, Catharine Prussing, Navjot Singh, John Kelly, Erasmus Schneider, Erica Lasek-Nesselquist |
| EPI_ISL_1482394, EPI_ISL_1482396, EPI_ISL_1482398, EPI_ISL_1482400, EPI_ISL_1482401 | WESTCHESTER MEDICAL CENTER | Wadsworth Center, New York State Department of Health | Kirsten St. George, Daryl M. Lamson, Alexis Russell, Matthew Shudt, Melissa A Leisner, Jonathan Plitnick, Catharine Prussing, Navjot Singh, John Kelly, Erasmus Schneider, Erica Lasek-Nesselquist |
| EPI_ISL_1482408 | BOSTON HEART DIAGNOSTICS CORP | Wadsworth Center, New York State Department of Health | Kirsten St. George, Daryl M. Lamson, Alexis Russell, Matthew Shudt, Melissa A Leisner, Jonathan Plitnick, Catharine Prussing, Navjot Singh, John Kelly, Erasmus Schneider, Erica Lasek-Nesselquist |
| EPI_ISL_1482414, EPI_ISL_1482418, EPI_ISL_1482427, EPI_ISL_1482428 | WESTCHESTER MEDICAL CENTER | Wadsworth Center, New York State Department of Health | Kirsten St. George, Daryl M. Lamson, Alexis Russell, Matthew Shudt, Melissa A Leisner, Jonathan Plitnick, Catharine Prussing, Navjot Singh, John Kelly, Erasmus Schneider, Erica Lasek-Nesselquist |
| EPI_ISL_1482456, EPI_ISL_1482462, EPI_ISL_1482463 | ALBANY MEDICAL CENTER | Wadsworth Center, New York State Department of Health | Kirsten St. George, Daryl M. Lamson, Alexis Russell, Matthew Shudt, Melissa A Leisner, Jonathan Plitnick, Catharine Prussing, Navjot Singh, John Kelly, Erasmus Schneider, Erica Lasek-Nesselquist |
| EPI_ISL_1482545, EPI_ISL_1482546, EPI_ISL_1482547, EPI_ISL_1482550, EPI_ISL_1482553 | WESTCHESTER MEDICAL CENTER | Wadsworth Center, New York State Department of Health | Kirsten St. George, Daryl M. Lamson, Alexis Russell, Matthew Shudt, Melissa A Leisner, Jonathan Plitnick, Catharine Prussing, Navjot Singh, John Kelly, Erasmus Schneider, Erica Lasek-Nesselquist |
| EPI_ISL_1482557, EPI_ISL_1482565, EPI_ISL_1482566, EPI_ISL_1482567 | NORTHWELL HEALTH LABORATORIES | Wadsworth Center, New York State Department of Health | Kirsten St. George, Daryl M. Lamson, Alexis Russell, Matthew Shudt, Melissa A Leisner, Jonathan Plitnick, Catharine Prussing, Navjot Singh, John Kelly, Erasmus Schneider, Erica Lasek-Nesselquist |
| EPI_ISL_1482576 | WHITE PLAINS HOSPITAL CENTER LABORATORY | Wadsworth Center, New York State Department of Health | Kirsten St. George, Daryl M. Lamson, Alexis Russell, Matthew Shudt, Melissa A Leisner, Jonathan Plitnick, Catharine Prussing, Navjot Singh, John Kelly, Erasmus Schneider, Erica Lasek-Nesselquist |
| EPI_ISL_1482583, EPI_ISL_1482603, EPI_ISL_1482608, EPI_ISL_1482609 | NORTHWELL HEALTH LABORATORIES | Wadsworth Center, New York State Department of Health | Kirsten St. George, Daryl M. Lamson, Alexis Russell, Matthew Shudt, Melissa A Leisner, Jonathan Plitnick, Catharine Prussing, Navjot Singh, John Kelly, Erasmus Schneider, Erica Lasek-Nesselquist |
| EPI_ISL_1482617, EPI_ISL_1482619, EPI_ISL_1482620, EPI_ISL_1482621, EPI_ISL_1482622, EPI_ISL_1482623 | WESTCHESTER MEDICAL CENTER | Wadsworth Center, New York State Department of Health | Kirsten St. George, Daryl M. Lamson, Alexis Russell, Matthew Shudt, Melissa A Leisner, Jonathan Plitnick, Catharine Prussing, Navjot Singh, John Kelly, Erasmus Schneider, Erica Lasek-Nesselquist |
| EPI_ISL_1483256 | Nevada State Public Health Laboratory | Nevada State Public Health Laboratory | Andrew Gorzalski, Mark Pandori |
| EPI_ISL_1483590 | Alaska State Virology Laboratory | Alaska State Virology Laboratory | Stephanie DeRonde, Elva House, Lisa Smith, Ph.D., Jack Chen, Ph.D. |
| EPI_ISL_1489661, EPI_ISL_1489662, EPI_ISL_1489667, EPI_ISL_1489675, EPI_ISL_1489680, EPI_ISL_1489688, EPI_ISL_1489691, EPI_ISL_1489700, EPI_ISL_1489707, EPI_ISL_1489709, EPI_ISL_1489712, EPI_ISL_1489714 | | | |
| see above | NJDOH, Public Health and Environmental Laboratories | New Jersey Public Health and Environmental Laboratories (NJ PHEL) | Lindsey Bodnar, Shiv K. Verma, Dana Woell, Byeong Jeong |
| EPI_ISL_1489728 | National Public Health Laboratory, National Centre for Infectious Diseases | National Public Health Laboratory, National Centre for Infectious Diseases | Tze Minn Mak, Zhenyang Zhou, Grace Jie Yin Ngan, Royce Ang, Lin Cui, Raymond Tzer Pin Lin |
| EPI_ISL_1489870 | Missouri State Public Health Laboratory | Missouri State Public Health Laboratory | Matthew Sinn, Joshua Barry, Ashley New |
| EPI_ISL_1491424 | University of Michigan Clinical Microbiology Laboratory | Lauring Lab, University of Michigan, Department of Microbiology and Immunology | Valesano |
| EPI_ISL_1491770, EPI_ISL_1491776, EPI_ISL_1491780, EPI_ISL_1491795, EPI_ISL_1491833, EPI_ISL_1491846, EPI_ISL_1491849, EPI_ISL_1491853, EPI_ISL_1491862, EPI_ISL_1491865, EPI_ISL_1491867, EPI_ISL_1491868, EPI_ISL_1491880, EPI_ISL_1491890, EPI_ISL_1491894, EPI_ISL_1491896, EPI_ISL_1491901, EPI_ISL_1491904, EPI_ISL_1491945, EPI_ISL_1491949 | | | |
| see above | Aegis Sciences Corporation | Centers for Disease Control and Prevention Division of Viral Diseases, Pathogen Discovery | Dakota Howard, Dhwani Batra, Peter W. Cook, Kara Moser, Adrian Paskey, Jason Caravas, Benjamin Rambo-Martin, Shatavia Morrison, Christopher Gulvick, Scott Sammons, Yvette Unoarumhi, Darlene Wagner, Matthew Schmerer, Cyndi Clark, Patrick Campbell, Rob Case, Vikramsinha Ghorpade, Holly Houdeshell, Ola Kvalvaag, Dillon Nall, Ethan Sanders, Alec Vest, Shaun Westlund, Matthew Hardison, Clinton R. Paden, Duncan MacCannell |
| EPI_ISL_1491953, EPI_ISL_1491954, EPI_ISL_1491959, EPI_ISL_1491963, EPI_ISL_1491975, EPI_ISL_1491994, EPI_ISL_1492005, EPI_ISL_1492006, EPI_ISL_1492007, EPI_ISL_1492013, EPI_ISL_1492018, EPI_ISL_1492026, EPI_ISL_1492118, EPI_ISL_1492131 | | | |
| see above | Quest Diagnostics Incorporated | Centers for Disease Control and Prevention Division of Viral Diseases, Pathogen Discovery | Dakota Howard, Dhwani Batra, Peter W. Cook, Kara Moser, Adrian Paskey, Jason Caravas, Benjamin Rambo-Martin, Shatavia Morrison, Christopher Gulvick, Scott Sammons, Yvette Unoarumhi, Darlene Wagner, Matthew Schmerer, S. H. Rosenthal, A. Gerasimova, R. M. Kagan, B. Anderson, M. Hua, Y. Liu, L.E. Bernstein, K.E. Livingston, A. Perez, I. A. Shlyakhter, R. V. Rolando, R. Owen, P. Tanpaiboon, F. Lacbawan, Clinton R. Paden, Duncan MacCannell |
| EPI_ISL_1492456 | LESP Jalisco | Instituto de diagnóstico y Referencia Epidemiologicos (INDRE) | Claudia Wong-Arambula, Abril Rodriguez-Maldonado, Vanessa Rivero-Arredondo, Ariadna Medina-Benitez, Joaquin Quiroz-Mercado, Sergio Rangel-Guerrero, Natividad Cruz-Ortiz, Tatiana Nunez-Garcia, Gisela Barrera-Badillo, Lucia Hernandez-Rivas, Irma Lopez-Martinez, Ernesto Ramirez-Gonzalez. |
| EPI_ISL_1492629 | Idaho Bureau of Laboratories | Idaho Bureau of Laboratories | "R. Beukelman, Matthew Charles Burns, Aimee Ceniseros, Robert L. Voermans, Christopher Ball" |
| EPI_ISL_1493364 | Oregon State Public Health Laboratory | Oregon State Public Health Laboratory | Rafia Razzaque, Eugene Yeboah, Vanda Makris, Laura Tsaknaridis, John Fontana and Shane Sevey |
| EPI_ISL_1493450, EPI_ISL_1493458, EPI_ISL_1493460, EPI_ISL_1493471, EPI_ISL_1493476, EPI_ISL_1493483, EPI_ISL_1493484, EPI_ISL_1493489, EPI_ISL_1493492 | NJDOH, Public Health and Environmental Laboratories | NJ_PHEL | Lindsey Bodnar, Shiv K. Verma, Dana Woell, Byeong Jeong |
| EPI_ISL_1500241, EPI_ISL_1500242 | Illinois Department of Public Health - Springfield Lab | Illinois Department of Public Health - Springfield Lab | Bryan Sim, Gordon McCall |
| EPI_ISL_1500596 | Clinical Microbiology Laboratory, NewYork Presbyterian Hospital/Columbia University Irving Medical Center | Uhlemann Laboratory, Columbia University Irving Medical Center | Medini K. Annavajhala, Anne-Catrin Uhlemann, Hiroshi Mohri, David Ho |
| EPI_ISL_1500650, EPI_ISL_1500768 | Sonora Quest Laboratories | TGen North | "Jolene Bowers, Heather Centner, Chris French, Hayley Yaglom, Ashlyn Pfeiffer, Darrin Lemmer, Dave Engelthaler, The Arizona COVID Genomics Union (ACGU)" |
| EPI_ISL_1501687 | Northwestern Memorial Hospital | Northwestern University - Ozer Lab | Ramon Lorenzo-Redondo, Lacy M. Simons, Taylor J. Dean, Chad J. Achenbach, Lawrence J. Jennings, Chao Qi, Michael G. Ison, Judd F. Hultquist, Egon A. Ozer |
| EPI_ISL_1503483, EPI_ISL_1503791 | Massachusetts State Public Health Laboratory | Massachusetts State Public Health Laboratory | Andrew Lang, Timelia Fink, Glen Gallagher, Sandra Smole |
| EPI_ISL_1509105, EPI_ISL_1509106, EPI_ISL_1509107, EPI_ISL_1509110, EPI_ISL_1509114, EPI_ISL_1509117, EPI_ISL_1509122, EPI_ISL_1509123, EPI_ISL_1509126, EPI_ISL_1509127, EPI_ISL_1509132, EPI_ISL_1509134, EPI_ISL_1509135, EPI_ISL_1509139, EPI_ISL_1509140, EPI_ISL_1509143, EPI_ISL_1509146, EPI_ISL_1509147, EPI_ISL_1509148, EPI_ISL_1509149, EPI_ISL_1509150, EPI_ISL_1509151, EPI_ISL_1509152, EPI_ISL_1509154, EPI_ISL_1509155, EPI_ISL_1509159, EPI_ISL_1509160, EPI_ISL_1509162, EPI_ISL_1509163, EPI_ISL_1509166, EPI_ISL_1509167, EPI_ISL_1509168, EPI_ISL_1509172, EPI_ISL_1509175, EPI_ISL_1509176, EPI_ISL_1509179, EPI_ISL_1509184, EPI_ISL_1509187, EPI_ISL_1509189, EPI_ISL_1509197, EPI_ISL_1509198, EPI_ISL_1509199, EPI_ISL_1509200, EPI_ISL_1509202, EPI_ISL_1509203, EPI_ISL_1509204, EPI_ISL_1509205, EPI_ISL_1509215, EPI_ISL_1509219, EPI_ISL_1509222, EPI_ISL_1509223, EPI_ISL_1509224, EPI_ISL_1509225, EPI_ISL_1509230, EPI_ISL_1509232, EPI_ISL_1509233, EPI_ISL_1509236, EPI_ISL_1509241, EPI_ISL_1509249, EPI_ISL_1509253, EPI_ISL_1509254, EPI_ISL_1509255, EPI_ISL_1509258 | | | |
| see above | Yale Clinical Virology Lab | Grubaugh Lab - Yale School of Public Health | Joseph Fauver, Mallery Breban, Isabell Ott, Tara Alpert, Mary Petrone, Anderson Brito, Chantal Vogels, Annie Watkins, Chaney Kalinich, Jessica Rothman, Marie L. Landry, Nathan Grubaugh |
| EPI_ISL_1509266, EPI_ISL_1509267 | Murphy Medical Association | Grubaugh Lab - Yale School of Public Health | Joseph Fauver, Mallery Breban, Isabel Ott, Tara Alpert, Mary Petrone, Anderson Brito, Chantal Vogels, Annie Watkins, Chaney Kalinich, Jessica Rothman, |

| | | | |
|---|---|---|---|
| | | | Caleb Neal, Eva Laszlo, Steven Murphy, Nathan Grubaugh |
| EPI_ISL_1509271 | Grubaugh Lab - Yale School of Public Health | Grubaugh Lab - Yale School of Public Health | Joseph Fauver, Mallery Breban, Isabell Ott, Tara Alpert, Mary Petrone, Anderson Brito, Chantal Vogels, Annie Watkins, Chaney Kalinich, Jessica Rothman, Nathan Grubaugh |
| EPI_ISL_1509275, EPI_ISL_1509276 | USC Clinical Lab | Los Angeles County PHL | P. Hemarajata et al. |
| EPI_ISL_1511688, EPI_ISL_1511694, EPI_ISL_1511717, EPI_ISL_1511770, EPI_ISL_1511776, EPI_ISL_1511779, EPI_ISL_1511781, EPI_ISL_1511789, EPI_ISL_1511791, EPI_ISL_1511794, EPI_ISL_1511804, EPI_ISL_1511808, EPI_ISL_1511812, EPI_ISL_1511817, EPI_ISL_1511824, EPI_ISL_1511826, EPI_ISL_1511830, EPI_ISL_1511834, EPI_ISL_1511836, EPI_ISL_1511847, EPI_ISL_1511850, EPI_ISL_1511852, EPI_ISL_1511856, EPI_ISL_1511858 | | | |
| see above | Quest Diagnostics Incorporated | Centers for Disease Control and Prevention Division of Viral Diseases, Pathogen Discovery | Dakota Howard, Dhwani Batra, Peter W. Cook, Kara Moser, Adrian Paskey, Jason Caravas, Benjamin Rambo-Martin, Shatavia Morrison, Christopher Gulvick, Scott Sammons, Yvette Unoarumhi, Darlene Wagner, Matthew Schmerer, S. H. Rosenthal, A. Gerasimova, R. M. Kagan, B. Anderson, M. Hua, Y. Liu, L.E. Bernstein, K.E. Livingston, A. Perez, I. A. Shlyakhter, R. V. Rolando, R. Owen, P. Tanpaiboon, F. Lacbawan, Clinton R. Paden, Duncan MacCannell |
| EPI_ISL_1511882, EPI_ISL_1511904, EPI_ISL_1511977, EPI_ISL_1511998, EPI_ISL_1511999, EPI_ISL_1512000, EPI_ISL_1512001, EPI_ISL_1512062, EPI_ISL_1512126, EPI_ISL_1512151, EPI_ISL_1512205, EPI_ISL_1512245, EPI_ISL_1512317, EPI_ISL_1512331, EPI_ISL_1512427, EPI_ISL_1512459, EPI_ISL_1512460, EPI_ISL_1512527, EPI_ISL_1512646, EPI_ISL_1512719, EPI_ISL_1512736 | | | |
| see above | Helix/Illumina | Centers for Disease Control and Prevention Division of Viral Diseases, Pathogen Discovery | Dakota Howard, Dhwani Batra, Peter W. Cook, Kara Moser, Adrian Paskey, Jason Caravas, Benjamin Rambo-Martin, Shatavia Morrison, Christopher Gulvick, Scott Sammons, Yvette Unoarumhi, Darlene Wagner, Matthew Schmerer, Eileen de Feo, Jan Antico, Christine Tran, Matthew Tolentino, Shannon Wickline, Kim Gietzen, Brad Sickler, Jingtao Liu, Eric Allen, Phil Febbo, Nicole L. Washington, Simon White, Geraint Levan, Kelly Schiabor Barrett, Elizabeth Cirulli, Alexandre Bolze, Ary Ascencio, Charlotte Rivera-Garcia, Ryan Cho, Jason Nguyen, Sherry Wang, Jimmy Ramirez, Tyler Cassens, Efren Sandoval, Magnus Isaksson, William Lee, David Becker, Marc Laurent, James Lu, Clinton R. Paden, Duncan MacCannell |
| EPI_ISL_1512822, EPI_ISL_1512840, EPI_ISL_1512841, EPI_ISL_1512847, EPI_ISL_1512854, EPI_ISL_1512886, EPI_ISL_1512888, EPI_ISL_1512889, EPI_ISL_1512892, EPI_ISL_1512901, EPI_ISL_1512904, EPI_ISL_1512911, EPI_ISL_1512923, EPI_ISL_1512934, EPI_ISL_1512948, EPI_ISL_1512949, EPI_ISL_1513022, EPI_ISL_1513037, EPI_ISL_1513050, EPI_ISL_1513055, EPI_ISL_1513060, EPI_ISL_1513068, EPI_ISL_1513071, EPI_ISL_1513073, EPI_ISL_1513083, EPI_ISL_1513086, EPI_ISL_1513101, EPI_ISL_1513111, EPI_ISL_1513118, EPI_ISL_1513128, EPI_ISL_1513129, EPI_ISL_1513131 | | | |
| see above | Infinity Biologix | Centers for Disease Control and Prevention Division of Viral Diseases, Pathogen Discovery | Dakota Howard, Dhwani Batra, Peter W. Cook, Kara Moser, Adrian Paskey, Jason Caravas, Benjamin Rambo-Martin, Shatavia Morrison, Christopher Gulvick, Scott Sammons, Yvette Unoarumhi, Darlene Wagner, Matthew Schmerer, Christian Bixby, Yihe Wang, Jonathan Schultz, Chirayu Goswami, Russ Hager, Robin Grimwood, Clinton R. Paden, Duncan MacCannell |
| EPI_ISL_1513141, EPI_ISL_1513151, EPI_ISL_1513156, EPI_ISL_1513166, EPI_ISL_1513212, EPI_ISL_1513223, EPI_ISL_1513231, EPI_ISL_1513235, EPI_ISL_1513329, EPI_ISL_1513331, EPI_ISL_1513338, EPI_ISL_1513390, EPI_ISL_1513482, EPI_ISL_1513486, EPI_ISL_1513519, EPI_ISL_1513562, EPI_ISL_1513564, EPI_ISL_1513567, EPI_ISL_1513568, EPI_ISL_1513575, EPI_ISL_1513577, EPI_ISL_1513581, EPI_ISL_1513602, EPI_ISL_1513605, EPI_ISL_1513616, EPI_ISL_1513620 | | | |
| see above | Aegis Sciences Corporation | Centers for Disease Control and Prevention Division of Viral Diseases, Pathogen Discovery | Dakota Howard, Dhwani Batra, Peter W. Cook, Kara Moser, Adrian Paskey, Jason Caravas, Benjamin Rambo-Martin, Shatavia Morrison, Christopher Gulvick, Scott Sammons, Yvette Unoarumhi, Darlene Wagner, Matthew Schmerer, Cyndi Clark, Patrick Campbell, Rob Case, Vikramsinha Ghorpade, Holly Houdeshell, Ola Kvalvaag, Dillon Nall, Ethan Sanders, Alec Vest, Shaun Westlund, Matthew Hardison, Clinton R. Paden, Duncan MacCannell |
| EPI_ISL_1513637, EPI_ISL_1513638, EPI_ISL_1513639, EPI_ISL_1513643, EPI_ISL_1513647, EPI_ISL_1513654, EPI_ISL_1513680, EPI_ISL_1513849, EPI_ISL_1513883, EPI_ISL_1513903, EPI_ISL_1513943, EPI_ISL_1513954, EPI_ISL_1513992, EPI_ISL_1513994, EPI_ISL_1514031, EPI_ISL_1514085, EPI_ISL_1514147, EPI_ISL_1514213, EPI_ISL_1514219, EPI_ISL_1514220, EPI_ISL_1514278, EPI_ISL_1514284, EPI_ISL_1514317, EPI_ISL_1514320, EPI_ISL_1514323, EPI_ISL_1514329, EPI_ISL_1514345, EPI_ISL_1514348, EPI_ISL_1514353, EPI_ISL_1514354, EPI_ISL_1514355, EPI_ISL_1514357, EPI_ISL_1514364, EPI_ISL_1514443, EPI_ISL_1514532, EPI_ISL_1514544, EPI_ISL_1514591, EPI_ISL_1514612, EPI_ISL_1514621, EPI_ISL_1514627, EPI_ISL_1514632, EPI_ISL_1514638, EPI_ISL_1514640, EPI_ISL_1514642, EPI_ISL_1514644, EPI_ISL_1514651, EPI_ISL_1514652, EPI_ISL_1514664, EPI_ISL_1514669, EPI_ISL_1514681, EPI_ISL_1514686, EPI_ISL_1514691, EPI_ISL_1514692, EPI_ISL_1514695, EPI_ISL_1514696, EPI_ISL_1514703, EPI_ISL_1514716, EPI_ISL_1514787, EPI_ISL_1514890, EPI_ISL_1514896, EPI_ISL_1514898, EPI_ISL_1514941, EPI_ISL_1514980, EPI_ISL_1515004, EPI_ISL_1515018, EPI_ISL_1515025, EPI_ISL_1515075, EPI_ISL_1515076, EPI_ISL_1515083, EPI_ISL_1515089, EPI_ISL_1515097, EPI_ISL_1515098, EPI_ISL_1515106, EPI_ISL_1515108, EPI_ISL_1515133, EPI_ISL_1515155, EPI_ISL_1515249, EPI_ISL_1515318, EPI_ISL_1515325, EPI_ISL_1515326, EPI_ISL_1515327, EPI_ISL_1515329, EPI_ISL_1515334, EPI_ISL_1515349, EPI_ISL_1515375, EPI_ISL_1515411, EPI_ISL_1515431, EPI_ISL_1515467, EPI_ISL_1515493, EPI_ISL_1515518, EPI_ISL_1515519, EPI_ISL_1515590, EPI_ISL_1515703 | | | |
| see above | Laboratory Corporation of America | Centers for Disease Control and Prevention Division of Viral Diseases, Pathogen Discovery | Dakota Howard, Dhwani Batra, Peter W. Cook, Kara Moser, Adrian Paskey, Jason Caravas, Benjamin Rambo-Martin, Shatavia Morrison, Christopher Gulvick, Scott Sammons, Yvette Unoarumhi, Darlene Wagner, Matthew Schmerer, Minoo Agarwal, Eyad Almasri, Debbie Boles, Ayla Burns, Nuthawin Charoensri, Oren Cohen, Susan Countryman, Mary Ann Cristobal, Bobbi Croy, Suzanne Dale, Hrushikesh Deshmukh, Amanda Douglas, Vincent Drouillon, Marcia Eisenberg, Howard Engler, Rama Ghatti, Prashant Gupta, Susan Hicks, Jake Humphrey, Lax Iyer, Manoj Jain, Mohan Kolli, Brian Krueger, Tim Kuphal, Stanley Letovsky, Michael Levandoski, Craig Lukasik, Jonathan Meltzer, Brian Norvell, Mindy Nye, Scott Parker, Christos Petropoulos, John Pruitt, Steven Ragan, Scott Ryan, Mike Sapeta, Jana Schroth, Suresh Babu Selvaraju, Goran Stevovic, Amanda Suchanek, Andrea Throop, Lyndon Tilson, Thomas Urban, Joe Voshell, Kimberly Wagner, Jonathan Williams, Mary Williamson, Qian Zeng, Tricia Zwiefelhofer, Clinton R. Paden, Duncan MacCannell |
| EPI_ISL_1515810, EPI_ISL_1515996 | State Laboratories Division, Hawaii State Department of Health | State Laboratories Division, Hawaii State Department of Health | Pamela O'Brien, Drew Kuwazaki, Ayana Garnet, Razvan Sultana, Edward Desmond |
| EPI_ISL_1516197 | TN Division of Laboratory Services | Centers for Disease Control and Prevention Division of Viral Diseases, Pathogen Discovery | Mili Sheth, Sarah Nobles, Jasmine Padilla, Mark Burroughs, Shoshona Le, Katie Dillon, Peter Cook, Clinton R. Paden, Dhwani Batra, Krista Queen, Kristen Knipe, Dakota Howard, Yvette Unoarumhi, Darlene Wagner, Matthew Schmerer, Ben L. Rambo-Martin, Kristine Lacek, Sam Shepard, Alison Laufer Halpin, Dave Wentworth, Vivien Dugan, Suxiang Tong, Justin Lee |
| EPI_ISL_1516207, EPI_ISL_1516209, EPI_ISL_1516212 | NYC Department of Health and Mental Hygiene | Centers for Disease Control and Prevention Division of Viral Diseases, Pathogen Discovery | Mili Sheth, Sarah Nobles, Jasmine Padilla, Mark Burroughs, Shoshona Le, Katie Dillon, Peter Cook, Clinton R. Paden, Dhwani Batra, Krista Queen, Kristen Knipe, Dakota Howard, Yvette Unoarumhi, Darlene Wagner, Matthew Schmerer, Ben L. Rambo-Martin, Kristine Lacek, Sam Shepard, Alison Laufer Halpin, Dave Wentworth, Vivien Dugan, Suxiang Tong, Justin Lee |
| EPI_ISL_1516269 | SC Dept of Health and Env. Control-Bureau of Laboratories | Centers for Disease Control and Prevention Division of Viral Diseases, Pathogen Discovery | Mili Sheth, Sarah Nobles, Jasmine Padilla, Mark Burroughs, Shoshona Le, Katie Dillon, Peter Cook, Clinton R. Paden, Dhwani Batra, Krista Queen, Kristen Knipe, Dakota Howard, Yvette Unoarumhi, Darlene Wagner, Matthew Schmerer, Ben L. Rambo-Martin, Kristine Lacek, Sam Shepard, Alison Laufer Halpin, Dave Wentworth, Vivien Dugan, Suxiang Tong, Justin Lee |
| EPI_ISL_1516300 | OK Public Health Laboratory, Oklahoma State DOH | Centers for Disease Control and Prevention Division of Viral Diseases, Pathogen Discovery | Mili Sheth, Sarah Nobles, Jasmine Padilla, Mark Burroughs, Shoshona Le, Katie Dillon, Peter Cook, Clinton R. Paden, Dhwani Batra, Krista Queen, Kristen Knipe, Dakota Howard, Yvette Unoarumhi, Darlene Wagner, Matthew Schmerer, Ben L. Rambo-Martin, Kristine Lacek, Sam Shepard, Alison Laufer Halpin, Dave Wentworth, Vivien Dugan, Suxiang Tong, Justin Lee |
| EPI_ISL_1516473, EPI_ISL_1516484, EPI_ISL_1516532, EPI_ISL_1516540, EPI_ISL_1516574, EPI_ISL_1516584, EPI_ISL_1516717 | Broad Institute Clinical Research Sequencing Platform | Infectious Disease Program, Broad Institute of Harvard and MIT | Siddle,K.J., Adams,G., Pearlman,L., Gladden-Young,A., Vicente,G., Blumenstiel,B., DeFelice,M., Lee,M., McGovern,S., Lagerborg,K., Rudy,M., DeRuff,K., Carter,A., Normandin,E., Bauer,M., Reilly,S., Tomkins-Tinch,C., Loreth,C., Chaluvadi,S., Meldrim,J., Granger,B., Lemieux,J.E., Birren,B.W., Sabeti,P.C., Larkin,K., Dodge,S., Lennon,N., Madoff,L., Brown,C., Gallagher,G., Smole,S., Park,D.J., Gabriel,S., and MacInnis,B.L. |
| EPI_ISL_1519356 | National Public Health Laboratory, National Centre for Infectious Diseases | National Public Health Laboratory, National Centre for Infectious Diseases | Tze Minn Mak, Zhenyang Zhou, Grace Jie Yin Ngan, Royce Ang, Lin Cui, Raymond Tzer Pin Lin |
| EPI_ISL_1522072, EPI_ISL_1522073, EPI_ISL_1522122, EPI_ISL_1522131 | Dutch COVID-19 response team | National Institute for Public Health and the Environment (RIVM) | Adam Meijer, Harry Vennema, Dirk Eggink, Jeroen Cremer, Sharon van den Brink, Bas van der Veer, AnneMarie van den Brandt, Lisa Wijsman, Kim Freriks, Ryanne Jaarsma, Euníce Then, Jolienke Hardeman, Lynn Aarts, Sanne Bos, Melissa van Tuil, Robert Kohl, Linda van de Nes, Sjoerd Kuiling, James Groot, Florian Zwagemaker, Dennis Schmitz, Annelies Kroneman, Karim Hajji, Chantal Reusken, on behalf of the national COVID-19 response team |
| EPI_ISL_1524801 | National Public Health Laboratory, National Centre for Infectious Diseases | National Public Health Laboratory, National Centre for Infectious Diseases | Tze Minn Mak, Zhenyang Zhou, Grace Jie Yin Ngan, Royce Ang, Lin Cui, Raymond Tzer Pin Lin |
| EPI_ISL_1524850 | Hospital General Universitario Gregorio Marañón | Hospital General Universitario Gregorio Marañón | Sergio Buenestado Serrano, Pedro Sola Campoy, Laura Pérez-Lago, Cristina Rodriguez-Grande, Pilar Catalán, Patricia Muñoz, Darío García de Viedma |
| EPI_ISL_1525019, EPI_ISL_1525020, EPI_ISL_1525022, EPI_ISL_1525028, EPI_ISL_1525029, EPI_ISL_1525038, EPI_ISL_1525043, EPI_ISL_1525046, EPI_ISL_1525057, EPI_ISL_1525062, EPI_ISL_1525068, EPI_ISL_1525080, EPI_ISL_1525112, EPI_ISL_1525151, EPI_ISL_1525182, EPI_ISL_1525206, EPI_ISL_1525207, EPI_ISL_1525219, EPI_ISL_1525236 | | | |
| see above | Aegis Sciences Corporation | Centers for Disease Control and Prevention Division of Viral Diseases, Pathogen Discovery | Dakota Howard, Dhwani Batra, Peter W. Cook, Kara Moser, Adrian Paskey, Jason Caravas, Benjamin Rambo-Martin, Shatavia Morrison, Christopher Gulvick, Scott Sammons, Yvette Unoarumhi, Darlene Wagner, Matthew Schmerer, Cyndi Clark, Patrick Campbell, Rob Case, Vikramsinha Ghorpade, Holly Houdeshell, Ola Kvalvaag, Dillon Nall, Ethan Sanders, Alec Vest, Shaun Westlund, Matthew Hardison, Clinton R. Paden, Duncan MacCannell |
| EPI_ISL_1525267, EPI_ISL_1525283, EPI_ISL_1525557, EPI_ISL_1525591, EPI_ISL_1525602, EPI_ISL_1525746, | Fulgent Genetics | Centers for Disease Control and Prevention Division of Viral Diseases, Pathogen Discovery | Dakota Howard, Dhwani Batra, Peter W. Cook, Kara Moser, Adrian Paskey, Jason Caravas, Benjamin Rambo-Martin, Shatavia Morrison, Christopher Gulvick, Scott Sammons, Yvette Unoarumhi, Darlene Wagner, Matthew Schmerer, Harry Gao, Mickey Li, John Gao, Joseph Fierro, Benafsh Sapra, Becky Tsai, Yan Meng, Doreen Ng, James Xie, Clinton R. Paden, Duncan MacCannell |

| | | | |
|---|---|---|---|
| EPI_ISL_1525773, EPI_ISL_1525785, EPI_ISL_1525884 | | | |
| EPI_ISL_1525934, EPI_ISL_1526035, EPI_ISL_1526046, EPI_ISL_1526056 | Quest Diagnostics Incorporated | Centers for Disease Control and Prevention Division of Viral Diseases, Pathogen Discovery | Dakota Howard, Dhwani Batra, Peter W. Cook, Kara Moser, Adrian Paskey, Jason Caravas, Benjamin Rambo-Martin, Shatavia Morrison, Christopher Gulvick, Scott Sammons, Yvette Unoarumhi, Darlene Wagner, Matthew Schmerer, S. H. Rosenthal, A. Gerasimova, R. M. Kagan, B. Anderson, M. Hua, Y. Liu, L.E. Bernstein, K.E. Livingston, A. Perez, I. A. Shlyakhter, R. V. Rolando, R. Owen, P. Tanpaiboon, F. Lacbawan, Clinton R. Paden, Duncan MacCannell |
| EPI_ISL_1527031, EPI_ISL_1527058 | University of Mississippi Medical Center, Department of Pathology | University of Mississippi Medical Center, Molecular and Genomics Core Facility | Ashley C. Johnson, Ithiel J. Frame, Krishna K. Ayyalasomayajula, Michael R. Garrett, D. Ashley Robinson |
| EPI_ISL_1527121 | Massachusetts State Public Health Laboratory | Massachusetts State Public Health Laboratory | Andrew Lang, Timelia Fink, Glen Gallagher, Sandra Smole |
| EPI_ISL_1527908 | Michigan Department of Health and Human Services, Bureau of Laboratories | Michigan Department of Health and Human Services, Bureau of Laboratories | Blankenship HM, Riner D, Soehnlen MK |
| EPI_ISL_1528247 | IN State Department of Health Laboratory Services | IN State Department of Health Laboratory Services | Cassandra Campion, Jamie Yeadon, Brian Pope, Lixia Liu, Kyle Brownlee, Melissa Hindenlang, Ankita Kashikar, Mark Glazier |
| EPI_ISL_1528275 | VA-Division of Consolidated Laboratory Services | Centers for Disease Control and Prevention Division of Viral Diseases, Pathogen Discovery | Mili Sheth, Sarah Nobles, Jasmine Padilla, Mark Burroughs, Shoshona Le, Katie Dillon, Peter Cook, Clinton R. Paden, Dhwani Batra, Krista Queen, Kristen Knipe, Dakota Howard, Yvette Unoarumhi, Darlene Wagner, Matthew Schmerer, Ben L. Rambo-Martin, Kristine Lacek, Sam Shepard, Alison Laufer Halpin, Dave Wentworth, Vivien Dugan, Suxiang Tong, Justin Lee |
| EPI_ISL_1528326, EPI_ISL_1528330, EPI_ISL_1528332, EPI_ISL_1528338 | NJ Public Health and Environmental Laboratories | Centers for Disease Control and Prevention Division of Viral Diseases, Pathogen Discovery | Mili Sheth, Sarah Nobles, Jasmine Padilla, Mark Burroughs, Shoshona Le, Katie Dillon, Peter Cook, Clinton R. Paden, Dhwani Batra, Krista Queen, Kristen Knipe, Dakota Howard, Yvette Unoarumhi, Darlene Wagner, Matthew Schmerer, Ben L. Rambo-Martin, Kristine Lacek, Sam Shepard, Alison Laufer Halpin, Dave Wentworth, Vivien Dugan, Suxiang Tong, Justin Lee |
| EPI_ISL_1528413 | MS Public Health Laboratory | Centers for Disease Control and Prevention Division of Viral Diseases, Pathogen Discovery | Mili Sheth, Sarah Nobles, Jasmine Padilla, Mark Burroughs, Shoshona Le, Katie Dillon, Peter Cook, Clinton R. Paden, Dhwani Batra, Krista Queen, Kristen Knipe, Dakota Howard, Yvette Unoarumhi, Darlene Wagner, Matthew Schmerer, Ben L. Rambo-Martin, Kristine Lacek, Sam Shepard, Alison Laufer Halpin, Dave Wentworth, Vivien Dugan, Suxiang Tong, Justin Lee |
| EPI_ISL_1528427 | IN State Department of Health Laboratory Services | Centers for Disease Control and Prevention Division of Viral Diseases, Pathogen Discovery | Mili Sheth, Sarah Nobles, Jasmine Padilla, Mark Burroughs, Shoshona Le, Katie Dillon, Peter Cook, Clinton R. Paden, Dhwani Batra, Krista Queen, Kristen Knipe, Dakota Howard, Yvette Unoarumhi, Darlene Wagner, Matthew Schmerer, Ben L. Rambo-Martin, Kristine Lacek, Sam Shepard, Alison Laufer Halpin, Dave Wentworth, Vivien Dugan, Suxiang Tong, Justin Lee |
| EPI_ISL_1530665 | Sonora Quest Laboratories | TGen North | Jolene Bowers, Heather Centner, Chris French, Hayley Yaglom, Ashlyn Pfeiffer, Darrin Lemmer, Dave Engelthaler, The Arizona COVID Genomics Union (ACGU) |
| EPI_ISL_1530879, EPI_ISL_1530892, EPI_ISL_1530920, EPI_ISL_1530925, EPI_ISL_1530929, EPI_ISL_1530930, EPI_ISL_1530936 | NJDOH, Public Health and Environmental Laboratories | New Jersey Public Health and Environmental Laboratories (NJPHEL) | Lindsey Bodnar, Shiv K. Verma, Jacquelyn Deverell, Matthew Scarnati, Mohammad M Ali, Dana Woell, Byeong Jeong |
| EPI_ISL_1531089, EPI_ISL_1531110 | Florida Bureau of Public Health Laboratories | Florida Bureau of Public Health Laboratories | Sarah Schmedes, Jason Blanton |
| EPI_ISL_1531645 | M Health Fairview | Minnesota Department of Health, Public Health Laboratory | Alexandra Lorentz, Jacob Garfin, Matt Plumb, and Xiong Wang |
| EPI_ISL_1531746, EPI_ISL_1531763 | University of Oregon COVID-19 MAP Laboratory | University of Oregon Genomics and Cell Characterization Core Facility (GC3F) | Douglas Turnbull, Ariana White, Jeff Bishop, Jason Carriere |
| EPI_ISL_1532128 | Santa Clara Valley Medical Center | Santa Clara County Public Health Laboratory | Santa Clara County Public Health Department |
| EPI_ISL_1532732, EPI_ISL_1532737, EPI_ISL_1532739, EPI_ISL_1532740, EPI_ISL_1532754, EPI_ISL_1532761, EPI_ISL_1532770, EPI_ISL_1532776, EPI_ISL_1532778, EPI_ISL_1532781 | NYU Langone Health | Departments of Pathology and Medicine, New York University School of Medicine | Adriana Heguy, Dacia Dimartino, Emily Guzman, Christian Marier, Peter Meyn, Sitharam Ramaswami, Gael Westby, Paul Zappile, Yutong Zhang, Paolo Cotzia, Guiqing Wang |
| EPI_ISL_1534526 | Ministry of Health Turkey | Ministry of Health Turkey | Fatma Bayrakdar, Yasemin Cosgun, Suleyman Yalcin, Gulay Korukluoglu |
| EPI_ISL_1540721, EPI_ISL_1540722, EPI_ISL_1540723, EPI_ISL_1540733, EPI_ISL_1540734, EPI_ISL_1540736, EPI_ISL_1540747, EPI_ISL_1540750, EPI_ISL_1540757, EPI_ISL_1540759, EPI_ISL_1540761, EPI_ISL_1540768, EPI_ISL_1540771, EPI_ISL_1540774, EPI_ISL_1540775, EPI_ISL_1540777, EPI_ISL_1540782, EPI_ISL_1540783, EPI_ISL_1540785, EPI_ISL_1540790, EPI_ISL_1540794, EPI_ISL_1540798, EPI_ISL_1540801, EPI_ISL_1540805, EPI_ISL_1540806, EPI_ISL_1540811, EPI_ISL_1540827, EPI_ISL_1540828, EPI_ISL_1540829, EPI_ISL_1540847, EPI_ISL_1540848, EPI_ISL_1540849, EPI_ISL_1540851, EPI_ISL_1540852 | | | |
| see above | The Jackson Laboratory | The Jackson Laboratory | Bergeron D, Renzette N, Adams M, Omerza G, Kelly K, Long J, Li L |
| EPI_ISL_1540973, EPI_ISL_1540974, EPI_ISL_1541188 | Pathology and Laboratory Medicine Institute, Cleveland Clinic, Ohio, USA | Pathology and Laboratory Medicine Institute, Cleveland Clinic, Ohio, USA | Joy Nakitandwe, Zheng Jin Tu, Jay Brock, Yu-Wei Cheng, Gary Procop, Daniel Rhoads, Daniel H. Farkas, David Bosler |
| EPI_ISL_1542074, EPI_ISL_1542075, EPI_ISL_1542085, EPI_ISL_1542088, EPI_ISL_1542090, EPI_ISL_1542091, EPI_ISL_1542101, EPI_ISL_1542104, EPI_ISL_1542106, EPI_ISL_1542111, EPI_ISL_1542113, EPI_ISL_1542123, EPI_ISL_1542125, EPI_ISL_1542129, EPI_ISL_1542132, EPI_ISL_1542133, EPI_ISL_1542146, EPI_ISL_1542152, EPI_ISL_1542167, EPI_ISL_1542174, EPI_ISL_1542183, EPI_ISL_1542189, EPI_ISL_1542190, EPI_ISL_1542203, EPI_ISL_1542208, EPI_ISL_1542211, EPI_ISL_1542215, EPI_ISL_1542219, EPI_ISL_1542222, EPI_ISL_1542237, EPI_ISL_1542240, EPI_ISL_1542253, EPI_ISL_1542255, EPI_ISL_1542256, EPI_ISL_1542265, EPI_ISL_1542269, EPI_ISL_1542275, EPI_ISL_1542276, EPI_ISL_1542277, EPI_ISL_1542279, EPI_ISL_1542282, EPI_ISL_1542285, EPI_ISL_1542287, EPI_ISL_1542288, EPI_ISL_1542296, EPI_ISL_1542299, EPI_ISL_1542305, EPI_ISL_1542308, EPI_ISL_1542311, EPI_ISL_1542312, EPI_ISL_1542317, EPI_ISL_1542321, EPI_ISL_1542327, EPI_ISL_1542332, EPI_ISL_1542336, EPI_ISL_1542346, EPI_ISL_1542347, EPI_ISL_1542349, EPI_ISL_1542353, EPI_ISL_1542364, EPI_ISL_1542368, EPI_ISL_1542369, EPI_ISL_1542371, EPI_ISL_1542378, EPI_ISL_1542387, EPI_ISL_1542392, EPI_ISL_1542396, EPI_ISL_1542397, EPI_ISL_1542413, EPI_ISL_1542419, EPI_ISL_1542422, EPI_ISL_1542426, EPI_ISL_1542431, EPI_ISL_1542433, EPI_ISL_1542434, EPI_ISL_1542436, EPI_ISL_1542446, EPI_ISL_1542450, EPI_ISL_1542461, EPI_ISL_1542463, EPI_ISL_1542466, EPI_ISL_1542468, EPI_ISL_1542477, EPI_ISL_1542482, EPI_ISL_1542486, EPI_ISL_1542495, EPI_ISL_1542496, EPI_ISL_1542498, EPI_ISL_1542501, EPI_ISL_1542502, EPI_ISL_1542506, EPI_ISL_1542531, EPI_ISL_1542539, EPI_ISL_1542543, EPI_ISL_1542548, EPI_ISL_1542555, EPI_ISL_1542567, EPI_ISL_1542575, EPI_ISL_1542587, EPI_ISL_1542589, EPI_ISL_1542590, EPI_ISL_1542592, EPI_ISL_1542595, EPI_ISL_1542598, EPI_ISL_1542621, EPI_ISL_1542623, EPI_ISL_1542636, EPI_ISL_1542637, EPI_ISL_1542641, EPI_ISL_1542645, EPI_ISL_1542649, EPI_ISL_1542659, EPI_ISL_1542668, EPI_ISL_1542670, EPI_ISL_1542675, EPI_ISL_1542678, EPI_ISL_1542682, EPI_ISL_1542689, EPI_ISL_1542706, EPI_ISL_1542709, EPI_ISL_1542717, EPI_ISL_1542723, EPI_ISL_1542736, EPI_ISL_1542738, EPI_ISL_1542743, EPI_ISL_1542751, EPI_ISL_1542755, EPI_ISL_1542757, EPI_ISL_1542762, EPI_ISL_1542773, EPI_ISL_1542776, EPI_ISL_1542777, EPI_ISL_1542779, EPI_ISL_1542787, EPI_ISL_1542794, EPI_ISL_1542801, EPI_ISL_1542802, EPI_ISL_1542805, EPI_ISL_1542811, EPI_ISL_1542814, EPI_ISL_1542821, EPI_ISL_1542822, EPI_ISL_1542823, EPI_ISL_1542826, EPI_ISL_1542835, EPI_ISL_1542836, EPI_ISL_1542837, EPI_ISL_1542838, EPI_ISL_1542840, EPI_ISL_1542844, EPI_ISL_1542863, EPI_ISL_1542868, EPI_ISL_1542882, EPI_ISL_1542884, EPI_ISL_1542891, EPI_ISL_1542901, EPI_ISL_1542922, EPI_ISL_1542928, EPI_ISL_1542934, EPI_ISL_1542935, EPI_ISL_1542940, EPI_ISL_1542957, EPI_ISL_1542958, EPI_ISL_1542961, EPI_ISL_1542967, EPI_ISL_1542972, EPI_ISL_1542973, EPI_ISL_1542977, EPI_ISL_1542987, EPI_ISL_1542988, EPI_ISL_1542993, EPI_ISL_1542998, EPI_ISL_1543001, EPI_ISL_1543011, EPI_ISL_1543014, EPI_ISL_1543017, EPI_ISL_1543020, EPI_ISL_1543033, EPI_ISL_1543039, EPI_ISL_1543040, EPI_ISL_1543041, EPI_ISL_1543046, EPI_ISL_1543047, EPI_ISL_1543050, EPI_ISL_1543051, EPI_ISL_1543054, EPI_ISL_1543064, EPI_ISL_1543066, EPI_ISL_1543068, EPI_ISL_1543080, EPI_ISL_1543087, EPI_ISL_1543094, EPI_ISL_1543095, EPI_ISL_1543096, EPI_ISL_1543104, EPI_ISL_1543106, EPI_ISL_1543111, EPI_ISL_1543112, EPI_ISL_1543116, EPI_ISL_1543128, EPI_ISL_1543131, EPI_ISL_1543135, EPI_ISL_1543152, EPI_ISL_1543154, EPI_ISL_1543157, EPI_ISL_1543161, EPI_ISL_1543168, EPI_ISL_1543169, EPI_ISL_1543174, EPI_ISL_1543183, EPI_ISL_1543184, EPI_ISL_1543192, EPI_ISL_1543193, EPI_ISL_1543198, EPI_ISL_1543210, EPI_ISL_1543221, EPI_ISL_1543226, EPI_ISL_1543227, EPI_ISL_1543228, EPI_ISL_1543232, EPI_ISL_1543234, EPI_ISL_1543235, EPI_ISL_1543242, EPI_ISL_1543250, EPI_ISL_1543253, EPI_ISL_1543261, EPI_ISL_1543263, EPI_ISL_1543265, EPI_ISL_1543269, EPI_ISL_1543272, EPI_ISL_1543276, EPI_ISL_1543285, EPI_ISL_1543291, EPI_ISL_1543300, EPI_ISL_1543302, EPI_ISL_1543304, EPI_ISL_1543309, EPI_ISL_1543319, EPI_ISL_1543326, EPI_ISL_1543329, EPI_ISL_1543333, EPI_ISL_1543334, EPI_ISL_1543339, EPI_ISL_1543348, EPI_ISL_1543356, EPI_ISL_1543369, EPI_ISL_1543373, EPI_ISL_1543374, EPI_ISL_1543383, EPI_ISL_1543384, EPI_ISL_1543388, EPI_ISL_1543401, EPI_ISL_1543405, EPI_ISL_1543407, EPI_ISL_1543409, EPI_ISL_1543417, EPI_ISL_1543427, EPI_ISL_1543437, EPI_ISL_1543439, EPI_ISL_1543446, EPI_ISL_1543449, EPI_ISL_1543452, EPI_ISL_1543453, EPI_ISL_1543455, EPI_ISL_1543458, EPI_ISL_1543464, EPI_ISL_1543465, EPI_ISL_1543469, EPI_ISL_1543476, EPI_ISL_1543478, EPI_ISL_1543482, EPI_ISL_1543492, EPI_ISL_1543499, EPI_ISL_1543500, EPI_ISL_1543503, EPI_ISL_1543504, EPI_ISL_1543505, EPI_ISL_1543515, EPI_ISL_1543516, EPI_ISL_1543530, EPI_ISL_1543536, EPI_ISL_1543541, EPI_ISL_1543542, EPI_ISL_1543546, EPI_ISL_1543556, EPI_ISL_1543560, EPI_ISL_1543577, EPI_ISL_1543589, EPI_ISL_1543596, EPI_ISL_1543599, EPI_ISL_1543600, EPI_ISL_1543604, EPI_ISL_1543610, EPI_ISL_1543611, EPI_ISL_1543612, EPI_ISL_1543626, EPI_ISL_1543637, EPI_ISL_1543643, EPI_ISL_1543650, EPI_ISL_1543652, EPI_ISL_1543658, EPI_ISL_1543668, EPI_ISL_1543671, EPI_ISL_1543682, EPI_ISL_1543683, EPI_ISL_1543687, EPI_ISL_1543707, EPI_ISL_1543712, EPI_ISL_1543713, EPI_ISL_1543714, EPI_ISL_1543718, EPI_ISL_1543722, EPI_ISL_1543730, EPI_ISL_1543738, EPI_ISL_1543742, EPI_ISL_1543745, EPI_ISL_1543750, EPI_ISL_1543760, EPI_ISL_1543765, EPI_ISL_1543767, EPI_ISL_1543771, EPI_ISL_1543776, EPI_ISL_1543777, EPI_ISL_1543783, EPI_ISL_1543784, EPI_ISL_1543800, EPI_ISL_1543805, EPI_ISL_1543812, EPI_ISL_1543815, EPI_ISL_1543816, EPI_ISL_1543819, EPI_ISL_1543824, EPI_ISL_1543831, EPI_ISL_1543834, EPI_ISL_1543835, EPI_ISL_1543843, EPI_ISL_1543844, EPI_ISL_1543849, EPI_ISL_1543859, EPI_ISL_1543860, EPI_ISL_1543861, EPI_ISL_1543883, EPI_ISL_1543884, EPI_ISL_1543890, EPI_ISL_1543905, EPI_ISL_1543910, EPI_ISL_1543917, EPI_ISL_1543918, EPI_ISL_1543920 | | | |
| see above | Pandemic Response Lab - NYC | Pandemic Response Lab, R&D | Henry Lee, Michael Hammerling, Melissa Hopkins, Cybill del Castillo, Shinyoung Clair Kang, William Ward, Pradeep Bugga, Sol Rey, Dylan Law, Katharine Nelson, Haiping Hao, Jon Laurent |
| EPI_ISL_1559340, EPI_ISL_1559358 | NJDOH, Public Health and Environmental Laboratories | NJ_PHEL | Lindsey Bodnar, Shiv K. Verma, Jacquelyn Deverell, Matthew Scarnati, Mohammad M Ali, Dana Woell, Byeong Jeong |
| EPI_ISL_1559422, EPI_ISL_1559441, EPI_ISL_1559514, EPI_ISL_1559541 | Quest Diagnostics Incorporated | Centers for Disease Control and Prevention Division of Viral Diseases, Pathogen Discovery | Dakota Howard, Dhwani Batra, Peter W. Cook, Kara Moser, Adrian Paskey, Jason Caravas, Benjamin Rambo-Martin, Shatavia Morrison, Christopher Gulvick, Scott Sammons, Yvette Unoarumhi, Darlene Wagner, Matthew Schmerer, S. H. Rosenthal, A. Gerasimova, R. M. Kagan, B. Anderson, M. Hua, Y. Liu, L.E. Bernstein, K.E. Livingston, A. Perez, I. A. Shlyakhter, R. V. Rolando, R. Owen, P. Tanpaiboon, F. Lacbawan, Clinton R. Paden, Duncan MacCannell |
| EPI_ISL_1559631, EPI_ISL_1559633, EPI_ISL_1559640, EPI_ISL_1559658, EPI_ISL_1559675, EPI_ISL_1559678, EPI_ISL_1559679, EPI_ISL_1559688, EPI_ISL_1559705, EPI_ISL_1559706, EPI_ISL_1559710, EPI_ISL_1559718, EPI_ISL_1559787, EPI_ISL_1559796, EPI_ISL_1559797, EPI_ISL_1559814, EPI_ISL_1559821, | | | |

| | | | |
|---|---|---|---|
| EPI_ISL_1559823, EPI_ISL_1559929, EPI_ISL_1559969, EPI_ISL_1560017, EPI_ISL_1560024, EPI_ISL_1560028, EPI_ISL_1560060, EPI_ISL_1560074, EPI_ISL_1560187, EPI_ISL_1560231, EPI_ISL_1560313, EPI_ISL_1560358, EPI_ISL_1560380, EPI_ISL_1560405, EPI_ISL_1560415, EPI_ISL_1560418, EPI_ISL_1560539, EPI_ISL_1560630, EPI_ISL_1560676, EPI_ISL_1560820, EPI_ISL_1560821, EPI_ISL_1560859, EPI_ISL_1560901, EPI_ISL_1560968, EPI_ISL_1561017, EPI_ISL_1561019, EPI_ISL_1561154, EPI_ISL_1561241, EPI_ISL_1561242, EPI_ISL_1561244, EPI_ISL_1561266, EPI_ISL_1561285, EPI_ISL_1561319, EPI_ISL_1561326, EPI_ISL_1561405, EPI_ISL_1561407, EPI_ISL_1561425, EPI_ISL_1561433, EPI_ISL_1561452, EPI_ISL_1561515, EPI_ISL_1561536, EPI_ISL_1561557, EPI_ISL_1561570, EPI_ISL_1561667, EPI_ISL_1561675, EPI_ISL_1561676, EPI_ISL_1561700, EPI_ISL_1561721, EPI_ISL_1561840, EPI_ISL_1561842, EPI_ISL_1561845, EPI_ISL_1561846, EPI_ISL_1561853, EPI_ISL_1561854, EPI_ISL_1561857, EPI_ISL_1561890, EPI_ISL_1561940, EPI_ISL_1561966, EPI_ISL_1561977, EPI_ISL_1562016, EPI_ISL_1562056, EPI_ISL_1562082, EPI_ISL_1562085, EPI_ISL_1562113, EPI_ISL_1562117, EPI_ISL_1562119, EPI_ISL_1562150, EPI_ISL_1562154, EPI_ISL_1562185, EPI_ISL_1562187, EPI_ISL_1562199, EPI_ISL_1562228, EPI_ISL_1562246, EPI_ISL_1562302, EPI_ISL_1562303, EPI_ISL_1562367, EPI_ISL_1562402, EPI_ISL_1562408, EPI_ISL_1562410, EPI_ISL_1562425, EPI_ISL_1562431, EPI_ISL_1562432, EPI_ISL_1562454, EPI_ISL_1562457, EPI_ISL_1562517, EPI_ISL_1562518, EPI_ISL_1562519, EPI_ISL_1562520, EPI_ISL_1562524, EPI_ISL_1562562, EPI_ISL_1562687, EPI_ISL_1562690, EPI_ISL_1562776, EPI_ISL_1562784, EPI_ISL_1562787, EPI_ISL_1562809, EPI_ISL_1562833, EPI_ISL_1562844, EPI_ISL_1562846, EPI_ISL_1562853, EPI_ISL_1562917, EPI_ISL_1562919, EPI_ISL_1562959, EPI_ISL_1562984, EPI_ISL_1563004, EPI_ISL_1563154, EPI_ISL_1563231, EPI_ISL_1563243, EPI_ISL_1563264, EPI_ISL_1563285, EPI_ISL_1563299, EPI_ISL_1563347, EPI_ISL_1563348, EPI_ISL_1563350, EPI_ISL_1563355, EPI_ISL_1563356, EPI_ISL_1563385, EPI_ISL_1563396, EPI_ISL_1563443, EPI_ISL_1563475, EPI_ISL_1563569, EPI_ISL_1563588 | | | |
| see above | Aegis Sciences Corporation | Centers for Disease Control and Prevention Division of Viral Diseases, Pathogen Discovery | Dakota Howard, Dhwani Batra, Peter W. Cook, Kara Moser, Adrian Paskey, Jason Caravas, Benjamin Rambo-Martin, Shatavia Morrison, Christopher Gulvick, Scott Sammons, Yvette Unoarumhi, Darlene Wagner, Matthew Schmerer, Cyndi Clark, Patrick Campbell, Rob Case, Vikramsinha Ghorpade, Holly Houdeshell, Ola Kvalvaag, Dillon Nall, Ethan Sanders, Alec Vest, Shaun Westlund, Matthew Hardison, Clinton R. Paden, Duncan MacCannell |
| EPI_ISL_1563646 | IN State Department of Health Laboratory Services | IN State Department of Health Laboratory Services | Cassandra Campion, Jamie Yeadon, Brian Pope, Lixia Liu, Kyle Brownlee, Melissa Hindenlang, Ankita Kashikar, Mark Glazier |
| EPI_ISL_1574987 | Edward Hines Jr VA Hospital | Illinois Department of Public Health - Chicago Lab | Vineet K. Dhiman, Ira Heimler |
| EPI_ISL_1575445 | SUNY UPSTATE MEDICAL UNIVERSITY | Wadsworth Center, New York State Department of Health | Kirsten St. George, Daryl M. Lamson, Alexis Russell, Matthew Shudt, Melissa A Leisner, Jonathan Plitnick, Catharine Prussing, Navjot Singh, John Kelly, Erasmus Schneider, Erica Lasek-Nesselquist |
| EPI_ISL_1575470 | MONTEFIORE MEDICAL CENTER LABORATORIES | Wadsworth Center, New York State Department of Health | Kirsten St. George, Daryl M. Lamson, Alexis Russell, Matthew Shudt, Melissa A Leisner, Jonathan Plitnick, Catharine Prussing, Navjot Singh, John Kelly, Erasmus Schneider, Erica Lasek-Nesselquist |
| EPI_ISL_1575501, EPI_ISL_1575538, EPI_ISL_1575573, EPI_ISL_1575742, EPI_ISL_1575818, EPI_ISL_1575826, EPI_ISL_1575998, EPI_ISL_1576164, EPI_ISL_1576277 | Helix/Illumina | Centers for Disease Control and Prevention Division of Viral Diseases, Pathogen Discovery | Dakota Howard, Dhwani Batra, Peter W. Cook, Kara Moser, Adrian Paskey, Jason Caravas, Benjamin Rambo-Martin, Shatavia Morrison, Christopher Gulvick, Scott Sammons, Yvette Unoarumhi, Darlene Wagner, Matthew Schmerer, Eileen de Feo, Jan Antico, Christine Tran, Matthew Tolentino, Shannon Wickline, Kim Gietzen, Brad Sickler, Jingtao Liu, Eric Allen, Phil Febbo, Nicole L. Washington, Simon White, Geraint Levan, Kelly Schiabor Barrett, Elizabeth Cirulli, Alexandre Bolze, Ary Ascencio, Charlotte Rivera-Garcia, Ryan Cho, Jason Nguyen, Sherry Wang, Jimmy Ramirez, Tyler Cassens, Efren Sandoval, Magnus Isaksson, William Lee, David Becker, Marc Laurent, James Lu, Clinton R. Paden, Duncan MacCannell |
| EPI_ISL_1576301 | Montana Public Health Laboratory | Montana Public Health Laboratory | Joy Ritter, Michelle Mozer, Carrie Biskupiak, Deborah Gibson |
| EPI_ISL_1576347, EPI_ISL_1576348, EPI_ISL_1576456, EPI_ISL_1576519, EPI_ISL_1576680, EPI_ISL_1576726, EPI_ISL_1576736 | Helix/Illumina | Centers for Disease Control and Prevention Division of Viral Diseases, Pathogen Discovery | Dakota Howard, Dhwani Batra, Peter W. Cook, Kara Moser, Adrian Paskey, Jason Caravas, Benjamin Rambo-Martin, Shatavia Morrison, Christopher Gulvick, Scott Sammons, Yvette Unoarumhi, Darlene Wagner, Matthew Schmerer, Eileen de Feo, Jan Antico, Christine Tran, Matthew Tolentino, Shannon Wickline, Kim Gietzen, Brad Sickler, Jingtao Liu, Eric Allen, Phil Febbo, Nicole L. Washington, Simon White, Geraint Levan, Kelly Schiabor Barrett, Elizabeth Cirulli, Alexandre Bolze, Ary Ascencio, Charlotte Rivera-Garcia, Ryan Cho, Jason Nguyen, Sherry Wang, Jimmy Ramirez, Tyler Cassens, Efren Sandoval, Magnus Isaksson, William Lee, David Becker, Marc Laurent, James Lu, Clinton R. Paden, Duncan MacCannell |
| EPI_ISL_1576824 | ALBANY MEDICAL CENTER | Wadsworth Center, New York State Department of Health | Kirsten St. George, Daryl M. Lamson, Alexis Russell, Matthew Shudt, Melissa A Leisner, Jonathan Plitnick, Catharine Prussing, Navjot Singh, John Kelly, Erasmus Schneider, Erica Lasek-Nesselquist |
| EPI_ISL_1576863, EPI_ISL_1576894, EPI_ISL_1576899, EPI_ISL_1576900, EPI_ISL_1576903, EPI_ISL_1576906, EPI_ISL_1576908, EPI_ISL_1576911, EPI_ISL_1576919, EPI_ISL_1576921, EPI_ISL_1576922, EPI_ISL_1576931, EPI_ISL_1576939, EPI_ISL_1576941, EPI_ISL_1576942 | | | |
| see above | NYU Langone Health | Departments of Pathology and Medicine, New York University School of Medicine | Adriana Heguy, Dacia Dimartino, Emily Guzman, Christian Marier, Peter Meyn, Sitharam Ramaswami, Gael Westby, Paul Zappile, Yutong Zhang, Paolo Cotzia, Guiqing Wang |
| EPI_ISL_1578053, EPI_ISL_1578081 | South Dakota Public Health Laboratory | South Dakota Public Health Laboratory | Jacob Garfin and Chris Carlson |
| EPI_ISL_1578180 | Flow Health | Infectious Disease Program, Broad Institute of Harvard and MIT | Lemieux,J.E., Siddle,K.J., Adams,G., Gladden-Young,A., Lagerborg,K., Rudy,M., DeRuff,K., Carter,A., Normandin,E., Bauer,M., Reilly,S., Tomkins-Tinch,C., Loreth,C., Chaluvadi,S., Birren,B.W., Gallagher,G., Smole,S., Park,D.J., MacInnis,B.L., and Sabeti,P.C. |
| EPI_ISL_1578211, EPI_ISL_1578358, EPI_ISL_1578359, EPI_ISL_1578360, EPI_ISL_1578367, EPI_ISL_1578373, EPI_ISL_1578374, EPI_ISL_1578376, EPI_ISL_1578388, EPI_ISL_1578396, EPI_ISL_1578401, EPI_ISL_1578403, EPI_ISL_1578404, EPI_ISL_1578405, EPI_ISL_1578406 | | | |
| see above | Broad Institute Clinical Research Sequencing Platform | Infectious Disease Program, Broad Institute of Harvard and MIT | Siddle,K.J., Adams,G., Pearlman,L., Gladden-Young,A., Vicente,G., Blumenstiel,B., DeFelice,M., Lee,M., McGovern,S., Lagerborg,K., Rudy,M., DeRuff,K., Carter,A., Normandin,E., Bauer,M., Reilly,S., Tomkins-Tinch,C., Loreth,C., Chaluvadi,S., Meldrim,J., Granger,B., Lemieux,J.E., Birren,B.W., Sabeti,P.C., Larkin,K., Dodge,S., Lennon,N., Madoff,L., Brown,C., Gallagher,G., Smole,S., Park,D.J., Gabriel,S., and MacInnis,B.L. |
| EPI_ISL_1580670 | BOSTON HEART DIAGNOSTICS CORP | Wadsworth Center, New York State Department of Health | Kirsten St. George, Daryl M. Lamson, Alexis Russell, Matthew Shudt, Melissa A Leisner, Jonathan Plitnick, Catharine Prussing, Navjot Singh, John Kelly, Erasmus Schneider, Erica Lasek-Nesselquist |
| EPI_ISL_1580689, EPI_ISL_1580690, EPI_ISL_1580693, EPI_ISL_1580696 | Columbia University Irving Medical Center | Wadsworth Center, New York State Department of Health | Kirsten St. George, Daryl M. Lamson, Alexis Russell, Matthew Shudt, Melissa A Leisner, Jonathan Plitnick, Catharine Prussing, Navjot Singh, John Kelly, Erasmus Schneider, Erica Lasek-Nesselquist |
| EPI_ISL_1580717 | URMC LABS | Wadsworth Center, New York State Department of Health | Kirsten St. George, Daryl M. Lamson, Alexis Russell, Matthew Shudt, Melissa A Leisner, Jonathan Plitnick, Catharine Prussing, Navjot Singh, John Kelly, Erasmus Schneider, Erica Lasek-Nesselquist |
| EPI_ISL_1580731, EPI_ISL_1580733, EPI_ISL_1580735 | NORTHWELL HEALTH LABORATORIES | Wadsworth Center, New York State Department of Health | Kirsten St. George, Daryl M. Lamson, Alexis Russell, Matthew Shudt, Melissa A Leisner, Jonathan Plitnick, Catharine Prussing, Navjot Singh, John Kelly, Erasmus Schneider, Erica Lasek-Nesselquist |
| EPI_ISL_1581078, EPI_ISL_1581086, EPI_ISL_1581107, EPI_ISL_1581143, EPI_ISL_1581711, EPI_ISL_1581775 | Helix/Illumina | Centers for Disease Control and Prevention Division of Viral Diseases, Pathogen Discovery | Dakota Howard, Dhwani Batra, Peter W. Cook, Kara Moser, Adrian Paskey, Jason Caravas, Benjamin Rambo-Martin, Shatavia Morrison, Christopher Gulvick, Scott Sammons, Yvette Unoarumhi, Darlene Wagner, Matthew Schmerer, Eileen de Feo, Jan Antico, Christine Tran, Matthew Tolentino, Shannon Wickline, Kim Gietzen, Brad Sickler, Jingtao Liu, Eric Allen, Phil Febbo, Nicole L. Washington, Simon White, Geraint Levan, Kelly Schiabor Barrett, Elizabeth Cirulli, Alexandre Bolze, Ary Ascencio, Charlotte Rivera-Garcia, Ryan Cho, Jason Nguyen, Sherry Wang, Jimmy Ramirez, Tyler Cassens, Efren Sandoval, Magnus Isaksson, William Lee, David Becker, Marc Laurent, James Lu, Clinton R. Paden, Duncan MacCannell |
| EPI_ISL_1581958, EPI_ISL_1582049, EPI_ISL_1582072, EPI_ISL_1582074, EPI_ISL_1582085, EPI_ISL_1582099, EPI_ISL_1582103, EPI_ISL_1582116, EPI_ISL_1582120, EPI_ISL_1582126 | Quest Diagnostics Incorporated | Centers for Disease Control and Prevention Division of Viral Diseases, Pathogen Discovery | Dakota Howard, Dhwani Batra, Peter W. Cook, Kara Moser, Adrian Paskey, Jason Caravas, Benjamin Rambo-Martin, Shatavia Morrison, Christopher Gulvick, Scott Sammons, Yvette Unoarumhi, Darlene Wagner, Matthew Schmerer, S. H. Rosenthal, A. Gerasimova, R. M. Kagan, B. Anderson, M. Hua, Y. Liu, L.E. Bernstein, K.E. Livingston, A. Perez, I. A. Shlyakhter, R. V. Rolando, R. Owen, P. Tanpaiboon, F. Lacbawan, Clinton R. Paden, Duncan MacCannell |
| EPI_ISL_1582154 | ALBANY MEDICAL CENTER | Wadsworth Center, New York State Department of Health | Kirsten St. George, Daryl M. Lamson, Alexis Russell, Matthew Shudt, Melissa A Leisner, Jonathan Plitnick, Catharine Prussing, Navjot Singh, John Kelly, Erasmus Schneider, Erica Lasek-Nesselquist |
| EPI_ISL_1582159 | MONTEFIORE MEDICAL CENTER LABORATORIES | Wadsworth Center, New York State Department of Health | Kirsten St. George, Daryl M. Lamson, Alexis Russell, Matthew Shudt, Melissa A Leisner, Jonathan Plitnick, Catharine Prussing, Navjot Singh, John Kelly, Erasmus Schneider, Erica Lasek-Nesselquist |
| EPI_ISL_1582169 | Quest Diagnostics Incorporated | Centers for Disease Control and Prevention Division of Viral Diseases, Pathogen Discovery | Dakota Howard, Dhwani Batra, Peter W. Cook, Kara Moser, Adrian Paskey, Jason Caravas, Benjamin Rambo-Martin, Shatavia Morrison, Christopher Gulvick, Scott Sammons, Yvette Unoarumhi, Darlene Wagner, Matthew Schmerer, S. H. Rosenthal, A. Gerasimova, R. M. Kagan, B. Anderson, M. Hua, Y. Liu, L.E. Bernstein, K.E. Livingston, A. Perez, I. A. Shlyakhter, R. V. Rolando, R. Owen, P. Tanpaiboon, F. Lacbawan, Clinton R. Paden, Duncan MacCannell |
| EPI_ISL_1582174, EPI_ISL_1582175, EPI_ISL_1582176 | MONTEFIORE MEDICAL CENTER LABORATORIES | Wadsworth Center, New York State Department of Health | Kirsten St. George, Daryl M. Lamson, Alexis Russell, Matthew Shudt, Melissa A Leisner, Jonathan Plitnick, Catharine Prussing, Navjot Singh, John Kelly, Erasmus Schneider, Erica Lasek-Nesselquist |
| EPI_ISL_1582181, EPI_ISL_1582183, EPI_ISL_1582184, EPI_ISL_1582188, EPI_ISL_1582200, EPI_ISL_1582204, EPI_ISL_1582207, EPI_ISL_1582208, EPI_ISL_1582210, EPI_ISL_1582213 | NYC Pandemic Response Lab | Wadsworth Center, New York State Department of Health | Kirsten St. George, Daryl M. Lamson, Alexis Russell, Matthew Shudt, Melissa A Leisner, Jonathan Plitnick, Catharine Prussing, Navjot Singh, John Kelly, Erasmus Schneider, Erica Lasek-Nesselquist |
| EPI_ISL_1582215 | Columbia University Irving Medical Center | Wadsworth Center, New York State Department of Health | Kirsten St. George, Daryl M. Lamson, Alexis Russell, Matthew Shudt, Melissa A Leisner, Jonathan Plitnick, Catharine Prussing, Navjot Singh, John Kelly, |

| | | | |
|---|---|---|---|
| | | | Erasmus Schneider, Erica Lasek-Nesselquist |
| EPI_ISL_1582229 | SARATOGA HOSPITAL LABORATORY | Wadsworth Center, New York State Department of Health | Kirsten St. George, Daryl M. Lamson, Alexis Russell, Matthew Shudt, Melissa A Leisner, Jonathan Plitnick, Catharine Prussing, Navjot Singh, John Kelly, Erasmus Schneider, Erica Lasek-Nesselquist |
| EPI_ISL_1582231 | WESTCHESTER MEDICAL CENTER | Wadsworth Center, New York State Department of Health | Kirsten St. George, Daryl M. Lamson, Alexis Russell, Matthew Shudt, Melissa A Leisner, Jonathan Plitnick, Catharine Prussing, Navjot Singh, John Kelly, Erasmus Schneider, Erica Lasek-Nesselquist |
| EPI_ISL_1582233, EPI_ISL_1582239, EPI_ISL_1582242, EPI_ISL_1582244 | TEMPUS LABS INC | Wadsworth Center, New York State Department of Health | Kirsten St. George, Daryl M. Lamson, Alexis Russell, Matthew Shudt, Melissa A Leisner, Jonathan Plitnick, Catharine Prussing, Navjot Singh, John Kelly, Erasmus Schneider, Erica Lasek-Nesselquist |
| EPI_ISL_1582248 | STONY BROOK UNIVERSITY HOSPITAL | Wadsworth Center, New York State Department of Health | Kirsten St. George, Daryl M. Lamson, Alexis Russell, Matthew Shudt, Melissa A Leisner, Jonathan Plitnick, Catharine Prussing, Navjot Singh, John Kelly, Erasmus Schneider, Erica Lasek-Nesselquist |
| EPI_ISL_1582255 | GOOD SAMARITAN HOSPITAL LABORATORY | Wadsworth Center, New York State Department of Health | Kirsten St. George, Daryl M. Lamson, Alexis Russell, Matthew Shudt, Melissa A Leisner, Jonathan Plitnick, Catharine Prussing, Navjot Singh, John Kelly, Erasmus Schneider, Erica Lasek-Nesselquist |
| EPI_ISL_1582257 | NORTH SHORE UNIVERSITY HOSPITAL | Wadsworth Center, New York State Department of Health | Kirsten St. George, Daryl M. Lamson, Alexis Russell, Matthew Shudt, Melissa A Leisner, Jonathan Plitnick, Catharine Prussing, Navjot Singh, John Kelly, Erasmus Schneider, Erica Lasek-Nesselquist |
| EPI_ISL_1582260 | ALBANY MEDICAL CENTER | Wadsworth Center, New York State Department of Health | Kirsten St. George, Daryl M. Lamson, Alexis Russell, Matthew Shudt, Melissa A Leisner, Jonathan Plitnick, Catharine Prussing, Navjot Singh, John Kelly, Erasmus Schneider, Erica Lasek-Nesselquist |
| EPI_ISL_1582263, EPI_ISL_1582268 | Columbia University Irving Medical Center | Wadsworth Center, New York State Department of Health | Kirsten St. George, Daryl M. Lamson, Alexis Russell, Matthew Shudt, Melissa A Leisner, Jonathan Plitnick, Catharine Prussing, Navjot Singh, John Kelly, Erasmus Schneider, Erica Lasek-Nesselquist |
| EPI_ISL_1582278, EPI_ISL_1582280 | WHITE PLAINS HOSPITAL CENTER LABORATORY | Wadsworth Center, New York State Department of Health | Kirsten St. George, Daryl M. Lamson, Alexis Russell, Matthew Shudt, Melissa A Leisner, Jonathan Plitnick, Catharine Prussing, Navjot Singh, John Kelly, Erasmus Schneider, Erica Lasek-Nesselquist |
| EPI_ISL_1582288 | NORTHWELL HEALTH LABORATORIES | Wadsworth Center, New York State Department of Health | Kirsten St. George, Daryl M. Lamson, Alexis Russell, Matthew Shudt, Melissa A Leisner, Jonathan Plitnick, Catharine Prussing, Navjot Singh, John Kelly, Erasmus Schneider, Erica Lasek-Nesselquist |
| EPI_ISL_1582309, EPI_ISL_1582310, EPI_ISL_1582313, EPI_ISL_1582315 | WESTCHESTER MEDICAL CENTER | Wadsworth Center, New York State Department of Health | Kirsten St. George, Daryl M. Lamson, Alexis Russell, Matthew Shudt, Melissa A Leisner, Jonathan Plitnick, Catharine Prussing, Navjot Singh, John Kelly, Erasmus Schneider, Erica Lasek-Nesselquist |
| EPI_ISL_1582319, EPI_ISL_1582320 | MONTEFIORE MEDICAL CENTER LABORATORIES | Wadsworth Center, New York State Department of Health | Kirsten St. George, Daryl M. Lamson, Alexis Russell, Matthew Shudt, Melissa A Leisner, Jonathan Plitnick, Catharine Prussing, Navjot Singh, John Kelly, Erasmus Schneider, Erica Lasek-Nesselquist |
| EPI_ISL_1582328, EPI_ISL_1582329, EPI_ISL_1582335, EPI_ISL_1582336 | NYC Pandemic Response Lab | Wadsworth Center, New York State Department of Health | Kirsten St. George, Daryl M. Lamson, Alexis Russell, Matthew Shudt, Melissa A Leisner, Jonathan Plitnick, Catharine Prussing, Navjot Singh, John Kelly, Erasmus Schneider, Erica Lasek-Nesselquist |
| EPI_ISL_1582362 | SUNY UPSTATE MEDICAL UNIVERSITY | Wadsworth Center, New York State Department of Health | Kirsten St. George, Daryl M. Lamson, Alexis Russell, Matthew Shudt, Melissa A Leisner, Jonathan Plitnick, Catharine Prussing, Navjot Singh, John Kelly, Erasmus Schneider, Erica Lasek-Nesselquist |
| EPI_ISL_1582369, EPI_ISL_1582370, EPI_ISL_1582374, EPI_ISL_1582376, EPI_ISL_1582385 | NORTHWELL HEALTH LABORATORIES | Wadsworth Center, New York State Department of Health | Kirsten St. George, Daryl M. Lamson, Alexis Russell, Matthew Shudt, Melissa A Leisner, Jonathan Plitnick, Catharine Prussing, Navjot Singh, John Kelly, Erasmus Schneider, Erica Lasek-Nesselquist |
| EPI_ISL_1582403 | TEMPUS LABS INC | Wadsworth Center, New York State Department of Health | Kirsten St. George, Daryl M. Lamson, Alexis Russell, Matthew Shudt, Melissa A Leisner, Jonathan Plitnick, Catharine Prussing, Navjot Singh, John Kelly, Erasmus Schneider, Erica Lasek-Nesselquist |
| EPI_ISL_1585481, EPI_ISL_1585563 | Centro de Investigación Biomédica de Occidente (CIBO) | Instituto Nacional de Enfermedades Respiratorias (INER): Centro de Investigación en Enfermedades Infecciosas (CIENI) | Consorcio Mexicano de Vigilancia Genómica (CoViGen-Mex). Authors (in alphabetical order): Julio Elias Alvarado-Yaah, Carlos F. Arias, Santiago Ávila-Ríos, Víctor Hugo Borja-Aburto, Celia Boukadida, Juan Bautista Chale-Dzul, Célida Duque Molina, José Antonio Enciso-Moreno, Gloria Elena Espinosa-Ayala, Fernando Fontove-Herrera, Victor Eduardo Garcia-Arias, Concepción Grajales-Muñiz, Ricardo Grande, Alfredo Herrera-Estrella, Carla Ivón Herrera-Najera, Pavel Isa, Brenda Irasema Maldonado-Meza, Bernardo Martínez-Miguel, Margarita Matías-Florentino, María Guadalupe de Jesús Mireles-Rivera, Gloria María Molina-Salinas, Hector Montoya-Fuentes, José Esteban Muñoz-Medina, José de Jesús Nuñez-Contreras, Alicia Ocaña-Mondragón, Luis Alberto Ochoa-Carrera, Hector Esteban Paz-Juárez, Francisco Pulido, Helen Haydee Fernanda Ramírez-Plascencia, Angel Gustavo Salas-Lais, Alejandro Sanchez-Flores, Clara Esperanza Santacruz-Tinoco, María Guadalupe Santiago-Mauricio, Nelly Sélem-Mojica, Blanca Taboada, Gloria Vazquez |
| EPI_ISL_1587261, EPI_ISL_1587267, EPI_ISL_1587270, EPI_ISL_1587271, EPI_ISL_1587272, EPI_ISL_1587274, EPI_ISL_1587278, EPI_ISL_1587280, EPI_ISL_1587282, EPI_ISL_1587285, EPI_ISL_1587291, EPI_ISL_1587292, EPI_ISL_1587297, EPI_ISL_1587298, EPI_ISL_1587309, EPI_ISL_1587311, EPI_ISL_1587312, EPI_ISL_1587317, EPI_ISL_1587320, EPI_ISL_1587321, EPI_ISL_1587328, EPI_ISL_1587329, EPI_ISL_1587330, EPI_ISL_1587334, EPI_ISL_1587339, EPI_ISL_1587341, EPI_ISL_1587342, EPI_ISL_1587343, EPI_ISL_1587344, EPI_ISL_1587347, EPI_ISL_1587348, EPI_ISL_1587350, EPI_ISL_1587354, EPI_ISL_1587357, EPI_ISL_1587358, EPI_ISL_1587361, EPI_ISL_1587371, EPI_ISL_1587375, EPI_ISL_1587377, EPI_ISL_1587378, EPI_ISL_1587381, EPI_ISL_1587382, EPI_ISL_1587383, EPI_ISL_1587384, EPI_ISL_1587385, EPI_ISL_1587386, EPI_ISL_1587390, EPI_ISL_1587395, EPI_ISL_1587398, EPI_ISL_1587403, EPI_ISL_1587404, EPI_ISL_1587409, EPI_ISL_1587410, EPI_ISL_1587411, EPI_ISL_1587412, EPI_ISL_1587418, EPI_ISL_1587421, EPI_ISL_1587423, EPI_ISL_1587424, EPI_ISL_1587426, EPI_ISL_1587427 | | | |
| see above | Yale Clinical Virology Lab | Grubaugh Lab - Yale School of Public Health | Joseph Fauver, Mallery Breban, Isabel Ott, Tara Alpert, Mary Petrone, Anderson Brito, Chantal Vogels, Annie Watkins, Chaney Kalinich, Jessica Rothman, Marie L. Landry, Nathan Grubaugh |
| EPI_ISL_1587589, EPI_ISL_1587593, EPI_ISL_1587594, EPI_ISL_1587601, EPI_ISL_1587619, EPI_ISL_1587660, EPI_ISL_1587715, EPI_ISL_1587716, EPI_ISL_1587744, EPI_ISL_1587750, EPI_ISL_1587898, EPI_ISL_1587917, EPI_ISL_1587919, EPI_ISL_1587952, EPI_ISL_1587956, EPI_ISL_1587976, EPI_ISL_1587987, EPI_ISL_1588003, EPI_ISL_1588019, EPI_ISL_1588020, EPI_ISL_1588023, EPI_ISL_1588211, EPI_ISL_1588288, EPI_ISL_1588298, EPI_ISL_1588301, EPI_ISL_1588363, EPI_ISL_1588371, EPI_ISL_1588405, EPI_ISL_1588412, EPI_ISL_1588415, EPI_ISL_1588418, EPI_ISL_1588435 | | | |
| see above | Infinity Biologix | Centers for Disease Control and Prevention Division of Viral Diseases, Pathogen Discovery | Dakota Howard, Dhwani Batra, Peter W. Cook, Kara Moser, Adrian Paskey, Jason Caravas, Benjamin Rambo-Martin, Shatavia Morrison, Christopher Gulvick, Scott Sammons, Yvette Unoarumhi, Darlene Wagner, Matthew Schmerer, Christian Bixby, Yihe Wang, Jonathan Schultz, Chirayu Goswami, Russ Hager, Robin Grimwood, Clinton R. Paden, Duncan MacCannell |
| EPI_ISL_1592148 | TGen North | TGen North | Jolene Bowers, Heather Centner, Chris French, Hayley Yaglom, Ashlyn Pfeiffer, Darrin Lemmer, Dave Engelthaler, The Arizona COVID Genomics Union (ACGU) |
| EPI_ISL_1592597, EPI_ISL_1592661, EPI_ISL_1592963, EPI_ISL_1593139, EPI_ISL_1593182 | Helix/Illumina | Centers for Disease Control and Prevention Division of Viral Diseases, Pathogen Discovery | Dakota Howard, Dhwani Batra, Peter W. Cook, Kara Moser, Adrian Paskey, Jason Caravas, Benjamin Rambo-Martin, Shatavia Morrison, Christopher Gulvick, Scott Sammons, Yvette Unoarumhi, Darlene Wagner, Matthew Schmerer, Eileen de Feo, Jan Antico, Christine Tran, Matthew Tolentino, Shannon Wickline, Kim Gietzen, Brad Sickler, Jingtao Liu, Eric Allen, Phil Febbo, Nicole L. Washington, Simon White, Geraint Levan, Kelly Schiabor Barrett, Elizabeth Cirulli, Alexandre Bolze, Ary Ascencio, Charlotte Rivera-Garcia, Ryan Cho, Jason Nguyen, Sherry Wang, Jimmy Ramirez, Tyler Cassens, Efren Sandoval, Magnus Isaksson, William Lee, David Becker, Marc Laurent, James Lu, Clinton R. Paden, Duncan MacCannell |
| EPI_ISL_1593720 | DOHMH Central Harlem | New York City Public Health Laboratory | Jade Wang, et al. |
| EPI_ISL_1593741 | DOHMH Crown Heights | New York City Public Health Laboratory | Jade Wang, et al. |
| EPI_ISL_1593748 | DOHMH Jamaica | New York City Public Health Laboratory | Jade Wang, et al. |
| EPI_ISL_1593750 | DOHMH Fort Greene | New York City Public Health Laboratory | Jade Wang, et al. |
| EPI_ISL_1593757 | OCME Office Of Chief Medical Examiner | New York City Public Health Laboratory | Jade Wang, et al. |
| EPI_ISL_1593770 | DOHMH Fort Greene | New York City Public Health Laboratory | Jade Wang, et al. |
| EPI_ISL_1593811 | Wyoming Public Health Laboratory | Wyoming Public Health Laboratory | Jim Mildenberger, Wanda Manley, Noah Hull, Taylor Fearing, Lynette Gumbleton, Channing Weber, Ashley Norberg, Chayse Rowley, Marley Goetz, Brian Dominguez, Elliot Thomasson, Sam Britz, Cari Sloma, and Rob Christensen |
| EPI_ISL_1593865 | WESTCHESTER MEDICAL CENTER | Wadsworth Center, New York State Department of Health | Kirsten St. George, Daryl M. Lamson, Alexis Russell, Matthew Shudt, Melissa A Leisner, Jonathan Plitnick, Catharine Prussing, Navjot Singh, John Kelly, Erasmus Schneider, Erica Lasek-Nesselquist |
| EPI_ISL_1593868 | MONTEFIORE MEDICAL CENTER LABORATORIES | Wadsworth Center, New York State Department of Health | Kirsten St. George, Daryl M. Lamson, Alexis Russell, Matthew Shudt, Melissa A Leisner, Jonathan Plitnick, Catharine Prussing, Navjot Singh, John Kelly, Erasmus Schneider, Erica Lasek-Nesselquist |

| | | | |
|---|---|---|---|
| EPI_ISL_1593883 | NYC Pandemic Response Lab | Wadsworth Center, New York State Department of Health | Kirsten St. George, Daryl M. Lamson, Alexis Russell, Matthew Shudt, Melissa A Leisner, Jonathan Plitnick, Catharine Prussing, Navjot Singh, John Kelly, Erasmus Schneider, Erica Lasek-Nesselquist |
| EPI_ISL_1593887 | NORTHWELL HEALTH LABORATORIES | Wadsworth Center, New York State Department of Health | Kirsten St. George, Daryl M. Lamson, Alexis Russell, Matthew Shudt, Melissa A Leisner, Jonathan Plitnick, Catharine Prussing, Navjot Singh, John Kelly, Erasmus Schneider, Erica Lasek-Nesselquist |
| EPI_ISL_1594026 | University of Wisconsin-Madison AIDS Vaccine Research Laboratories | University of Wisconsin-Madison AIDS Vaccine Research Laboratories | Gage Moreno, Katarina Braun, et al. AIDS Vaccine Research Laboratories |
| EPI_ISL_1597697, EPI_ISL_1597705, EPI_ISL_1597708, EPI_ISL_1597710, EPI_ISL_1597715, EPI_ISL_1597717, EPI_ISL_1597727, EPI_ISL_1597729, EPI_ISL_1597731, EPI_ISL_1597746, EPI_ISL_1597748, EPI_ISL_1597749, EPI_ISL_1597751, EPI_ISL_1597755, EPI_ISL_1597757, EPI_ISL_1597758, EPI_ISL_1597760, EPI_ISL_1597765, EPI_ISL_1597770, EPI_ISL_1597771, EPI_ISL_1597773 | | | |
| see above | Montefiore Medical Center | Abbott | Yitz Goldstein, Amy Fox, Ana Olivo, Ana Vallari, Barbara Harris, Mary Rodgers, Todd Meyer, Gavin Cloherty |
| EPI_ISL_1608212 | North Dakota Department of Health, Public Health Laboratory | North Dakota Department of Health, Public Health Laboratory | Lisa Wingerter |
| EPI_ISL_1608805, EPI_ISL_1608880, EPI_ISL_1609009, EPI_ISL_1609049, EPI_ISL_1609071, EPI_ISL_1609076, EPI_ISL_1609077, EPI_ISL_1609110, EPI_ISL_1609150, EPI_ISL_1609154, EPI_ISL_1609168, EPI_ISL_1609170, EPI_ISL_1609177, EPI_ISL_1609204, EPI_ISL_1609238, EPI_ISL_1609240, EPI_ISL_1609241, EPI_ISL_1609244, EPI_ISL_1609249, EPI_ISL_1609253, EPI_ISL_1609255, EPI_ISL_1609259, EPI_ISL_1609265, EPI_ISL_1609276, EPI_ISL_1609291, EPI_ISL_1609306, EPI_ISL_1609307, EPI_ISL_1609309, EPI_ISL_1609315, EPI_ISL_1609316, EPI_ISL_1609319, EPI_ISL_1609325, EPI_ISL_1609329, EPI_ISL_1609331, EPI_ISL_1609332, EPI_ISL_1609337, EPI_ISL_1609341, EPI_ISL_1609354, EPI_ISL_1609367, EPI_ISL_1609385, EPI_ISL_1609402, EPI_ISL_1609412, EPI_ISL_1609417, EPI_ISL_1609419, EPI_ISL_1609424, EPI_ISL_1609435, EPI_ISL_1609450, EPI_ISL_1609451, EPI_ISL_1609457, EPI_ISL_1609465, EPI_ISL_1609472, EPI_ISL_1609495, EPI_ISL_1609497, EPI_ISL_1609498, EPI_ISL_1609499, EPI_ISL_1609501, EPI_ISL_1609504, EPI_ISL_1609511, EPI_ISL_1609513, EPI_ISL_1609521, EPI_ISL_1609527, EPI_ISL_1609533, EPI_ISL_1609537, EPI_ISL_1609600, EPI_ISL_1609603, EPI_ISL_1609612, EPI_ISL_1609622, EPI_ISL_1609626, EPI_ISL_1609656, EPI_ISL_1609668, EPI_ISL_1609680, EPI_ISL_1609693, EPI_ISL_1609703, EPI_ISL_1609779, EPI_ISL_1609796, EPI_ISL_1609797, EPI_ISL_1609800, EPI_ISL_1609808, EPI_ISL_1609812, EPI_ISL_1609824, EPI_ISL_1609876, EPI_ISL_1609877, EPI_ISL_1609880, EPI_ISL_1609890, EPI_ISL_1609903, EPI_ISL_1609914, EPI_ISL_1609922, EPI_ISL_1609928, EPI_ISL_1609931, EPI_ISL_1609943, EPI_ISL_1609952, EPI_ISL_1609956, EPI_ISL_1609962, EPI_ISL_1610003, EPI_ISL_1610034, EPI_ISL_1610036, EPI_ISL_1610113, EPI_ISL_1610156, EPI_ISL_1610206, EPI_ISL_1610218, EPI_ISL_1610238, EPI_ISL_1610249, EPI_ISL_1610320, EPI_ISL_1610367, EPI_ISL_1610441, EPI_ISL_1610521, EPI_ISL_1610602, EPI_ISL_1610675, EPI_ISL_1610721, EPI_ISL_1610741, EPI_ISL_1610866, EPI_ISL_1610951, EPI_ISL_1610953, EPI_ISL_1611115, EPI_ISL_1611219, EPI_ISL_1611221, EPI_ISL_1611244, EPI_ISL_1611250, EPI_ISL_1611252, EPI_ISL_1611269, EPI_ISL_1611296, EPI_ISL_1611314, EPI_ISL_1611327, EPI_ISL_1611332, EPI_ISL_1611353, EPI_ISL_1611365, EPI_ISL_1611366, EPI_ISL_1611368, EPI_ISL_1611376, EPI_ISL_1611378, EPI_ISL_1611381, EPI_ISL_1611399, EPI_ISL_1611428, EPI_ISL_1611433, EPI_ISL_1611435, EPI_ISL_1611437, EPI_ISL_1611438, EPI_ISL_1611442, EPI_ISL_1611464, EPI_ISL_1611467, EPI_ISL_1611528, EPI_ISL_1611530, EPI_ISL_1611538, EPI_ISL_1611539, EPI_ISL_1611543, EPI_ISL_1611547, EPI_ISL_1611549, EPI_ISL_1611552, EPI_ISL_1611564, EPI_ISL_1611569, EPI_ISL_1611570, EPI_ISL_1611571, EPI_ISL_1611611, EPI_ISL_1611626, EPI_ISL_1611631, EPI_ISL_1611632, EPI_ISL_1611664, EPI_ISL_1611689, EPI_ISL_1611691, EPI_ISL_1611693, EPI_ISL_1611711, EPI_ISL_1611726, EPI_ISL_1611744, EPI_ISL_1611760, EPI_ISL_1611788, EPI_ISL_1611795, EPI_ISL_1611814, EPI_ISL_1611820, EPI_ISL_1611823, EPI_ISL_1611844, EPI_ISL_1611848, EPI_ISL_1611851, EPI_ISL_1611865, EPI_ISL_1611897, EPI_ISL_1611898, EPI_ISL_1611908, EPI_ISL_1611916, EPI_ISL_1611917, EPI_ISL_1611922, EPI_ISL_1611923, EPI_ISL_1611926, EPI_ISL_1611928, EPI_ISL_1611931, EPI_ISL_1611932, EPI_ISL_1611940, EPI_ISL_1611957, EPI_ISL_1611961, EPI_ISL_1612038, EPI_ISL_1612111, EPI_ISL_1612119, EPI_ISL_1612170, EPI_ISL_1612212, EPI_ISL_1612284, EPI_ISL_1612329, EPI_ISL_1612453, EPI_ISL_1612471, EPI_ISL_1612677, EPI_ISL_1612747, EPI_ISL_1612750, EPI_ISL_1612806 | | | |
| see above | Laboratory Corporation of America | Centers for Disease Control and Prevention Division of Viral Diseases, Pathogen Discovery | Dakota Howard, Dhwani Batra, Peter W. Cook, Kara Moser, Adrian Paskey, Jason Caravas, Benjamin Rambo-Martin, Shatavia Morrison, Christopher Gulvick, Scott Sammons, Yvette Unoarumhi, Darlene Wagner, Matthew Schmerer, Minoo Agarwal, Eyad Almasri, Debbie Boles, Ayla Burns, Nuthawin Charoensri, Oren Cohen, Susan Countryman, Mary Ann Cristobal, Bobbi Croy, Suzanne Dale, Hrushikesh Deshmukh, Amanda Douglas, Vincent Drouillon, Marcia Eisenberg, Howard Engler, Rama Ghatti, Prashant Gupta, Susan Hicks, Jake Humphrey, Lax Iyer, Manoj Jain, Mohan Kolli, Brian Krueger, Tim Kuphal, Stanley Letovsky, Michael Levandoski, Craig Lukasik, Jonathan Meltzer, Brian Norvell, Mindy Nye, Scott Parker, Christos Petropoulos, John Pruitt, Steven Ragan, Scott Ryan, Mike Sapeta, Jana Schroth, Suresh Babu Selvaraju, Goran Stevovic, Amanda Suchanek, Andrea Throop, Lyndon Tilson, Thomas Urban, Joe Voshell, Kimberly Wagner, Jonathan Williams, Mary Williamson, Qian Zeng, Tricia Zwiefelhofer, Clinton R. Paden, Duncan MacCannell |
| EPI_ISL_1612978, EPI_ISL_1613224, EPI_ISL_1613304, EPI_ISL_1613363, EPI_ISL_1613369, EPI_ISL_1613384, EPI_ISL_1613395, EPI_ISL_1613493, EPI_ISL_1613502, EPI_ISL_1613514, EPI_ISL_1613517, EPI_ISL_1613952, EPI_ISL_1614014, EPI_ISL_1614015, EPI_ISL_1614051, EPI_ISL_1614093, EPI_ISL_1614099, EPI_ISL_1614133, EPI_ISL_1614147, EPI_ISL_1614162, EPI_ISL_1614524, EPI_ISL_1614533, EPI_ISL_1614602, EPI_ISL_1614620, EPI_ISL_1614669, EPI_ISL_1614674, EPI_ISL_1614686, EPI_ISL_1614796, EPI_ISL_1614840, EPI_ISL_1614850, EPI_ISL_1614903 | | | |
| see above | Fulgent Genetics | Centers for Disease Control and Prevention Division of Viral Diseases, Pathogen Discovery | Dakota Howard, Dhwani Batra, Peter W. Cook, Kara Moser, Adrian Paskey, Jason Caravas, Benjamin Rambo-Martin, Shatavia Morrison, Christopher Gulvick, Scott Sammons, Yvette Unoarumhi, Darlene Wagner, Matthew Schmerer, Harry Gao, Mickey Li, John Gao, Joseph Fierro, Benafsh Sapra, Becky Tsai, Yan Meng, Doreen Ng, James Xie, Clinton R. Paden, Duncan MacCannell |
| EPI_ISL_1614967, EPI_ISL_1615009, EPI_ISL_1615021, EPI_ISL_1615039 | Quest Diagnostics Incorporated | Centers for Disease Control and Prevention Division of Viral Diseases, Pathogen Discovery | Dakota Howard, Dhwani Batra, Peter W. Cook, Kara Moser, Adrian Paskey, Jason Caravas, Benjamin Rambo-Martin, Shatavia Morrison, Christopher Gulvick, Scott Sammons, Yvette Unoarumhi, Darlene Wagner, Matthew Schmerer, S. H. Rosenthal, A. Gerasimova, R. M. Kagan, B. Anderson, M. Hua, Y. Liu, L.E. Bernstein, K.E. Livingston, A. Perez, I. A. Shlyakhter, R. V. Rolando, R. Owen, P. Tanpaiboon, F. Lacbawan, Clinton R. Paden, Duncan MacCannell |
| EPI_ISL_1615154, EPI_ISL_1615256, EPI_ISL_1615414, EPI_ISL_1615468 | Helix/Illumina | Centers for Disease Control and Prevention Division of Viral Diseases, Pathogen Discovery | Dakota Howard, Dhwani Batra, Peter W. Cook, Kara Moser, Adrian Paskey, Jason Caravas, Benjamin Rambo-Martin, Shatavia Morrison, Christopher Gulvick, Scott Sammons, Yvette Unoarumhi, Darlene Wagner, Matthew Schmerer, Eileen de Feo, Jan Antico, Christine Tran, Matthew Tolentino, Shannon Wickline, Kim Gietzen, Brad Sickler, Jingtao Liu, Eric Allen, Phil Febbo, Nicole L. Washington, Simon White, Geraint Levan, Kelly Schiabor Barrett, Elizabeth Cirulli, Alexandre Bolze, Ary Ascencio, Charlotte Rivera-Garcia, Ryan Cho, Jason Nguyen, Sherry Wang, Jimmy Ramirez, Tyler Cassens, Efren Sandoval, Magnus Isaksson, William Lee, David Becker, Marc Laurent, James Lu, Clinton R. Paden, Duncan MacCannell |
| EPI_ISL_1620452 | National Virus Reference Laboratory | National Virus Reference Laboratory | Zoe Yandle, Charlene Bennett, Gabriel Gonzalez, Michael Carr, Jonathan Dean, Cillian F De Gascun |
| EPI_ISL_1620596, EPI_ISL_1620598, EPI_ISL_1620607 | Puerto Rico Department of Health | Centers for Disease Control and Prevention, Dengue Branch | Gilberto A. Santiago, Glenda Gonzalez, Betzabel Flores, Keyla Charriez, Gabriela Paz-Bailey, Jorge L. Munoz-Jordan |
| EPI_ISL_1620677 | North Dakota Department of Health, Public Health Laboratory | North Dakota Department of Health, Public Health Laboratory | Lisa Wingerter |
| EPI_ISL_1621723, EPI_ISL_1621856, EPI_ISL_1621871, EPI_ISL_1621875, EPI_ISL_1621879, EPI_ISL_1621880, EPI_ISL_1621883, EPI_ISL_1621906, EPI_ISL_1621921, EPI_ISL_1621928, EPI_ISL_1621929, EPI_ISL_1621934, EPI_ISL_1621942, EPI_ISL_1621958, EPI_ISL_1621992, EPI_ISL_1622003, EPI_ISL_1622030, EPI_ISL_1622032, EPI_ISL_1622076, EPI_ISL_1622095, EPI_ISL_1622103 | | | |
| see above | Quest Diagnostics Incorporated | Centers for Disease Control and Prevention Division of Viral Diseases, Pathogen Discovery | Dakota Howard, Dhwani Batra, Peter W. Cook, Kara Moser, Adrian Paskey, Jason Caravas, Benjamin Rambo-Martin, Shatavia Morrison, Christopher Gulvick, Scott Sammons, Yvette Unoarumhi, Darlene Wagner, Matthew Schmerer, S. H. Rosenthal, A. Gerasimova, R. M. Kagan, B. Anderson, M. Hua, Y. Liu, L.E. Bernstein, K.E. Livingston, A. Perez, I. A. Shlyakhter, R. V. Rolando, R. Owen, P. Tanpaiboon, F. Lacbawan, Clinton R. Paden, Duncan MacCannell |
| EPI_ISL_1623702 | Oregon State Public Health Laboratory | Oregon State Public Health Laboratory | Rafia Razzaque, Eugene Yeboah, Vanda Makris, Laura Tsaknaridis, John Fontana and Shane Sevey |
| EPI_ISL_1623955, EPI_ISL_1623956, EPI_ISL_1623960, EPI_ISL_1623963, EPI_ISL_1623965, EPI_ISL_1623967, EPI_ISL_1623968, EPI_ISL_1623971 | MONTEFIORE MEDICAL CENTER LABORATORIES | Wadsworth Center, New York State Department of Health | Kirsten St. George, Daryl M. Lamson, Alexis Russell, Matthew Shudt, Melissa A Leisner, Jonathan Plitnick, Catharine Prussing, Navjot Singh, John Kelly, Erasmus Schneider, Erica Lasek-Nesselquist |
| EPI_ISL_1623973 | NYC Pandemic Response Lab | Wadsworth Center, New York State Department of Health | Kirsten St. George, Daryl M. Lamson, Alexis Russell, Matthew Shudt, Melissa A Leisner, Jonathan Plitnick, Catharine Prussing, Navjot Singh, John Kelly, Erasmus Schneider, Erica Lasek-Nesselquist |
| EPI_ISL_1623975, EPI_ISL_1623977, EPI_ISL_1623979, EPI_ISL_1623983, EPI_ISL_1623984, EPI_ISL_1623987, EPI_ISL_1623988 | MONTEFIORE MEDICAL CENTER LABORATORIES | Wadsworth Center, New York State Department of Health | Kirsten St. George, Daryl M. Lamson, Alexis Russell, Matthew Shudt, Melissa A Leisner, Jonathan Plitnick, Catharine Prussing, Navjot Singh, John Kelly, Erasmus Schneider, Erica Lasek-Nesselquist |
| EPI_ISL_1623994, EPI_ISL_1623997 | MEMORIAL SLOAN KETTERING CANCER CENTER | Wadsworth Center, New York State Department of Health | Kirsten St. George, Daryl M. Lamson, Alexis Russell, Matthew Shudt, Melissa A Leisner, Jonathan Plitnick, Catharine Prussing, Navjot Singh, John Kelly, Erasmus Schneider, Erica Lasek-Nesselquist |
| EPI_ISL_1624010 | Columbia University Irving Medical Center | Wadsworth Center, New York State Department of Health | Kirsten St. George, Daryl M. Lamson, Alexis Russell, Matthew Shudt, Melissa A Leisner, Jonathan Plitnick, Catharine Prussing, Navjot Singh, John Kelly, Erasmus Schneider, Erica Lasek-Nesselquist |
| EPI_ISL_1624015, EPI_ISL_1624023, EPI_ISL_1624029, EPI_ISL_1624066, EPI_ISL_1624073 | New York Presbyterian Hospital | Wadsworth Center, New York State Department of Health | Kirsten St. George, Daryl M. Lamson, Alexis Russell, Matthew Shudt, Melissa A Leisner, Jonathan Plitnick, Catharine Prussing, Navjot Singh, John Kelly, Erasmus Schneider, Erica Lasek-Nesselquist |
| EPI_ISL_1624086 | NYC Pandemic Response Lab | Wadsworth Center, New York State Department of Health | Kirsten St. George, Daryl M. Lamson, Alexis Russell, Matthew Shudt, Melissa A Leisner, Jonathan Plitnick, Catharine Prussing, Navjot Singh, John Kelly, Erasmus Schneider, Erica Lasek-Nesselquist |
| EPI_ISL_1624167 | Illinois Department of Public Health - Springfield Lab | Illinois Department of Public Health - Springfield Lab | Bryan Sim, Gordon McCall |

| | | | |
|---|---|---|---|
| EPI_ISL_1624487 | CHAMPLAIN VALLEY PHYSICIANS HOSPITAL MEDICAL CENTER | Wadsworth Center, New York State Department of Health | Kirsten St. George, Daryl M. Lamson, Alexis Russell, Matthew Shudt, Melissa A Leisner, Jonathan Plitnick, Catharine Prussing, Navjot Singh, John Kelly, Erasmus Schneider, Erica Lasek-Nesselquist |
| EPI_ISL_1624704 | Massachusetts State Public Health Laboratory | Massachusetts State Public Health Laboratory | Andrew Lang, Timelia Fink, Glen Gallagher, Sandra Smole |
| EPI_ISL_1625544, EPI_ISL_1625616, EPI_ISL_1625617, EPI_ISL_1625618, EPI_ISL_1625619, EPI_ISL_1625620, EPI_ISL_1625621, EPI_ISL_1625622, EPI_ISL_1625623, EPI_ISL_1625624, EPI_ISL_1625625, EPI_ISL_1625630, EPI_ISL_1625635, EPI_ISL_1625636, EPI_ISL_1625637, EPI_ISL_1625638, EPI_ISL_1625639, EPI_ISL_1625640, EPI_ISL_1625641, EPI_ISL_1625643, EPI_ISL_1625644, EPI_ISL_1625645, EPI_ISL_1625646, EPI_ISL_1625647, EPI_ISL_1625648, EPI_ISL_1625649, EPI_ISL_1625650, EPI_ISL_1625651, EPI_ISL_1625653, EPI_ISL_1625654, EPI_ISL_1625655, EPI_ISL_1625656, EPI_ISL_1625657, EPI_ISL_1625659, EPI_ISL_1625661 | | | |
| see above | The Jackson Laboratory | The Jackson Laboratory | Bergeron D, Renzette N, Adams M, Omerza G, Kelly K, Long J, Li L |
| EPI_ISL_1625773 | Pathology and Laboratory Medicine Institute, Cleveland Clinic, Ohio, USA | Pathology and Laboratory Medicine Institute, Cleveland Clinic, Ohio, USA | Jennifer Starbuck, Joy Nakitandwe, Kristen McDonnell, David Plunkett, Zheng Jin Tu, Jay Brock, Yu-Wei Cheng, Gary Procop, Daniel Rhoads, Daniel H. Farkas, David Bosler |
| EPI_ISL_1625849, EPI_ISL_1625850, EPI_ISL_1625857, EPI_ISL_1625859, EPI_ISL_1625869, EPI_ISL_1625870, EPI_ISL_1625883, EPI_ISL_1625906, EPI_ISL_1625911 | NJDOH, Public Health and Environmental Laboratories | NJ_PHEL | Lindsey Bodnar, Shiv K. Verma, Jacquelyn Deverell, Matthew Scarnati, Mohammad M Ali, Dana Woell, Byeong Jeong |
| EPI_ISL_1626322 | Wisconsin State Laboratory of Hygiene Communicable Disease Division | Wisconsin State Laboratory of Hygiene Communicable Disease Division | Kelsey R. Florek, Abigail C. Shockey |
| EPI_ISL_1626595 | IN State Department of Health Laboratory Services | IN State Department of Health Laboratory Services | Cassandra Campion, Jamie Yeadon, Brian Pope, Lixia Liu, Kyle Brownlee, Melissa Hindenlang, Ankita Kashikar, Mark Glazier |
| EPI_ISL_1627276 | North Dakota Department of Health, Public Health Laboratory | North Dakota Department of Health, Public Health Laboratory | Lisa Wingerter |
| EPI_ISL_1627313 | Idaho Bureau of Laboratories | Idaho Bureau of Laboratories | R. Beukelman, Matthew Charles Burns, Aimee Ceniseros, Robert L. Voermans, Christopher Ball |
| EPI_ISL_1628494, EPI_ISL_1628495, EPI_ISL_1628496 | University of Mississippi Medical Center, Department of Pathology | University of Mississippi Medical Center, Molecular and Genomics Core Facility | Ashley C. Johnson, Ithiel J. Frame, Krishna K. Ayyalasomayajula, Michael R. Garrett, D. Ashley Robinson |
| EPI_ISL_1628685 | Baylor Scott & White - Temple | Baylor Scott & White - Temple | Ari Rao, Linden Morales, Kimberly Walker, Marcus Volz, Shelby Hendrickson, Caitlin Maloney |
| EPI_ISL_1632695 | Virginia Division of Consolidated Laboratory Services | Virginia Division of Consolidated Laboratory Services | Virginia DCLS |
| EPI_ISL_1633254, EPI_ISL_1633255, EPI_ISL_1633257, EPI_ISL_1633258 | Pathology and Laboratory Medicine Institute, Cleveland Clinic, Ohio, USA | Pathology and Laboratory Medicine Institute, Cleveland Clinic, Ohio, USA | Concetta Peck, Joy Nakitandwe, Kristen McDonnell, David Plunkett, Zheng Jin Tu, Jay Brock, Yu-Wei Cheng, Gary Procop, Daniel Rhoads, Daniel H. Farkas, David Bosler |
| EPI_ISL_1633492 | Genetica Molecular and Subdepartamento de Virologia ISP Chile | Instituto de Salud Publica de Chile | Javier Tognarelli, Karen Orostica, Barbara Parra, Loredana Arata, Jaime Lagos, Gisselle Barra, Patricia Bustos, Rodrigo Fasce, Andres Castillo, Jorge Fernandez |
| EPI_ISL_1633686, EPI_ISL_1633687, EPI_ISL_1633699, EPI_ISL_1633704, EPI_ISL_1633724, EPI_ISL_1633732, EPI_ISL_1633742, EPI_ISL_1633744, EPI_ISL_1633746, EPI_ISL_1633761, EPI_ISL_1633762, EPI_ISL_1633767, EPI_ISL_1633779, EPI_ISL_1633781, EPI_ISL_1633786, EPI_ISL_1633791, EPI_ISL_1633792, EPI_ISL_1633793, EPI_ISL_1633797, EPI_ISL_1633799, EPI_ISL_1633804, EPI_ISL_1633820, EPI_ISL_1633827, EPI_ISL_1633832, EPI_ISL_1633843, EPI_ISL_1633844, EPI_ISL_1633855, EPI_ISL_1633872, EPI_ISL_1633892, EPI_ISL_1633899, EPI_ISL_1633911, EPI_ISL_1633914, EPI_ISL_1633921, EPI_ISL_1633949, EPI_ISL_1633975, EPI_ISL_1633992, EPI_ISL_1633997, EPI_ISL_1634001, EPI_ISL_1634021, EPI_ISL_1634032, EPI_ISL_1634034, EPI_ISL_1634052, EPI_ISL_1634060, EPI_ISL_1634068, EPI_ISL_1634074, EPI_ISL_1634077, EPI_ISL_1634079, EPI_ISL_1634080, EPI_ISL_1634086, EPI_ISL_1634093, EPI_ISL_1634100, EPI_ISL_1634110, EPI_ISL_1634113, EPI_ISL_1634118, EPI_ISL_1634123, EPI_ISL_1634135, EPI_ISL_1634152, EPI_ISL_1634157, EPI_ISL_1634161, EPI_ISL_1634162, EPI_ISL_1634171, EPI_ISL_1634180, EPI_ISL_1634182, EPI_ISL_1634187, EPI_ISL_1634188, EPI_ISL_1634191, EPI_ISL_1634194, EPI_ISL_1634200, EPI_ISL_1634203, EPI_ISL_1634204, EPI_ISL_1634205, EPI_ISL_1634210, EPI_ISL_1634214, EPI_ISL_1634215, EPI_ISL_1634222, EPI_ISL_1634224, EPI_ISL_1634253, EPI_ISL_1634261, EPI_ISL_1634267, EPI_ISL_1634271, EPI_ISL_1634307, EPI_ISL_1634311, EPI_ISL_1634313, EPI_ISL_1634322, EPI_ISL_1634323, EPI_ISL_1634328, EPI_ISL_1634330 | | | |
| see above | Pandemic Response Lab - NYC | Pandemic Response Lab, R&D | Henry Lee, Michael Hammerling, Melissa Hopkins, Cybill del Castillo, Shinyoung Clair Kang, William Ward, Pradeep Bugga, Sol Rey, Dylan Law, Katharine Nelson, Haiping Hao, Jon Laurent |
| EPI_ISL_1635351 | Lighthouse Lab in Cambridge | Wellcome Sanger Institute for the COVID-19 Genomics UK (COG-UK) Consortium | Rob Howes, The Lighthouse Lab in Cambridge and Alex Alderton, Roberto Amato, Jeffrey Barrett, Sonia Goncalves, Ewan Harrison, David K. Jackson, Ian Johnston, Dominic Kwiatkowski, Cordelia Langford, John Sillitoe on behalf of the Wellcome Sanger Institute COVID-19 Surveillance Team |
| EPI_ISL_1635451, EPI_ISL_1635463, EPI_ISL_1635466, EPI_ISL_1635476, EPI_ISL_1635479, EPI_ISL_1635485, EPI_ISL_1635488, EPI_ISL_1635494, EPI_ISL_1635499, EPI_ISL_1635504, EPI_ISL_1635508, EPI_ISL_1635511, EPI_ISL_1635521, EPI_ISL_1635527, EPI_ISL_1635546, EPI_ISL_1635547, EPI_ISL_1635551, EPI_ISL_1635565, EPI_ISL_1635572, EPI_ISL_1635574, EPI_ISL_1635579, EPI_ISL_1635580, EPI_ISL_1635587, EPI_ISL_1635588, EPI_ISL_1635597, EPI_ISL_1635598, EPI_ISL_1635601, EPI_ISL_1635602, EPI_ISL_1635605, EPI_ISL_1635606, EPI_ISL_1635610, EPI_ISL_1635633, EPI_ISL_1635641, EPI_ISL_1635642, EPI_ISL_1635651, EPI_ISL_1635657, EPI_ISL_1635661, EPI_ISL_1635664, EPI_ISL_1635669, EPI_ISL_1635677, EPI_ISL_1635683, EPI_ISL_1635684, EPI_ISL_1635693, EPI_ISL_1635694, EPI_ISL_1635699, EPI_ISL_1635705, EPI_ISL_1635707, EPI_ISL_1635714, EPI_ISL_1635717, EPI_ISL_1635718, EPI_ISL_1635722, EPI_ISL_1635735, EPI_ISL_1635739, EPI_ISL_1635742, EPI_ISL_1635751, EPI_ISL_1635755, EPI_ISL_1635758, EPI_ISL_1635773, EPI_ISL_1635774, EPI_ISL_1635778, EPI_ISL_1635779, EPI_ISL_1635780, EPI_ISL_1635787, EPI_ISL_1635789, EPI_ISL_1635791, EPI_ISL_1635795, EPI_ISL_1635797, EPI_ISL_1635799, EPI_ISL_1635803, EPI_ISL_1635804, EPI_ISL_1635816, EPI_ISL_1635824, EPI_ISL_1635833, EPI_ISL_1635836, EPI_ISL_1635839, EPI_ISL_1635841, EPI_ISL_1635843, EPI_ISL_1635848, EPI_ISL_1635849, EPI_ISL_1635851, EPI_ISL_1635854, EPI_ISL_1635868, EPI_ISL_1635874, EPI_ISL_1635876, EPI_ISL_1635889, EPI_ISL_1635898, EPI_ISL_1635903, EPI_ISL_1635905, EPI_ISL_1635918, EPI_ISL_1635928, EPI_ISL_1635945, EPI_ISL_1635946, EPI_ISL_1635955, EPI_ISL_1635958, EPI_ISL_1635962, EPI_ISL_1635965, EPI_ISL_1635969, EPI_ISL_1635977, EPI_ISL_1635987, EPI_ISL_1635991, EPI_ISL_1635994, EPI_ISL_1636003, EPI_ISL_1636016, EPI_ISL_1636026, EPI_ISL_1636027, EPI_ISL_1636029, EPI_ISL_1636042, EPI_ISL_1636051, EPI_ISL_1636056, EPI_ISL_1636063, EPI_ISL_1636078, EPI_ISL_1636079, EPI_ISL_1636104, EPI_ISL_1636105, EPI_ISL_1636106, EPI_ISL_1636108, EPI_ISL_1636116, EPI_ISL_1636118, EPI_ISL_1636120, EPI_ISL_1636123, EPI_ISL_1636127, EPI_ISL_1636134, EPI_ISL_1636138, EPI_ISL_1636143, EPI_ISL_1636149, EPI_ISL_1636152, EPI_ISL_1636156, EPI_ISL_1636164, EPI_ISL_1636165, EPI_ISL_1636170, EPI_ISL_1636171, EPI_ISL_1636193, EPI_ISL_1636206, EPI_ISL_1636207, EPI_ISL_1636219, EPI_ISL_1636221, EPI_ISL_1636229, EPI_ISL_1636230, EPI_ISL_1636232, EPI_ISL_1636237, EPI_ISL_1636244, EPI_ISL_1636247, EPI_ISL_1636252, EPI_ISL_1636258, EPI_ISL_1636271, EPI_ISL_1636273, EPI_ISL_1636275, EPI_ISL_1636276, EPI_ISL_1636278, EPI_ISL_1636288, EPI_ISL_1636290, EPI_ISL_1636297, EPI_ISL_1636303, EPI_ISL_1636309, EPI_ISL_1636310, EPI_ISL_1636311, EPI_ISL_1636320, EPI_ISL_1636327, EPI_ISL_1636329, EPI_ISL_1636338, EPI_ISL_1636340, EPI_ISL_1636346, EPI_ISL_1636354, EPI_ISL_1636356, EPI_ISL_1636368, EPI_ISL_1636376, EPI_ISL_1636380, EPI_ISL_1636388, EPI_ISL_1636389 | | | |
| see above | Pandemic Response Lab - NYC | Pandemic Response Lab, R&D | Henry Lee, Michael Hammerling, Melissa Hopkins, Cybill del Castillo, Shinyoung Clair Kang, William Ward, Pradeep Bugga, Sol Rey, Dylan Law, Katharine Nelson, Haiping Hao, Jon Laurent |
| EPI_ISL_1636461 | Microbiology Department, Laboratori Clínic Metropolitana Nord. Hospital Universitari Germans Trias i Pujol. | Can Ruti SARS-CoV-2 Sequencing Hub (HUGTiP/IrsiCaixa/IGTP) | Marc Noguera-Julian, Pilar Armengol, Ignacio Blanco, Antoni E Bordoy, Francesc Catala-Moll, Pere-Joan Cardona, Maria Casadellà, Cristina Casañ, Gemma Clara, Bonaventura Clotet, Cristina Esteban, Montserrat Giménez, Mercedes Guerrero, Anna Not, Roger Paredes, Mariona Parera, Verónica Saludes, Alba Sánchez, and Elisa Martró on behalf of the Can Ruti SARS-CoV-2 Sequencing Hub. |
| EPI_ISL_1637959 | Wales Specialist Virology Centre Sequencing lab: Pathogen Genomics Unit | Public Health Wales Microbiology Cardiff Wales Specialist Virology Centre | Catherine Moore, Johnathan Evans, Laura Gifford, Malorie Perry, Simon Cottrell, Angela Marchbank, Alec Birchley, Alexander Adams, Amy Gaskin, Bree Gatica-Wilcox, Jason Coombes, Joel Southgate, Lauren Gilbert, Lee Graham, Nicole Pacchiarini, Sara Kumziene-Summerhayes, Sarah Taylor, Sophie Jones, Sara Rey, Matthew Bull, Joanne Watkins, Sally Corden, Tom Connor |
| EPI_ISL_1638578 | UniversitÃ¤tsklinikum Bonn - Institut fÃ¼r Virologie | Robert Koch Institute | unknown |
| EPI_ISL_1647028 | NCSLPH | NCSLPH | Chase K, Miller MC, Greene S, Glover W |
| EPI_ISL_1647857 | Hospital General Universitario Gregorio Marañón | Hospital General Universitario Gregorio Marañón | Sergio Buenestado Serrano, Pedro Sola Campoy, Laura Pérez-Lago, Cristina Rodriguez-Grande, Pilar Catalán, Patricia Muñoz, Darío García de Viedma |
| EPI_ISL_1648094, EPI_ISL_1648105, EPI_ISL_1648106, EPI_ISL_1648109, EPI_ISL_1648110, EPI_ISL_1648111, EPI_ISL_1648118 | NJDOH, Public Health and Environmental Laboratories | NJ_PHEL | Lindsey Bodnar, Shiv K. Verma, Jacquelyn Deverell, Matthew Scarnati, Dana Woell, Byeong Jeong |
| EPI_ISL_1648161 | NCSLPH | NCSLPH | Chase K, Miller MC, Greene S, Glover W |
| EPI_ISL_1648187, EPI_ISL_1648213, EPI_ISL_1648260, EPI_ISL_1648341, EPI_ISL_1648359, EPI_ISL_1648367, EPI_ISL_1648372, EPI_ISL_1648384, EPI_ISL_1648387, EPI_ISL_1648430, EPI_ISL_1648538, EPI_ISL_1648691, EPI_ISL_1648728, EPI_ISL_1648763, EPI_ISL_1648772, EPI_ISL_1648784, EPI_ISL_1648829, EPI_ISL_1648837 | | | |
| see above | Quest Diagnostics Incorporated | Centers for Disease Control and Prevention Division of Viral Diseases, Pathogen Discovery | Dakota Howard, Dhwani Batra, Peter W. Cook, Kara Moser, Adrian Paskey, Jason Caravas, Benjamin Rambo-Martin, Shatavia Morrison, Christopher Gulvick, Scott Sammons, Yvette Unoarumhi, Darlene Wagner, Matthew Schmerer, S. H. Rosenthal, A. Gerasimova, R. M. Kagan, B. Anderson, M. Hua, Y. Liu, L.E. Bernstein, K.E. Livingston, A. Perez, I. A. Shlyakhter, R. V. Rolando, R. Owen, P. Tanpaiboon, F. Lacbawan, Clinton R. Paden, Duncan MacCannell |
| EPI_ISL_1648977, EPI_ISL_1649066, EPI_ISL_1649103, EPI_ISL_1649104, EPI_ISL_1649155, EPI_ISL_1649159, EPI_ISL_1649228, EPI_ISL_1649235, EPI_ISL_1649240, EPI_ISL_1649248, EPI_ISL_1649252, EPI_ISL_1649309, EPI_ISL_1649326, EPI_ISL_1649358, EPI_ISL_1649399, EPI_ISL_1649429, EPI_ISL_1649430, EPI_ISL_1649438, EPI_ISL_1649503, EPI_ISL_1649504, EPI_ISL_1649539, EPI_ISL_1649568, EPI_ISL_1649615, EPI_ISL_1649663, EPI_ISL_1649675, EPI_ISL_1649692, EPI_ISL_1649725, EPI_ISL_1649777, EPI_ISL_1649820, EPI_ISL_1649834, EPI_ISL_1649846, EPI_ISL_1649861, EPI_ISL_1649956, EPI_ISL_1650003, | | | |

| | | | |
|---|---|---|---|
| EPI_ISL_1650006, EPI_ISL_1650056, EPI_ISL_1650061, EPI_ISL_1650094, EPI_ISL_1650118, EPI_ISL_1650200, EPI_ISL_1650237, EPI_ISL_1650252, EPI_ISL_1650287, EPI_ISL_1650433, EPI_ISL_1650472, EPI_ISL_1650482, EPI_ISL_1650541, EPI_ISL_1650558, EPI_ISL_1650559, EPI_ISL_1650601, EPI_ISL_1650608, EPI_ISL_1650634, EPI_ISL_1650676, EPI_ISL_1650779, EPI_ISL_1650811, EPI_ISL_1650819, EPI_ISL_1650825, EPI_ISL_1650834, EPI_ISL_1650839, EPI_ISL_1650854, EPI_ISL_1650931, EPI_ISL_1650949, EPI_ISL_1650953, EPI_ISL_1650985, EPI_ISL_1650987, EPI_ISL_1650990, EPI_ISL_1651069, EPI_ISL_1651077, EPI_ISL_1651092, EPI_ISL_1651098, EPI_ISL_1651154, EPI_ISL_1651183, EPI_ISL_1651185 | | | |
| see above | Aegis Sciences Corporation | Centers for Disease Control and Prevention Division of Viral Diseases, Pathogen Discovery | Dakota Howard, Dhwani Batra, Peter W. Cook, Kara Moser, Adrian Paskey, Jason Caravas, Benjamin Rambo-Martin, Shatavia Morrison, Christopher Gulvick, Scott Sammons, Yvette Unoarumhi, Darlene Wagner, Matthew Schmerer, Cyndi Clark, Patrick Campbell, Rob Case, Vikramsinha Ghorpade, Holly Houdeshell, Ola Kvalvaag, Dillon Nall, Ethan Sanders, Alec Vest, Shaun Westlund, Matthew Hardison, Clinton R. Paden, Duncan MacCannell |
| EPI_ISL_1652490 | Illinois Department of Public Health | Gagnon Lab, Southern Illinois University | Keith Gagnon |
| EPI_ISL_1652527 | Simple Laboratories | Gagnon Lab, Southern Illinois University | Keith Gagnon |
| EPI_ISL_1653130 | Lighthouse Lab in Cambridge | Wellcome Sanger Institute for the COVID-19 Genomics UK (COG-UK) Consortium | Rob Howes, The Lighthouse Lab in Cambridge and Alex Alderton, Roberto Amato, Jeffrey Barrett, Sonia Goncalves, Ewan Harrison, David K. Jackson, Ian Johnston, Dominic Kwiatkowski, Cordelia Langford, John Sillitoe on behalf of the Wellcome Sanger Institute COVID-19 Surveillance Team |
| EPI_ISL_1653774, EPI_ISL_1653783 | Southern Nevada Public Health Laboratory | Southern Nevada Public Health Laboratory | Michael Picker |
| EPI_ISL_1654582, EPI_ISL_1654591, EPI_ISL_1654592, EPI_ISL_1654600, EPI_ISL_1654603, EPI_ISL_1654606, EPI_ISL_1654613, EPI_ISL_1654621, EPI_ISL_1654626, EPI_ISL_1654633, EPI_ISL_1654634, EPI_ISL_1654635, EPI_ISL_1654644, EPI_ISL_1654646 | | | |
| see above | NYU Langone Health | Departments of Pathology and Medicine, New York University School of Medicine | Adriana Heguy, Dacia Dimartino, Emily Guzman, Christian Marier, Peter Meyn, Sitharam Ramaswami, Gael Westby, Paul Zappile, Yutong Zhang, Paolo Cotzia, Guiqing Wang |
| EPI_ISL_1656455 | University of Michigan Clinical Microbiology Laboratory | Lauring Lab, University of Michigan, Department of Microbiology and Immunology | Valesano |
| EPI_ISL_1656734 | Synlab Haut de France | UMR 8199/1283 EGID | Mehdi Derhourhi |
| EPI_ISL_1658632 | Viollier AG | Department of Biosystems Science and Engineering, ETH Zurich | Chaoran Chen, Sarah Nadeau, Catharine Aquino, Ivan Topolsky, Philipp Jablonski, Lara Fuhrmann, David Dreifuss, Katharina Jahn, Andreia Cabral de Gouvea, Maria Domenica Moccia, Simon Gruter, Timothy Sykes, Lennart Opitz, Griffin White, Laura Neff, Doris Popovic, Andrea Patrignani, Jay Tracy, Ralph Schlapbach, Christiane Beckmann, Maurice Redondo, Olivier Kobel, Christoph Noppen, Sophie Seidel, Noemie Santamaria de Souza, Niko Beerenwinkel, Tanja Stadler |
| EPI_ISL_1659640 | National Institute of Public Health - National Institute of Hygiene | 1. Academic Center for Pathomorphological and Genetic-Molecular Diagnostics ltd, Bialystok, Poland 2. National Institute of Public Health - National Institute of Hygiene, Warsaw, Poland | Radosaw Charkiewicz, Jacek Nikliski, Przemysaw Biecek. Joanna Resze, Piotr Majewski, Anetta Sulewska, Piotr Karabowicz, Joanna Kiluk, Janusz Dzicio, Konrad Raczkowski, Katarzyna Zacharczuk, Magdalena Nowakowska, Magorzata Sadkowska-Todys, Tomasz Wokowicz |
| EPI_ISL_1660383 | Houston Health Dept. | Houston Health Dept. | Ryker Penn, Pamela Brown, Adolpho Lara, Yanlai Lai |
| EPI_ISL_1660516 | Orebro University Hospital | Orebro University Hospital | unknown |
| EPI_ISL_1662655 | Delaware Public Health Lab | Delaware Public Health Lab | Rebecca Savage |
| EPI_ISL_1662727 | Marshfield Labs | Pathogen Discovery, Respiratory Viruses Branch, Division of Viral Diseases, Centers for Disease Control and Prevention | Ying Tao, Yan Li, Anna Kelleher, Jing Zhang, Anna Montmayeur, Brian Lynch, Krista Queen, Anna Uehara, Peter Cook, Han Jia Justin Ng, Rachel Marine, Clinton R. Paden, Haibin Wang, Mark Burroughs, Justin Lee, Adam Retchless, Suxiang Tong |
| EPI_ISL_1663075 | Illinois Department of Public Health | Illinois Department of Public Health - Chicago Lab | Vineet K. Dhiman, Ira Heimler |
| EPI_ISL_1664327, EPI_ISL_1664345, EPI_ISL_1664458, EPI_ISL_1664476 | Clinical Molecular Microbiology Laboratory, UNC Hospitals | Jeremy Wang | Jeremy Wang, Alexander Rubinsteyn, Colleen Rice, Jason Smedberg, Shawn Hawken, Melissa Miller, Corbin Jones, Robert Hagan |
| EPI_ISL_1665176 | CONSEJERIA DE SANIDAD Y ASUNTOS SOCIALES | Instituto de Salud Carlos III | Sandonís,V. Vázquez-Morón, S. Iglesias-Caballero, M. Camarero, S. Pozo, F. Casas, I. Jiménez, P. Zaballos, A. Monzón, S. Varona, S. Cuesta, I. GUTIERREZ AVILA, GONZALO |
| EPI_ISL_1665982, EPI_ISL_1666035, EPI_ISL_1666068, EPI_ISL_1666070, EPI_ISL_1666168, EPI_ISL_1666182, EPI_ISL_1666200, EPI_ISL_1666232, EPI_ISL_1666245, EPI_ISL_1666283, EPI_ISL_1666305, EPI_ISL_1666319, EPI_ISL_1666356, EPI_ISL_1666363, EPI_ISL_1666369, EPI_ISL_1666371, EPI_ISL_1666490, EPI_ISL_1666532, EPI_ISL_1666573, EPI_ISL_1666624, EPI_ISL_1666988, EPI_ISL_1667017, EPI_ISL_1667114, EPI_ISL_1667129, EPI_ISL_1667157, EPI_ISL_1667197, EPI_ISL_1667219, EPI_ISL_1667253, EPI_ISL_1667269, EPI_ISL_1667274, EPI_ISL_1667278, EPI_ISL_1667357, EPI_ISL_1667373 | | | |
| see above | Fulgent Genetics | Centers for Disease Control and Prevention Division of Viral Diseases, Pathogen Discovery | Dakota Howard, Dhwani Batra, Peter W. Cook, Kara Moser, Adrian Paskey, Jason Caravas, Benjamin Rambo-Martin, Shatavia Morrison, Christopher Gulvick, Scott Sammons, Yvette Unoarumhi, Darlene Wagner, Matthew Schmerer, Harry Gao, Mickey Li, John Gao, Joseph Fierro, Benafsh Sapra, Becky Tsai, Yan Meng, Doreen Ng, James Xie, Clinton R. Paden, Duncan MacCannell |
| EPI_ISL_1667529, EPI_ISL_1667544, EPI_ISL_1667546, EPI_ISL_1667547, EPI_ISL_1667549, EPI_ISL_1667574, EPI_ISL_1667754, EPI_ISL_1667787, EPI_ISL_1667840, EPI_ISL_1667875, EPI_ISL_1668015, EPI_ISL_1668075, EPI_ISL_1668096, EPI_ISL_1668112, EPI_ISL_1668126, EPI_ISL_1668130, EPI_ISL_1668136, EPI_ISL_1668161, EPI_ISL_1668181, EPI_ISL_1668232, EPI_ISL_1668256, EPI_ISL_1668335, EPI_ISL_1668337, EPI_ISL_1668350, EPI_ISL_1668354, EPI_ISL_1668388 | | | |
| see above | Aegis Sciences Corporation | Centers for Disease Control and Prevention Division of Viral Diseases, Pathogen Discovery | Dakota Howard, Dhwani Batra, Peter W. Cook, Kara Moser, Adrian Paskey, Jason Caravas, Benjamin Rambo-Martin, Shatavia Morrison, Christopher Gulvick, Scott Sammons, Yvette Unoarumhi, Darlene Wagner, Matthew Schmerer, Cyndi Clark, Patrick Campbell, Rob Case, Vikramsinha Ghorpade, Holly Houdeshell, Ola Kvalvaag, Dillon Nall, Ethan Sanders, Alec Vest, Shaun Westlund, Matthew Hardison, Clinton R. Paden, Duncan MacCannell |
| EPI_ISL_1668520, EPI_ISL_1668705, EPI_ISL_1668723, EPI_ISL_1668757, EPI_ISL_1668791, EPI_ISL_1669014, EPI_ISL_1669087 | Quest Diagnostics Incorporated | Centers for Disease Control and Prevention Division of Viral Diseases, Pathogen Discovery | Dakota Howard, Dhwani Batra, Peter W. Cook, Kara Moser, Adrian Paskey, Jason Caravas, Benjamin Rambo-Martin, Shatavia Morrison, Christopher Gulvick, Scott Sammons, Yvette Unoarumhi, Darlene Wagner, Matthew Schmerer, S. H. Rosenthal, A. Gerasimova, R. M. Kagan, B. Anderson, M. Hua, Y. Liu, L.E. Bernstein, K.E. Livingston, A. Perez, I. A. Shlyakhter, R. V. Rolando, R. Owen, P. Tanpaiboon, F. Lacbawan, Clinton R. Paden, Duncan MacCannell |
| EPI_ISL_1671148, EPI_ISL_1671152, EPI_ISL_1671153, EPI_ISL_1671157 | NYC Pandemic Response Lab | Wadsworth Center, New York State Department of Health | Kirsten St. George, Daryl M. Lamson, Alexis Russell, Matthew Shudt, Melissa A Leisner, Jonathan Plitnick, Catharine Prussing, Navjot Singh, John Kelly, Erasmus Schneider, Erica Lasek-Nesselquist |
| EPI_ISL_1671160 | Columbia University Irving Medical Center | Wadsworth Center, New York State Department of Health | Kirsten St. George, Daryl M. Lamson, Alexis Russell, Matthew Shudt, Melissa A Leisner, Jonathan Plitnick, Catharine Prussing, Navjot Singh, John Kelly, Erasmus Schneider, Erica Lasek-Nesselquist |
| EPI_ISL_1671170, EPI_ISL_1671171 | MONTEFIORE MEDICAL CENTER LABORATORIES | Wadsworth Center, New York State Department of Health | Kirsten St. George, Daryl M. Lamson, Alexis Russell, Matthew Shudt, Melissa A Leisner, Jonathan Plitnick, Catharine Prussing, Navjot Singh, John Kelly, Erasmus Schneider, Erica Lasek-Nesselquist |
| EPI_ISL_1671172, EPI_ISL_1671173 | NYC Pandemic Response Lab | Wadsworth Center, New York State Department of Health | Kirsten St. George, Daryl M. Lamson, Alexis Russell, Matthew Shudt, Melissa A Leisner, Jonathan Plitnick, Catharine Prussing, Navjot Singh, John Kelly, Erasmus Schneider, Erica Lasek-Nesselquist |
| EPI_ISL_1671176 | Columbia University Irving Medical Center | Wadsworth Center, New York State Department of Health | Kirsten St. George, Daryl M. Lamson, Alexis Russell, Matthew Shudt, Melissa A Leisner, Jonathan Plitnick, Catharine Prussing, Navjot Singh, John Kelly, Erasmus Schneider, Erica Lasek-Nesselquist |
| EPI_ISL_1671180, EPI_ISL_1671184 | NYC Pandemic Response Lab | Wadsworth Center, New York State Department of Health | Kirsten St. George, Daryl M. Lamson, Alexis Russell, Matthew Shudt, Melissa A Leisner, Jonathan Plitnick, Catharine Prussing, Navjot Singh, John Kelly, Erasmus Schneider, Erica Lasek-Nesselquist |
| EPI_ISL_1671186, EPI_ISL_1671190 | MONTEFIORE MEDICAL CENTER LABORATORIES | Wadsworth Center, New York State Department of Health | Kirsten St. George, Daryl M. Lamson, Alexis Russell, Matthew Shudt, Melissa A Leisner, Jonathan Plitnick, Catharine Prussing, Navjot Singh, John Kelly, Erasmus Schneider, Erica Lasek-Nesselquist |
| EPI_ISL_1671194 | New York Presbyterian Hospital | Wadsworth Center, New York State Department of Health | Kirsten St. George, Daryl M. Lamson, Alexis Russell, Matthew Shudt, Melissa A Leisner, Jonathan Plitnick, Catharine Prussing, Navjot Singh, John Kelly, Erasmus Schneider, Erica Lasek-Nesselquist |
| EPI_ISL_1671314, EPI_ISL_1671332, EPI_ISL_1671335, EPI_ISL_1671342, EPI_ISL_1671345 | TEMPUS LABS INC | Wadsworth Center, New York State Department of Health | Kirsten St. George, Daryl M. Lamson, Alexis Russell, Matthew Shudt, Melissa A Leisner, Jonathan Plitnick, Catharine Prussing, Navjot Singh, John Kelly, Erasmus Schneider, Erica Lasek-Nesselquist |
| EPI_ISL_1671349 | STONY BROOK UNIVERSITY HOSPITAL | Wadsworth Center, New York State Department of Health | Kirsten St. George, Daryl M. Lamson, Alexis Russell, Matthew Shudt, Melissa A Leisner, Jonathan Plitnick, Catharine Prussing, Navjot Singh, John Kelly, Erasmus Schneider, Erica Lasek-Nesselquist |
| EPI_ISL_1671461 | WESTCHESTER MEDICAL CENTER | Wadsworth Center, New York State Department of Health | Kirsten St. George, Daryl M. Lamson, Alexis Russell, Matthew Shudt, Melissa A Leisner, Jonathan Plitnick, Catharine Prussing, Navjot Singh, John Kelly, |

| | | | |
|---|---|---|---|
| | | | Erasmus Schneider, Erica Lasek-Nesselquist |
| EPI_ISL_1671477 | MONTEFIORE MEDICAL CENTER LABORATORIES | Wadsworth Center, New York State Department of Health | Kirsten St. George, Daryl M. Lamson, Alexis Russell, Matthew Shudt, Melissa A Leisner, Jonathan Plitnick, Catharine Prussing, Navjot Singh, John Kelly, Erasmus Schneider, Erica Lasek-Nesselquist |
| EPI_ISL_1671480, EPI_ISL_1671482 | WESTCHESTER MEDICAL CENTER | Wadsworth Center, New York State Department of Health | Kirsten St. George, Daryl M. Lamson, Alexis Russell, Matthew Shudt, Melissa A Leisner, Jonathan Plitnick, Catharine Prussing, Navjot Singh, John Kelly, Erasmus Schneider, Erica Lasek-Nesselquist |
| EPI_ISL_1671484 | ST. FRANCIS HOSPITAL LABORATORY | Wadsworth Center, New York State Department of Health | Kirsten St. George, Daryl M. Lamson, Alexis Russell, Matthew Shudt, Melissa A Leisner, Jonathan Plitnick, Catharine Prussing, Navjot Singh, John Kelly, Erasmus Schneider, Erica Lasek-Nesselquist |
| EPI_ISL_1671488, EPI_ISL_1671490 | WESTCHESTER MEDICAL CENTER | Wadsworth Center, New York State Department of Health | Kirsten St. George, Daryl M. Lamson, Alexis Russell, Matthew Shudt, Melissa A Leisner, Jonathan Plitnick, Catharine Prussing, Navjot Singh, John Kelly, Erasmus Schneider, Erica Lasek-Nesselquist |
| EPI_ISL_1671493 | NORTH SHORE UNIVERSITY HOSPITAL | Wadsworth Center, New York State Department of Health | Kirsten St. George, Daryl M. Lamson, Alexis Russell, Matthew Shudt, Melissa A Leisner, Jonathan Plitnick, Catharine Prussing, Navjot Singh, John Kelly, Erasmus Schneider, Erica Lasek-Nesselquist |
| EPI_ISL_1671495 | BOSTON HEART DIAGNOSTICS CORP | Wadsworth Center, New York State Department of Health | Kirsten St. George, Daryl M. Lamson, Alexis Russell, Matthew Shudt, Melissa A Leisner, Jonathan Plitnick, Catharine Prussing, Navjot Singh, John Kelly, Erasmus Schneider, Erica Lasek-Nesselquist |
| EPI_ISL_1671497 | MONTEFIORE MEDICAL CENTER LABORATORIES | Wadsworth Center, New York State Department of Health | Kirsten St. George, Daryl M. Lamson, Alexis Russell, Matthew Shudt, Melissa A Leisner, Jonathan Plitnick, Catharine Prussing, Navjot Singh, John Kelly, Erasmus Schneider, Erica Lasek-Nesselquist |
| EPI_ISL_1671505, EPI_ISL_1671508 | WESTCHESTER MEDICAL CENTER | Wadsworth Center, New York State Department of Health | Kirsten St. George, Daryl M. Lamson, Alexis Russell, Matthew Shudt, Melissa A Leisner, Jonathan Plitnick, Catharine Prussing, Navjot Singh, John Kelly, Erasmus Schneider, Erica Lasek-Nesselquist |
| EPI_ISL_1671603 | NORTHWELL HEALTH LABORATORIES | Wadsworth Center, New York State Department of Health | Kirsten St. George, Daryl M. Lamson, Alexis Russell, Matthew Shudt, Melissa A Leisner, Jonathan Plitnick, Catharine Prussing, Navjot Singh, John Kelly, Erasmus Schneider, Erica Lasek-Nesselquist |
| EPI_ISL_1671607 | WHITE PLAINS HOSPITAL CENTER LABORATORY | Wadsworth Center, New York State Department of Health | Kirsten St. George, Daryl M. Lamson, Alexis Russell, Matthew Shudt, Melissa A Leisner, Jonathan Plitnick, Catharine Prussing, Navjot Singh, John Kelly, Erasmus Schneider, Erica Lasek-Nesselquist |
| EPI_ISL_1671646 | WESTCHESTER MEDICAL CENTER | Wadsworth Center, New York State Department of Health | Kirsten St. George, Daryl M. Lamson, Alexis Russell, Matthew Shudt, Melissa A Leisner, Jonathan Plitnick, Catharine Prussing, Navjot Singh, John Kelly, Erasmus Schneider, Erica Lasek-Nesselquist |
| EPI_ISL_1671655 | WHITE PLAINS HOSPITAL CENTER LABORATORY | Wadsworth Center, New York State Department of Health | Kirsten St. George, Daryl M. Lamson, Alexis Russell, Matthew Shudt, Melissa A Leisner, Jonathan Plitnick, Catharine Prussing, Navjot Singh, John Kelly, Erasmus Schneider, Erica Lasek-Nesselquist |
| EPI_ISL_1671659 | WESTCHESTER MEDICAL CENTER | Wadsworth Center, New York State Department of Health | Kirsten St. George, Daryl M. Lamson, Alexis Russell, Matthew Shudt, Melissa A Leisner, Jonathan Plitnick, Catharine Prussing, Navjot Singh, John Kelly, Erasmus Schneider, Erica Lasek-Nesselquist |
| EPI_ISL_1671664 | MONTEFIORE MEDICAL CENTER LABORATORIES | Wadsworth Center, New York State Department of Health | Kirsten St. George, Daryl M. Lamson, Alexis Russell, Matthew Shudt, Melissa A Leisner, Jonathan Plitnick, Catharine Prussing, Navjot Singh, John Kelly, Erasmus Schneider, Erica Lasek-Nesselquist |
| EPI_ISL_1671665 | WHITE PLAINS HOSPITAL CENTER LABORATORY | Wadsworth Center, New York State Department of Health | Kirsten St. George, Daryl M. Lamson, Alexis Russell, Matthew Shudt, Melissa A Leisner, Jonathan Plitnick, Catharine Prussing, Navjot Singh, John Kelly, Erasmus Schneider, Erica Lasek-Nesselquist |
| EPI_ISL_1671666 | WESTCHESTER MEDICAL CENTER | Wadsworth Center, New York State Department of Health | Kirsten St. George, Daryl M. Lamson, Alexis Russell, Matthew Shudt, Melissa A Leisner, Jonathan Plitnick, Catharine Prussing, Navjot Singh, John Kelly, Erasmus Schneider, Erica Lasek-Nesselquist |
| EPI_ISL_1671667 | URMC LABS | Wadsworth Center, New York State Department of Health | Kirsten St. George, Daryl M. Lamson, Alexis Russell, Matthew Shudt, Melissa A Leisner, Jonathan Plitnick, Catharine Prussing, Navjot Singh, John Kelly, Erasmus Schneider, Erica Lasek-Nesselquist |
| EPI_ISL_1671668, EPI_ISL_1671670, EPI_ISL_1671673, EPI_ISL_1671675 | NORTHWELL HEALTH LABORATORIES | Wadsworth Center, New York State Department of Health | Kirsten St. George, Daryl M. Lamson, Alexis Russell, Matthew Shudt, Melissa A Leisner, Jonathan Plitnick, Catharine Prussing, Navjot Singh, John Kelly, Erasmus Schneider, Erica Lasek-Nesselquist |
| EPI_ISL_1672290 | M Health Fairview | Minnesota Department of Health, Public Health Laboratory | Alexandra Lorentz, Jacob Garfin, Matt Plumb, and Xiong Wang |
| EPI_ISL_1674536, EPI_ISL_1674539, EPI_ISL_1674554, EPI_ISL_1674556, EPI_ISL_1674559, EPI_ISL_1674564, EPI_ISL_1674568, EPI_ISL_1674578, EPI_ISL_1674583, EPI_ISL_1674585, EPI_ISL_1674587, EPI_ISL_1674588, EPI_ISL_1674596, EPI_ISL_1674600, EPI_ISL_1674603, EPI_ISL_1674610, EPI_ISL_1674612, EPI_ISL_1674619, EPI_ISL_1674621, EPI_ISL_1674626, EPI_ISL_1674628, EPI_ISL_1674631, EPI_ISL_1674634, EPI_ISL_1674635, EPI_ISL_1674636, EPI_ISL_1674637, EPI_ISL_1674654, EPI_ISL_1674657, EPI_ISL_1674661, EPI_ISL_1674667, EPI_ISL_1674668, EPI_ISL_1674673, EPI_ISL_1674675, EPI_ISL_1674677, EPI_ISL_1674690, EPI_ISL_1674693, EPI_ISL_1674694, EPI_ISL_1674696, EPI_ISL_1674697 | | | |
| see above | Yale Clinical Virology Lab | Grubaugh Lab - Yale School of Public Health | Joseph Fauver, Mallery Breban, Isabel Ott, Tara Alpert, Mary Petrone, Anderson Brito, Chantal Vogels, Annie Watkins, Chaney Kalinich, Jessica Rothman, Marie L. Landry, Nathan Grubaugh |
| EPI_ISL_1674702, EPI_ISL_1674704 | Murphy Medical Association | Grubaugh Lab - Yale School of Public Health | Joseph Fauver, Mallery Breban, Isabel Ott, Tara Alpert, Mary Petrone, Anderson Brito, Chantal Vogels, Annie Watkins, Chaney Kalinich, Jessica Rothman, Caleb Neal, Eva Laszlo, Steven Murphy, Nathan Grubaugh |
| EPI_ISL_1674970 | Florida Bureau of Public Health Laboratories | Florida Bureau of Public Health Laboratories | Sarah Schmedes, Jason Blanton |
| EPI_ISL_1678061, EPI_ISL_1678062, EPI_ISL_1678063, EPI_ISL_1678066, EPI_ISL_1678067, EPI_ISL_1678069, EPI_ISL_1678070, EPI_ISL_1678071, EPI_ISL_1678072, EPI_ISL_1678075, EPI_ISL_1678076, EPI_ISL_1678078, EPI_ISL_1678080 | | | |
| see above | NYC Department of Health and Mental Hygiene | Centers for Disease Control and Prevention Division of Viral Diseases, Pathogen Discovery | Mili Sheth, Sarah Nobles, Jasmine Padilla, Mark Burroughs, Shoshona Le, Katie Dillon, Peter Cook, Clinton R. Paden, Dhwani Batra, Krista Queen, Kristen Knipe, Dakota Howard, Yvette Unoarumhi, Darlene Wagner, Matthew Schmerer, Ben L. Rambo-Martin, Kristine Lacek, Sam Shepard, Alison Laufer Halpin, Dave Wentworth, Vivien Dugan, Suxiang Tong, Justin Lee |
| EPI_ISL_1678134, EPI_ISL_1678135, EPI_ISL_1678141 | SC Dept of Health and Env. Control-Bureau of Laboratories | Centers for Disease Control and Prevention Division of Viral Diseases, Pathogen Discovery | Mili Sheth, Sarah Nobles, Jasmine Padilla, Mark Burroughs, Shoshona Le, Katie Dillon, Peter Cook, Clinton R. Paden, Dhwani Batra, Krista Queen, Kristen Knipe, Dakota Howard, Yvette Unoarumhi, Darlene Wagner, Matthew Schmerer, Ben L. Rambo-Martin, Kristine Lacek, Sam Shepard, Alison Laufer Halpin, Dave Wentworth, Vivien Dugan, Suxiang Tong, Justin Lee |
| EPI_ISL_1678161 | "CDPH, Viral and Rickettsial Disease Laboratory" | Centers for Disease Control and Prevention Division of Viral Diseases, Pathogen Discovery | Mili Sheth, Sarah Nobles, Jasmine Padilla, Mark Burroughs, Shoshona Le, Katie Dillon, Peter Cook, Clinton R. Paden, Dhwani Batra, Krista Queen, Kristen Knipe, Dakota Howard, Yvette Unoarumhi, Darlene Wagner, Matthew Schmerer, Ben L. Rambo-Martin, Kristine Lacek, Sam Shepard, Alison Laufer Halpin, Dave Wentworth, Vivien Dugan, Suxiang Tong, Justin Lee |
| EPI_ISL_1678228 | GA Department of Public Health Laboratory | Centers for Disease Control and Prevention Division of Viral Diseases, Pathogen Discovery | Mili Sheth, Sarah Nobles, Jasmine Padilla, Mark Burroughs, Shoshona Le, Katie Dillon, Peter Cook, Clinton R. Paden, Dhwani Batra, Krista Queen, Kristen Knipe, Dakota Howard, Yvette Unoarumhi, Darlene Wagner, Matthew Schmerer, Ben L. Rambo-Martin, Kristine Lacek, Sam Shepard, Alison Laufer Halpin, Dave Wentworth, Vivien Dugan, Suxiang Tong, Justin Lee |
| EPI_ISL_1678894, EPI_ISL_1679008, EPI_ISL_1679014, EPI_ISL_1679020, EPI_ISL_1679021, EPI_ISL_1679023, EPI_ISL_1679052, EPI_ISL_1679621, EPI_ISL_1679741, EPI_ISL_1679760, EPI_ISL_1679776 | | | |
| see above | Helix/Illumina | Centers for Disease Control and Prevention Division of Viral Diseases, Pathogen Discovery | Dakota Howard, Dhwani Batra, Peter W. Cook, Kara Moser, Adrian Paskey, Jason Caravas, Benjamin Rambo-Martin, Shatavia Morrison, Christopher Gulvick, Scott Sammons, Yvette Unoarumhi, Darlene Wagner, Matthew Schmerer, Eileen de Feo, Jan Antico, Christine Tran, Matthew Tolentino, Shannon Wickline, Kim Gietzen, Brad Sickler, Jingtao Liu, Eric Allen, Phil Febbo, Nicole L. Washington, Simon White, Geraint Levan, Kelly Schiabor Barrett, Elizabeth Cirulli, Alexandre Bolze, Ary Ascencio, Charlotte Rivera-Garcia, Ryan Cho, Jason Nguyen, Sherry Wang, Jimmy Ramirez, Tyler Cassens, Efren Sandoval, Magnus Isaksson, William Lee, David Becker, Marc Laurent, James Lu, Clinton R. Paden, Duncan MacCannell |
| EPI_ISL_1680545, EPI_ISL_1680550, EPI_ISL_1680552, EPI_ISL_1680603, EPI_ISL_1680647, EPI_ISL_1680687, EPI_ISL_1680692, EPI_ISL_1680698, EPI_ISL_1680701, EPI_ISL_1680765, EPI_ISL_1680776, EPI_ISL_1680778, EPI_ISL_1680780, EPI_ISL_1680785, EPI_ISL_1680790, EPI_ISL_1680792, EPI_ISL_1680850, EPI_ISL_1680858, EPI_ISL_1680865, EPI_ISL_1680867, EPI_ISL_1680868, EPI_ISL_1681143, EPI_ISL_1681173, EPI_ISL_1681188, EPI_ISL_1681201, EPI_ISL_1681203, EPI_ISL_1681204, EPI_ISL_1681249, EPI_ISL_1681257, EPI_ISL_1681259, EPI_ISL_1681265, EPI_ISL_1681266, EPI_ISL_1681269, EPI_ISL_1681274, EPI_ISL_1681276, EPI_ISL_1681280, EPI_ISL_1681298, EPI_ISL_1681318, EPI_ISL_1681322, EPI_ISL_1681333, EPI_ISL_1681335, EPI_ISL_1681336, EPI_ISL_1681341, EPI_ISL_1681342, EPI_ISL_1681344, EPI_ISL_1681415, EPI_ISL_1681416, EPI_ISL_1681419, EPI_ISL_1681421, EPI_ISL_1681431, EPI_ISL_1681432, EPI_ISL_1681433, EPI_ISL_1681455, EPI_ISL_1681473, EPI_ISL_1681495, EPI_ISL_1681504, EPI_ISL_1681547, EPI_ISL_1681560, EPI_ISL_1681565, EPI_ISL_1681577, EPI_ISL_1681584, EPI_ISL_1681602, EPI_ISL_1681746, EPI_ISL_1681777, EPI_ISL_1681846, EPI_ISL_1681848, EPI_ISL_1681849, EPI_ISL_1681865, EPI_ISL_1682025, EPI_ISL_1682037, EPI_ISL_1682061, EPI_ISL_1682098, EPI_ISL_1682107, EPI_ISL_1682109, EPI_ISL_1682121, EPI_ISL_1682161, EPI_ISL_1682293, EPI_ISL_1682342, EPI_ISL_1682420, EPI_ISL_1682558, EPI_ISL_1682763, EPI_ISL_1682823, EPI_ISL_1682929, EPI_ISL_1682996, EPI_ISL_1683049, EPI_ISL_1683096, EPI_ISL_1683100, EPI_ISL_1683102, EPI_ISL_1683108, EPI_ISL_1683113, EPI_ISL_1683131, EPI_ISL_1683168, EPI_ISL_1683200, EPI_ISL_1683201, EPI_ISL_1683214, EPI_ISL_1683220, EPI_ISL_1683272, EPI_ISL_1683273, EPI_ISL_1683282, EPI_ISL_1683283, EPI_ISL_1683299, EPI_ISL_1683308, EPI_ISL_1683333, EPI_ISL_1683344, EPI_ISL_1683345, EPI_ISL_1683382, EPI_ISL_1683383, EPI_ISL_1683409, EPI_ISL_1683425, EPI_ISL_1683459, EPI_ISL_1683485, EPI_ISL_1683493, EPI_ISL_1683552, EPI_ISL_1683597, EPI_ISL_1683635, EPI_ISL_1683648, EPI_ISL_1683655, EPI_ISL_1683664, EPI_ISL_1683668, | | | |

| | | | |
|---|---|---|---|
| EPI_ISL_1683673, EPI_ISL_1683685, EPI_ISL_1683708, EPI_ISL_1683711, EPI_ISL_1683721, EPI_ISL_1683732, EPI_ISL_1683737, EPI_ISL_1683749, EPI_ISL_1683757, EPI_ISL_1683760, EPI_ISL_1683771, EPI_ISL_1683772, EPI_ISL_1683781, EPI_ISL_1683787, EPI_ISL_1683789, EPI_ISL_1683812, EPI_ISL_1683823, EPI_ISL_1683825, EPI_ISL_1683894, EPI_ISL_1684065, EPI_ISL_1684066, EPI_ISL_1684077, EPI_ISL_1684082, EPI_ISL_1684087, EPI_ISL_1684102, EPI_ISL_1684122, EPI_ISL_1684146, EPI_ISL_1684149, EPI_ISL_1684156, EPI_ISL_1684173, EPI_ISL_1684174, EPI_ISL_1684201, EPI_ISL_1684243, EPI_ISL_1684244, EPI_ISL_1684245, EPI_ISL_1684249, EPI_ISL_1684260, EPI_ISL_1684269, EPI_ISL_1684270, EPI_ISL_1684275, EPI_ISL_1684276, EPI_ISL_1684295, EPI_ISL_1684303, EPI_ISL_1684306, EPI_ISL_1684307, EPI_ISL_1684310, EPI_ISL_1684319, EPI_ISL_1684323, EPI_ISL_1684448, EPI_ISL_1684469, EPI_ISL_1684565, EPI_ISL_1684601, EPI_ISL_1684634, EPI_ISL_1684660, EPI_ISL_1684775, EPI_ISL_1684814, EPI_ISL_1684944, EPI_ISL_1685187, EPI_ISL_1685198, EPI_ISL_1685202 | | | |
| see above | Laboratory Corporation of America | Centers for Disease Control and Prevention Division of Viral Diseases, Pathogen Discovery | Dakota Howard, Dhwani Batra, Peter W. Cook, Kara Moser, Adrian Paskey, Jason Caravas, Benjamin Rambo-Martin, Shatavia Morrison, Christopher Gulvick, Scott Sammons, Yvette Unoarumhi, Darlene Wagner, Matthew Schmerer, Minoo Agarwal, Eyad Almasri, Debbie Boles, Ayla Burns, Nuthawin Charoensri, Oren Cohen, Susan Countryman, Mary Ann Cristobal, Bobbi Croy, Suzanne Dale, Hrushikesh Deshmukh, Amanda Douglas, Vincent Drouillon, Marcia Eisenberg, Howard Engler, Rama Ghatti, Prashant Gupta, Susan Hicks, Jake Humphrey, Lax Iyer, Manoj Jain, Mohan Kolli, Brian Krueger, Tim Kuphal, Stanley Letovsky, Michael Levandoski, Craig Lukasik, Jonathan Meltzer, Brian Norvell, Mindy Nye, Scott Parker, Christos Petropoulos, John Pruitt, Steven Ragan, Scott Ryan, Mike Sapeta, Jana Schroth, Suresh Babu Selvaraju, Goran Stevovic, Amanda Suchanek, Andrea Throop, Lyndon Tilson, Thomas Urban, Joe Voshell, Kimberly Wagner, Jonathan Williams, Mary Williamson, Qian Zeng, Tricia Zwiefelhofer, Clinton R. Paden, Duncan MacCannell |
| EPI_ISL_1685473, EPI_ISL_1685519, EPI_ISL_1685562, EPI_ISL_1685574, EPI_ISL_1685578, EPI_ISL_1685596, EPI_ISL_1685608, EPI_ISL_1685623, EPI_ISL_1685690, EPI_ISL_1685699, EPI_ISL_1685749, EPI_ISL_1685806, EPI_ISL_1685807, EPI_ISL_1685813, EPI_ISL_1685820, EPI_ISL_1685821, EPI_ISL_1685824, EPI_ISL_1685841, EPI_ISL_1685890, EPI_ISL_1685916, EPI_ISL_1685917, EPI_ISL_1685928, EPI_ISL_1685954, EPI_ISL_1685960, EPI_ISL_1685992, EPI_ISL_1685999, EPI_ISL_1686135, EPI_ISL_1686138, EPI_ISL_1686173, EPI_ISL_1686233, EPI_ISL_1686282, EPI_ISL_1686307, EPI_ISL_1686393, EPI_ISL_1686397, EPI_ISL_1686559, EPI_ISL_1686588, EPI_ISL_1686747, EPI_ISL_1686771, EPI_ISL_1686774, EPI_ISL_1686776, EPI_ISL_1686779, EPI_ISL_1686785, EPI_ISL_1686792, EPI_ISL_1686804, EPI_ISL_1686805, EPI_ISL_1686972, EPI_ISL_1686973, EPI_ISL_1686991, EPI_ISL_1687031, EPI_ISL_1687050, EPI_ISL_1687091, EPI_ISL_1687139, EPI_ISL_1687140, EPI_ISL_1687149, EPI_ISL_1687195, EPI_ISL_1687205, EPI_ISL_1687224, EPI_ISL_1687280, EPI_ISL_1687310, EPI_ISL_1687385, EPI_ISL_1687391, EPI_ISL_1687430, EPI_ISL_1687443, EPI_ISL_1687491, EPI_ISL_1687510, EPI_ISL_1687553, EPI_ISL_1687600, EPI_ISL_1687650, EPI_ISL_1687653, EPI_ISL_1687699, EPI_ISL_1687730, EPI_ISL_1687748, EPI_ISL_1687786, EPI_ISL_1687833, EPI_ISL_1687835, EPI_ISL_1687871, EPI_ISL_1687874, EPI_ISL_1687904, EPI_ISL_1687961, EPI_ISL_1688002, EPI_ISL_1688049, EPI_ISL_1688077, EPI_ISL_1688095, EPI_ISL_1688155, EPI_ISL_1688245, EPI_ISL_1688392, EPI_ISL_1688557, EPI_ISL_1688620, EPI_ISL_1688634, EPI_ISL_1688638, EPI_ISL_1688670, EPI_ISL_1688700, EPI_ISL_1688738, EPI_ISL_1688766, EPI_ISL_1688957, EPI_ISL_1688968, EPI_ISL_1689219, EPI_ISL_1689298, EPI_ISL_1689385, EPI_ISL_1689515, EPI_ISL_1689626, EPI_ISL_1689629, EPI_ISL_1689634, EPI_ISL_1689636, EPI_ISL_1689723, EPI_ISL_1689729, EPI_ISL_1689735, EPI_ISL_1689750, EPI_ISL_1689823, EPI_ISL_1689854, EPI_ISL_1689855, EPI_ISL_1689856, EPI_ISL_1689862, EPI_ISL_1689863, EPI_ISL_1689865, EPI_ISL_1689887, EPI_ISL_1689890, EPI_ISL_1689998, EPI_ISL_1690043, EPI_ISL_1690046, EPI_ISL_1690059, EPI_ISL_1690115, EPI_ISL_1690121, EPI_ISL_1690180, EPI_ISL_1690189, EPI_ISL_1690250 | | | |
| see above | Aegis Sciences Corporation | Centers for Disease Control and Prevention Division of Viral Diseases, Pathogen Discovery | Dakota Howard, Dhwani Batra, Peter W. Cook, Kara Moser, Adrian Paskey, Jason Caravas, Benjamin Rambo-Martin, Shatavia Morrison, Christopher Gulvick, Scott Sammons, Yvette Unoarumhi, Darlene Wagner, Matthew Schmerer, Cyndi Clark, Patrick Campbell, Rob Case, Vikramsinha Ghorpade, Holly Houdeshell, Ola Kvalvaag, Dillon Nall, Ethan Sanders, Alec Vest, Shaun Westlund, Matthew Hardison, Clinton R. Paden, Duncan MacCannell |
| EPI_ISL_1690401, EPI_ISL_1690455, EPI_ISL_1690499, EPI_ISL_1690719, EPI_ISL_1690890, EPI_ISL_1690956, EPI_ISL_1690970, EPI_ISL_1691015, EPI_ISL_1691212, EPI_ISL_1691228, EPI_ISL_1691323, EPI_ISL_1691338 | | | |
| see above | Helix/Illumina | Centers for Disease Control and Prevention Division of Viral Diseases, Pathogen Discovery | Dakota Howard, Dhwani Batra, Peter W. Cook, Kara Moser, Adrian Paskey, Jason Caravas, Benjamin Rambo-Martin, Shatavia Morrison, Christopher Gulvick, Scott Sammons, Yvette Unoarumhi, Darlene Wagner, Matthew Schmerer, Eileen de Feo, Jan Antico, Christine Tran, Matthew Tolentino, Shannon Wickline, Kim Gietzen, Brad Sickler, Jingtao Liu, Eric Allen, Phil Febbo, Nicole L. Washington, Simon White, Geraint Levan, Kelly Schiabor Barrett, Elizabeth Cirulli, Alexandre Bolze, Ary Ascencio, Charlotte Rivera-Garcia, Ryan Cho, Jason Nguyen, Sherry Wang, Jimmy Ramirez, Tyler Cassens, Efren Sandoval, Magnus Isaksson, William Lee, David Becker, Marc Laurent, James Lu, Clinton R. Paden, Duncan MacCannell |
| EPI_ISL_1691424, EPI_ISL_1691436, EPI_ISL_1691450 | Quest Diagnostics Incorporated | Centers for Disease Control and Prevention Division of Viral Diseases, Pathogen Discovery | Dakota Howard, Dhwani Batra, Peter W. Cook, Kara Moser, Adrian Paskey, Jason Caravas, Benjamin Rambo-Martin, Shatavia Morrison, Christopher Gulvick, Scott Sammons, Yvette Unoarumhi, Darlene Wagner, Matthew Schmerer, S. H. Rosenthal, A. Gerasimova, R. M. Kagan, B. Anderson, M. Hua, Y. Liu, L.E. Bernstein, K.E. Livingston, A. Perez, I. A. Shlyakhter, R. V. Rolando, R. Owen, P. Tanpaiboon, F. Lacbawan, Clinton R. Paden, Duncan MacCannell |
| EPI_ISL_1692049, EPI_ISL_1692052, EPI_ISL_1692062, EPI_ISL_1692067, EPI_ISL_1692082, EPI_ISL_1692088, EPI_ISL_1692090, EPI_ISL_1692103, EPI_ISL_1692104, EPI_ISL_1692126, EPI_ISL_1692137, EPI_ISL_1692146, EPI_ISL_1692151, EPI_ISL_1692159, EPI_ISL_1692165, EPI_ISL_1692183, EPI_ISL_1692188, EPI_ISL_1692205, EPI_ISL_1692215, EPI_ISL_1692222, EPI_ISL_1692237, EPI_ISL_1692243, EPI_ISL_1692265, EPI_ISL_1692283, EPI_ISL_1692285, EPI_ISL_1692299, EPI_ISL_1692300, EPI_ISL_1692308, EPI_ISL_1692309, EPI_ISL_1692310, EPI_ISL_1692322, EPI_ISL_1692333, EPI_ISL_1692336, EPI_ISL_1692342, EPI_ISL_1692365, EPI_ISL_1692371, EPI_ISL_1692377, EPI_ISL_1692379, EPI_ISL_1692380, EPI_ISL_1692390, EPI_ISL_1692397, EPI_ISL_1692402, EPI_ISL_1692403, EPI_ISL_1692428, EPI_ISL_1692429, EPI_ISL_1692444, EPI_ISL_1692485, EPI_ISL_1692503, EPI_ISL_1692575, EPI_ISL_1692581, EPI_ISL_1692669, EPI_ISL_1692889, EPI_ISL_1693025, EPI_ISL_1693108, EPI_ISL_1693115, EPI_ISL_1693125, EPI_ISL_1693167, EPI_ISL_1693175, EPI_ISL_1693229, EPI_ISL_1693274, EPI_ISL_1693275, EPI_ISL_1693307, EPI_ISL_1693320, EPI_ISL_1693375, EPI_ISL_1693670, EPI_ISL_1693927, EPI_ISL_1693966, EPI_ISL_1693968, EPI_ISL_1694015, EPI_ISL_1694347, EPI_ISL_1694388, EPI_ISL_1694412, EPI_ISL_1694520 | | | |
| see above | Infinity Biologix | Centers for Disease Control and Prevention Division of Viral Diseases, Pathogen Discovery | Dakota Howard, Dhwani Batra, Peter W. Cook, Kara Moser, Adrian Paskey, Jason Caravas, Benjamin Rambo-Martin, Shatavia Morrison, Christopher Gulvick, Scott Sammons, Yvette Unoarumhi, Darlene Wagner, Matthew Schmerer, Christian Bixby, Yihe Wang, Jonathan Schultz, Chirayu Goswami, Russ Hager, Robin Grimwood, Clinton R. Paden, Duncan MacCannell |
| EPI_ISL_1694671 | Quest Diagnostics Incorporated | Centers for Disease Control and Prevention Division of Viral Diseases, Pathogen Discovery | Dakota Howard, Dhwani Batra, Peter W. Cook, Kara Moser, Adrian Paskey, Jason Caravas, Benjamin Rambo-Martin, Shatavia Morrison, Christopher Gulvick, Scott Sammons, Yvette Unoarumhi, Darlene Wagner, Matthew Schmerer, S. H. Rosenthal, A. Gerasimova, R. M. Kagan, B. Anderson, M. Hua, Y. Liu, L.E. Bernstein, K.E. Livingston, A. Perez, I. A. Shlyakhter, R. V. Rolando, R. Owen, P. Tanpaiboon, F. Lacbawan, Clinton R. Paden, Duncan MacCannell |
| EPI_ISL_1694699 | DOHMH Jamaica | New York City Public Health Laboratory | Jade Wang, et al. |
| EPI_ISL_1694701 | DOHMH Riverside | New York City Public Health Laboratory | Jade Wang, et al. |
| EPI_ISL_1694712 | DOHMH Jamaica | New York City Public Health Laboratory | Jade Wang, et al. |
| EPI_ISL_1694713 | DOHMH Corona | New York City Public Health Laboratory | Jade Wang, et al. |
| EPI_ISL_1694719 | DOHMH Fort Greene | New York City Public Health Laboratory | Jade Wang, et al. |
| EPI_ISL_1694722, EPI_ISL_1694726 | OCME Office Of Chief Medical Examiner | New York City Public Health Laboratory | Jade Wang, et al. |
| EPI_ISL_1694728 | DOHMH Crown Heights | New York City Public Health Laboratory | Jade Wang, et al. |
| EPI_ISL_1694729 | DOHMH Morrisania | New York City Public Health Laboratory | Jade Wang, et al. |
| EPI_ISL_1694735 | DOHMH Corona | New York City Public Health Laboratory | Jade Wang, et al. |
| EPI_ISL_1694740, EPI_ISL_1694754, EPI_ISL_1694755, EPI_ISL_1694756 | DOHMH Jamaica | New York City Public Health Laboratory | Jade Wang, et al. |
| EPI_ISL_1694762 | Department of Homeless Services | New York City Public Health Laboratory | Jade Wang, et al. |
| EPI_ISL_1695015 | Laboratory Corporation of America | Centers for Disease Control and Prevention Division of Viral Diseases, Pathogen Discovery | Dakota Howard, Dhwani Batra, Peter W. Cook, Kara Moser, Adrian Paskey, Jason Caravas, Benjamin Rambo-Martin, Shatavia Morrison, Christopher Gulvick, Scott Sammons, Yvette Unoarumhi, Darlene Wagner, Matthew Schmerer, Minoo Agarwal, Eyad Almasri, Debbie Boles, Ayla Burns, Nuthawin Charoensri, Oren Cohen, Susan Countryman, Mary Ann Cristobal, Bobbi Croy, Suzanne Dale, Hrushikesh Deshmukh, Amanda Douglas, Vincent Drouillon, Marcia Eisenberg, Howard Engler, Rama Ghatti, Prashant Gupta, Susan Hicks, Jake Humphrey, Lax Iyer, Manoj Jain, Mohan Kolli, Brian Krueger, Tim Kuphal, Stanley Letovsky, Michael Levandoski, Craig Lukasik, Jonathan Meltzer, Brian Norvell, Mindy Nye, Scott Parker, Christos Petropoulos, John Pruitt, Steven Ragan, Scott Ryan, Mike Sapeta, Jana Schroth, Suresh Babu Selvaraju, Goran Stevovic, Amanda Suchanek, Andrea Throop, Lyndon Tilson, Thomas Urban, Joe Voshell, Kimberly Wagner, Jonathan Williams, Mary Williamson, Qian Zeng, Tricia Zwiefelhofer, Clinton R. Paden, Duncan MacCannell |
| EPI_ISL_1695113, EPI_ISL_1695237, EPI_ISL_1695305, EPI_ISL_1695382, EPI_ISL_1695392, EPI_ISL_1695469, EPI_ISL_1695539, EPI_ISL_1695554 | Aegis Sciences Corporation | Centers for Disease Control and Prevention Division of Viral Diseases, Pathogen Discovery | Dakota Howard, Dhwani Batra, Peter W. Cook, Kara Moser, Adrian Paskey, Jason Caravas, Benjamin Rambo-Martin, Shatavia Morrison, Christopher Gulvick, Scott Sammons, Yvette Unoarumhi, Darlene Wagner, Matthew Schmerer, Cyndi Clark, Patrick Campbell, Rob Case, Vikramsinha Ghorpade, Holly Houdeshell, Ola Kvalvaag, Dillon Nall, Ethan Sanders, Alec Vest, Shaun Westlund, Matthew Hardison, Clinton R. Paden, Duncan MacCannell |
| EPI_ISL_1695787 | NJ Public Health and Environmental Laboratories | Centers for Disease Control and Prevention Division of Viral Diseases, Pathogen Discovery | Mili Sheth, Sarah Nobles, Jasmine Padilla, Mark Burroughs, Shoshona Le, Katie Dillon, Peter Cook, Clinton R. Paden, Dhwani Batra, Krista Queen, Kristen Knipe, Dakota Howard, Yvette Unoarumhi, Darlene Wagner, Matthew Schmerer, Ben L. Rambo-Martin, Kristine Lacek, Sam Shepard, Alison Laufer Halpin, Dave Wentworth, Vivien Dugan, Suxiang Tong, Justin Lee |
| EPI_ISL_1695825 | UT-Unified State Labs: Public Health Utah DOH | Centers for Disease Control and Prevention Division of Viral Diseases, Pathogen Discovery | Mili Sheth, Sarah Nobles, Jasmine Padilla, Mark Burroughs, Shoshona Le, Katie Dillon, Peter Cook, Clinton R. Paden, Dhwani Batra, Krista Queen, Kristen Knipe, Dakota Howard, Yvette Unoarumhi, Darlene Wagner, Matthew Schmerer, Ben L. Rambo-Martin, Kristine Lacek, Sam Shepard, Alison Laufer Halpin, Dave Wentworth, Vivien Dugan, Suxiang Tong, Justin Lee |
| EPI_ISL_1695889, EPI_ISL_1695891 | NYSDOH Wadsworth Center, Virology Lab | Centers for Disease Control and Prevention Division of Viral Diseases, Pathogen Discovery | Mili Sheth, Sarah Nobles, Jasmine Padilla, Mark Burroughs, Shoshona Le, Katie Dillon, Peter Cook, Clinton R. Paden, Dhwani Batra, Krista Queen, Kristen Knipe, Dakota Howard, Yvette Unoarumhi, Darlene Wagner, Matthew Schmerer, Ben L. Rambo-Martin, Kristine Lacek, Sam Shepard, Alison Laufer Halpin, |

| | | | |
|---|---|---|---|
| | | | Dave Wentworth, Vivien Dugan, Suxiang Tong, Justin Lee |
| EPI_ISL_1695908, EPI_ISL_1695910 | NJ Public Health and Environmental Laboratories | Centers for Disease Control and Prevention Division of Viral Diseases, Pathogen Discovery | Mili Sheth, Sarah Nobles, Jasmine Padilla, Mark Burroughs, Shoshona Le, Katie Dillon, Peter Cook, Clinton R. Paden, Dhwani Batra, Krista Queen, Kristen Knipe, Dakota Howard, Yvette Unoarumhi, Darlene Wagner, Matthew Schmerer, Ben L. Rambo-Martin, Kristine Lacek, Sam Shepard, Alison Laufer Halpin, Dave Wentworth, Vivien Dugan, Suxiang Tong, Justin Lee |
| EPI_ISL_1697384 | Wichita State University - Molecular Diagnostics Lab | Kansas Health and Environmental Lab | Mike Grose, Jonathan Barnell, Ben Olsen, and Phil Adam |
| EPI_ISL_1697994, EPI_ISL_1698159 | Lighthouse Lab in Milton Keynes | Wellcome Sanger Institute for the COVID-19 Genomics UK (COG-UK) Consortium | The Lighthouse Lab in Milton Keynes and Alex Alderton, Roberto Amato, Jeffrey Barrett, Sonia Goncalves, Ewan Harrison, David K. Jackson, Ian Johnston, Dominic Kwiatkowski, Cordelia Langford, John Sillitoe on behalf of the Wellcome Sanger Institute COVID-19 Surveillance Team |
| EPI_ISL_1698285, EPI_ISL_1698346 | Lighthouse Lab in Cambridge | Wellcome Sanger Institute for the COVID-19 Genomics UK (COG-UK) Consortium | Rob Howes, The Lighthouse Lab in Cambridge and Alex Alderton, Roberto Amato, Jeffrey Barrett, Sonia Goncalves, Ewan Harrison, David K. Jackson, Ian Johnston, Dominic Kwiatkowski, Cordelia Langford, John Sillitoe on behalf of the Wellcome Sanger Institute COVID-19 Surveillance Team |
| EPI_ISL_1700669, EPI_ISL_1700673 | NYC Department of Health and Mental Hygiene | Pathogen Discovery, Respiratory Viruses Branch, Division of Viral Diseases, Centers for Disease Control and Prevention | Ying Tao, Yan Li, Anna Kelleher, Jing Zhang, Anna Montmayeur, Brian Lynch, Krista Queen, Anna Uehara, Peter Cook, Han Jia Justin Ng, Rachel Marine, Clinton R. Paden, Haibin Wang, Mark Burroughs, Justin Lee, Adam Retchless, Suxiang Tong |
| EPI_ISL_1700702, EPI_ISL_1700706, EPI_ISL_1700707, EPI_ISL_1700709 | NJDOH, Public Health and Environmental Laboratories | NJ_PHEL | Lindsey Bodnar, Shiv K. Verma, Jacquelyn Deverell, Matthew Scarnati, Dana Woell, Byeong Jeong |
| EPI_ISL_1700788 | NJDOH, Public Health and Environmental Laboratories | NJ Public Health and Environmental Laboratories | Lindsey Bodnar, Shiv K. Verma, Jacquelyn Deverell, Matthew Scarnati, Dana Woell, Byeong Jeong |
| EPI_ISL_1701854, EPI_ISL_1701863 | Helix/Illumina | Centers for Disease Control and Prevention Division of Viral Diseases, Pathogen Discovery | Dakota Howard, Dhwani Batra, Peter W. Cook, Kara Moser, Adrian Paskey, Jason Caravas, Benjamin Rambo-Martin, Shatavia Morrison, Christopher Gulvick, Scott Sammons, Yvette Unoarumhi, Darlene Wagner, Matthew Schmerer, Eileen de Feo, Jan Antico, Christine Tran, Matthew Tolentino, Shannon Wickline, Kim Gietzen, Brad Sickler, Jingtao Liu, Eric Allen, Phil Febbo, Nicole L. Washington, Simon White, Geraint Levan, Kelly Schiabor Barrett, Elizabeth Cirulli, Alexandre Bolze, Ary Ascencio, Charlotte Rivera-Garcia, Ryan Cho, Jason Nguyen, Sherry Wang, Jimmy Ramirez, Tyler Cassens, Efren Sandoval, Magnus Isaksson, William Lee, David Becker, Marc Laurent, James Lu, Clinton R. Paden, Duncan MacCannell |
| EPI_ISL_1701884, EPI_ISL_1701900, EPI_ISL_1701915, EPI_ISL_1701949, EPI_ISL_1701952, EPI_ISL_1701972, EPI_ISL_1701976, EPI_ISL_1701978, EPI_ISL_1701996, EPI_ISL_1701997, EPI_ISL_1702037, EPI_ISL_1702055, EPI_ISL_1702076 | | | |
| see above | Aegis Sciences Corporation | Centers for Disease Control and Prevention Division of Viral Diseases, Pathogen Discovery | Dakota Howard, Dhwani Batra, Peter W. Cook, Kara Moser, Adrian Paskey, Jason Caravas, Benjamin Rambo-Martin, Shatavia Morrison, Christopher Gulvick, Scott Sammons, Yvette Unoarumhi, Darlene Wagner, Matthew Schmerer, Cyndi Clark, Patrick Campbell, Rob Case, Vikramsinha Ghorpade, Holly Houdeshell, Ola Kvalvaag, Dillon Nall, Ethan Sanders, Alec Vest, Shaun Westlund, Matthew Hardison, Clinton R. Paden, Duncan MacCannell |
| EPI_ISL_1702189, EPI_ISL_1702190, EPI_ISL_1702192, EPI_ISL_1702194, EPI_ISL_1702267, EPI_ISL_1702272, EPI_ISL_1702275, EPI_ISL_1702278, EPI_ISL_1702534, EPI_ISL_1702542 | Infinity Biologix | Centers for Disease Control and Prevention Division of Viral Diseases, Pathogen Discovery | Dakota Howard, Dhwani Batra, Peter W. Cook, Kara Moser, Adrian Paskey, Jason Caravas, Benjamin Rambo-Martin, Shatavia Morrison, Christopher Gulvick, Scott Sammons, Yvette Unoarumhi, Darlene Wagner, Matthew Schmerer, Christian Bixby, Yihe Wang, Jonathan Schultz, Chirayu Goswami, Russ Hager, Robin Grimwood, Clinton R. Paden, Duncan MacCannell |
| EPI_ISL_1702580, EPI_ISL_1702602, EPI_ISL_1702611, EPI_ISL_1702612, EPI_ISL_1702614, EPI_ISL_1702615, EPI_ISL_1702636, EPI_ISL_1702652, EPI_ISL_1702663, EPI_ISL_1702664, EPI_ISL_1702675, EPI_ISL_1702681, EPI_ISL_1702706, EPI_ISL_1702712 | | | |
| see above | Quest Diagnostics Incorporated | Centers for Disease Control and Prevention Division of Viral Diseases, Pathogen Discovery | Dakota Howard, Dhwani Batra, Peter W. Cook, Kara Moser, Adrian Paskey, Jason Caravas, Benjamin Rambo-Martin, Shatavia Morrison, Christopher Gulvick, Scott Sammons, Yvette Unoarumhi, Darlene Wagner, Matthew Schmerer, S. H. Rosenthal, A. Gerasimova, R. M. Kagan, B. Anderson, M. Hua, Y. Liu, L.E. Bernstein, K.E. Livingston, A. Perez, I. A. Shlyakhter, R. V. Rolando, R. Owen, P. Tanpaiboon, F. Lacbawan, Clinton R. Paden, Duncan MacCannell |
| EPI_ISL_1702788, EPI_ISL_1702799, EPI_ISL_1702800, EPI_ISL_1702802, EPI_ISL_1702847, EPI_ISL_1702855, EPI_ISL_1702867, EPI_ISL_1702876, EPI_ISL_1702912 | Aegis Sciences Corporation | Centers for Disease Control and Prevention Division of Viral Diseases, Pathogen Discovery | Dakota Howard, Dhwani Batra, Peter W. Cook, Kara Moser, Adrian Paskey, Jason Caravas, Benjamin Rambo-Martin, Shatavia Morrison, Christopher Gulvick, Scott Sammons, Yvette Unoarumhi, Darlene Wagner, Matthew Schmerer, Cyndi Clark, Patrick Campbell, Rob Case, Vikramsinha Ghorpade, Holly Houdeshell, Ola Kvalvaag, Dillon Nall, Ethan Sanders, Alec Vest, Shaun Westlund, Matthew Hardison, Clinton R. Paden, Duncan MacCannell |
| EPI_ISL_1703434 | Dutch COVID-19 response team | National Institute for Public Health and the Environment (RIVM) | Adam Meijer, Harry Vennema, Dirk Eggink, Jeroen Cremer, Sharon van den Brink, Bas van der Veer, AnneMarie van den Brandt, Lisa Wijsman, Kim Freriks, Ryanne Jaarsma, Euníce Then, Jolienke Hardeman, Lynn Aarts, Sanne Bos, Melissa van Tuil, Robert Kohl, Linda van de Nes, Sjoerd Kuiling, James Groot, Florian Zwagemaker, Dennis Schmitz, Annelies Kroneman, Karim Hajji, Chantal Reusken, on behalf of the national COVID-19 response team |
| EPI_ISL_1704708 | Northwestern Memorial Hospital | Northwestern University - Ozer Lab | Ramon Lorenzo-Redondo, Lacy M. Simons, Taylor J. Dean, Chad J. Achenbach, Lawrence J. Jennings, Chao Qi, Michael G. Ison, Judd F. Hultquist, Egon A. Ozer |
| EPI_ISL_1704740 | Lurie Children's Hospital of Chicago | Northwestern University - Ozer Lab | Ramon Lorenzo-Redondo, Lacy M. Simons, Taylor J. Dean, Michael G. Ison, Xiaotian, Zheng, William J. Muller, Larry K. Kociolek, Judd F. Hultquist, Egon A. Ozer |
| EPI_ISL_1704775 | Northwestern Memorial Hospital | Northwestern University - Ozer Lab | Ramon Lorenzo-Redondo, Lacy M. Simons, Taylor J. Dean, Chad J. Achenbach, Lawrence J. Jennings, Chao Qi, Michael G. Ison, Judd F. Hultquist, Egon A. Ozer |
| EPI_ISL_1707095 | USC Clinical Lab | Los Angeles County PHL | P. Hemarajata et al. |
| EPI_ISL_1707373 | Sonora Quest Laboratories | TGen North | "Jolene Bowers, Heather Centner, Chris French, Hayley Yaglom, Ashlyn Pfeiffer, Darrin Lemmer, Dave Engelthaler, The Arizona COVID Genomics Union (ACGU)" |
| EPI_ISL_1708250 | Illinois Department of Public Health - Springfield Lab | Illinois Department of Public Health - Springfield Lab | Bryan Sim, Gordon McCall |
| EPI_ISL_1708542, EPI_ISL_1708573 | Gundersen Clinical Microbiology Laboratory | Kabara Cancer Research Institute | Craig S. Richmond, Paraic A. Kenny |
| EPI_ISL_671745 | DOHMH Riverside | New York City Public Health Laboratory | Jade Wang, et al. |
| EPI_ISL_765493 | MONTEFIORE MEDICAL CENTER LABORATORIES | Wadsworth Center, New York State Department.of Health | Kirsten St. George, Daryl M. Lamson, Alexis Russel, Matthew Shudt, Melissa A Leisner, Jonathan Plitnick, Navjot Singh, John Kelly, Sara Griesemer, Erasmus Schneider, Erica Lasek-Nesselquist |
| EPI_ISL_794288 | Columbia University Irving Medical Center | Wadsworth Center, New York State Department.of Health | Kirsten St. George, Daryl M. Lamson, Alexis Russel, Matthew Shudt, Melissa A Leisner, Jonathan Plitnick, Navjot Singh, John Kelly, Sara Griesemer, Erasmus Schneider, Erica Lasek-Nesselquist |
| EPI_ISL_801975 | MSHS Clinical Microbiology Laboratories | MSHS Pathogen Surveillance Program | Ana S. Gonzalez-Reiche, Hala Alshammary, Mitchell J. Sullivan, Brianne Ciferri, Ajay Obla, Angela Amoako, Mahmoud Awawda, Elena Hirsch, Ashley S. Salimbangon, Levy Sominsky, Katherine Beach, Kayla Russo, Charles Gleason, Shelcie Fabre, Giulio Kleiner, Zenab Khan, Bremy Alburquerque, Adriana van de Guchte, Komal Srivastava, Matthew M. Hernandez, Jayeeta Dutta, Denise Jurczyszak, Emily Ferreri, Rachel Chernet, Nancy Francoeur, Betsaida Salom Melo, Irina Oussenko, Gintaras Deikus, Juan Soto, Shwetha Hara Sridhar, Ying-Chih Wang, Kathryn Twyman, Andrew Kasarskis, Deena R. Altman, Robert Sebra, Adolfo García-Sastre, Marta Luksza, Gopi Patel, Sarah Schaefer, Melissa Gitman, Michael D. Nowak, Alberto Paniz-Mondolfi, Emilia Mia Sordillo, Viviana Simon, Harm van Bakel |
| EPI_ISL_802788 | Columbia University Irving Medical Center | Wadsworth Center, New York State Department.of Health | Kirsten St. George, Daryl M. Lamson, Alexis Russel, Matthew Shudt, Melissa A Leisner, Jonathan Plitnick, Navjot Singh, John Kelly, Sara Griesemer, Erasmus Schneider, Erica Lasek-Nesselquist |
| EPI_ISL_812733 | DOHMH Corona | New York City Public Health Laboratory | Jade Wang, et al. |
| EPI_ISL_833137 | Laboratorio de Ecologia de Doencas Transmissiveis na Amazonia, Instituto Leonidas e Maria Deane - Fiocruz Amazonia | Laboratorio de Ecologia de Doencas Transmissiveis na Amazonia, Instituto Leonidas e Maria Deane - Fiocruz Amazonia | Valdinete Nascimento, Victor Souza, André Corado, Fernanda Nascimento, George Silva, Ágatha Costa, Debora Duarte, Karina Pessoa, Matilde Mejía, Luciana Gonçalves, Maria Júlia Brandão, Michele Jesus, Felipe Naveca on behalf of the Fiocruz COVID-19 Genomic Surveillance Network |
| EPI_ISL_854454, EPI_ISL_854455, EPI_ISL_854457 | MONTEFIORE MEDICAL CENTER LABORATORIES | Wadsworth Center, New York State Department of Health | Kirsten St. George, Daryl M. Lamson, Alexis Russel, Matthew Shudt, Melissa A Leisner, Jonathan Plitnick, Navjot Singh, John Kelly, Erasmus Schneider, Erica Lasek-Nesselquist |

| | | | |
|---|---|---|---|
| EPI_ISL_857056, EPI_ISL_857060 | OCME Office Of Chief Medical Examiner | New York City Public Health Laboratory | Jade Wang, et al. |
| EPI_ISL_857163 | DOHMH Central Harlem | New York City Public Health Laboratory | Jade Wang, et al. |
| EPI_ISL_857164 | DOHMH Morrisania | New York City Public Health Laboratory | Jade Wang, et al. |
| EPI_ISL_861112 | New York Presbyterian Hospital | Wadsworth Center, New York State Department of Health | Kirsten St. George, Daryl M. Lamson, Alexis Russel, Matthew Shudt, Melissa A Leisner, Jonathan Plitnick, Navjot Singh, John Kelly, Erasmus Schneider, Erica Lasek-Nesselquist |
| EPI_ISL_861244, EPI_ISL_861267, EPI_ISL_861278, EPI_ISL_861280, EPI_ISL_861291, EPI_ISL_861308, EPI_ISL_861313 | MONTEFIORE MEDICAL CENTER LABORATORIES | Wadsworth Center, New York State Department of Health | Kirsten St. George, Daryl M. Lamson, Alexis Russel, Matthew Shudt, Melissa A Leisner, Jonathan Plitnick, Navjot Singh, John Kelly, Erasmus Schneider, Erica Lasek-Nesselquist |
| EPI_ISL_861353, EPI_ISL_861361 | WESTCHESTER MEDICAL CENTER | Wadsworth Center, New York State Department of Health | Kirsten St. George, Daryl M. Lamson, Alexis Russel, Matthew Shudt, Melissa A Leisner, Jonathan Plitnick, Navjot Singh, John Kelly, Erasmus Schneider, Erica Lasek-Nesselquist |
| EPI_ISL_883333 | DOHMH Riverside | New York City Public Health Laboratory | Jade Wang, et al. |
| EPI_ISL_883339 | DOHMH Chelsea | New York City Public Health Laboratory | Jade Wang, et al. |
| EPI_ISL_883340, EPI_ISL_883342 | DOHMH PHL | New York City Public Health Laboratory | Jade Wang, et al. |
| EPI_ISL_883344 | OCME Office Of Chief Medical Examiner | New York City Public Health Laboratory | Jade Wang, et al. |
| EPI_ISL_883346 | DOHMH PHL | New York City Public Health Laboratory | Jade Wang, et al. |
| EPI_ISL_884080 | Wadsworth Center, New York State Department of Health | Wadsworth Center, New York State Department of Health | Kirsten St. George, Daryl M. Lamson, Alexis Russel, Matthew Shudt, Melissa A Leisner, Jonathan Plitnick, Navjot Singh, John Kelly, Erasmus Schneider, Erica Lasek-Nesselquist |
| EPI_ISL_886531, EPI_ISL_888376 | Labcorp | Genomics and Discovery, Respiratory Viruses Branch, Division of Viral Diseases, Centers for Disease Control and Prevention | Peter W. Cook,Dhwani Batra,Ben L. Rambo-Martin,Summer Galloway,Brian Krueger,Minoo Agarwal,Eyad Almasri,Debbie Boles,Ayla Burns,Nuthawin Charoensri,Oren Cohen,Susan Countryman,Mary Ann Cristobal,Bobbi Croy,Suzanne Dale,Hrushikesh Deshmukh,Amanda Douglas,Vincent Drouillon,Marcia Eisenberg,Howard Engler,Rama Ghatti,Prashant Gupta,Susan Hicks,Jake Humphrey,Lax Iyer,Manoj Jain,Mohan Kolli,Tim Kuphal,Stanley Letovsky,Michael Levandoski,Craig Lukasik,Jonathan Meltzer,Brian Norvell,Mindy Nye,Scott Parker,Christos Petropoulos,John Pruitt,Steven Ragan,Scott Ryan,Mike Sapeta,Jana Schroth,Suresh Babu Selvaraju,Goran Stevovic,Amanda Suchanek,Andrea Throop,Lyndon Tilson,Thomas Urban,Joe Voshell,Kimberly Wagner,Jonathan Williams,Mary Williamson,Qian Zeng,Tricia Zwiefelhofer,Clinton R. Paden,Suxiang Tong,Duncan MacCannell, |
| EPI_ISL_896227, EPI_ISL_896232, EPI_ISL_896233, EPI_ISL_896395 | Columbia University Irving Medical Center | Wadsworth Center, New York State Department of Health | Kirsten St. George, Daryl M. Lamson, Alexis Russel, Matthew Shudt, Melissa A Leisner, Jonathan Plitnick, Navjot Singh, John Kelly, Erasmus Schneider, Erica Lasek-Nesselquist |
| EPI_ISL_896532 | New York Presbyterian Hospital | Wadsworth Center, New York State Department of Health | Kirsten St. George, Daryl M. Lamson, Alexis Russel, Matthew Shudt, Melissa A Leisner, Jonathan Plitnick, Navjot Singh, John Kelly, Erasmus Schneider, Erica Lasek-Nesselquist |
| EPI_ISL_906837, EPI_ISL_906838, EPI_ISL_906839 | National Public Health Laboratory, National Centre for Infectious Diseases | National Public Health Laboratory, National Centre for Infectious Diseases | Tze Minn Mak, Zhenyang Zhou, Lin Cui, Raymond Tzer Pin Lin |
| EPI_ISL_936256, EPI_ISL_936261, EPI_ISL_936276, EPI_ISL_936292, EPI_ISL_936299 | MONTEFIORE MEDICAL CENTER LABORATORIES | Wadsworth Center, New York State Department of Health | Kirsten St. George, Daryl M. Lamson, Alexis Russel, Matthew Shudt, Melissa A Leisner, Jonathan Plitnick, Navjot Singh, John Kelly, Erasmus Schneider, Erica Lasek-Nesselquist |
| EPI_ISL_937126 | DOHMH Riverside | New York City Public Health Laboratory | Jade Wang, et al. |
| EPI_ISL_937155 | DOHMH PHL | New York City Public Health Laboratory | Jade Wang, et al. |
| EPI_ISL_937190 | OCME Office Of Chief Medical Examiner | New York City Public Health Laboratory | Jade Wang, et al. |
| EPI_ISL_937191 | Department of Homeless Services | New York City Public Health Laboratory | Jade Wang, et al. |
| EPI_ISL_937201 | DOHMH Fort Greene | New York City Public Health Laboratory | Jade Wang, et al. |
| EPI_ISL_937206 | DOHMH Morrisania | New York City Public Health Laboratory | Jade Wang, et al. |
| EPI_ISL_937211, EPI_ISL_937215 | DOHMH PHL | New York City Public Health Laboratory | Jade Wang, et al. |
| EPI_ISL_937218 | DOHMH Chelsea | New York City Public Health Laboratory | Jade Wang, et al. |
| EPI_ISL_937236 | DOHMH Morrisania | New York City Public Health Laboratory | Jade Wang, et al. |
| EPI_ISL_937247, EPI_ISL_937252, EPI_ISL_937255 | OCME Office Of Chief Medical Examiner | New York City Public Health Laboratory | Jade Wang, et al. |
| EPI_ISL_937272 | NYC HH Elmhurst Hospital Medical Center | New York City Public Health Laboratory | Jade Wang, et al. |
| EPI_ISL_966264 | NYU Langone Health | Departments of Pathology and Medicine, New York University School of Medicine | Adriana Heguy, Dacia Dimartino, Emily Guzman, Christian Marier, Peter Meyn, Sitharam Ramaswami, Gael Westby, Paul Zappile, Yutong Zhang, Paolo Cotzia, Guiqing Wang |
| EPI_ISL_966387 | DOHMH Jamaica | New York City Public Health Laboratory | Jade Wang, et al. |
| EPI_ISL_966413 | DOHMH Central Harlem | New York City Public Health Laboratory | Jade Wang, et al. |
| EPI_ISL_966415 | DOHMH Corona | New York City Public Health Laboratory | Jade Wang, et al. |
| EPI_ISL_966420 | DOHMH Morrisania | New York City Public Health Laboratory | Jade Wang, et al. |
| EPI_ISL_966424 | DOHMH Jamaica | New York City Public Health Laboratory | Jade Wang, et al. |
| EPI_ISL_979499 | Eurofins Diatherix | Hudsonalpha Genome Sequencing Center | Jane Grimwood, Melissa Williams, Lori H. Handley, Joshua Stough, Leslie Malone, Stefan Brzezinski, Ada Stewart, Teresa Jones, Jenell Webber, John Lovell, Jennifer Cart, and Jeremy Schmutz |
| EPI_ISL_982461, EPI_ISL_982490, EPI_ISL_983103, EPI_ISL_983114 | MONTEFIORE MEDICAL CENTER LABORATORIES | Wadsworth Center, New York State Department of Health | Kirsten St. George, Daryl M. Lamson, Alexis Russel, Matthew Shudt, Melissa A Leisner, Jonathan Plitnick, Navjot Singh, John Kelly, Erasmus Schneider, Erica Lasek-Nesselquist |
| EPI_ISL_984771, EPI_ISL_984816, EPI_ISL_984817, EPI_ISL_984883, EPI_ISL_984917, EPI_ISL_984927, EPI_ISL_984930, EPI_ISL_984945, EPI_ISL_994803, EPI_ISL_994804, EPI_ISL_994824, EPI_ISL_994826, EPI_ISL_994830, EPI_ISL_994844, EPI_ISL_994853, EPI_ISL_994862, EPI_ISL_994870, EPI_ISL_994872, EPI_ISL_994876, EPI_ISL_994880, EPI_ISL_994890, EPI_ISL_994895, EPI_ISL_994925, EPI_ISL_994943, EPI_ISL_994959, EPI_ISL_994960, EPI_ISL_994966, EPI_ISL_994968, EPI_ISL_994970, EPI_ISL_994974, EPI_ISL_994986, EPI_ISL_994988, EPI_ISL_995010, EPI_ISL_995023, EPI_ISL_995026, EPI_ISL_995040, EPI_ISL_995046, EPI_ISL_995048, EPI_ISL_995060, EPI_ISL_995072, EPI_ISL_995094, EPI_ISL_995108, EPI_ISL_995110, EPI_ISL_995111, EPI_ISL_995123, EPI_ISL_995131, EPI_ISL_995147, EPI_ISL_995154, EPI_ISL_995155 | | | |
| see above | Pandemic Response Lab - NYC | Pandemic Response Lab, R&D | Henry Lee, Michael Hammerling, Melissa Hopkins, Cybill del Castillo, William Ward, Pradeep Bugga, Haiping Hao, Jon Laurent |
