## Supplementary figures and images for "Combining genomic and epidemiological data to compare the transmissibility of SARS-CoV-2 lineages"

### Supplementary Figure 1

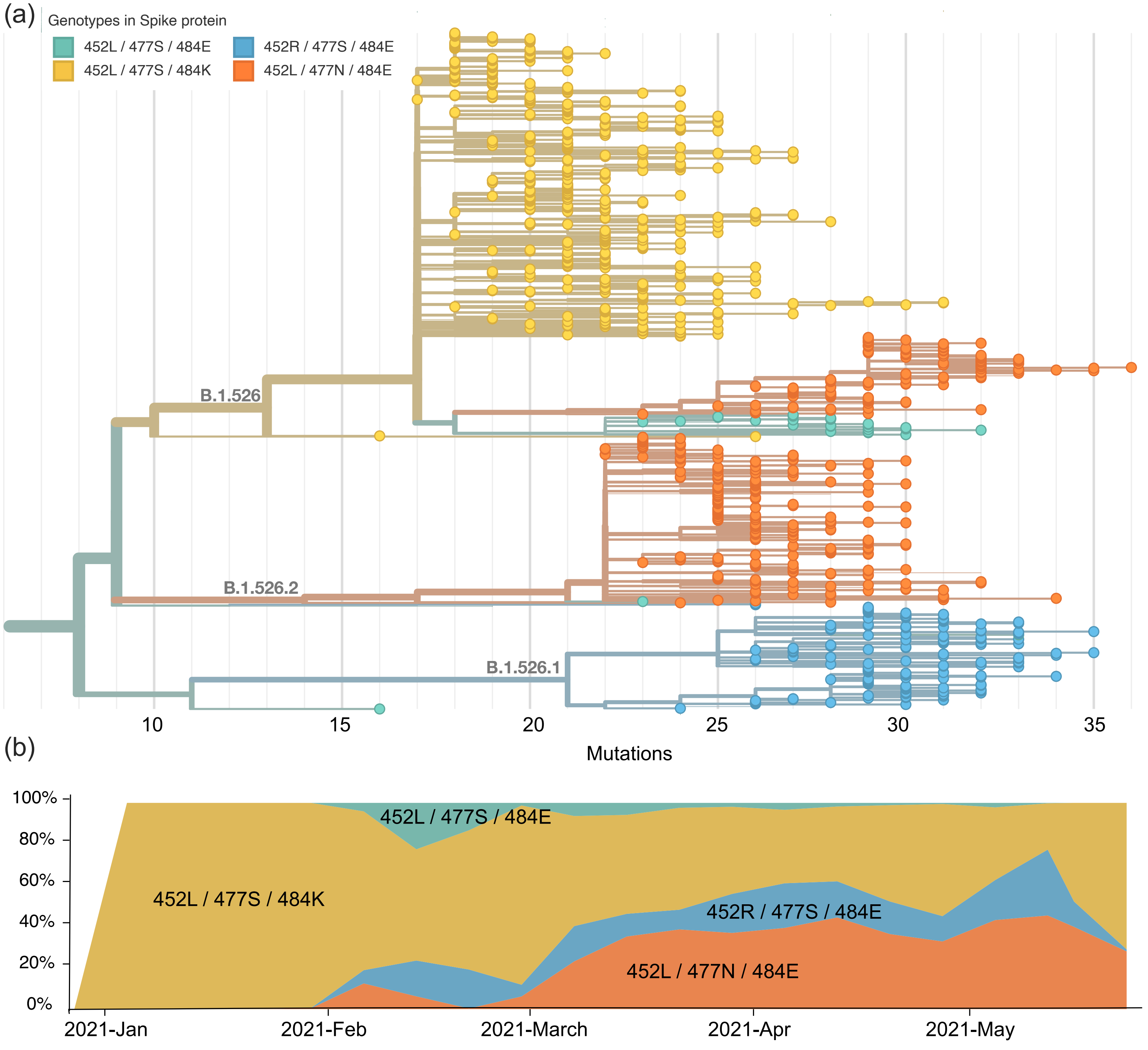

### Supplementary Figure 2

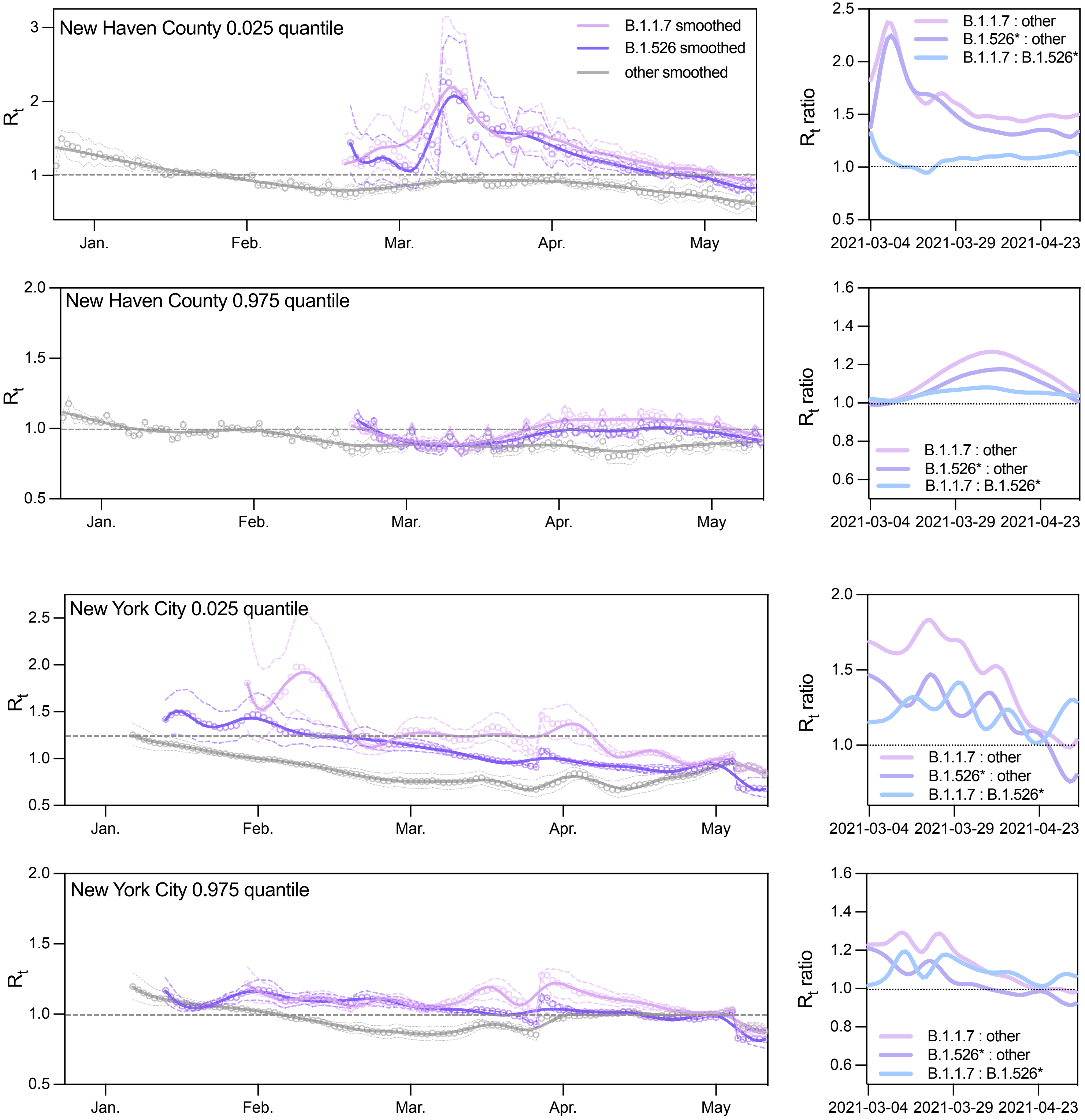

### Supplementary Figure 3

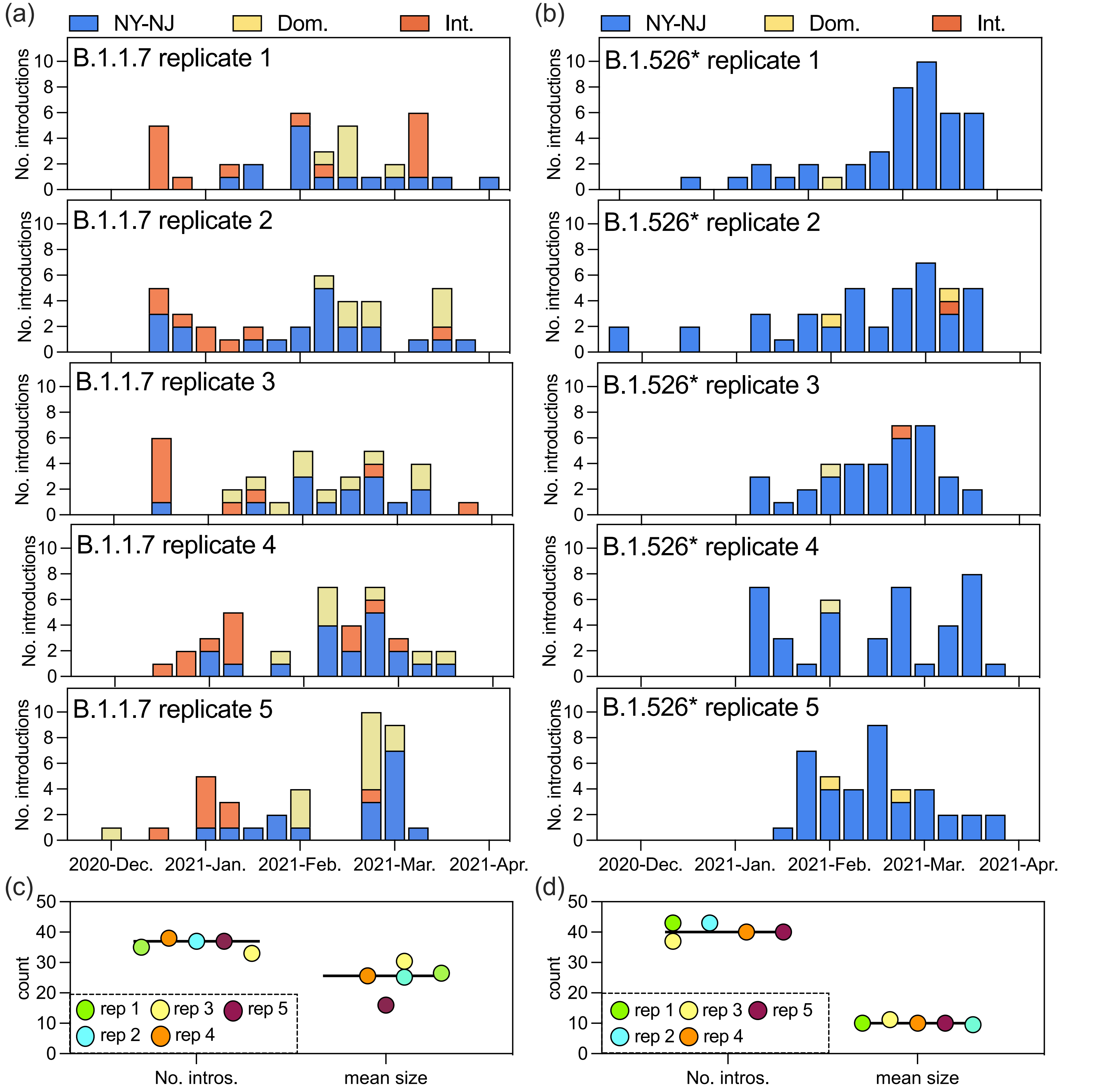
