## Supplementary Table 1 for "Combining genomic and epidemiological data to compare the transmissibility of SARS-CoV-2 lineages"

| Lineage | Count | | Lineage | Count |
|---|---|---|---|---|
| A | 1 | | B.1.36.10 | 1 |
| A.2.5 | 1 | | B.1.361 | 1 |
| A.23.1 | 1 | | B.1.369 | 1 |
| B | 1 | | B.1.37 | 1 |
| B.1 | 25 | | B.1.375 | 2 |
| B.1.1 | 12 | | B.1.400 | 2 |
| B.1.1.1 | 3 | | B.1.409 | 1 |
| B.1.1.192 | 7 | | B.1.420 | 1 |
| B.1.1.28 | 3 | | **B.1.427** | **21** |
| B.1.1.348 | 2 | | **B.1.429** | **42** |
| B.1.1.372 | 1 | | B.1.509 | 3 |
| B.1.1.416 | 1 | | B.1.517 | 106 |
| B.1.1.420 | 1 | | B.1.517.1 | 1 |
| B.1.1.434 | 16 | | B.1.523 | 2 |
| B.1.1.486 | 19 | | **B.1.525** | **4** |
| B.1.1.519 | 17 | | B.1.543 | 1 |
| B.1.110.3 | 4 | | B.1.551 | 1 |
| B.1.111 | 10 | | B.1.568 | 6 |
| B.1.177 | 1 | | B.1.575 | 55 |
| B.1.2 | 159 | | B.1.575.1 | 1 |
| B.1.234 | 2 | | B.1.577 | 2 |
| B.1.240 | 2 | | B.1.588 | 1 |
| B.1.243 | 46 | | B.1.596 | 9 |
| B.1.265 | 1 | | B.1.604 | 1 |
| B.1.280 | 1 | | **B.1.617.2** | **2** |
| B.1.298 | 1 | | B.1.621 | 1 |
| B.1.311 | 2 | | C.37 | 5 |
| B.1.314 | 1 | | **P.1** | **40** |
| B.1.320 | 1 | | R.1 | 33 |
| B.1.349 | 4 | | R.2 | 3 |
| **B.1.351** | **17** | | | |
